## Supplementary Appendix for "Performing arts as a health resource? An umbrella review of the health impacts of music and dance participation"

***Table of Contents***

**Component 1 – Systematic review of systematic reviews of the health benefits of performing arts participation**

*Supplementary methods – GRADE Quality of evidence appraisal* 3

*Table S1. Specific outcome measures assigned to each health domain* 5

*Table S2. Umbrella review level evidence synthesis – all outcomes and GRADE Certainty of Evidence ratings* 10

*Table S3. Systematic review level evidence synthesis for each outcome measure from each included review of dance participation* 20

*Table S4. Systematic review level evidence synthesis for each outcome measure from each included review of music participation* 34

*Table S5. Summary of included systematic reviews* 50

*Table S6. GRADE publication bias ratings for included systematic reviews containing quantitative synthesis (meta-analysis)* 58

*Table S7. Details of primary studies in included systematic reviews of dance participation* 59

*Table S8. Details of primary studies in included systematic reviews of music participation* 104

*Table S9. GRADE Certainty of Evidence ratings for each outcome measure from primary randomized controlled trials of dance participation contained in included reviews* 161

*Table S10. GRADE Certainty of Evidence ratings for each outcome measure from primary non-randomized studies of dance participation contained in included reviews* 181

*Table S11. GRADE Certainty of Evidence ratings for each outcome measure from primary randomized controlled trials of music participation contained in included reviews* 194

*Table S12. GRADE Certainty of Evidence ratings for each outcome measure from primary non-randomized studies of music participation contained in included reviews* 210

*Table S13. Review articles excluded after full-text review, with reasons* 245

**Component 2 – Systematic review of observational studies investigating the impact of performing arts participation on mortality and non-communicable disease risk**

*Table S14. Details of included studies evaluating the impact of performing arts participation on mortality and non-communicable disease risk* 247

*Table S15. GRADE Certainty of Evidence ratings for each outcome measure from included studies evaluating the impact of performing arts participation on mortality and non-communicable disease risk* 249

*Table S16. Studies of mortality and non-communicable disease risk excluded after full-text review* 251

**Component 3 – Systematic review of studies of heart rate responses to performing arts participation**

*Table S17. Details of included studies reporting mean heart rate data during dance participation* 252

*Table S18. Details of included studies reporting mean heart rate data during music participation* 259

*Table S19. Studies of the physical demands of performing arts participation excluded after full-text review, with reasons* 262

**Reference list of included reviews and studies** *(Components 1-3)* 269

#### *Supplementary Methods – GRADE Quality of Evidence Appraisal*

GRADE quality of evidence determinations are based on study design and assessments of five factors which may negatively impact evidence quality: risk of bias; inconsistency; indirectness; imprecision; publication bias. Studies can also be rated higher if a large effect, dose response gradient and/or robust control for all plausible confounding is present. Randomized controlled trials begin the rating process as high-quality evidence, with observational and non-randomized studies beginning as low-quality evidence. For this review, assessment of these factors were conducted in the following manner:

*Study limitations / risk of bias* – assessed for each outcome for each primary study meeting umbrella review inclusion criteria in included reviews, as per GRADE and Cochrane guidelines.[1,2] Specific assessment criteria are per GRADE[3] and detailed in Tables S9-S12.

*Imprecision* – Omitted. Given the general, descriptive and imprecise nature of outcome classifications used in this review (positive, negative, no effect, unclear), assessment of imprecision is irrelevant to evidence quality appraisals in this review.

*Indirectness* – The impact of using surrogate outcomes for health impacts of interest was determined for each outcome at the individual study level. Indirectness related to population, intervention and head-to-head comparisons were addressed by umbrella review inclusion/ exclusion criteria and/or irrelevant to this review.[4]

*Inconsistency* – Omitted. The descriptive methods of outcome classification used in this review serve in lieu of analyses of inconsistency, which are based on analyses of heterogeneity within a pooled effect.[5]

*Publication Bias* – Assessed for included reviews presenting quantitively synthesized results (meta-analysis) only. Assessment of publications bias conducted using the validated criteria developed by Meader et al. for appraisal of publication bias in systematic reviews within the GRADE framework.[6] Rigorous assessment of publication bias for narratively synthesized evidence was precluded by the restriction of inclusion criteria to systematic reviews and further potential exclusion of primary studies from included reviews for not meeting umbrella review inclusion criteria.

*Up-rating* – Conducted as appropriate per GRADE guidelines.[7]

Appraisals were conducted, except for publication bias, first for each outcome at the primary study level. When GRADE was used to assess quality of evidence of primary studies in included reviews, these GRADE ratings were also included in this umbrella review. Otherwise, all GRADE ratings were determined for the purposes of this review.

GRADE ratings for each outcome at the primary study level were then synthesized for each included review, with a GRADE rating assigned to each outcome at the review level. Where applicable, down rating of evidence quality for publication bias was conducted at this stage. GRADE ratings for each outcome at the review level were then further synthesized and a GRADE rating was assigned to each outcome at the umbrella review level.

*Table S1. Specific outcome measures assigned to each health domain*

| *Domain* | *Outcome* |
|---|---|
| ***Auditory*** | Auditory processing |
| | Auditory working memory |
| | Hearing threshold |
| | Melodic contour identification |
| | Pitch discrimination |
| | Speech in noise |
| | Vocal emotion identification |
| ***Autonomic Tone*** | Adrenaline |
| | Noradrenaline |
| ***Blood pressure*** | Blood pressure |
| ***Body Composition*** | % body fat |
| | Body composition (BMI, body composition) |
| | Body mass |
| | Skinfold measurements |
| | Total fat mass |
| | Waist circumference |
| ***Bone Health*** | Bone mineral density |
| ***Cognitive*** | Attention |
| | BDNF |
| | Brain structure/plasticity |
| | Cognitive function/Global cognition |
| | Confusion |
| | Executive function |
| | Fluid intelligence |
| | IQ |
| | IQ (nonverbal) |
| | Memory (long-/short-term, working) |
| | Mental status (Blessed Information Memory Concentration test) |
| | Mental flexibility |
| | Perceptual speed |
| | Spatial ability/reasoning |
| | Vocabulary |
| ***Developmental*** | Foot anthropometrics |
| | Growth |
| | Musculoskeletal development |
| | Puberty onset |
| ***Educational*** | Language |
| | Mathematics |
| | Phonological awareness |
| | Reading |
| | Writing |
| ***Endothelial function*** | Cortisol |

| | |
|---|---|
| | Cortisol:DHEA ratio |
| | DHEA |
| ***Glucose/Insulin*** | Glucose |
| | Insulin |
| ***Immune function*** | C-Reactive Protein |
| | IFN-alpha |
| | IL-2 |
| | IL-6 |
| | Immunological profile |
| | Immunoglobulin A |
| | LAK cell activity |
| | NK cell activity |
| | TNF-alpha |
| | White blood cell count |
| ***Lipid lipoprotein profile*** | HbA1c |
| | HDL cholesterol |
| | Total cholesterol |
| | Triglycerides |
| ***Mental health*** | Anxiety |
| | Burnout |
| | Compassion fatigue |
| | Depersonalization |
| | Depression |
| | Emotional exhaustion |
| | Emotional regulation |
| | Mood |
| | Psychological wellbeing/distress |
| | Self-actualization |
| | Self-esteem |
| | Self-perception |
| ***Non-communicable disease risk*** | All-cause mortality |
| | Asthma |
| | Cardiovascular disease |
| | Dementia |
| | Diabetes |
| | Hypertension |
| | Metabolic syndrome |
| | Osteoarthritis |
| | Rheumatoid arthritis |
| | Tinnitus |
| ***Physical fitness*** | Abdominal strength/endurance (sit ups) |
| | Cardiovascular capacity (VO2 max) |
| | Cardiovascular/cardiorespiratory fitness (Physical Working Capacity 130, Senior fitness test, VO2 peak) |

| | |
|---|---|
| | Endurance (6-minute walk test) |
| | Endurance (muscular - knee extensions) |
| | Endurance (1-mile walk/run) |
| | Heart rate variability |
| | Power (muscular/aerobic) |
| | Peak ventilation |
| | Respiratory exchange ratio |
| | Resting HR |
| | Strength |
| | VO2 at anaerobic threshold |
| ***Physical function*** | Agility |
| | Balance |
| | Falls |
| | Fine motor performance |
| | Flexibility/range of motion |
| | Gait (Gaitrite) |
| | Gait speed |
| | Lung function |
| | Mobility (timed up & go; sit to stand) |
| | Mobility (Step test) |
| | Oestrogen |
| | Proprioception |
| | Reaction time |
| | Sit and Reach (flexibility) |
| | Tactile performance |
| ***Self-reported health/wellbeing*** | Alcohol Consumption |
| | Balance confidence |
| | Body satisfaction (Cathexis) |
| | Daytime somnolence |
| | Drowsiness |
| | Fatigue |
| | Fear of falling |
| | Functional autonomy |
| | Health responsibility |
| | Health system utilization |
| | Lack of coordination |
| | Life satisfaction/Quality of Life/Wellbeing |
| | Loneliness |
| | Medication usage |
| | Morale |
| | Negative affect |
| | Nutrition |
| | Overall/general health |

| | |
|---|---|
| | Perceived self-efficacy |
| | Personal accomplishment |
| | Positive affect |
| | Quality of life |
| | Sexual activity |
| | Sleep quality |
| | Smoking |
| | Somatic symptoms |
| | Spiritual growth/wellbeing |
| | Stress |
| | Stress management |
| | Vigor |
| | Wellbeing |
| ***Social functioning*** | Aggression |
| | Anger |
| | Job engagement |
| | Social functioning |
| | Social activity level |
| | Social anxiety |
| | Social Support (perceived) |

*Table S2. Evidence Synthesis – All GRADE Certainty of Evidence Ratings*

| *Domain* | *Outcome* | *GRADE* | *Effect* | *# reviews* | *# studies/ outcomes* | *Sex* | *Age group* | *Music/ Dance?* | *Style/instrument* | *Participation length* |
|---|---|---|---|---|---|---|---|---|---|---|
| ***Auditory*** | Auditory processing | Moderate | Positive | 1 | 13 | Mixed | Adults | Music | Instrumental | Sustained |
| | Auditory working memory | Very Low | Mixed (positive, no effect) | 1 | 2 | Mixed | Adults | Music | Piano & violin, unspecified | Sustained |
| | Hearing threshold | Very Low | Negative | 1 | 1 | Mixed | Adults, Older adults | Music | Singing | Sustained |
| | Melodic contour identification | Low | Positive | 1 | 1 | Mixed | Adults | Music | Instrumental | Sustained |
| | Pitch discrimination | Moderate | Positive | 1 | 7 | Mixed | Adults | Music | Instrumental, unspecified | Sustained |
| | Speech in noise | Moderate | Positive | 1 | 21 | Mixed | Children, Adolescents, Adults | Music | Instrumental, vocal, unspecified | Sustained |
| | Vocal emotion identification | Low | Positive | 1 | 1 | Mixed | Adults | Music | Instrumental | Sustained |
| ***Autonomic Tone*** | Adrenaline | Very low | No effect | 1 | 1 | Mixed | Adults, Older Adults | Music | Drums | Acute |
| | Noradrenaline | Very low | Mixed (positive, no effect) | 1 | 1 | Mixed | Adults, Older adults | Music | Drums | Acute |
| ***Blood pressure*** | Blood pressure | Very Low | Mixed (positive, no effect) | 3 | 8 | Mixed | Adolescents, Adults | Dance | Ballroom, Dance Team, Zumba | Sustained |
| | Blood pressure | Very Low | Mixed (positive, no effect) | 2 | 2 | Mixed | Adults | Music | Drums, Singing | Acute |
| ***Body Composition*** | % body fat | Very Low | Mixed (positive, no effect) | 2 | 6 | Female | Adolescents, Adults | Dance | Dance team, Zumba | Sustained |
| | Body composition (BMI, body composition) | Very Low | Mixed (positive, no effect) | 5 | 18 | Mixed | Children, adolescents, adults, older adults | Dance | Aerobic dance, African Dance, Ballet, Ballroom, Bhangra, Greek traditional, Hip hop, Zumba | Sustained |
| | Body mass | Very low | Mixed (positive, no effect) | 3 | 16 | Mixed | Children, Adolescents, Adults, Older Adults | Dance | Aerobic dance, Ballroom, Dance team, Greek folk/traditional dance, Zumba | Sustained |
| | Skinfold measurements | Moderate | Positive | 1 | 3 | Female, Unspecified | Children, Adolescents, Adults | Dance | Aerobic dance | Sustained |
| | Total fat mass | Moderate | Positive | 1 | 4 | Female | Children, Adolescents, Adults | Dance | Aerobic dance, Zumba | Sustained |

| | | | | | | | | | | |
|---|---|---|---|---|---|---|---|---|---|---|
| | Waist circumference | Very Low | Mixed (positive, no effect) | 3 | 6 | Mixed | Children, Adults | Dance | Aerobic dance, African dance, Ballroom, Greek folk/traditional dance, Hip-hop, Zumba | Sustained |
| ***Bone Health*** | Bone mineral density | Very Low | Mixed (positive, no effect) | 3 | 6 | Female | Adults, Older Adults | Dance | Aerobic dance, Folk, Waltz, Zumba | Sustained |
| | Bone mineral density | Low | Positive | 1 | 2 | Female | Children, Adolescents | Dance | Ballet | Sustained |
| ***Cognitive*** | Attention (selective, visual) | Low | No effect | 1 | 3 | Mixed | Adults | Music | Instrumental, Unspecified | Sustained |
| | Attention/'Mental status' | Moderate | Positive | 1 | 2 | Mixed | Older adults | Dance | Agilando, Multiple styles – line/jazz/rock'n'roll/square | Sustained |
| | BDNF | Moderate | Positive | 1 | 1 | Mixed | Older adults | Dance | Multiple styles – line/jazz/rock'n'roll/square | Sustained |
| | Brain structure/plasticity | Moderate | Positive | 1 | 3 | Mixed | Adults, Older adults | Dance | Multiple styles – line/jazz/rock'n'roll/square | Sustained |
| | Cognitive function/Global cognition | High | Positive | 2 | 10 | Mixed | Older adults | Dance | Agilando, Ballroom, Jazz, Latin, Tango, Square dance | Sustained |
| | Confusion | Very Low | Mixed (positive, no effect) | 2 | 2 | Mixed | Adults, Older Adults | Music | Drums | Acute |
| | Executive function | Low | No effect | 1 | 1 | Unspecified | Adults | Music | Instrumental | Unclear |
| | Executive function | High | No effect | 3 | 11 | Mixed | Older adults | Dance | Ballroom, Contemporary, Folk, Latin, Social, Tango, Waltz | Sustained |
| | Fluid intelligence | Low | No effect | 1 | 2 | Mixed | Adults, Older adults | Dance | Agilando, unspecified | Sustained |
| | IQ | Moderate | Positive | 2 | 5 | Mixed | Children, Adolescents, Adults | Music | Instrumental, Music education | Sustained |
| | IQ (nonverbal) | Low | No effect | 2 | 9 | Mixed | Children, Adolescents, Adults | Music | Instrumental, Music education | Sustained |
| | IQ (nonverbal) | Low | No effect | 1 | 1 | Mixed | Older adults | Dance | Agilando | Sustained |
| | Memory | Very Low | Mixed (positive, no effect | 3 | 6 | Mixed | Older adults | Dance | Ballroom, Social, Waltz | Sustained |
| | Memory (long-/short-term, working) | Moderate | Positive | 1 | 42 | Mixed | Adults | Music | Instrumental | Sustained |
| | Mental flexibility | Low | Positive | 1 | 1 | Unspecified | Adults | Music | Instrumental | Sustained |
| | Mental status (Blessed Information | Low | No effect | 1 | 1 | Mixed | Older adults | Dance | Social | Sustained |

| | | | | | | | | | | |
|---|---|---|---|---|---|---|---|---|---|---|
| | Memory Concentration test) | | | | | | | | | |
| | Perceptual speed | High | No effect | 1 | 2 | Mixed | Adults, Older adults | Dance | Social, unspecified | Sustained |
| | Spatial ability/reasoning | High | Positive | 1 | 23 | Mixed | Children, Adolescents | Music | Music education (general, Kodaly, Kindermusik, snare drum, piano, vocal) | Sustained |
| | Vocabulary | High | No effect | 1 | 1 | Mixed | Older adults | Dance | Social | Sustained |
| ***Developmental*** | Foot anthropometrics | Low | No effect | 1 | 1 | Female | Adolescents | Dance | Ballet | Sustained |
| | Growth | Very Low | Mixed | 1 | 1 | Female | Adolescents | Dance | Ballet | Sustained |
| | Musculoskeletal development | Low | No effect | 1 | 1 | Female | Children, Adolescents | Dance | Ballet | Sustained |
| | Puberty onset | Very Low | Negative | 1 | 1 | Female | Children | Dance | Ballet | Sustained |
| ***Educational*** | Language | Very Low | Mixed (negative, no effect) | 2 | 2 | Mixed | Children, Adolescents | Music | Music education, instrumental | Sustained |
| | Mathematics | Very Low | Mixed (positive, no effect) | 2 | 26 | Mixed | Children, Adolescents | Music | Music education (general, Kodaly, Suzuki, piano) | Sustained |
| | Phonological awareness | Very Low | Positive | 2 | 18 | Mixed | Children | Music | Music education (general, rhythm intervention, music & movement) | Sustained |
| | Reading | Very Low | Mixed (positive, no effect) | 2 | 37 | Mixed | Children, Adolescents | Music | Orchestra, music education (general, instrumental, vocal, Suzuki) | Sustained |
| | Writing | Very Low | Mixed (positive, no effect) | 1 | 2 | Mixed | Children, Adolescents | Music | Music education (general) | Sustained |
| ***Endothelial function / Stress response*** | Cortisol | Very low | Mixed (positive, no effect) | 4 | 7 | Mixed | Adolescents, Adults | Music | Singing, music education, drums | Acute |
| | Cortisol:DHEA ratio | Low | No effect | 2 | 2 | Mixed | Adults | Music | Singing, Drums | Acute |
| | DHEA | Low | No effect | 2 | 2 | Mixed | Adults | Music | Singing, Drums | Acute |
| ***Glucose/Insulin*** | Glucose | High | No effect | 3 | 6 | Mixed | Adults | Dance | Aerobic Dance, Ballroom, Bhangra, Zumba | Sustained |
| | Insulin | High | No effect | 2 | 2 | Female | Adults | Dance | Bhangra, Zumba | Sustained |
| ***Inflammation / Immune function*** | CRP | Low | No effect | 1 | 1 | Female | Adults | Dance | Zumba | Sustained |
| | IFN-alpha | Very Low | Mixed (positive, no effect) | 1 | 2 | Mixed | Adults, Older adults | Music | Drums | Acute |
| | IL-2 | Low | No effect | 1 | 2 | Mixed | Adults, Older adults | Music | Drums | Acute |

| | | | | | | | | | | |
|---|---|---|---|---|---|---|---|---|---|---|
| | IL-6 | Low | Unclear | 1 | 1 | Female | Adults | Dance | Zumba | Sustained |
| | Immunoglobulin A | High | Positive | 3 | 4 | Mixed | Adults, Older adults | Music | Singing, Drums | Acute |
| | Immunological profile | High | Positive | 2 | 2 | Mixed | Adults | Music | Singing, Drums | Acute |
| | LAK cell activity | Low | Positive | 1 | 1 | Mixed | Adults | Music | Drums | Acute |
| | NK cell activity | Very Low | Mixed (positive, no effect | 2 | 3 | Mixed | Adults, Older adults | Music | Drums | Acute |
| | TNF-alpha | Very Low | Mixed (positive, no effect) | 1 | 1 | Mixed | Adults | Music | Singing | Acute |
| | White blood cell count | Low | No effect | 1 | 2 | Mixed | Adults, Older adults | Music | Drums | Acute |
| | White blood cell count | Low | Negative | 1 | 1 | Female | Adults | Dance | Zumba | Sustained |
| ***Lipid lipoprotein profile*** | HbA1c | Very Low | No effect | 1 | 1 | Female | Adults | Dance | Zumba | Sustained |
| | HDL cholesterol | Very Low | Mixed (positive, no effect | 2 | 6 | Mixed | Adults | Dance | Aerobic dance, Ballroom, Waltz, Zumba | Sustained |
| | Total cholesterol | Very Low | Mixed (positive, no effect) | 3 | 7 | Mixed | Adults | Dance | Aerobic dance, Ballroom, Waltz, Zumba | Sustained |
| | Triglycerides | Very Low | Mixed (positive, no effect) | 3 | 7 | Mixed | Adults | Dance | Aerobic dance, Ballroom, Waltz, Zumba | Sustained |
| ***Mental health*** | Anxiety | Very Low | Mixed (positive, no effect) | 6 | 9 | Mixed | Adults, older adults | Music | Drums, Singing | Acute/Sustained |
| | Anxiety | Low | Positive | 2 | 2 | Mixed | Adults, Older Adults | Dance | Greek traditional dance, Modern | Acute/Sustained |
| | Burnout | Low | Positive | 1 | 1 | Mixed | Adults | Music | Drums | Sustained |
| | Compassion fatigue | Very Low | No effect | 1 | 1 | Mixed | Adults | Music | Instrumental | Sustained |
| | Depersonalization | Low | No effect | 1 | 1 | Mixed | Adults | Music | Drums | Sustained |
| | Depression | Very Low | Mixed (positive, no effect) | 5 | 9 | Mixed | Adolescents, Adults, Older Adults | Music | Drums, keyboard, singing | Acute/Sustained |
| | Depression | High | No effect | 1 | 1 | Mixed | Older adults | Dance | Turkish folk dance, Jazz, Social | Sustained |
| | Emotional exhaustion | Low | Positive | 1 | 1 | Mixed | Adults | Music | Drums | Sustained |
| | Emotional regulation | Low | Positive | 1 | 14 | Mixed | Adults | Music | Singing, piano, guitar | Acute |

| | | | | | | | | | | |
|---|---|---|---|---|---|---|---|---|---|---|
| | Mood | Very Low | Mixed (positive, no effect) | 6 | 10 | Mixed | Adults, Older Adults | Music | Drums, Singing | Acute/Sustained |
| | Mood | Moderate | Positive | 1 | 1 | Mixed | Adults | Dance | Hip hop | Acute |
| | Psychological wellbeing/ distress | Low | Positive | 1 | 1 | Mixed | Older adults | Dance | Greek traditional dance | Acute |
| | Self-actualization | Very Low | Unclear | 1 | 1 | Mixed | Older adults | Music | Singing | Unspecified |
| | Self-esteem | Very Low | Mixed (positive, no effect) | 2 | 2 | Mixed | Adolescents | Music | Guitar, Music Education (general) | Sustained |
| | Self-esteem | Low | No effect | 1 | 2 | Female | Children, Adolescents | Dance | Dance team | Sustained |
| | Self-perception | Moderate | Positive | 1 | 2 | Female | Adolescents | Dance | Aerobic dance | Sustained |
| ***Non-communicable disease risk*** | All-cause mortality | Very Low | Mixed (positive, no effect) | N/A | 2 | Mixed | Adults, older adults | Music | Instrumental, singing | Sustained |
| | Asthma | Very Low | No effect | N/A | 1 | Mixed | Older adults | Music | Instrumental | Sustained |
| | Cardiovascular disease | Very Low | Mixed (positive, no effect) | N/A | 1 | Mixed | Adults, Older adults | Dance | Unspecified | Sustained |
| | Dementia | Low | Positive | N/A | 2 | Mixed | Older adults | Music | Instrumental | Sustained |
| | Dementia | Very Low | No effect | N/A | 1 | Mixed | Older adults | Music | Instrumental | Sustained |
| | Dementia | Low | Positive | N/A | 1 | Mixed | Older adults | Dance | Unspecified | Sustained |
| | Diabetes | Very Low | No effect | N/A | 1 | Mixed | Older adults | Music | Instrumental | Sustained |
| | Hypertension | Very Low | No effect | N/A | 1 | Mixed | Older adults | Music | Instrumental | Sustained |
| | Metabolic syndrome | Very Low | Positive | N/A | 2 | Mixed | Adults, Older adults | Dance | Unspecified, International standard | Sustained |
| | Osteoarthritis | Very Low | No effect | N/A | 1 | Mixed | Older adults | Music | Instrumental | Sustained |
| | Rheumatoid arthritis | Very Low | No effect | N/A | 1 | Mixed | Older adults | Music | Instrumental | Sustained |
| | Tinnitus | Very Low | Negative | N/A | 1 | Mixed | Older adults | Music | Instrumental | Sustained |
| ***Physical fitness*** | Abdominal strength/endurance (sit ups) | Moderate | Positive | 2 | 5 | Female/ Unspecified | Children, Adolescents, Adults | Dance | Aerobic dance | Sustained |
| | Cardiovascular capacity (VO2 max) | High | Positive | 3 | 12 | Mixed | Adolescents, Adults, Older Adults | Dance | Aerobic dance, Balinese, Dance Team, Greek folk/traditional dance, Waltz, Zumba | Sustained |
| | Cardiovascular/ cardiorespiratory fitness (Physical Working Capacity | Very Low | Mixed (positive, no effect) | 2 | 5 | Mixed | Adults, Older adults | Dance | Agilando, Bhangra, Creative, Social, multiple styles-line/jazz/rock‘n‘roll/ square | Sustained |

| | | | | | | | | | |
|---|---|---|---|---|---|---|---|---|---|
| | 130, Senior fitness test, VO2 peak) | | | | | | | | |
| | Endurance (1-mile walk/run) | Very Low | Mixed (positive, no effect) | 2 | 4 | Unspecified | Children, Adolescents, Adults | Dance | Aerobic dance, African dance, Hip-hop | Sustained |
| | Endurance (6MWT) | High | Positive | 3 | 6 | Mixed | Adults, Older adults | Dance | Aerobic Dance, Ballroom, Thai, Turkish folk, Zumba | Sustained |
| | Endurance (muscular/isokinetic) | Low | No effect | 1 | 1 | Female | Adults | Dance | Ballet | Sustained |
| | Heart rate variability | Very Low | Mixed | 1 | 1 | Mixed | Adults | Music | Singing | Acute |
| | Peak ventilation | High | Positive | 1 | 4 | Mixed | Adults, Older adults | Dance | Aerobic dance, Greek folk/traditional dance, Zumba | Sustained |
| | Power (aerobic) | Low | Positive | 1 | 1 | Female | Children, Adolescents | Dance | Aerobic dance | Sustained |
| | Power (muscular/aerobic) | High | No effect | 2 | 2 | Mixed | Adults, Older adults | Dance | Ballet, Salsa | Sustained |
| | Respiratory exchange ratio | Moderate | No effect | 1 | 2 | Mixed | Adults | Dance | Aerobic dance, Zumba | Sustained |
| | Resting HR | Very Low | No effect | 1 | 1 | Female | Adults | Dance | Zumba | Sustained |
| | Strength | Moderate | Positive | 3 | 8 | Mixed | Children, Adolescents, Adults, Older adults | Dance | Aerobic Dance, Dance Team, Social | Sustained |
| | Strength | Low | No effect | 1 | 1 | Female | Adults | Dance | Ballet | Sustained |
| | VO2 at anaerobic threshold | Low | Positive | 1 | 1 | Female | Adults | Dance | Aerobic dance | Sustained |
| ***Physical function*** | Agility | Low | Positive | 1 | 1 | Female | Children, Adolescents | Dance | Aerobic dance | Sustained |
| | Balance | High | Positive | 6 | 47 | Mixed | Adolescents, Adults, Older adults | Dance | Aerobic Dance, Agilando, Ballet, Ballroom, Caribbean, Contemporary, Greek traditional, Latin, Lebed Method, Line dance, Modern, Multiple-line/jazz/rock‘n‘roll/square, Opera, Salsa, Thai, Turkish folk, Unspecified, Zumba | Sustained |
| | Balance | Very Low | Mixed (positive, no effect | 2 | 4 | Mixed | Children, Adolescents | Dance | Ballet, Modern | Sustained |
| | Falls | Very Low | Mixed (positive, no effect | 1 | 3 | Mixed | Adults, Older adults | Dance | Ballroom, Folk, Low impact aerobic dance | Sustained |

| | | | | | | | | | | |
|---|---|---|---|---|---|---|---|---|---|---|
| | Falls incidence | Very Low | Mixed (positive, no effect) | 2 | 2 | Mixed | Older adults | Music | Singing | Sustained |
| | Fine motor performance | Low | Positive | 2 | 3 | Mixed | Older adults | Dance | Agilando, Ballroom, unspecified | Sustained |
| | Flexibility/range of motion | High | Positive | 6 | 19 | Mixed | Children, Adolescents, Adults, Older Adults | Dance | Aerobic dance, Ballroom, Ballet, Folk/traditional dance, Social, Thai, Zumba | Sustained |
| | Gait (GaitRite) | Low | Positive | 1 | 2 | Mixed | Older adults | Dance | Social | Sustained |
| | Gait speed | Low | No effect | 1 | 5 | Mixed | Older adults | Dance | Line dance, Salsa, multiple-line/jazz/ square, Argentine, Folk dance | Sustained |
| | Lung function | Low | No effect | 1 | 1 | Mixed | Adults | Music | Singing | Sustained |
| | Mobility (Step test) | Very Low | Mixed | 1 | 2 | Female | Children, Adolescents, Adults | Dance | Aerobic dance | Sustained |
| | Mobility (timed up & go; sit to stand) | Moderate | Positive | 2 | 12 | Mixed | Older adults | Dance | Aerobic dance, Argentine Tango, Ballroom, Folk, Lebed method, Turkish, Thai | Sustained |
| | Oestrogen | Low | No effect | 1 | 1 | Female | Adults | Dance | Aerobic | Sustained |
| | Proprioception | High | Positive | 1 | 1 | Mixed | Older adults | Dance | Creative Dance | Sustained |
| | Reaction time | Low | Positive | 2 | 4 | Mixed | Older adults | Dance | Agilando, Ballroom, Social, Unspecified | Sustained |
| | Flexibility (Sit and reach) | Very Low | No effect | 1 | 1 | Female | Adolescents | Dance | Dance team | Sustained |
| | Tactile performance | Low | Positive | 2 | 3 | Mixed | Older adults | Dance | Agilando, Ballroom, Unspecified | Sustained |
| ***Self-reported health/wellbeing*** | Alcohol Consumption | Moderate | No effect | 1 | 1 | Mixed | Older adults | Dance | Caribbean | Sustained |
| | Balance confidence | High | Positive | 1 | 1 | Mixed | Older adults | Dance | Argentine tango | Sustained |
| | Body satisfaction (Cathexis) | Very Low | No effect | 1 | 1 | Female | Adolescents | Dance | Dance team | Sustained |
| | Daytime somnolence | Low | No effect | 1 | 1 | Mixed | Adults | Music | Singing | Sustained |
| | Drowsiness | Very Low | Positive | 1 | 1 | Mixed | Adults | Music | Drums | Acute |
| | Fatigue | Very low | No effect | 1 | 1 | Mixed | Older adults | Music | Drums | Acute |
| | Fatigue | High | Positive | 2 | 2 | Mixed | Adults | Music | Drums | Acute |
| | Fatigue | Low | Positive | 1 | 1 | Mixed | Older adults | Dance | Greek traditional dance | Acute |
| | Fear of falling | Very Low | Mixed | 1 | 2 | Mixed | Adults, Older adults | Dance | Video game dance, Low impact dance | Sustained |
| | Functional autonomy | High | Positive | 1 | 1 | Unspecified | Older adults | Dance | Ballroom | Sustained |
| | General health | Very Low | Mixed | 1 | 2 | Mixed | Older adults | Dance | Turkish folk, low impact aerobic | Sustained |

| | | | | | | | | | | |
|---|---|---|---|---|---|---|---|---|---|---|
| | Health responsibility | Low | No effect | 1 | 1 | Mixed | Older adults | Music | Instrumental | Sustained |
| | Health system utilization | Very Low | Positive | 2 | 2 | Mixed | Older adults | Music | Singing | Sustained |
| | Lack of coordination | Low | No effect | 1 | 1 | Mixed | Adults | Music | Drums | Acute |
| | Life satisfaction | High | Positive | 1 | 1 | Mixed | Older adults | Dance | Creative Dance | Sustained |
| | Life satisfaction/ Quality of Life/ Wellbeing | Very Low | No effect | 1 | 1 | Mixed | Older adults | Music | Singing | Unspecified |
| | Lifestyle/quality of life | Low | Positive | 1 | 1 | Mixed | Older adults | Dance | Agilando | Sustained |
| | Loneliness | Very Low | Mixed (positive, no effect) | 2 | 2 | Mixed | Older Adults | Music | Singing | Sustained |
| | Medication usage | Very Low | Positive | 2 | 2 | Mixed | Older adults | Music | Singing | Sustained |
| | Morale | Very Low | No effect | 1 | 1 | Mixed | Older adults | Music | Singing | Sustained |
| | Negative affect | Very Low | Mixed (positive, no effect) | 2 | 2 | Mixed | Adults | Music | Singing, Drums | Acute |
| | Negative affect | Low | No effect | 1 | 1 | Female | Adults | Music | Drums | Sustained |
| | Nutrition | Low | No effect | 1 | 1 | Mixed | Older adults | Music | Instrumental | Sustained |
| | Overall health | Very Low | Mixed (positive, no effect) | 2 | 2 | Mixed | Older adults | Music | Singing | Sustained |
| | Perceived self-efficacy | Low | No effect | 1 | 1 | Female | Adults | Music | Drums | Sustained |
| | Personal accomplishment | Low | No effect | 1 | 1 | Mixed | Adults | Music | Drums | Sustained |
| | Positive affect | Low | Positive | 5 | 5 | Mixed | Adults | Music | Drums, Singing | Acute/Sustained |
| | Quality of life | High | Positive | 2 | 2 | Mixed | Adults, Older Adults | Music | Singing | Sustained |
| | Sexual activity | Moderate | Positive | 1 | 1 | Mixed | Older adults | Dance | Caribbean | Sustained |
| | Sleep quality | Moderate | Positive | 1 | 1 | Mixed | Older adults | Dance | Caribbean | Sustained |
| | Smoking | Moderate | No effect | 1 | 1 | Mixed | Older adults | Dance | Caribbean | Sustained |
| | Spiritual growth/wellbeing | Low | Positive | 2 | 2 | Mixed | Adults | Music | Singing | Sustained |
| | Stress | Low | Positive | 2 | 2 | Mixed | Adults | Music | Singing, Drums | Sustained/Acute |
| | Stress | Moderate | Positive | 1 | 1 | Mixed | Older adults | Dance | Social | Sustained |
| | Stress management | Low | No effect | 1 | 1 | Mixed | Older adults | Music | Instrumental | Sustained |

| | | | | | | | | | | |
|---|---|---|---|---|---|---|---|---|---|---|
| | Vigor | Very Low | Mixed (positive, no effect) | 2 | 2 | Mixed | Adults, older adults | Music | Drums | Acute |
| | Wellbeing | Low | No effect | 1 | 2 | Mixed | Adults, Older adults | Music | Singing, instrumental | Acute |
| ***Social functioning*** | Aggression | Very Low | Positive | 1 | 1 | Mixed | Adults | Music | Drums | Acute |
| | Anger | Very Low | Mixed (positive, no effect) | 2 | 2 | Mixed | Adolescents, Adults | Music | Percussion, keyboard, singing | Sustained |
| | Anger | High | Positive | 2 | 2 | Mixed | Adults, Older adults | Music | Drums | Acute |
| | Job engagement | Low | No effect | 1 | 1 | Female | Adults | Music | Drums | Sustained |
| | Social activity level | Very Low | Positive | 2 | 2 | Mixed | Older Adults | Music | Singing | Sustained |
| | Social anxiety | Low | No effect | 1 | 1 | Mixed | Adolescents | Music | Music education (general) | Sustained |
| | Social functioning | Very Low | Mixed (positive, no effect) | 5 | 6 | Mixed | Adolescents, Older Adults | Music | Drums, keyboard, Singing, Instrumental, Music education (general) | Sustained |
| | Social Support (perceived) | Moderate | Positive | 1 | 1 | Mixed | Older adults | Dance | Social | Sustained |
| | Somatic symptoms | Low | No effect | 1 | 1 | Mixed | Older adults | Music | Singing | Sustained |

*Table S3. Synthesized evidence for each outcome measure from each included review of dance participation. Outcome measures grouped by health domain as per Table S1.*

| *Outcome* | *GRADE* | *Effect* | *Author* | *Year* | *Type of evidence synthesis* | *# studies* | *Study design* | *Country of origin* | *Total N* | *Participant demographics* | *Intervention length* | *Frequency/duration of intervention sessions* | *Style/instrument* |
|---|---|---|---|---|---|---|---|---|---|---|---|---|---|
| ***Domain: Auditory*** | | | | | | | | | | | | | |
| ***Domain: Autonomic Tone*** | | | | | | | | | | | | | |
| ***Domain: Blood pressure*** | | | | | | | | | | | | | |
| Blood pressure | Moderate | No effect | Fong Yan et al. | 2018 | Narrative | 2 | RCT, non-randomized experimental study | Italy, Norway | 172 | Hospital employees & Type II diabetic/obese adults, mean age 45-59, mixed sex | 6 months – 40 weeks | 1-2 hours/2x weekly | Ballroom |
| Blood pressure | Very Low | Positive | Burkhardt & Brennan | 2012 | Narrative | 1 | Non-randomized experimental study | USA | 16 | Young females (age range 14-15; 8 on dance team, 8 physical education control) | 5 months | 1 hour/5x weekly | Dance team |
| Blood pressure | Very Low | Mixed (positive & no effect) | Cugusi et al | 2019 | Narrative | 5 | RCT, Single group experimental study | Italy, Switzerland, UK, USA | 118 | Overweight & obese women (mean age 34-54) | 8-16 weeks | 50-60 min/1-3x weekly | Zumba |
| ***Domain: Body Composition*** | | | | | | | | | | | | | |
| % body fat | Very Low | Positive | Burkhardt & Brennan | 2012 | Narrative | 1 | Non-randomized experimental study | USA | 16 | Young females (age range 14-15; 8 on dance team, 8 physical education control | 5 months | 1 hour/5x weekly | Dance team |
| % body fat | Very Low | Mixed (positive, no effect | Cugusi et al | 2019 | Narrative | 5 | RCT, Single group experimental study | Italy, Switzerland, UK, USA | 149 | Healthy & overweight women (mean age 27-54) | 8-16 weeks | 50-60 min/1-3x weekly | Zumba |
| BMI | High | Positive | Fong Yan et al. | 2018 | Meta-analysis | 6 | RCT, non-randomized experimental study | Greece, Italy, Japan, Norway, UK, USA | 341 | Hospital/university employees, schoolgirls, type II diabetic/obese individuals, chronic heart failure (mean age 14-67), mixed sex | 6-40 weeks | 50 min-2 hours/2-3x weekly | Aerobic dance, Ballroom, Greek folk/traditional dance, Zumba |
| BMI | Very Low | Mixed (positive & no effect) | Burkhardt & Brennan | 2012 | Narrative | 1 | RCT | USA | 142 | Children (aged 8-13), mixed sex | 12 weeks | 50 min/3x weekly | Hip-hop, African dance, & aerobics |
| BMI | Very Low | Mixed (positive, no effect | Cugusi et al | 2019 | Narrative | 6 | RCT, Single group experimental study | Italy, Malta, Switzerland, UK, USA | 171 | Healthy & overweight women (mean age 21-54) | 8-16 weeks | 50-60 min/1-3x weekly | Zumba |

| | | | | | | | | | | | | | |
|---|---|---|---|---|---|---|---|---|---|---|---|---|---|
| Body composition | Very Low | Mixed (positive, no effect) | Rodrigues-Krause, Krause, Reischak-Oliveira | 2019 | Narrative | 4 | RCT, cross-sectional | Canada, Hong Kong, Taiwan | 242 | Older adults (mean age = 56.4 – 69.1), mixed sex | 12-16 weeks; at least 3 years experience | 50-60 min/2-3 weekly; at least 150 min weekly | Bhangra, Low impact aerobic dance |
| Body composition | Low | No effect | Letton, Thom, & Ward | 2020 | Narrative | 1 | Non-randomized experimental study | Israel | 30 | 1st-3rd grade girls (age range 6-9) | 6 months | 90 min/2x weekly | Ballet |
| Body Mass | High | Positive | Fong Yan et al. | 2018 | Meta-analysis | 8 | RCT, non-randomized experimental study | Croatia, Greece, Italy, Japan, Norway, Turkey, USA | 541 | University/hospital employees; 3rd grade, 4th grade & university students, Type II diabetic/obese individuals, chronic heart failure (mean age 9-67), mixed sex | 8 weeks – 8 months | 45 min – 2 hours / 2-3x weekly | Aerobic dance, Ballroom, Greek folk/traditional dance, Zumba |
| Body mass | Very Low | No effect | Burkhardt & Brennan | 2012 | Narrative | 1 | Non-randomized experimental study | USA | 16 | Young females (age range 14-15; 8 on dance team, 8 physical education control | 5 months | 1 hour/5x weekly | Dance team |
| Body mass | Very Low | Mixed (positive, no effect | Cugusi et al | 2019 | Narrative | 7 | RCT, Single group experimental study | Italy, Malta, Switzerland, UK, USA | 198 | healthy & overweight women (mean age 27-54) | 8-16 weeks | 50-60 min/1-3x weekly | Zumba |
| Skinfold measurements | Moderate | Positive | Fong Yan et al. | 2018 | Narrative | 3 | RCT, non-randomized experimental study | Greece, India, Japan | 175 | 1st grade & university students, moderately obese women, mean age 7-52, female/sex unspecified | 6-12 weeks | 30-60 min/2-3x weekly | Aerobic dance |
| Total fat mass | Moderate | Positive | Fong Yan et al. | 2018 | Meta-analysis | 4 | RCT, non-randomized experimental study, longitudinal cohort study | Croatia, Japan, Norway, USA | 440 | Female 3rd-4th graders/hospital employees/university students/moderately obese Japanese women (mean age ~9/10 – 52) | 8-40 weeks | 1 hour/2-3x weekly | Aerobic dance, Zumba |
| Waist circumference | Low | Positive | Fong Yan et al. | 2018 | Narrative | 2 | Non-randomized experimental cohort study | Italy, Japan | 160 | Obese/Type II diabetic adults, mean age 49-59, mixed sex | 12 weeks – 6 months | 1-2 hours/2-3x weekly | Aerobic dance, Ballroom, Greek folk/traditional dance, Zumba |
| Waist circumference | Low | No effect | Burkhardt & Brennan | 2012 | Narrative | 1 | RCT | USA | 61 | African American girls, age 8-10 | 12 weeks | 3x weekly | Hip hop, African dance & step aerobics |
| Waist circumference | Very Low | Mixed (positive & no effect) | Cugusi et al | 2019 | Narrative | 3 | Single group experimental study | Italy, Switzerland, USA | 85 | Overweight/obese women (mean age 39-54) | 12-16 weeks | 50-60 min/2-3x weekly | Zumba |
| ***Domain: Bone Health*** | | | | | | | | | | | | | |

| | | | | | | | | | | | | | |
|---|---|---|---|---|---|---|---|---|---|---|---|---|---|
| Bone density (peripheral and spinal) | Very Low | Mixed (positive, no effect) | Keogh et al. | 2009 | Narrative | 1 | Single group experimental study | Austria | 28 | Female members of a senior dancing group, mean age = 66 | 12 months | Unspecified | Folk, waltz, aerobic dance |
| Bone density/composition | Very Low | Mixed (positive, no effect) | Rodrigues-Krause, Krause, Reischak-Oliveira | 2019 | Narrative | 3 | RCT, cross-sectional | Taiwan, USA | 166 | Female older adults (mean age = 56 - 59.1) | 16 weeks - 6 months; at least 3 years | 60 min/2-4x weekly; at least 150 min weekly | Aerobic dance |
| Bone mineral density | Moderate | No effect | Fong Yan et al. | 2018 | Narrative | 2 | RCT, non-randomized experimental study | Norway, UK | 101 | Female hospital employees & postmenopausal women (mean age 45-58) | 10-40 weeks | 1 hour/1-2x weekly | Aerobic dance, Zumba |
| Bone mineral density | Low | Positive | Burkhardt & Brennan | 2012 | Narrative | 2 | Cross-sectional, prospective cohort study | Australia | 173 | Young girls, mean age 8-14 | 3 years; unspecified | Unspecified | Ballet |
| ***Domain: Cognitive*** | | | | | | | | | | | | | |
| Attention | Moderate | Positive | Teixeira-Machado, Arida & de Jesus Mari | 2019 | Narrative | 2 | RCT | Germany | 87 | Healthy older adults, mean age 68-70, mixed sex | 24 weeks - 18 months | 60-90 min/1-2x weekly | Agilando, multiple styles – line/jazz/rock‘n‘roll/square |
| BDNF | Moderate | Positive | Teixeira-Machado, Arida & de Jesus Mari | 2019 | Narrative | 1 | RCT | Germany | 52 | Healthy older adults, mean age 68, mixed sex | 18 months | 90 min/twice weekly (1st 6 months); 90 min/weekly (last 12 months) | Multiple styles – line/jazz/rock‘n‘roll/square |
| Brain structure/plasticity | Moderate | Positive | Teixeira-Machado, Arida & de Jesus Mari | 2019 | Narrative | 3 | RCT, Cross-sectional | Germany, USA | 92 | University students & healthy older adults, mean age 22 – 68, mixed sex | 18 months; mean 12 years experience | 60 - 90 min/1-2x week; mean 14 hours/week | Multiple styles – line/jazz/rock‘n‘roll/square |
| Cognitive performance | Low | Positive | Rodrigues-Krause, Krause, Reischak-Oliveira | 2019 | Narrative | 3 | Cross-sectional, Single group experimental study | Germany, USA | 124 | Older adults (mean age = 68 - ~71), mixed sex | 15 weeks; Mean 16.5 - 22.1 years dance experience | Unspecified; Mean 1.3 - 4.6 hours weekly dancing | Ballroom, jazz, unspecified |
| Executive function | High | No effect | Teixeira-Machado, Arida & de Jesus Mari | 2019 | Narrative | 1 | RCT | USA | 188 | Healthy older adults, mean age 65, mixed sex | 6 months | 3x/week | Social |
| Executive function | Low | No effect | Rodrigues-Krause, Krause, Reischak-Oliveira | 2019 | Narrative | 1 | Cross-sectional | USA | 48 | Older adults (mean age = 80.4), mixed sex | Mean 36.5 years social dance experience | Unspecified | 'social dancing' |
| Executive function | High | No effect | Meng et al. | 2020 | Meta-analysis | 9 | RCT, Non-randomized experimental study | Australia, Brazil, Croatia, France, Greece, Japan, South Korea USA | 1286 | Older adults (age 60+)mixed sex | 10 weeks - 12 months | 45-120 min/1-2x weekly | Ballroom, Contemporary, Folk, Latin, Tango, Waltz |

| | | | | | | | | | | | | |
|---|---|---|---|---|---|---|---|---|---|---|---|---|
| Fluid intelligence | Low | No effect | Teixeira-Machado, Arida & de Jesus Mari | 2019 | Narrative | 2 | RCT, Cross-sectional | Germany, USA | 75 | Healthy older adults & female university students, mean age 22-68, mixed sex | 24 weeks; mean 12 years experience | 1 hour/weekly, mean 14 hours weekly dancing | Agilando, unspecified |
| Global cognition | High | Positive | Meng et al. | 2020 | Meta-analysis | 7 | RCT, non-randomized experimental study | China, Germany, Greece, Japan, South Korea, USA | 677 | Older adults (age 60+), mixed sex | 12-48 weeks | 30-90 min/1-5x weekly | Agilando, Ballroom, Latin, Tango, Square dance |
| Intelligence (nonverbal) | Low | No effect | Teixeira-Machado, Arida & de Jesus Mari | 2019 | Narrative | 1 | RCT | Germany | 35 | Healthy older adults, mean age 70, mixed sex | 24 weeks | 1 hour/weekly | Agilando |
| Memory | Very Low | Mixed | Teixeira-Machado, Arida & de Jesus Mari | 2019 | Narrative | 3 | RCT, cross-sectional | Germany, USA | 280 | Healthy older adults & female university students, mean age 22-68, mixed sex | 6 - 18 months; mean 12 years dance experience | 60-90 min/1-3x weekly; mean 14 hours/week | Social, multiple styles – line/jazz/rock‘n‘roll/square, unspecified |
| Memory | Very Low | Mixed (positive, no effect) | Meng et al. | 2020 | Narrative | 2 | RCT | Australia, Croatia | 139 | Healthy older adults (age 60+), mixed sex | 10 weeks - 8 months | 45-60 min/1-2x weekly | Ballroom, Waltz |
| Memory (episodic) | Low | No effect | Rodrigues-Krause, Krause, Reischak-Oliveira | 2019 | Narrative | 1 | Cross-sectional | USA | 48 | Older adults (mean age = 80.4), mixed sex | Mean 36.5 years social dance experience | Unspecified | Social |
| Mental status | Low | No effect | Rodrigues-Krause, Krause, Reischak-Oliveira | 2019 | Narrative | 1 | Cross-sectional | USA | 48 | Older adults (mean age = 80.4), mixed sex | Mean 36.5 years social dance experience | Unspecified | Social |
| Perceptual speed | High | No effect | Teixeira-Machado, Arida & de Jesus Mari | 2019 | Narrative | 2 | RCT, Cross-sectional | USA | 228 | Healthy older adults & female university students, mean age 22-66, mixed sex | 6 months; mean 12 years experience | 3x/week; mean 14 hours/week | Social, unspecified |
| Vocabulary | High | No effect | Teixeira-Machado, Arida & de Jesus Mari | 2019 | Narrative | 1 | RCT | USA | 188 | Healthy older adults, mean age 65, mixed sex | 6 months | 3x/week | Social |
| ***Domain: Developmental*** | | | | | | | | | | | | | |
| Foot anthropometrics | Low | No effect | Letton, Thom, & Ward | 2020 | Narrative | 1 | Cross-sectional | Turkey | 67 | Female adolescents (mean age = ~11) | Mean 4.4 years ballet experience | Unspecified | Ballet |
| Growth | Very Low | Mixed | Letton, Thom, & Ward | 2020 | Narrative | 1 | Longitudinal cohort study | France | 127 | Young girls (mean age 12.6) | 5 years | Mean 8.8 hours weekly ballet practice | Ballet |
| Musculoskeletal development | Low | No effect | Burkhardt & Brennan | 2012 | Narrative | 1 | Cross-sectional | Israel | 1708 | Young females, age 8-16 | Unspecified | mean 2.4-11.3 hours weekly dance | Ballet |
| Puberty onset | Very Low | Negative | Letton, Thom, & Ward | 2020 | Narrative | 1 | Non-randomized experimental study | Israel | 30 | 1st-3rd grade girls (age range 6-9) | 6 months | 90 min/2x weekly | Ballet |

| | | | | | | | | | | | | |
|---|---|---|---|---|---|---|---|---|---|---|---|---|
| ***Domain: Educational*** | | | | | | | | | | | | |
| ***Domain: Endothelial function*** | | | | | | | | | | | | |
| ***Domain: Glucose/Insulin*** | | | | | | | | | | | | |
| Glucose | High | No effect | Rodrigues-Krause, Krause, Reischak-Oliveira | 2019 | Narrative | 2 | RCT | Canada, Taiwan | 107 | Adult Women (mean age = 56.4 - 59) | 12-16 weeks | 60 min/3x weekly | Bhangra, Low impact aerobic dance |
| Glucose | Moderate | No effect | Fong Yan et al. | 2018 | Narrative | 2 | RCT, non-randomized experimental study | Italy, Norway | 172 | Hospital employees & Type II diabetic/obese adults, mean age 45-59, mixed sex | 6 months – 40 weeks | 1-2 hours/2x weekly | Ballroom dance, Zumba |
| Glucose | Very Low | No effect | Cugusi et al | 2019 | Narrative | 2 | Single group experimental study | USA | 54 | Overweight/obese women (mean age = 48-52) | 12-16 weeks | 60 min/2-3x weekly | Zumba |
| Insulin | High | No effect | Rodrigues-Krause, Krause, Reischak-Oliveira | 2019 | Narrative | 1 | RCT | Canada | 75 | Post-menopausal obese south east Asian women (mean age = 56 - 57) | 12 weeks | 60 min/3x weekly | Bhangra |
| Insulin | Very Low | No effect | Cugusi et al | 2019 | Narrative | 1 | Single group experimental study | USA | 41 | Obese women (mean age = 49.3) | 16 weeks | 60min/3x weekly | Zumba |
| ***Domain: Immune function*** | | | | | | | | | | | | |
| CRP | Low | No effect | Cugusi et al | 2019 | Narrative | 1 | RCT | UK | 20 | Overweight and sedentary women (mean age = 34) | 8 weeks | 60 min/1-2x weekly | Zumba |
| IL-6 | Low | Unclear | Cugusi et al | 2019 | Narrative | 1 | RCT | UK | 20 | Overweight and sedentary women (mean age = 34) | 8 weeks | 60 min/1-2x weekly | Zumba |
| White blood cell count | Low | Negative | Cugusi et al | 2019 | Narrative | 1 | RCT | UK | 20 | Overweight and sedentary women (mean age = 34) | 8 weeks | 60 min/1-2x weekly | Zumba |
| ***Domain: Lipid lipoprotein profile*** | | | | | | | | | | | | |
| Cholesterol | High | Positive | Rodrigues-Krause, Krause, Reischak-Oliveira | 2019 | Narrative | 1 | RCT | Taiwan | 32 | Sedentary females (mean age 59) | 16 weeks | 60 min/3x weekly | Low impact aerobic dance |
| HbA1c | Very Low | No effect | Cugusi et al | 2019 | Narrative | 1 | Single group experimental study | USA | 41 | Obese women (mean age = 49.3) | 16 weeks | 60min/3x weekly | Zumba |
| HDL cholesterol | High | Positive | Fong Yan et al. | 2018 | Meta-analysis | 4 | RCT, non-randomized experimental study | Italy, Norway, Turkey | 290 | Female hospital employees & university students, | 8-40 weeks | 21 min-2 hours/2-3x weekly | Aerobic dance, Ballroom, Waltz, Zumba |

| | | | | | | | | | | | | |
|---|---|---|---|---|---|---|---|---|---|---|---|---|
| | | | | | | | | | Type II diabetic/obese adults, chronic heart failure (mean age 21 - 59.5), mixed sex | | | |
| HDL cholesterol | Very Low | Mixed (no effect, unclear) | Cugusi et al | 2019 | Narrative | 2 | Single group experimental study | USA | 54 | Overweight/obese women (mean age = 49-52) | 12-16 weeks | 60 min/2-3x weekly | Zumba |
| Total cholesterol | High | Positive | Fong Yan et al. | 2018 | Meta-analysis | 4 | RCT, non-randomized experimental study | Italy, Norway, Turkey | 290 | Female hospital employees & university students, Type II diabetic/obese adults, chronic heart failure (mean age 21 - 59.5), mixed sex | 8-40 weeks | 21 min-2 hours/2-3x weekly | Aerobic dance, Ballroom, Waltz, Zumba |
| Total cholesterol | Low | No effect | Cugusi et al | 2019 | Narrative | 2 | RCT, Single group experimental study | UK, USA | 61 | Overweight/obese women (mean age 34-49) | 8-16 weeks | 60 min/1-3x weekly | Zumba |
| Triglycerides | High | Positive | Rodrigues-Krause, Krause, Reischak-Oliveira | 2019 | Narrative | 1 | RCT | Taiwan | 32 | Sedentary females (mean age 59) | 16 weeks | 60 min/3x weekly | Low impact aerobic dance |
| Triglycerides | High | Positive | Fong Yan et al. | 2018 | Meta-analysis | 4 | RCT, non-randomized experimental study | Italy, Norway, Turkey | 290 | Female hospital employees & university students, Type II diabetic/obese adults, chronic heart failure (mean age 21 - 59.5), mixed sex | 8-40 weeks | 21 min-2 hours/2-3x weekly | Aerobic dance, Ballroom, Waltz, Zumba |
| Triglycerides | Very Low | Mixed (positive & no effect) | Cugusi et al | 2019 | Narrative | 2 | Single group experimental study | USA | 54 | Overweight/obese women (mean age = 49-52) | 12-16 weeks | 60 min/2-3x weekly | Zumba |
| ***Domain: Mental health*** | | | | | | | | | | | | | |
| Anxiety | Low | Positive | Rodrigues-Krause, Krause, Reischak-Oliveira | 2019 | Narrative | 1 | RCT | Greece | 111 | Healthy older adults (mean age = 69.8), mixed sex | 60 min | Once | Greek traditional dance |
| Anxiety | Low | Positive | Burkhardt & Brennan | 2012 | Narrative | 1 | Non-randomized experimental study | UK | 84 | University students, mean age 20, mixed sex | 12 weeks | Unspecified | Modern |
| Depression | High | No effect | Rodrigues-Krause, Krause, | 2019 | Narrative | 3 | RCT, Single group experimental | Turkey, USA | 98 | Older adults (mean age = 68 – 80.4), mixed sex | 8-15 weeks; mean 36.5 years | 1 hour/3x weekly; unspecified | Turkish folk dance, Jazz, Social |

| | | | | | | | | | | | | | |
|---|---|---|---|---|---|---|---|---|---|---|---|---|---|
| | | | Reischak-Oliveira | | | | study, cross-sectional | | | | dancing experience | | |
| Mood | Moderate | Positive | Burkhardt & Brennan | 2012 | Narrative | 1 | RCT | South Korea | 277 | High school & undergraduate students, mean age 21, mixed sex | 50 min | Once | Hip hop |
| Psychological distress | Low | Positive | Rodrigues-Krause, Krause, Reischak-Oliveira | 2019 | Narrative | 1 | Non-randomized experimental study | Greece | 111 | Healthy older adults (mean age = 69.8), mixed sex | 60 min | Once | Greek traditional dance |
| Psychological wellbeing | Low | Positive | Rodrigues-Krause, Krause, Reischak-Oliveira | 2019 | Narrative | 1 | Non-randomized experimental study | Greece | 111 | Healthy older adults (mean age = 69.8), mixed sex | 60 min | Once | Greek traditional dance |
| Self-esteem | Low | No effect | Burkhardt & Brennan | 2012 | Narrative | 2 | Non-randomized experimental study, RCT | USA | 77 | Young females, mean age 8-15 | 12 weeks - 5 months | 1 hour/3-5x weekly | Dance team |
| Self-perception | Moderate | Positive | Burkhardt & Brennan | 2012 | Narrative | 2 | Non-randomized experimental study, RCT | UK | 163 | Adolescent girls, mean age 14-16 | 5-6 weeks | 50 - 60 min/1-2x weekly | Aerobic dance |
| ***Domain: Physical fitness*** | | | | | | | | | | | | | |
| 6MWT | High | Positive | Cugusi et al | 2019 | Narrative | 1 | RCT | Switzerland | 30 | Healthy women (mean age = 21) | 8 weeks | 60 min/2x weekly | Zumba |
| Abdominal strength/endurance (sit ups) | Moderate | Positive | Fong Yan et al. | 2018 | Meta-analysis | 4 | RCT, longitudinal cohort study, non-randomized experimental study | Greece, India, USA | 260 | 1st grade, 5th grade & university students, mean age 7-20, female/sex unspecified | 6-12 weeks | 30-60 min/3x weekly | Aerobic dance |
| Cardiovascular fitness (VO2max) | High | Positive | Fong Yan et al. | 2018 | Meta-analysis | 6 | RCT, non-randomized experimental study | Greece, Italy, Japan, Norway, USA | 313 | Hospital/university employees, moderately obese women, female university students, chronic heart failure (mean age 19-67), mixed sex | 7-40 weeks | 21-60 min/2-4x weekly | Aerobic dance, Greek folk/traditional dance, Waltz, Zumba |
| Cardiovascular/ cardiorespiratory fitness (Physical Working Capacity 130, VO2 peak) | High | No effect | Teixeira-Machado, Arida & de Jesus Mari | 2019 | Narrative | 3 | RCT | Germany, USA | 275 | Healthy older adults, mean age 65-70, mixed sex | 24 weeks - 18 months | 60-90 min/1-3x weekly | Social, Agilando, multiple styles-line/jazz/rock‘n‘roll/square |
| Core strength | Low | Positive | Burkhardt & Brennan | 2012 | Narrative | 1 | Non-randomized experimental study | Croatia | 220 | 3rd & 4th grade girls | 60 high school periods | Unspecified | Aerobic dance |

| | | | | | | | | | | | | | |
|---|---|---|---|---|---|---|---|---|---|---|---|---|---|
| Endurance | Low | No effect | Letton, Thom, & Ward | 2020 | Narrative | 1 | Cross-sectional | USA | 27 | Female university students (age range 18-35) | 2+ years ballet experience | Unspecified | Ballet |
| Endurance (1-mile walk/run) | Moderate | Positive | Fong Yan et al. | 2018 | Narrative | 3 | RCT, non-randomized experimental study | Greece, India | 172 | Students (1st grade, 5th grade & university), mean age 6-~20, sex unspecified | | 6-12 weeks | Aerobic dance |
| Endurance (6-min walk test) | Moderate | Positive | Liu, Shen & Tsai | 2020 | Meta-analysis | 4 | RCT | Brazil, Hong Kong, Thailand, Turkey | 206 | Healthy older adults, mean age 65-74, mixed sex | 6 weeks - 3 months | 40-60 min/2-3x weekly | Ballroom, Turkish folk, low impact aerobic, Thai dance |
| Endurance (6-min walk test) | Low | Positive | Fong Yan et al. | 2018 | Narrative | 1 | Non-randomized experimental study | Italy | 100 | Obese/Type II diabetic adults, mean age 59, mixed sex | 6 months | 2 hours/2x weekly | Ballroom dance |
| Fitness (general/aerobic) | High | Positive | Rodrigues-Krause, Krause, Reischak-Oliveira | 2019 | Narrative | 2 | RCT | Canada, Portugal | 132 | Older women (mean age 57.1 - 72.8) | 12-24 weeks | 50-60 min/3x weekly | Bhangra, Creative Dance |
| Muscular power | High | No effect | Rodrigues-Krause, Krause, Reischak-Oliveira | 2019 | Narrative | 1 | RCT | Germany | 28 | Older adults (69-72), mixed sex | 8 weeks | 1 hour/2x weekly | Salsa |
| Peak ventilation | High | Positive | Fong Yan et al. | 2018 | Meta-analysis | 4 | RCT, non-randomized experimental study | Greece, Norway, USA | 165 | University students & employees, hospital employees, chronic heart failure patients (mean age 19-67), mixed sex | 7 weeks-8 months | 30-60 min/2-4x weekly | Aerobic dance, Greek folk/traditional dance, Zumba |
| Power | Low | No effect | Letton, Thom, & Ward | 2020 | Narrative | 1 | Cross-sectional | USA | 27 | Female university students (age range 18-35) | 2+ years ballet experience | Unspecified | Ballet |
| Power | Low | Positive | Burkhardt & Brennan | 2012 | Narrative | 1 | Non-randomized experimental study | Croatia | 220 | 3rd & 4th grade girls | 60 high school periods | Unspecified | Aerobic dance |
| Respiratory exchange ratio | Moderate | No effect | Fong Yan et al. | 2018 | Narrative | 2 | RCT | Norway, USA | 97 | University & hospital employees, mean age 37-45, mixed sex | 8-40 weeks | 50-60 min/2-3x weekly | Aerobic dance, Zumba |
| Resting HR | Very Low | No effect | Cugusi et al | 2019 | Narrative | 1 | Single group experimental study | Switzerland | 17 | Overweight women (median age = 54) | 12 weeks | 60 min/3x weekly | Zumba |
| Strength | Moderate | Positive | Rodrigues-Krause, Krause, Reischak-Oliveira | 2019 | Narrative | 5 | RCT, Cross-sectional | Hong Kong, Taiwan, USA | 311 | Older adults (mean age = 56 – 80.4), mixed sex | 3-6 months; 3+ years experience | 50-60 min/2-3x weekly; 2-5 dances/2-4x weekly; 150+ min weekly | Aerobic dance, social |
| Strength | Low | No effect | Letton, Thom, & Ward | 2020 | Narrative | 1 | Cross-sectional | USA | 27 | Female university students (age range 18-35) | 2+ years ballet experience | Unspecified | Ballet |

| | | | | | | | | | | | | |
|---|---|---|---|---|---|---|---|---|---|---|---|---|
| Strength | Very Low | Positive | Burkhardt & Brennan | 2012 | Narrative | 1 | Non-randomized experimental study | USA | 16 | Young females (age range 14-15; 8 on dance team, 8 physical education control | 5 months | 1 hour/5x weekly | Dance team |
| Strength (upper body - push-ups) | Low | Positive | Fong Yan et al. | 2018 | Narrative | 2 | Non-randomized experimental cohort study | Greece | 97 | 1st – 5th grade students, mean age 7-11, sex unspecified | 8-12 weeks | 45 min/3x weekly | Aerobic dance |
| Timed mile run | Moderate | No effect | Burkhardt & Brennan | 2012 | Narrative | 1 | RCT | USA | 81 | Adolescents, mean age 10-13, sex unspecified | 12 weeks | 50 min/3x weekly | Hip-hop, African dance, & aerobics |
| VO2 at anaerobic threshold | Low | Positive | Fong Yan et al. | 2018 | Narrative | 1 | Non-randomized experimental study | Japan | 60 | Moderately obese Japanese women, mean age = 52 | 12 weeks | 1 hour/2-3x weekly | Aerobic dance |
| VO2 max | Low | Positive | Burkhardt & Brennan | 2012 | Narrative | 2 | Non-randomized experimental study | Indonesia, USA | 76 | Adolescents, age 14-20, mixed sex | 8 weeks - 5 months | 50-60 min/3-5x weekly | Balinese, Dance team |
| VO2 max | High | Positive | Cugusi et al | 2019 | Narrative | 4 | RCT, Single group experimental study | Switzerland, UK, USA | 122 | Healthy & overweight women (mean age 27-54) | 8-16 weeks | 60 min/1-3x weekly | Zumba |
| ***Domain: Physical function*** | | | | | | | | | | | | | |
| Agility | Low | Positive | Burkhardt & Brennan | 2012 | Narrative | 1 | Non-randomized experimental study | Croatia | 220 | 3rd & 4th grade girls | 60 high school periods | Unspecified | Aerobic dance |
| Balance | Low | Positive | Clarke et al. | 2018 | Narrative | 27 | Cross-sectional | Australia, France, Germany, Italy, Spain, Taiwan, Thailand, USA | 1018 | Individuals (dancers & non-dancers) aged 16-35; mixed sex | 5+ years dance training | Variable, unspecified | Ballet, modern, Thai, opera |
| Balance | Moderate | Positive | Teixeira-Machado, Arida & de Jesus Mari | 2019 | Narrative | 3 | RCT, Cross-sectional | Germany, USA | 127 | Older adults & University students, mean age 22 – 70, mixed sex | 24 weeks - 18 months; mean 12 years experience | 60 - 90 min/1-2x week; mean 14 hours/week | Unspecified, Agilando, multiple styles |
| Balance | High | Positive | Liu, Shen & Tsai | 2020 | Narrative | 4 | RCT | Germany, Greece, Turkey, USA | 114 | Healthy older adults, mean age 69-74, mixed sex | 8 - 10 weeks | 1 hour/2-3x weekly | Line dance, Turkish folk, Salsa, Greek traditional |
| Balance | Moderate | Positive | Rodrigues-Krause, Krause, Reischak-Oliveira | 2019 | Narrative | 10 | RCT, cross-sectional, single group experimental study | Brazil, China, Germany, Hong Kong, Italy, USA | 811 | Older adults (mean age = 61.4-80.4), mixed sex | 6 weeks - 8 months; mean 6.1-36.5 years experience | 45-90 min/1-3x weekly; mean 1.3-4.8 hours weekly dancing | Ballroom, Caribbean, Jazz, Low impact aerobic dance, Lebed Method, Social |
| Balance | Low | Positive | Hwang & Braun | 2015 | Narrative | 1 | Non-randomized experimental study | France | 41 | Healthy older adults - mean age = 73-74, mixed sex | 4.4 months (average) | 1 hour/weekly | Contemporary |

| | | | | | | | | | | | | |
|---|---|---|---|---|---|---|---|---|---|---|---|---|
| Balance | Very Low | Mixed (positive, no effect) | Costa, Ferreira & Felicio | 2013 | Narrative | 3 | Cross-sectional | Brazil, France, Taiwan | 51 | Adolescents & young adults (dancers & non-dancers), mean age 14-19, mixed sex | Mean 31-84 months experience | 3+ hours/weekly; unspecified | Ballet, Modern |
| Falls | Very Low | Mixed | Veronese et al. | 2017 | Narrative | 3 | RCT | Australia, Brazil, Taiwan | 621 | Older adults, mean age 59-68, mixed sex | 12 weeks - 12 months | 50-60 min/2-3x weekly | Ballroom, Folk, Low impact aerobic dance |
| Fine motor performance | Low | Positive | Teixeira-Machado, Arida & de Jesus Mari | 2019 | Narrative | 1 | RCT | Germany | 35 | Healthy older adults, mean age 70, mixed sex | 24 weeks | 1 hour/weekly | Agilando |
| Fine motor performance | Low | Positive | Rodrigues-Krause, Krause, Reischak-Oliveira | 2019 | Narrative | 2 | Cross-sectional | Germany | 111 | Older adults (mean age = ~71), mixed sex | Mean 16.5 - 22.1 | Mean 1.3 - 4.6 hours weekly dancing | Ballroom, unspecified |
| Flexibility | Moderate | Positive | Liu, Shen & Tsai | 2020 | Meta-analysis | 4 | RCT | Czech Republic, Hong Kong, Thailand | 239 | Healthy older adults, mean age 65-82, mixed sex | 6 weeks - 3 months | 40-75 min/1-3x weekly | Ballroom, low impact aerobic, Thai dance |
| Flexibility | Moderate | Positive | Rodrigues-Krause, Krause, Reischak-Oliveira | 2019 | Narrative | 3 | RCT, cross-sectional | China, Taiwan | 474 | Older adults (mean age = 59-62), mixed sex | 16 weeks; 3+ years dance experience | 60 min/3x weekly; 150+ min weekly | Low impact aerobic dance, social |
| Flexibility | Low | Positive | Letton, Thom, & Ward | 2020 | Narrative | 2 | Cross-sectional, non-randomized experimental study | Finland, Israel | 65 | Female children & adolescents (age range 6-12) | 6 months; 2+ years ballet experience | 90 min/2x weekly; Mean 7.25 - 11.1 hours weekly training | Ballet |
| Flexibility (sit & reach – each leg separately) | Low | Positive | Fong Yan et al. | 2018 | Narrative | 2 | Non-randomized experimental cohort study | Greece | 97 | 1st – 5th grade students, mean age 7-11, sex unspecified | 8-12 weeks | 45 min/3x weekly | Aerobic dance |
| Flexibility (sit & reach) | High | Positive | Fong Yan et al. | 2018 | Meta-analysis | 5 | RCT, longitudinal cohort study, non-randomized experimental study | Croatia, India, Norway, Thailand, USA | 493 | 3rd & 4th grade & university students, hospital employees, older women (mean age 9/10 - 67), female/unspecified sex | 6-40 weeks | 30-60 min/2-3x weekly | Aerobic dance, Folk/traditional dance, Zumba |
| Gait | Low | Positive | Rodrigues-Krause, Krause, Reischak-Oliveira | 2019 | Narrative | 2 | Cross-sectional | China, USA | 452 | Older adults (mean age 61.4 – 80.4), mixed sex | Mean 6.1 - 36.5 years dance experience | Mean 4.8 weekly dancing sessions; unspecified | Social |
| Gait Speed | Low | No effect | Liu, Shen & Tsai | 2020 | Meta-analysis | 5 | RCT | Australia, Canada, Germany, USA | 641 | Healthy older adults, mean age 67-78, mixed sex | 8 weeks - 12 months | 60-120 min/2x weekly | Line dance, Salsa, multiple-line/jazz/square, Argentine, Folk dance |
| Mobility | Low | Positive | Rodrigues-Krause, Krause, Reischak-Oliveira | 2019 | Narrative | 3 | Cross-sectional, single group experimental study | Germany, USA | 122 | Older adults (mean age = ~71), mixed sex | 6 weeks; mean 16.5-22.1 years dance experience | 45 min/3x weekly; mean 1.3-4.6 hours weekly dance | Ballroom, Lebed Method, Unspecified |

| | | | | | | | | | | | | | |
|---|---|---|---|---|---|---|---|---|---|---|---|---|---|
| Mobility (Step test) | Very Low | Mixed | Fong Yan et al. | 2018 | Narrative | 2 | Non-randomized experimental cohort study, longitudinal cohort study | Croatia, USA | 308 | 3rd & 4th grade & university female students (mean age 9/10-20) | 8 weeks – 60 high school periods | 1 hour/3x weekly | Aerobic dance |
| Mobility (timed up & go; sit-to-stand) | Moderate | Positive | Liu, Shen & Tsai | 2020 | Meta-analysis | 9 | RCT | Australia, Brazil, Canada, Czech Republic, Hong Kong, Spain, Thailand, Turkey | 917 | Healthy older adults, mean age 65-81, mixed sex | 6 weeks - 12 months | 40-75 min/1-3x weekly | Ballroom, Turkish folk, Low impact aerobic, Thai dance, Argentine Tango, Folk, Multiple-Flamenco, Sevillanas, ballet |
| Oestrogen | Low | No effect | Rodrigues-Krause, Krause, Reischak-Oliveira | 2019 | Narrative | 1 | RCT | USA | 96 | Older Caucasian women (mean age = ~56) | 6 months | 2-5 dances, 2-4 days/week | Aerobic |
| Postural stability | High | Positive | Fong Yan et al. | 2018 | Narrative | 2 | RCT | Greece, Norway | 96 | Older adults & female hospital employees, mean age 45-71, mixed sex | 12-40 weeks | 1 hour/2x weekly | Latin, Zumba |
| Posture | Low | No effect | Letton, Thom, & Ward | 2020 | Narrative | 1 | Non-randomized experimental study | Israel | 30 | 1st-3rd grade girls (age range 6-9) | 6 months | 90 min/2x weekly | Ballet |
| Proprioception | High | Positive | Hwang & Braun | 2015 | Narrative | 1 | RCT | Portugal | 37 | Inactive older adult - mean age = 64-65, mixed sex | 12 weeks | 90 min/3x weekly | Creative Dance |
| Range of motion | Low | Positive | Fong Yan et al. | 2018 | Narrative | 3 | Non-randomized experimental cohort study | Croatia, Greece | 317 | 1st – 5th grade students, mean age 7-11, sex female/unspecified | 8-12 weeks | 45 min/3x weekly | Aerobic dance |
| Reaction time | Low | Positive | Teixeira-Machado, Arida & de Jesus Mari | 2019 | Narrative | 1 | RCT | Germany | 35 | Healthy older adults, mean age 70, mixed sex | 24 weeks | 1 hour/weekly | Agilando |
| Reaction time | Low | Positive | Rodrigues-Krause, Krause, Reischak-Oliveira | 2019 | Narrative | 3 | Cross-sectional | China, Germany | 515 | Older adults (mean age = 61 – 77), mixed sex | Mean 6.1 - 22.1 years dancing experience | Mean 1.3 - 48 hours weekly dancing | Ballroom, social, unspecified |
| Sit and reach | Very Low | No effect | Burkhardt & Brennan | 2012 | Narrative | 1 | Non-randomized experimental study | USA | 16 | Young females (age range 14-15; 8 on dance team, 8 physical education control | 5 months | 1 hour/5x weekly | Dance team |
| Tactile performance | Low | Positive | Teixeira-Machado, Arida & de Jesus Mari | 2019 | Narrative | 1 | RCT | Germany | 35 | Healthy older adults, mean age 70, mixed sex | 24 weeks | 1 hour/weekly | Agilando |
| Tactile performance | Low | Positive | Rodrigues-Krause, Krause, | 2019 | Narrative | 2 | Cross-sectional | Germany | 111 | Older adults (mean age = ~71), mixed sex | Mean 16.5 - 22.1 | Mean 1.3 - 4.6 hours weekly dancing | Ballroom, unspecified |

| | | | | | | | | | | | | |
|---|---|---|---|---|---|---|---|---|---|---|---|---|
| | | | Reischak-Oliveira | | | | | | | | | |
| ***Domain: Self-reported health/ wellbeing*** | | | | | | | | | | | | |
| Alcohol Consumption | Moderate | No effect | Rodrigues-Krause, Krause, Reischak-Oliveira | 2019 | Narrative | 1 | RCT | Italy | 40 | Older adults (mean age = 61.3), mixed sex | 3 months | 35-60 min/2x weekly (35 min 1st 2 weeks, then 60 min) | Caribbean |
| Balance confidence | High | Positive | Rodrigues-Krause, Krause, Reischak-Oliveira | 2019 | Narrative | 1 | RCT | Canada | 25 | Older adults (mean age = 74-78), mixed sex | 10 weeks | 2 hours/2x weekly | Argentine tango |
| Body satisfaction (Cathexis) | Very Low | No effect | Burkhardt & Brennan | 2012 | Narrative | 1 | Non-randomized experimental study | USA | 16 | Young females (age range 14-15; 8 on dance team, 8 physical education control | 5 months | 1 hour/5x weekly | Dance team |
| Fatigue | Low | Positive | Rodrigues-Krause, Krause, Reischak-Oliveira | 2019 | Narrative | 1 | Non-randomized experimental study | Greece | 111 | Healthy older adults (mean age = 69.8), mixed sex | 60 min | Once | Greek traditional dance |
| Fear of falling | Very Low | Mixed | Veronese et al. | 2017 | Narrative | 2 | RCT | Switzerland, Taiwan | 63 | Healthy older adults, mean age 59-86, mixed sex | 12-16 weeks | 10-60 min/2-3x weekly | Video game dance, Low impact dance |
| Functional autonomy | High | Positive | Rodrigues-Krause, Krause, Reischak-Oliveira | 2019 | Narrative | 1 | RCT | Brazil | 75 | Sedentary older subjects (mean age = 67-68), sex unspecified | 8 months | 50 min/3x weekly | Ballroom |
| General health | Very Low | Mixed | Liu, Shen & Tsai | 2020 | Narrative | 2 | RCT | Hong Kong, Turkey | 134 | Healthy older adults, mean age 68-74, mixed sex | 8 weeks - 3 months | 50-60 min/2-3x weekly | Turkish folk, low impact aerobic |
| Life satisfaction | High | Positive | Rodrigues-Krause, Krause, Reischak-Oliveira | 2019 | Narrative | 1 | RCT | Portugal | 57 | Older women (mean age 71-73) | 24 weeks | 50 min/3x weekly | Creative Dance |
| Lifestyle/quality of life | Low | Positive | Teixeira-Machado, Arida & de Jesus Mari | 2019 | Narrative | 1 | RCT | Germany | 35 | Healthy older adults, mean age 70, mixed sex | 24 weeks | 1 hour/weekly | Agilando |
| Sexual activity | Moderate | Positive | Rodrigues-Krause, Krause, Reischak-Oliveira | 2019 | Narrative | 1 | RCT | Italy | 40 | Older adults (mean age = 63.1), mixed sex | 3 months | 35-60 min/2x weekly (35 min 1st 2 weeks, then 60 min) | Caribbean |
| Sleep quality | Moderate | Positive | Rodrigues-Krause, Krause, Reischak-Oliveira | 2019 | Narrative | 1 | RCT | Italy | 40 | Older adults (mean age = 63.1), mixed sex | 3 months | 35-60 min/2x weekly (35 min 1st 2 weeks, then 60 min) | Caribbean |

| | | | | | | | | | | | | | |
|---|---|---|---|---|---|---|---|---|---|---|---|---|---|
| Smoking | Moderate | No effect | Rodrigues-Krause, Krause, Reischak-Oliveira | 2019 | Narrative | 1 | RCT | Italy | 40 | Older adults (mean age = 63.1), mixed sex | 3 months | 35-60 min/2x weekly (35 min 1st 2 weeks, then 60 min) | Caribbean |
| Stress | Moderate | Positive | Teixeira-Machado, Arida & de Jesus Mari | 2019 | Narrative | 1 | RCT | USA | 247 | Healthy older adults, mean age 65, mixed sex | 6 months | 3x/week | Social |
| ***Domain: Social functioning*** | | | | | | | | | | | | | |
| Social Support (perceived) | Moderate | Positive | Teixeira-Machado, Arida & de Jesus Mari | 2019 | Narrative | 1 | RCT | USA | 247 | Healthy older adults, mean age 65, mixed sex | 6 months | 3x/week | Social |

*Table S4. Synthesized evidence for each outcome measure from each included review of music participation. Outcome measures grouped by health domain as per Table S1.*

| Outcome | GRADE | Effect | Author | Year | Type of evidence synthesis | # studies | Study design | Country of origin | Total N | Participant demographics | Intervention length | Frequency/duration of intervention sessions | Style/instrument |
|---|---|---|---|---|---|---|---|---|---|---|---|---|---|
| ***Domain: Auditory*** | | | | | | | | | | | | | |
| Auditory processing | Moderate | Positive | Coffey, Mogilever & Zatorre | 2017 | Narrative | 13 | Cross-sectional, nonrandomized longitudinal cohort study | Canada, France, UK, USA | 624 | Adults & adolescents (musicians & nonmusicians); mean age = 15=56, mixed sex | 3 years + | Variable, unspecified | Instrumental |
| Auditory working memory | Very Low | Mixed | Coffey, Mogilever & Zatorre | 2017 | Narrative | 2 | Cross-sectional | USA | 64 | Adults (musicians & nonmusicians), age 18-40, mixed sex | 10+ years musical experience | At least 3 hours/week | Piano & violin, unspecified |
| Hearing threshold | Very Low | Negative | Clift et al | 2010 | Narrative | 1 | Cross-sectional | Austria | 89 | Adults (musicians & nonmusicians), age range 18-61, mixed sex | Mean 24 (male) – 27 (female) years performing experience | Mean weekly singing of 15 (male) – 29 (female) hours | Singing |
| Melodic contour identification | Low | Positive | Coffey, Mogilever & Zatorre | 2017 | Narrative | 1 | Cross-sectional | Netherlands | 50 | Adults (musicians & nonmusicians), mean age = 23, mixed sex | Average 14.6 years musical training | Unspecified ('regular musical training within the last 3 years') | Instrumental |
| Pitch discrimination | Moderate | Positive | Coffey, Mogilever & Zatorre | 2017 | Narrative | 7 | Cross-sectional | Canada, UK, USA | 351 | Adults (musicians & nonmusicians), mean age 23-31, mixed sex | 6+ years music training | 3+ hours/week; unspecified | Instrumental, unspecified |
| Speech in noise | Moderate | Positive | Coffey, Mogilever & Zatorre | 2017 | Narrative | 21 | RCT, cross-sectional | Canada, France, Netherlands, UK, USA | 863 | Adults & children, mean age 8 - 56, mixed sex | 1+ years | 2+ hours/week; unspecified | Instrumental, vocal, unspecified |
| Vocal emotion identification | Low | Positive | Coffey, Mogilever & Zatorre | 2017 | Narrative | 1 | Cross-sectional | Netherlands | 50 | Adults (musicians & nonmusicians), mean age = 23, mixed sex | Average 14.6 years musical training | Unspecified ('regular musical training within the last 3 years') | Instrumental |
| ***Domain: Autonomic Tone*** | | | | | | | | | | | | | |
| Adrenaline | Very low | No effect | Fancourt, Ockelford & Belai | 2014 | Narrative | 1 | Single group study (uncontrolled) | Japan | 54 | Adult & older adult nonmusicians (mean age 27-70, mixed sex) | 1 hour | Once | Drums |
| Noradrenaline | Very low | Positive | Fancourt, Ockelford & Belai | 2014 | Narrative | 1 | Single group study (uncontrolled) | Japan | 27 | Older adult nonmusicians (mean age 70.3), mixed sex | 1 hour | Once | Drums |
| Noradrenaline | Very low | No effect | Fancourt, Ockelford & Belai | 2014 | Narrative | 1 | Single group study (uncontrolled) | Japan | 27 | Younger adult nonmusicians (mean age 27.9), mixed sex | 1 hour | Once | Drums |
| ***Domain: Blood pressure*** | | | | | | | | | | | | | |
| Blood pressure | Very Low | Mixed (positive- | Yap, Kwan & Ang | 2017 | Narrative | 1 | Single group experimental | South Africa | 34 | Young adult novice drummers & middle | 40 minutes | Once | Drums |

| | | | | | | | | | | | | |
|---|---|---|---|---|---|---|---|---|---|---|---|---|
| | | middle aged adults, no effect young adults) | | | | | study (pre-post analysis) | | | aged experienced drummers, mean age 23 - 53, mixed sex | | | |
| Blood pressure | Very Low | No effect | Daykin et al | 2018 | Narrative | 1 | Nonrandomized experimental cohort study | UK | 23 | University singers, mean age 21, mixed sex | 30 minutes | Once | Singing |
| ***Domain: Body Composition*** | | | | | | | | | | | | | |
| ***Domain: Bone Health*** | | | | | | | | | | | | | |
| ***Domain: Cognitive*** | | | | | | | | | | | | | |
| Confusion | Very low | No effect | Fancourt, Ockelford & Belai | 2014 | Narrative | 1 | Single group study (uncontrolled) | Japan | 27 | Older adult nonmusicians (mean age 70.3), mixed sex | 1 hour | Once | Drums |
| Confusion | Very low | Positive | Fancourt, Ockelford & Belai | 2014 | Narrative | 1 | Single group study (uncontrolled) | Japan | 27 | Younger adult nonmusicians (mean age 27.9), mixed sex | 1 hour | Once | Drums |
| Confusion | Very Low | Positive | Yap, Kwan & Ang | 2017 | Narrative | 1 | Nonrandomized experimental study | USA | 50 | University students, mean age 21, mixed sex | 45 min - 1 hour | Once | Drums |
| Executive function | Low | No effect | Coffey, Mogilever & Zatorre | 2017 | Narrative | 1 | Cross-sectional | USA | 34 | Adults (musicians & nonmusicians), mean age = 24, sex unspecified | Unclear | Unspecified | Instrumental |
| Intelligence (nonverbal/logical – Raven's Standard Progressive Matrices) | Low | No effect | Hetland | 2000 | Meta-analysis | 3 | Longitudinal cohort study, Retrospective cross sectional analysis | Hungary, Switzerland, USA | 694 | Children (age 6-15), mixed sex | 7 months – 3 years | 4-6 classes, weekly | Music education (general, Kodaly) |
| IQ | Moderate (1 high quality study showing positive effects; rated down for publication bias) | Positive | Jaschke et al. | 2013 | Narrative | 4 | RCT, cross-sectional & longitudinal cohort study | Hong Kong, Canada, Australia | 399 | Children (age 6-13), mixed sex | 6.5 months - 5 years | 1-3 hours, weekly; multiple studies unspecified | Music education |

| | | | | | | | | | | | | | |
|---|---|---|---|---|---|---|---|---|---|---|---|---|---|
| IQ | Low | Positive | Coffey, Mogilever & Zatorre | 2017 | Narrative | 1 | Cross-sectional | USA | 33 | University students (musicians and nonmusicians), mean age = 21, mixed sex | At least 10 years of musical training, beginning before age 10 | Currently playing at least 5 hours/week | Instrumental |
| IQ (nonverbal) | Low | No effect | Coffey, Mogilever & Zatorre | 2017 | Narrative | 6 | Cross-sectional | UK, USA | 250 | Adults (musicians & nonmusicians), mean age 22-56, mixed sex | 10 + years musical experience | 3+ hours/weekly; unspecified | Instrumental, vocal, unspecified |
| Long-term memory | Moderate | Positive | Talamini et al | 2017 | Meta-analysis | 9 | Cross-sectional | Canada, China, Germany, Hong Kong, UK, USA | 411 | Adult musicians and nonmusicians (mean age 19-28, mixed sex) | 4+ years music training | Largely unspecified | Instrumental |
| Mental flexibility | Low | Positive | Coffey, Mogilever & Zatorre | 2017 | Narrative | 1 | Cross-sectional | UK, USA | 50 | Adults (musicians & nonmusicians) mean age = 22, sex unspecified | average 22.7 years of musical training | practice at least 3x/weekly over the 3 years before testing | Instrumental |
| Selective attention | Low | No effect | Coffey, Mogilever & Zatorre | 2017 | Narrative | 2 | Cross-sectional | UK, USA | 84 | Adults (musicians & nonmusicians), mean age 24-27, sex unspecified | Average 14-22 years musical training | 3+ hours/week; unclear | Instrumental |
| Short-term memory | Moderate | Positive | Talamini et al | 2017 | Meta-analysis | 17 | Cross-sectional | Australia, Brazil, Canada, Denmark, Finland, France, Germany, Israel, Italy, Singapore, Taiwan, UK, Ukraine, USA | 756 | Adult musicians and nonmusicians (mean age 20-32, mixed sex) | 5+ years music training | Variable, unspecified | Instrumental, singing |
| Spatial abilities (recognition, memory, mental rotation, spatial visualization) | Low (‚equivocal results of meta analysis‘) | Positive | Hetland | 2000 | Meta-analysis | 8 | RCT, Non randomized experimental cohort study, Longitudinal cohort study, Retrospective cross sectional analysis | Switzerland, USA | 655 | Children (age 3-15), mixed sex | 4 weeks – 8 months | 10 – 75 min | Music education (general, Kodaly, Kindermusik, snare drum, piano, vocal) |

| | | | | | | | | | | | | |
|---|---|---|---|---|---|---|---|---|---|---|---|---|
| Spatial Reasoning | High | Positive | Hetland | 2000 | Meta-analysis | 15 | RCT, Non randomized experimental cohort study, Longitudinal cohort study, Retrospective cross sectional analysis | Canada, USA | 701 | Children (age 4-9), mixed sex | 7 weeks – 8 months | 10 – 60 minutes, 1-5x/week | Music education (general, Kodaly, piano, voice) |
| Visual attention | Low | No effect | Coffey, Mogilever & Zatorre | 2017 | Narrative | 1 | Cross-sectional | USA | 33 | University students (musicians and nonmusicians), mean age = 21, mixed sex | At least 10 years musical experience beginning before age 11 | At least 3 hours/week | Unspecified |
| Working memory | Moderate | Positive | Talamini et al | 2017 | Meta-analysis | 16 | Cross-sectional | Australia, Canada, Denmark, Finland, France, Germany, Italy, Singapore, Taiwan, UK, USA | 536 | Adult musicians and nonmusicians (mean age 20-50, mixed sex) | 9+ years music training | Variable, unspecified | Instrumental, singing |
| ***Domain: Developmental*** | | | | | | | | | | | | | |
| ***Domain: Educational*** | | | | | | | | | | | | | |
| Language expression (not otherwise specified) | Moderate | Negative | Jaschke et al. | 2013 | Narrative | 1 | RCT | Canada | 117 | Children (aged 8-12), mixed sex | 3 years | 30-45 min, weekly | Music education (general) |
| Language skills | Very low | No effect | Coffey, Mogilever & Zatorre | 2017 | Narrative | 1 | Nonrandomized longitudinal cohort study | USA | 68 | High school students, mean age 15, sex unspecified | 3 years | 2.33-3 hours/week | Instrumental |
| Mathematics | Very low (mixed no effect, positive results from moderate/high quality studies) | Mixed | Jaschke et al. | 2013 | Narrative | 3 | RCT | Australia, Canada | 330 | Children (aged 6-12), mixed sex | 5 months - 3 years | 30 min - 1 hour, weekly; 1 study unspecified | Music education (general) |
| Mathematics (causation - experimental) | Low | Positive | Vaughn | 2000 | Meta-analysis | 5 | Non-randomized longitudinal | Canada, USA | 339 | Students (kindergarten - 6th grade), mixed sex | 14 months - 3 years | 30 minutes+, 1-3/x weekly | Music education (general, Kodaly, Suzuki, piano) |

| | | | | | | | | | | | |
|---|---|---|---|---|---|---|---|---|---|---|---|
| | | | | | | | cohort study, longitudinal cohort study | | | | | |
| Mathematics (correlation with music instruction) | Moderate | Positive | Vaughn | 2000 | Meta-analysis | 18 | Cross-sectional, retrospective cohort study, unclear | USA | 5738142 | Students (3rd grade - college), mixed sex | 8 months - 5 years+ | 70 min+, weekly; unspecified | Music education (general, Kodaly, Suzuki, piano) |
| Phonological awareness | Very Low | Positive | Gordon, Fehd & McCandliss | 2015 | Meta-analysis | 18 | RCT, Non-randomized experimental study, cross-sectional | Brazil, Canada, Germany, Spain, UK, USA | 1230 | Children (mean age 4.5-9.1; mixed sex) | 4-26 weeks | 10-120 min/1-5x weekly | Music education (general, rhythm intervention, music & movement) |
| Reading | Very Low (mixed negative, no effect & positive results from moderate & high quality studies) | Mixed | Jaschke et al. | 2013 | Narrative | 7 | RCT, cross-sectional & longitudinal cohort study | Germany, USA, Canada, Australia | 463 | Children (</= age 13), mixed sex | 20 weeks - 1.27 years; unspecified duration in multiple studies | Minimum 30 min/week; multiple studies unspecified | Music education (general) |
| Reading abilities (causation) | Low | No effect | Butzlaff | 2000 | Meta-analysis | 6 | Cross-sectional, longitudinal cohort study, non-randomized experimental study, unclear (experimental) | UK, USA | 253 | Students (1st - 4th grade), mixed sex | 3-6 months | 30 - 60 min, 2-3x/week | Orchestra, music education (instrumental, vocal, Suzuki) |
| Reading abilities (correlation) | Moderate | Positive | Butzlaff | 2000 | Meta-analysis | 24 | Non-randomized experimental cohort study, cross-sectional, retrospective cohort study, longitudinal cohort study, unclear (correlational) | USA | 5734913 | Students (1st - 12th grade), mixed sex | 4 months - 3 years | 30 - 90 min, 1-2x week; 7-8 hours/weekly | Orchestra, music education (instrumental, vocal, Suzuki) |
| Reading Fluency | Low | No effect | Gordon, Fehd & McCandliss | 2015 | Meta-analysis | 5 | RCT, Non-randomized experimental study | Brazil, Portugal, UK, USA | 434 | Children aged 5-9, mixed sex | 6-24 weeks | 30-75 min/1-3x weekly | Music education (general, rhythm intervention) |

| | | | | | | | | | | | | | |
|---|---|---|---|---|---|---|---|---|---|---|---|---|---|
| Writing | Very Low (mixed no effect, positive results from Low/Very Low quality studies) | Mixed | Jaschke et al. | 2013 | Narrative | 2 | pseudo-RCT; non-randomized | USA, Australia | 134 | Children (age 4-13), mixed sex | 30 weeks - 6.5 months | 30 minutes, twice weekly; unspecified | Music education (general) |
| ***Domain: Endothelial Function / Stress Response*** | | | | | | | | | | | | | |
| Cortisol | Low | No effect | Fancourt, Ockelford & Belai | 2014 | | 3 | RCT, non-randomized longitudinal cohort study, non-randomized intervention study | German, Sweden, USA | 151 | Adolescents & adults with mixed levels of musical experience (mean age 13-30), mixed sex | 1 hour (2 studies); 8 months (1 study) | Once (2 studies); 1 hour/week (1 study) | Singing, music education, drums |
| Cortisol | Very Low | Mixed | Clift et al | 2010 | Narrative | 1 | Single group experimental study (pre-post analysis) | USA | 42 | Professional singers, mean age 46, mixed sex | 1.5-2.5 hours | Once | Singing |
| Cortisol | Very Low | No effect | Gick | 2012 | Narrative | 2 | Cross-sectional, single group cohort study | Sweden, USA | 26 | Professional & amateur singers, mean age 21-40, mixed sex | 45 minutes – one rehearsal | Once; 4-11 data collections | Singing |
| Cortisol/Cortisol:DHEA ratio | Low | No effect | Daykin et al | 2018 | Narrative | 1 | Nonrandomized experimental trial | Germany | 21 | Adults participating in choir for research purposes, median age ~50, mixed sex | 30 minutes | Once | Singing |
| DHEA | Low | No effect | Fancourt, Ockelford & Belai | 2014 | Narrative | 1 | RCT | USA | 60 | Adults (unspecified musical experience; mean age = 30, mixed sex) | 1 hour | Once | Drums |
| DHEA | Low | No effect | Daykin et al | 2018 | Narrative | 1 | Nonrandomized experimental trial | Germany | 21 | Adults participating in choir for research purposes, median age ~50, mixed sex | 30 minutes | Once | Singing |
| DHEA to cortisol ratio | Low | No effect | Fancourt, Ockelford & Belai | 2014 | Narrative | 1 | RCT | USA | 60 | Adults (unspecified musical experience; mean age = 30, mixed sex) | 1 hour | Once | Drums |
| ***Domain: Glucose/Insulin*** | | | | | | | | | | | | | |

| | | | | | | | | | | | | |
|---|---|---|---|---|---|---|---|---|---|---|---|---|
| ***Domain: Immune function*** | | | | | | | | | | | | |
| IFN-alpha | Very low (mixed effects from very low & low quality evidence) | Mixed | Fancourt, Ockelford & Belai | 2014 | Narrative | 2 | RCT, Single group study | Japan, USA | 114 | Adults (varying levels of musical experience), mean age 28-70, mixed sex | 1 hour | Once | Drums |
| IL-2 | Low | No effect | Fancourt, Ockelford & Belai | 2014 | Narrative | 2 | RCT, Single group study | Japan, USA | 114 | Adults (varying levels of musical experience), mean age 28-70, mixed sex | 1 hour | Once | Drums |
| Immune function (B cells; CD4/CD8 ratio; CD8+ T cells; IL-10 production; IL-4 production; Naive T cells; Naive:memory T cell; Neutrophils; NK cell count) | Very low | No effect | Fancourt, Ockelford & Belai | 2014 | Narrative | 1 | Single group study (uncontrolled) | Japan | 54 | Adult & older adult nonmusicians (mean age 27-70, mixed sex) | 1 hour | Once | Drums |
| Immune function (CD4+ T cells; IL-6 production; Lymphocytes; Memory T cells; T cells) | Very low | Positive | Fancourt, Ockelford & Belai | 2014 | Narrative | 1 | Single group study (uncontrolled) | Japan | 27 | Older adult nonmusicians (mean age 70.3), mixed sex | 1 hour | Once | Drums |
| Immune function (CD4+ T cells; IL-6 production; Lymphocytes; Memory T cells; T cells) | Very low | No effect | Fancourt, Ockelford & Belai | 2014 | Narrative | 1 | Single group study (uncontrolled) | Japan | 27 | Younger adult nonmusicians (mean age 27.9), mixed sex | 1 hour | Once | Drums |
| Immune function (IFN-gamma gene expression (mRNA), IL-10 gene expression (mRNA)) | High | Positive | Yap, Kwan & Ang | 2017 | Narrative | 1 | RCT | Japan | 40 | Healthy males, mean age 38 | 60 minutes | Once | Drums |
| Immune function (IL-2 gene expression (mRNA), IL-6 gene expression (mRNA), CD-56+ cells, leukocyte counts) | High | No effect | Yap, Kwan & Ang | 2017 | Narrative | 1 | RCT | Japan | 40 | Healthy males, mean age 38 | 60 minutes | Once | Drums |
| Immunoglobulin A | Very Low | Positive | Clift et al | 2010 | Narrative | 1 | Single group experimental study (pre-post analysis) | USA | 42 | Professional singers, mean age 46, mixed sex | 1.5 (performance) – 2.5 (rehearsal) hours | Once (3 pre-post tests; 2 rehearsal, 1 performance) | Singing |

| | | | | | | | | | | | | |
|---|---|---|---|---|---|---|---|---|---|---|---|---|
| Immunoglobulin A | Very Low | Positive | Gick | 2012 | Narrative | 1 | Single group cohort study | USA | 10 | Conservatory singers, mean age 21, sex unspecified | One rehearsal/ performance | Data collections from 4-11 rehearsals/ performances over a 10 week period | Singing |
| Immunoglobulin A | High | Positive | Fancourt, Ockelford & Belai | 2014 | Narrative | 2 | RCT, non-randomized intervention study | USA, Germany | 64 | University students and amateur singers (age range 18-74), mixed sex | 30 min – 1 hour | Once | Drums, Singing |
| LAK cell activity | Low | Positive | Fancourt, Ockelford & Belai | 2014 | Narrative | 1 | RCT | USA | 60 | Adults (unspecified musical experience; mean age = 30, mixed sex) | 1 hour | Once | Drums |
| NK cell activity | Very low (mixed effects from very low & low quality evidence) | Mixed | Fancourt, Ockelford & Belai | 2014 | Narrative | 2 | RCT, Single group study | Japan, USA | 114 | Adults (varying levels of musical experience), mean age 28-70, mixed sex | 1 hour | Once | Drums |
| NK cell activity | Very Low | Mixed | Yap, Kwan & Ang | 2017 | Narrative | 1 | RCT | Japan | 40 | Healthy males, mean age 38 | 60 minutes | Once | Drums |
| TNF-alpha | Very Low | Mixed | Gick | 2012 | Narrative | 1 | Cross-sectional | Sweden | 16 | Amateur/professiona l singers, mean age 40, mixed sex | 45 min | Once | Singing |
| White blood cell count | Low | No effect | Fancourt, Ockelford & Belai | 2014 | Narrative | 2 | RCT, Single group study | Japan, USA | 114 | Adults (varying levels of musical experience), mean age 28-70, mixed sex | 1 hour | Once | Drums |
| | | | | | | | ***Domain: Lipid lipoprotein profile*** | | | | | | |
| | | | | | | | ***Domain: Mental health*** | | | | | | |
| Anxiety | Very low | Mixed (positive, no effect) | Fancourt, Ockelford & Belai | 2014 | Narrative | 2 | RCT, Single group study | Japan, USA | 114 | Adults (varying levels of musical experience), mean age 28-70, mixed sex | 1 hour | Once | Drums |
| Anxiety | Very Low | Positive | Yap, Kwan & Ang | 2017 | Narrative | 1 | Nonrandomized experimental study | USA | 50 | University students, mean age 21, mixed sex | 45 min - 1 hour | Once | Drums |
| Anxiety | High | Positive | Daykin et al | 2018 | Narrative | 2 | RCT | Germany, UK | 412 | Adults, mean age 25-69, mixed sex | 41 minutes, 14 weeks | Once, 90 minutes (weekly | Drums, singing |
| Anxiety | Low | No effect | Raglio et al. | 2019 | Narrative | 1 | RCT (only comparative, not inactive control group) | South Korea | 20 | Graduate female music therapy students, mean age 28 | 4 weeks | 60 minutes, weekly | Drums |

| | | | | | | | | | | | | | |
|---|---|---|---|---|---|---|---|---|---|---|---|---|---|
| Anxiety | Low | Positive | Clift et al | 2010 | Narrative | 1 | Non randomized experimental cohort study | UK | 61 | Residents of retirement homes (mean age 84), mixed sex | 4 weeks | 1 hour, weekly | Singing |
| Anxiety (state) | Very Low | No effect | Phillips & Becker | 2019 | Narrative | 1 | Single group study (uncontrolled) | USA | 62 | Healthcare workers, mean age 48, mixed sex | 5 weeks | 90 minutes, weekly | Singing |
| Anxiety (trait) | Very Low | Positive | Phillips & Becker | 2019 | Narrative | 1 | Single group study (uncontrolled) | USA | 62 | Healthcare workers, mean age 48, mixed sex | 5 weeks | 90 minutes, weekly | Singing |
| Burnout | Low | Positive | Phillips & Becker | 2019 | Narrative | 1 | RCT | USA | 125 | Healthcare workers, mean age 44, mixed sex | 6 weeks | 1 hour, weekly | Drums |
| Compassion fatigue | Very Low | No effect | Phillips & Becker | 2019 | Narrative | 1 | Nonrandomized experimental study (2 experimental groups) | USA | 17 | Hospice workers, age range 28-60, mixed sex | 6 weeks | 60 minutes, weekly | Instrumental |
| Depersonalization | Low | No effect | Yap, Kwan & Ang | 2017 | Narrative | 1 | Nonrandomized crossover experimental study | USA | 75 | Nursing students, mean age 28, mixed sex | 6 weeks | 1 hour, weekly | Drums |
| Depression | Moderate | Positive | Daykin et al | 2013 | Narrative | 1 | RCT | USA | 52 | at risk' youth (aged 14-18), mixed sex | | 1 hour, weekly | Music (Percussion & keyboard education) |
| Depression | Very low (mixed effects from very low & low quality evidence) | Mixed | Fancourt, Ockelford & Belai | 2014 | Narrative | 2 | RCT, Single group study | Japan, USA | 114 | Adults (varying levels of musical experience), mean age 28-70, mixed sex | 1 hour | Once | Drums |
| Depression | Very Low | Positive | Yap, Kwan & Ang | 2017 | Narrative | 1 | Nonrandomized experimental study | USA | 50 | University students, mean age 21, mixed sex | 45 min - 1 hour | Once | Drums |
| Depression | High (when measured immediately post-intervention; depression measured ~6-months post intervention in | Positive | Daykin et al | 2018 | Narrative | 3 | RCT, Nonrandomized experimental study | Germany, UK, USA | 578 | Adults, mean age 25-69, mixed sex | 41 minutes, 14-30 weeks | Once, Weekly (duration unspecified - 90 minutes) | Singing |

| | | | | | | | | | | | | | |
|---|---|---|---|---|---|---|---|---|---|---|---|---|---|
| | observational study) | | | | | | | | | | | | |
| Depression | Very Low | Mixed | Clift et al | 2010 | Narrative | 2 | Nonrandomized experimental study | UK, USA | 189 | Older adults (over 64), mean age 79-84, mixed sex | 4-30 weeks | Once weekly | Singing |
| Emotional exhaustion | Low | Positive | Yap, Kwan & Ang | 2017 | Narrative | 1 | Nonrandomized crossover experimental study | USA | 75 | Nursing students, mean age 28, mixed sex | 6 weeks | 1 hour, weekly | Drums |
| Emotional regulation | Low | Positive | Moore | 2013 | Narrative | 14 | Cross-sectional | Canada, Germany, Japan, Sweden, USA | 255 | Adults with varying musical experience (non-musicians - professional), mean age 22-38, mixed sex | <2 hours | Once | Singing, piano, guitar |
| Mood | Very low | Positive | Fancourt, Ockelford & Belai | 2014 | Narrative | 1 | Single group study (uncontrolled) | Japan | 54 | Adult & older adult nonmusicians (mean age 27-70, mixed sex) | 1 hour | Once | Drums |
| Mood | High | Positive | Yap, Kwan & Ang | 2017 | Narrative | 2 | RCT, Nonrandomized crossover experimental study | USA, Japan | 115 | Nursing students & healthy males, mean age 21 - 38, mixed sex | 60 minutes - 6 weeks | Once - 1 hour (weekly) | Drums |
| Mood | High | Positive | Daykin et al | 2018 | Narrative | 4 | RCT, nonrandomized experimental study | Germany, UK, USA | 364 | Adults, university students, mean age 21-50, mixed sex | 30-41 minutes; 30 weeks | Once; once/week (duration unspecified) | Singing, drums |
| Mood | Low | Positive | Phillips & Becker | 2019 | Narrative | 1 | RCT | USA | 125 | Healthcare workers, mean age 44, mixed sex | 6 weeks | 1 hour, weekly | Drums |
| Mood | Low | No effect | Raglio et al. | 2019 | Narrative | 1 | RCT (only comparative, not inactive control group) | South Korea | 20 | Graduate female music therapy students, mean age 28 | 4 weeks | 60 minutes, weekly | Drums |
| Mood | Very Low | Mixed | Clift et al | 2010 | Narrative | 1 | RCT | Australia | 107 | Adults (varying levels of musical experience), age range 18-73, mixed sex | 30 minutes | Once | Singing |
| Self-actualization | Very Low | Unclear | Clift et al | 2010 | Narrative | 1 | Cross-sectional | USA | 98 | Retirees, mean age 65, mixed sex | Unspecified (chorus members generally with more robust musical histories) | Unspecified | Singing |

| | | | | | | | | | | | | | |
|---|---|---|---|---|---|---|---|---|---|---|---|---|---|
| Self-esteem | High | Positive | Daykin et al | 2013 | Narrative | 1 | RCT | USA | 45 | Juvenile delinquents / 'at risk' youth (aged 9-19), sex unspecified | 3 months | 1 hour, weekly | Guitar |
| Self-esteem | Low | No effect | Fancourt, Ockelford & Belai | 2014 | Narrative | 1 | Non-randomized longitudinal cohort study | Sweden | 60 | 5th & 6th graders, sex unspecified (likely mixed) | 8 months | 1 hour, weekly | Music education (general) |
| ***Domain: Physical fitness*** | | | | | | | | | | | | | |
| Heart rate variability | Very Low | Mixed | Gick | 2012 | Narrative | 1 | Cross-sectional | Sweden | 16 | Amateur/professional singers, mean age 40, mixed sex | 45 min | Once | Singing |
| ***Domain: Physical function*** | | | | | | | | | | | | | |
| Falls incidence | Very Low | Positive | Daykin et al | 2018 | Narrative | 1 | Nonrandomized experimental study | USA | 166 | Older adults, mean age 74, sex unspecified | 30 weeks | Once per week (duration unspecified) | Singing |
| Falls incidence | Very Low | No effect | Clift et al | 2010 | Narrative | 1 | Nonrandomized experimental study | USA | 128 | Older adults (over 64), mean age 79, mixed sex | 2 periods of 30 weeks over 2 years | Once per week (duration unspecified) | Singing |
| Lung function | Low | No effect | Clift et al | 2010 | Narrative | 1 | Cross-sectional | USA | 113 | Professional musicians, mean age 41-47, mixed sex | Mean years performing range 18.6-27.5 across groups | Unspecified | Singing |
| ***Domain: Self-reported health/wellbeing*** | | | | | | | | | | | | | |
| Daytime somnolence | Low | No effect | Gick | 2012 | Narrative | 1 | Cross-sectional | UK | 107 | Adults (musicians & nonmusicians), mean age 45, mixed sex | Mean 30.8 years singing | Unspecified | Singing |
| Drowsiness | Very Low | Positive | Yap, Kwan & Ang | 2017 | Narrative | 1 | Nonrandomized experimental study | USA | 50 | University students, mean age 21, mixed sex | 45 min - 1 hour | Once | Drums |
| Fatigue | Very low | No effect | Fancourt, Ockelford & Belai | 2014 | Narrative | 1 | Single group study (uncontrolled) | Japan | 27 | Older adult nonmusicians (mean age 70.3), mixed sex | 1 hour | Once | Drums |
| Fatigue | Very low | Positive | Fancourt, Ockelford & Belai | 2014 | Narrative | 1 | Single group study (uncontrolled) | Japan | 27 | Younger adult nonmusicians (mean age 27.9), mixed sex | 1 hour | Once | Drums |
| Fatigue | High | Positive | Daykin et al | 2018 | Narrative | 1 | RCT | Germany | 154 | Adults, mean age 25, mixed sex | 41 minutes | Once | Drums |
| Health responsibility | Low | No effect | Daykin et al | 2018 | Narrative | 1 | Non randomized longitudinal intervention study | UK | 98 | Older adults, mean age 68, mixed sex | 10 weeks | 60-90 mins, weekly | Instrumental |

| | | | | | | | | | | | | | |
|---|---|---|---|---|---|---|---|---|---|---|---|---|---|
| Health system utilization | Very Low | Positive | Daykin et al | 2018 | Narrative | 1 | Nonrandomized experimental study | USA | 166 | Older adults, mean age 74, sex unspecified | 30 weeks | Once per week (duration unspecified) | Singing |
| Health system utilization | Very Low | Positive | Clift et al | 2010 | Narrative | 1 | Nonrandomized experimental study | USA | 128 | Older adults (over 64), mean age 79, mixed sex | 2 periods of 30 weeks over 2 years | Once per week (duration unspecified) | Singing |
| Lack of coordination | Low | No effect | Yap, Kwan & Ang | 2017 | Narrative | 1 | Nonrandomized experimental study | USA | 50 | University students, mean age 21, mixed sex | 45 min - 1 hour | Once | Drums |
| Life satisfaction | Very Low | No effect | Clift et al | 2010 | Narrative | 1 | Cross-sectional | USA | 98 | Retirees, mean age 65, mixed sex | Unspecified (chorus members generally with more robust musical histories) | Unspecified | Singing |
| Loneliness | Low | Positive | Daykin et al | 2018 | Narrative | 1 | Nonrandomized experimental study | USA | 166 | Older adults, mean age 74, sex unspecified | 30 weeks | Once per week (duration unspecified) | Singing |
| Loneliness | Very Low | No effect | Clift et al | 2010 | Narrative | 1 | Nonrandomized experimental study | USA | 128 | Older adults (over 64), mean age 79, mixed sex | 2 periods of 30 weeks over 2 years | Once per week (duration unspecified) | Singing |
| Medication usage | Very Low | Positive | Daykin et al | 2018 | Narrative | 1 | Nonrandomized experimental study | USA | 166 | Older adults, mean age 74, sex unspecified | 30 weeks | Once per week (duration unspecified) | Singing |
| Medication usage (over the counter + prescription) | Very Low | Positive | Clift et al | 2010 | Narrative | 1 | Nonrandomized experimental study | USA | 128 | Older adults (over 64), mean age 79, mixed sex | 2 periods of 30 weeks over 2 years | Once per week (duration unspecified) | Singing |
| Morale | Very Low | No effect | Clift et al | 2010 | Narrative | 1 | Nonrandomized experimental study | USA | 128 | Older adults (over 64), mean age 79, mixed sex | 2 periods of 30 weeks over 2 years | Once per week (duration unspecified) | Singing |
| Negative affect | Low | Positive | Fancourt, Ockelford & Belai | 2014 | Narrative | 1 | Non-randomized intervention study | Germany | 31 | Adult amateur singers (mean age 56.9, range 29-74), mixed sex | 1 hour | Once | Singing |
| Negative affect | Low | No effect | Yap, Kwan & Ang | 2017 | Narrative | 1 | Nonrandomized experimental study | UK | 32 | Adults, mean age 25 - 44, mixed sex | 30 minutes | Once | Drums |
| Negative affect | Low | No effect | Raglio et al. | 2019 | Narrative | 1 | RCT (only comparative, not inactive control group) | South Korea | 20 | Graduate female music therapy students, mean age 28 | 4 weeks | 60 minutes, weekly | Drums |
| Nutrition | Low | No effect | Daykin et al | 2018 | Narrative | 1 | Non randomized longitudinal intervention study | UK | 98 | Older adults, mean age 68, mixed sex | 10 weeks | 60-90 mins, weekly | Instrumental |

| | | | | | | | | | | | | | |
|---|---|---|---|---|---|---|---|---|---|---|---|---|---|
| Overall health | Very Low | Positive | Daykin et al | 2018 | Narrative | 1 | Nonrandomized experimental study | USA | 166 | Older adults, mean age 74, sex unspecified | 30 weeks | Once/week (duration unspecified | Singing |
| Overall health | Very Low | No effect | Clift et al | 2010 | Narrative | 1 | Nonrandomized experimental study | USA | 128 | Older adults (over 64), mean age 79, mixed sex | 2 periods of 30 weeks over 2 years | Once per week (duration unspecified) | Singing |
| Perceived self-efficacy | Low | No effect | Raglio et al. | 2019 | Narrative | 1 | RCT (only comparative, not inactive control group) | South Korea | 20 | Graduate female music therapy students, mean age 28 | 4 weeks | 60 minutes, weekly | Drums |
| Personal accomplishment | Low | No effect | Yap, Kwan & Ang | 2017 | Narrative | 1 | Nonrandomized crossover experimental study | USA | 75 | Nursing students, mean age 28, mixed sex | 6 weeks | 1 hour, weekly | Drums |
| Positive affect | Low | Positive | Fancourt, Ockelford & Belai | 2014 | Narrative | 1 | Non-randomized intervention study | Germany | 31 | Adult amateur singers (mean age 56.9, range 29-74), mixed sex | 1 hour | Once | Singing |
| Positive affect | Low | Positive | Yap, Kwan & Ang | 2017 | Narrative | 1 | Nonrandomized experimental study | UK | 32 | Adults, mean age 25 - 44, mixed sex | 30 minutes | Once | Drums |
| Positive affect | Low | Positive | Raglio et al. | 2019 | Narrative | 1 | RCT (only comparative, not inactive control group) | South Korea | 20 | Graduate female music therapy students, mean age 28 | 4 weeks | 60 minutes, weekly | Drums |
| Positive affect | Very Low | Positive | Clift et al | 2010 | Narrative | 1 | Single group experimental study (pre-post analysis) | Finland | 212 | Amateur/advanced choir members, mean age 53, mixed sex | 1 rehearsal (duration unspecified) | Once | Singing |
| Positive affect | Very Low | Positive | Gick | 2012 | Narrative | 1 | Cross-sectional | Sweden | 16 | Amateur/professional singers, mean age 40, mixed sex | 45 min | Once | Singing |
| Quality of life | Very Low | Positive | Phillips & Becker | 2019 | Narrative | 1 | Single group study (uncontrolled) | USA | 62 | Healthcare workers, mean age 48, mixed sex | 5 weeks | 90 minutes, weekly | Singing |
| Quality of life (mental) | High | Positive | Daykin et al | 2018 | Narrative | 1 | RCT | UK | 258 | Older adults, mean age 69, mixed sex | 14 weeks | 90 minutes, weekly | Singing |
| Quality of life (physical) | High | No effect | Daykin et al | 2018 | Narrative | 1 | RCT | UK | 258 | Older adults, mean age 69, mixed sex | 14 weeks | 90 minutes, weekly | Singing |
| Spiritual growth | Low | Positive | Daykin et al | 2018 | Narrative | 1 | Non randomized longitudinal intervention study | UK | 98 | Older adults, mean age 68, mixed sex | 10 weeks | 60-90 minutes, weekly | Instrumental |

| | | | | | | | | | | | | | |
|---|---|---|---|---|---|---|---|---|---|---|---|---|---|
| Spiritual Wellbeing | Very Low | Positive | Phillips & Becker | 2019 | Narrative | 1 | Single group study (uncontrolled) | USA | 62 | Healthcare workers, mean age 48, mixed sex | 5 weeks | 90 minutes, weekly | Singing |
| Stress | Very Low | Positive | Phillips & Becker | 2019 | Narrative | 1 | Single group study (uncontrolled) | USA | 62 | Healthcare workers, mean age 48, mixed sex | 5 weeks | 90 minutes, weekly | Singing |
| Stress management | Low | No effect | Daykin et al | 2018 | Narrative | 1 | Non randomized longitudinal intervention study | UK | 98 | Older adults, mean age 68, mixed sex | 10 weeks | 60-90 minutes, weekly | Instrumental |
| Stress/anxiety | Low | Positive | Yap, Kwan & Ang | 2017 | Narrative | 1 | Single group experimental study (pre-post analysis) | South Africa | 34 | Young adult novice drummers & middle aged experienced drummers, mean age 23 - 53, mixed sex | 40 minutes | Once | Drums |
| Vigor | Very low | No effect | Fancourt, Ockelford & Belai | 2014 | Narrative | 1 | Single group study (uncontrolled) | Japan | 54 | Adult & older adult nonmusicians (mean age 27-70, mixed sex) | 1 hour | Once | Drums |
| Vigor/arousal | High | Positive | Daykin et al | 2018 | Narrative | 1 | RCT | Germany | 154 | Adults, mean age 25, mixed sex | 41 minutes | Once | Drums |
| Wellbeing | Low | No effect | Daykin et al | 2018 | Narrative | 2 | Nonrandomized experimental study | UK, USA | 166 | Male inmates; older adults, mean age 34-68, mixed sex | One performance; 10 weeks | Once; 60-90 mins/week | Singing, instrumental |
| ***Domain: Social functioning*** | | | | | | | | | | | | | |
| Aggression | Very Low | Positive | Yap, Kwan & Ang | 2017 | Narrative | 1 | Nonrandomized experimental study | USA | 50 | University students, mean age 21, mixed sex | 45 min - 1 hour | Once | Drums |
| Anger | Moderate | No effect | Daykin et al | 2013 | Narrative | 1 | RCT | USA | 52 | at risk' youth (aged 14-18), mixed sex | 6 weeks | 1 hour, weekly | Music (Percussion & keyboard education) |
| Anger | Very low | Positive | Fancourt, Ockelford & Belai | 2014 | Narrative | 1 | Single group study (uncontrolled) | Japan | 54 | Adult & older adult nonmusicians (mean age 27-70, mixed sex) | 1 hour | Once | Drums |
| Anger | High | Positive | Daykin et al | 2018 | Narrative | 1 | RCT | Germany | 154 | Adults, mean age 25, mixed sex | 41 minutes | Once | Drums |
| Anger (state) | Very Low | No effect | Phillips & Becker | 2019 | Narrative | 1 | Single group study (uncontrolled) | USA | 62 | Healthcare workers, mean age 48, mixed sex | 5 weeks | 90 minutes, weekly | Singing |
| Anger (trait) | Very Low | Positive | Phillips & Becker | 2019 | Narrative | 1 | Single group study (uncontrolled) | USA | 62 | Healthcare workers, mean age 48, mixed sex | 5 weeks | 90 minutes, weekly | Singing |

| | | | | | | | | | | | | | |
|---|---|---|---|---|---|---|---|---|---|---|---|---|---|
| Job engagement | Low | No effect | Raglio et al. | 2019 | Narrative | 1 | RCT (only comparative, not inactive control group) | South Korea | 20 | Graduate female music therapy students, mean age 28 | 4 weeks | 60 minutes, weekly | Drums |
| Psychosocial function | Moderate | No effect | Daykin et al | 2013 | Narrative | 1 | RCT | USA | 52 | at risk' youth (aged 14-18), mixed sex | | 1 hour, weekly | Music (Percussion & keyboard education) |
| Social activity level | Very Low | Positive | Clift et al | 2010 | Narrative | 1 | Nonrandomized experimental study | USA | 128 | Older adults (over 64), mean age 79, mixed sex | 2 periods of 30 weeks over 2 years | Once per week (duration unspecified) | Singing |
| Social anxiety | Low | No effect | Fancourt, Ockelford & Belai | 2014 | Narrative | 1 | Non-randomized longitudinal cohort study | Sweden | 60 | 5th & 6th graders, sex unspecified (likely mixed) | 8 months | 1 hour, weekly | Music education (general) |
| Social engagement (activity level) | Very Low | Positive | Daykin et al | 2018 | Narrative | 1 | Nonrandomized experimental study | USA | 166 | Older adults, mean age 74, sex unspecified | 30 weeks | Once per week (duration unspecified) | Singing |
| Social functioning | Moderate | No effect | Daykin et al | 2013 | Narrative | 1 | RCT | USA | 52 | at risk' youth (aged 14-18), mixed sex | 6 weeks | 1 hour, weekly | Percussion & keyboard |
| Social functioning | Low | No effect | Fancourt, Ockelford & Belai | 2014 | Narrative | 1 | Non-randomized longitudinal cohort study | Sweden | 60 | 5th & 6th graders, sex unspecified (likely mixed) | 8 months | 1 hour, weekly | Music education (general) |
| Social functioning | Low | Positive | Yap, Kwan & Ang | 2017 | Narrative | 1 | Nonrandomized intervention study | USA | 101 | 5th graders, mean age 11, mixed sex | 12 weeks | 40-45 minutes, weekly | Drums |
| Social functioning | Low | No effect | Daykin et al | 2018 | Narrative | 1 | Non randomized longitudinal intervention study | UK | 98 | Older adults, mean age 68, mixed sex | 10 weeks | 60-90 minutes, weekly | Instrumental |
| Social functioning | Low | No effect | Clift et al | 2010 | Narrative | 2 | Nonrandomized experimental cohort study; cross-sectional | UK, USA | 159 | Retirees (amateur musicians & nonmusicians), mean age 65-84, mixed sex | 4 weeks; unspecified | 1 hour, weekly; unspecified | Singing |
| Somatic symptoms | Low | No effect | Clift et al | 2010 | Narrative | 1 | Non randomized experimental cohort study | UK | 61 | Residents of retirement homes (mean age 84), mixed sex | 4 weeks | 1 hour | Singing |

*Table S5. Summary of included reviews*

| Author | Year | Music / Dance? | Type of synthesis | Review objective(s) | # databases searched | Additional manual search? | Search date range | Inclusion criteria | Exclusion criteria | Domain(s) |
|---|---|---|---|---|---|---|---|---|---|---|
| Burkhardt & Brennan | 2012 | Dance | Narrative | Explore the effects of recreational dance interventions on the health and well-being of children and young people | 6 | No | 1947-2009 | 1) studies reporting the effects of a recreational dance intervention on the physical, psychological or social health of young people aged 5-21; 2) controlled trials, cohort studies, case control studies, cross-sectional studies with control group for comparison | 1) Dance Movement Therapy or video dance games; 2) dance programs leading to professional dance; 3) participants were professional dancers; 4) non-controlled studies, observational studies, studies with no quantitative or statistical analysis of results | Blood pressure; Body composition; Bone health; Developmental; Mental health; Physical fitness; Physical function; Self-reported health/wellbeing |
| Butzlaff | 2000 | Music | Meta-analysis | Can music be used to enhance reading abilities? | 7 | Yes | Inception-1998 | 1) standardized measure of reading ability was used as the dependent variable; 2) a test of reading followed music 'instruction'; 3) statistical information was sufficient to allow for the calculation of an effect size | None specified | Educational |
| Clarke et al. | 2018 | Dance | Narrative | Identify all relevant literature on balance and theatrical styles of dance, involving adult participants who were either in full-time dance training or professional dancers | 7 | Yes | 1980-2016 | 1) experimental study; 2) referred to theatrical dance forms; 3) involved professional dancers and/or dance students in vocational and university training; 4) examined balance | 1) recreational or competition dance; 2) participants aged younger than 17 and/or older than 45 years old | Physical function |
| Clift et al | 2010 | Music | Narrative | Systematically map the research literature on group singing, wellbeing and health | 4 | Yes | 1985-2009 | 1) research which involves clearly identified singing groups (amateur & professional) & outcomes related to their wellbeing effects | 1) studies concerned with individual singing/individual music therapy interventions involving singing, studies of song writing as a form of singing, care-giver singing; 2) singing combined with other forms of music-making or creative activity | Auditory; Endothelial function/stress response; Immune function/inflammation; Mental health; Physical function; Self-reported health/wellbeing; Social functioning |
| Coffey, Mogilever & Zatorre | 2017 | Music | Narrative | 1) Consider how speech in noise tasks may vary; 2) consider coverage of the research area by different neuroimaging measures that offer complementary views of the neurophysiological basis of speech in noise differences between musicians and non-musicians; 3) suggest a means of comparing and planning future studies; 4) highlight specific research areas for future study | 1 | No | Inception-2016 | Studies of musician effects on speech-in-noise perception within neurologically normal populations | None specified | Auditory; Cognitive; Educational |

| | | | | | | | | | | |
|---|---|---|---|---|---|---|---|---|---|---|
| Costa, Ferreira & Felicio | 2013 | Dance | Narrative | Review studies about static and dynamic postural balance of ballet dancers, characterizing visual dependency in the postural control of these athletes to maintain balance | 5 | No | 1997-2013 | 1) studies involving assessment of static or dynamic balance in ballet dancers | None specified | Physical function |
| Cugusi et al | 2019 | Dance | Narrative | Systematically summarize, analyze and interpret the cardiovascular benefits of Zumba Fitness in women, in order to outline its potential roles in primary prevention of cardiovascular disease in both healthy subjects and women presenting with specific cardiovascular risk factors | 5 | Yes | Inception-2017 | 1) women of all ages with or without specific cardiovascular risk factors; 2) analyses of cardiovascular outcomes arising from a Zumba fitness program lasting 2+ weeks | None specified | Blood pressure; Body composition; Glucose/insulin; Immune function/inflammation; lipid lipoprotein profile; Physical fitness |
| Daykin et al | 2013 | Music | Narrative | What is the evidence surrounding the impact of music-making on the well-being, health and behavior of young offenders? | 11 | Yes | 1996-2013 | Interventions with young people aged 11-25 in a range of youth justice settings (custodial settings + 'at risk' populations) | 1) Studies conducted in the general population; 2) studies of disadvantaged and clinical populations who share some characteristics of young offenders but who were not targeted on the basis of having committed/being at risk of committing an offence | Mental health; Social functioning |
| Daykin et al | 2018 | Music | Narrative | What are the wellbeing outcomes of music and singing for adults and what are the processes by which wellbeing outcomes are achieved? | 9 | Yes | 1996-2016 | Empirical studies in any language from countries economically similar to the UK; studies assessed individual/group music interventions and any recognized measure of subjective wellbeing in adults; quantitative studies only included with a concurrent comparator | 1) Research subjects are paid professionals; 2) music used for clinical purposes (pain management, symptom relief); 3) music therapy interventions seeking to deliver wellbeing outcomes | Blood pressure; Endothelial function/stress response; Mental health; Physical function; Self-reported health/wellbeing; Social functioning |
| Eells | 2014 | Music | Narrative | Explore the literature on the effects of music and singing as a nursing intervention on the symptoms of anxiety in older adults | 3 | Yes | Inception-2011 | 1) participants over the age of 65; 2) related to anxiety and the use of music/singing as a nursing intervention; 3) Related to service user experiences of anxiety and the use of music/singing interventions | None specified | ***All studies meeting umbrella review inclusion criteria included in other reviews*** |
| Fancourt, Ockelford & Belai | 2014 | Music | Narrative | Consolidate key findings to date, compare theories concerning the mechanisms behind music‘s effect, and highlight gaps in current knowledge, helping to guide the focus of future studies | 6 | Yes | 1953-2013 | 1) new study; 2) ,controlled in order that the significance of alterations in biomarkers could be accurately assessed; 3) studies pairing music with other stimuli (i.e. exercise) were only included if they also contained a test incorporating just music on its own; 4) studies had to be | Animal studies | Autonomic tone; Endothelial function/stress response; Immune function/inflammation; Mental health; Self-reported health/wellbeing; Social functioning |

| | | | | | | | | | | |
|---|---|---|---|---|---|---|---|---|---|---|
| | | | | | | | | testing for potential positive effects of music | | |
| Fernandez-Arguelles et al. | 2015 | Dance | Narrative | Determine the therapeutic effects of dancing as a physical exercise modality on balance, flexibility, gait and muscle strength in older adults | 5 | Unspecified | 2000 - 2013 | 1) Studies analyzing the effects of dance (ballroom dance and/or dance based exercise) in older adults over 60 years of age with no disabling disease on balance, gait, dynamic mobility, strength, and flexibility outcomes; 2) Studies in English or Spanish | 1) Qualitative studies that did not quantify results; 2) Studies in which participants presented with a cognitive deficit and/or mental or neurological (physical or psychological) condition | ***All studies meeting umbrella review inclusion criteria included in other reviews*** |
| Fong Yan et al. | 2018 | Dance | Meta-analysis<br>Narrative | Systematically review the literature to investigate the effectiveness of structured dance interventions, in comparison to structured exercise programs, on physical health outcome measures | 7 | Yes | Inception-2017 | 1) original study comparing forms of dance intervention to a structured exercise program; 2) dance classes must have been used in the intervention; 3) minimum 4 week intervention duration; 4) measurable physical health outcomes; 5) comparison group that performed a structured exercise program | 1) literature reviews or case studies; 2) investigated psychological and/or mental health outcome measures only; 3) examined the effect of dance therapy, movement therapy, or creative movement; 4) dance intervention combined with other types of structured exercise; 5) control group did not exercise and/or were given unstructured physical activity | Body composition; Lipid lipoprotein profile; Physical fitness; Physical function<br>Blood pressure; Body composition; Bone health; Glucose/insulin; Lipid lipoprotein profile; Physical fitness; Physical function |
| Gick | 2012 | Music | Narrative | Consolidate research on singing, health and well-being into a common conceptual framework in order to facilitate its evaluation, consideration of possible mechanisms by which singing may have its effects, and suggestions for further research | 4 | Yes | Inception-2012 | 1) articles investigating active singing and some health or well-being outcome; 2) English language articles | 1) studies which investigated a multi-component treatment including singing with other interventions (musical or other); No data reported; attempts to obtain the article failed | Endothelial function/stress response; Immune function/inflammation; physical fitness; self-reported health/wellbeing |
| Gordon, Fehd & McCandliss | 2015 | Music | Meta-analysis | Synthesize previous research on music training and reading-related outcomes | 14 | Unspecified | Inception-2014 | 1) intervention with a control group; 2) peer-reviewed publication; 3) reported phonological or reading-related outcomes; 4) assessed outcomes pre- and post-intervention; 5) provided sufficient data to extract effect sizes; 6) intervention group received more music instruction than control group; 7) studies need to provide equivalent amounts of reading instruction across the intervention & control groups | None specified | Educational |
| Hetland | 2000 | Music | Meta-analysis | Does active instruction in music enhance preschool and elementary students‘ performance on spatial tasks? | 7 | Yes | Inception-1998 | 1) experiments reported in English that included a condition in which participants were taught to make music (vocal or instrumental); 2) one or more control conditions in which participants did not receive instruction in actively making | None specified | Cognitive |

| | | | | | | | | | |
|---|---|---|---|---|---|---|---|---|---|
| | | | | | | | music; 3) outcome measure of performance on one or more spatial tasks that required mental rotation and/or spatial visualization; 4) studies had to provide statistics necessary to compute an effect size | | |
| Hwang & Braun | 2015 | Dance | Narrative | Examine the effectiveness of dance programs in improving the physical health of all older adults, both those with health conditions and those considered healthy | 1 | Yes | Inception-2013 | 1) evaluate the benefits of a dance intervention to the physical health of older adults (aged 65+) lacking prior dance experience; 2) present results in quantitative format | 1) 'aerobic fitness classes taught to music' (i.e. Zumba, step-aerobics); 2) non-English publications | Physical function |
| Jaschke et al. | 2013 | Music | Narrative | To reflect the variation in published results in the field of music education and the far transfer effect and to show how difficult it is to interpret these results when different methods are used to measure them and their discussion is hampered by the absence of a proper classification of far transfer and the lack of a structured understanding of music and musicality' | 6 | Yes | 2001-2013 | Studies (RCT or non-randomized) of the effect of music education or participation on reading, spatial reasoning, writing, mathematics and intelligence in youth aged 4 to 13 | 1) Studies excluded when effects measured on the basis of study size rather than sample size; 2) no control group; 3) non-randomized longitudinal studies lasting <12 months; 4) meta-analyses; 5) analyzed near transfer | Cognitive; Educational |
| Keogh et al. | 2009 | Dance | Narrative | Review the physical benefits of dancing for healthy older adults | 5 | Yes | Inception-2009 | 1) apparently healthy older adults 60+ years old; 2) Compare a group of older dancers with an age matched group of nondancers or involve a dance intervention that lasted at least 8 weeks | None specified | Bone health |
| Letton, Thom, & Ward | 2020 | Dance | Narrative | Systematically identify and synthesize literature investigating the effectiveness of classical ballet training interventions on physical healthy and psychological outcome measures | 10 | Yes | Inception-2019 | Original research investigating a classical ballet training intervention (>/= 4 weeks) or reporting on observations of preexisting ballet classes or reporting on observations of pre-exiting ballet classes; reporting on measurable health outcomes relating to physical health, psychological functioning, or physiological health | 1) Review articles, single case studies; 2) non-English articles; 3) combined ballet training with other exercise or types of dance | Body composition; Developmental; Physical fitness; Physical function |
| Liu, Shen & Tsai | 2020 | Dance | Meta-analysis | Conduct a systematic analysis of random controlled research on the effectiveness of dance intervention in the physical function of healthy older adults | 5 | Yes | Inception-2018 | 1) RCT; 2) intervention duration at least 6 weeks; 3) healthy older adults aged at least 65; 4) English | 1) Uncertain study design/data; 2) dance intervention combined with other exercise | Physical function; Self-reported health/wellbeing |
| | | | Narrative | | | | | | | Physical fitness; Physical function |

| | | | | | | | | | | |
|---|---|---|---|---|---|---|---|---|---|---|
| Meng et al. | 2020 | Dance | Meta-analysis; Narrative | Assess the effects of dance interventions on global cognition, executive function and memory in older adults | 9 | Yes | Inception-2018 | 1) RCT or quasi-experimental; 2) participants were aged 60+; 3) Dance intervention (any style) longer than 4 weeks; 4) cognitive function reported as the primary or secondary outcome | 1) combined dance & other intervention | Cognitive; Cognitive |
| Moore | 2013 | Music | Narrative | Explore and synthesize results examining the effects of music on neural structures implicated in emotion regulation; Create preliminary clinical considerations for structuring the music stimulus when facilitating emotion regulation | 8 | No | Inception-2011 | 1) Primary research study; 2) participants are typically-developing humans, with no restrictions as to age, gender, ethnicity, or type of setting; 3) music was the primary stimulus, regardless of how it was implemented, the genre of music, or the music instruments incorporated; 4) study results reported on the impact of music on one or more of the following structures (amygdala, anterior cingulate cortex, orbitofrontal cortex, and lateral prefrontal cortex); 5) articles published in English language peer-reviewed journals | 1) Review articles/theoretical paper; 2) participants had a disorder, brain damage, or a syndrome; 3) study of rhythm was associated with circadian rhythm, cardiac rhythm, respiratory rhythm, dietary rhythm, and other nonmusical references to rhythm | Mental health |
| Phillips & Becker | 2019 | Music *(no studies of dance meeting inclusion criteria)* | Narrative | To synthesize evidence about the effectiveness of expressive arts interventions used to reduce psychosocial stress among healthcare workers | 4 | No | 1997 - 2017 | 1) intervention studies that address workplace stress, professional grief, compassion fatigue, secondary traumatic stress, or burnout; 2) interventions that used the expressive arts (music, poetry, narrative, storytelling, writing, singing, dancing, visual, or tactile art; 3) Quantitative and mixed methods studies that evaluated the effectiveness or acceptability of the intervention; 4) intervention studies that targeted any healthcare worker and/or any healthcare setting; 5) articles written in English | 1) intervention not aimed at workplace stress and/or did not use the expressive arts; 2) intervention was for non-healthcare workers and/or students; 3) article was a systematic review, meta-analysis, primarily qualitative study, case report, dissertation/thesis or conference abstract | Mental health; Self-reported health/wellbeing |
| Raglio et al. | 2019 | Music | Narrative | Select the evidence-based studies that took advantage of music to improve work-related psychological conditions and cognitive performances, highlighting perspectives in the implementation of music intervention in occupational medicine | 2 | No | 2000 - 2017 | RCTs or clinical controlled trials in English from peer-reviewed journals | None specified | Mental health; Self-reported health/wellbeing; Social functioning |
| Rodrigues-Krause et al | 2016 | Dance | Narrative; Meta-analysis | Verify the level of evidence regarding the adaptations of dance interventions on CVR factors in the elderly | 5 | Yes | 1980 - 2015 | 1) RCTs and nRCTs comparing dance interventions with a non-exercising control group and/or other exercising groups receiving another intervention; 2) | 1) Less than 4 weeks of intervention; 2) elderly patients with neurodegenerative diseases; 3) combined interventions (exercise + | ***All studies meeting umbrella review inclusion criteria included in other reviews*** |

| | | | | | | | | | | |
|---|---|---|---|---|---|---|---|---|---|---|
| | | | | | | | | Elderly subjects (mean age >60 years) with or without health conditions; 3) Designed to improve outcomes of interest; 4) Data reported as means or difference in between means and respective dispersion values at baseline and after the intervention period; 5) English, Portuguese or Spanish full text | dance); 3) duplicate publications/sub-studies; 5) studies including outcomes of no interest (i.e. aerobic fitness estimated by the 6-minute walk test) | |
| Rodrigues-Krause, Krause, Reischak-Oliveira | 2019 | Dance | Narrative | Review the literature on the use of dance as a form of intervention to promote functional and metabolic healthy in older adults | 5 | Yes | 1980 - 2016 | 1) RCT; 2) quasi-experimental trials, including pre- and postintervention studies with 1 group, nonrandomized controlled trials and studies with other nonrandom distributions; 3) observational designs - cross-sectional or cohorts | 1) evaluated older patients with cognitive and/or emotional impairment; 2) dance environment categorized as a dance videogame or dance on the water; 3) had study designs that did not fit the current study's protocols or were case studies | Bone health; Cognitive; Glucose/insulin; Lipid lipoprotein profile; Mental health; Physical fitness; Physical function; Self-reported health/wellbeing; |
| Skingley & Vella-Burrows | 2010 | Music | Narrative | Can music and singing contribute to the health and wellbeing of older people and are they relevant to the role of nurses? | 2 | Yes | 1997-2010 | 1) Empirical research studies or reviews of music or singing interventions for older people (65 years+); 2) Qualitative studies exploring music and singing for older people; 3) Studies published in English; 4) Studies appearing in nursing and other arts and health journals | 1) Studies of music therapy; 2) Studies unrelated to older people; 3) Studies where the intervention is primarily relevant to health professionals other than nurses | ***All studies meeting umbrella review inclusion criteria included in other reviews*** |
| Talamini et al | 2017 | Music | Meta-analysis | Whether there really is a difference between musicians and nonmusicians and whether the magnitude of any difference varies as a function of the memory system involved, and the type of stimuli presented | 9 | Yes | Inception-2017 | 1) adult participants; 2) study included a group of expert musicians ('participants who attended music conservatories or music schools') and a group of nonmusicians; 3) studies that administered a memory task to both groups related to long-term, short-term or working memory; 4) studies using verbal, visual, spatial or tonal stimuli; 5) studies published in English | 1) Missing data; 2) tasks not comparable with other studies considered | Cognitive |
| Teixeira-Machado, Arida & de Jesus Mari | 2019 | Dance | Narrative | What is the influence of dance practice on neuroplasticity in already mature brains? | 3 | No | Inception-2018 | 1) randomized clinical trial studies that observed the maturation and activation of the central nervous system through dance; 2) studies on humans of all ages and both genders included; 3) dance interventions and research comparing dancing with other interventions (i.e. physical exercise, sports) | None specified | Cognitive; Physical fitness; Physical function; Self-reported health/wellbeing; Social functioning |
| Vaughn | 2000 | Music | Meta-analysis | Conduct meta-analyses investigating the relationship between music and mathematics | 7 | Yes | Inception-1998 | 1) studies testing an association between music and mathematics; 2) studies with a measured math outcome; 3) studies with a control group; 4) sufficient statistical | 1) studies in which music was used as a reward for good performance in math; 2) studies in which musical jingles were used as memory aids to | Educational |

| | | | | | | | | | | |
|---|---|---|---|---|---|---|---|---|---|---|
| | | | | | | | | information for an effect size to be computed | teach math; 3) studies assessing whether individuals who have a high musical aptitude also have a high mathematical aptitude | |
| Veronese et al. | 2017 | Dance | Narrative | Investigate if dancing is more effective than usual care in preventing falling and improving fear of falling through a systematic revision of the RCTs available | 5 | Yes | Inception - 1 March 2017 | 1) RCT; 2) investigated the effect of a dancing program; 3) included data regarding falls or fear of falls using validated scales | 1) did not include humans; 2) did not include a control group; 3) did not use validated scales for assessing fear of falls; 4) did not report data at follow-up evaluation regarding falls/fear of falling | Physical function |
| Yap, Kwan & Ang | 2017 | Music | Narrative | To provide a systematic review on the current literature of the effects of rhythm-based music making on different aspects of health | 2 | Yes | Inception-October 2015 | English-language peer reviewed journal articles that studies the effects of drumming or percussion music on an individual | 1) Review articles, meta-analyses, case series, case reports; 2) studies without full articles; 3) studies that included drumming or percussing music making as part of a protocol with the inclusion of other intervention modalities | Blood pressure; Immune function/inflammation; Mental health; Self-reported health/wellbeing; Social functioning |

*Table S6. GRADE publication bias ratings for included reviews containing quantitative synthesis (meta-analysis). NB: publication bias ratings of 'Serious' and 'Very Serious' resulted in a down-rating of the certainty of corresponding review-level evidence by 1 or 2 levels, respectively, as per GRADE guidelines. Evidence evaluated as per criteria established and validated in:*

Meader et al. (2014) A checklist designed to aid consistency and reproducibility of GRADE assessments: development and pilot validation. *Systematic Reviews.*

| | | **GRADE publication bias evaluation** | | | | | | |
|---|---|---|---|---|---|---|---|---|
| **Author** | **Year** | *Did the authors conduct a comprehensive search?* | *Did the authors search for grey literature* | *Authors did NOT apply restrictions to study selection on the basis of language?* | *There was no industry influence on included studies?* | *(meta-analysis only) There was no evidence of funnel plot asymmetry?* | *There was no discrepancy in findings between published and unpublished trials?* | **GRADE publication bias** |
| Butzlaff | 2000 | Yes | Yes | No | Yes | Yes | N/A (not analyzed) | Uncertain |
| Fong Yan et al. | 2018 | Yes | Yes | Yes | Yes | Yes | N/A (not analyzed) | Uncertain |
| Gordon, Fehd & McCandliss | 2015 | No | No (explicit exclusion of grey literature) | No | Yes | Somewhat (no funnel plot asymmetry, but significant heterogeneity) | N/A (not analyzed) | Very serious (significant heterogeneity, plus no hand searches of included articles, exclusion of grey literature, and language restriction) |
| Hetland | 2000 | Yes | Yes | No | Yes | Yes | N/A (not analyzed) | Uncertain |
| Liu, Shen & Tsai | 2020 | Yes | No | No | Yes | Yes | N/A (not analyzed) | Serious (restricted to English studies + no grey literature) |
| Meng et al. | 2020 | Yes | Yes | Yes | Yes | Unclear | N/A (not analyzed) | Uncertain |
| Talamini et al | 2017 | Yes | No (but statistical corrections for potential publication bias from this exclusion) | No | Yes | Yes | N/A (not analyzed) | Uncertain |
| Vaughn | 2000 | Yes | Yes | No | Yes | Yes | N/A (not analyzed) | Uncertain |

*Table S7. Details of individual studies in included reviews of dance participation.*

| *Author* | *Year* | *In other review / other exclusion?* | *Industry support?* | *Design* | *Subjects* | *Study country* | *Type of dance participation* | *Length of participation* | *Frequency/ duration of each session* | *Outcome* | *Effect of dance participation* | *Outcome details* | *GRADE Certainty of Evidence* |
|---|---|---|---|---|---|---|---|---|---|---|---|---|---|
| ***Burkhardt & Brennan, 2012*** | | | | | | | | | | | | | |
| Adiputra et al | 1996 | | No | Non-randomized experimental study | 60 male students of the Senior Dancing School of Denpasar, Bali (age range 17-19 years; mean 19.6) | Indonesia | Balinese dance | 8 weeks | 50 min/3x weekly | VO2 max | Positive | Significant increase in VO2 max in dance vs control group | Low |
| Bennell et al | 2000 | | No | Cross-sectional | 130 females aged 8-11 (mean age = 9.6; 78 pre-and early pubertal novice dancers, 52 age-matched controls) | Australia | Ballet + various | Unspecified | Unspecified | Bone mineral density | Positive | Significantly greater bone mineral density in dancers vs controls (hip & femoral neck) | Low |
| Blackman et al | 1988 | | No | Non-randomized experimental study | 16 females (age range 14-15; 8 on dance team, 8 physical education control | USA | Dance team | 5 months | 1 hour/5x weekly | VO2 max | Positive | Significant pre-post increase in VO2 max in dance group | Very Low |
| | | | | | | | | | | Sit and reach | No effect | No pre-post effect | Very Low |
| | | | | | | | | | | Strength | Positive | Significant pre-post increase in dance group (1-RM bench press) | Very Low |
| | | | | | | | | | | % body fat | Mixed | Significant pre-post difference in total skinfold measurements, but not hydrostatic weighing (skinfolds, hydrostatic weighing) | Very Low |
| | | | | | | | | | | Self esteem | NO effect | No significant effects (Coopersmith Self-Esteem Inventory, Tennessee Self-Concept Scale) | Very Low |
| | | | | | | | | | | Body satisfaction (Cathexis) | No effect | No significant effects (Body Cathexis Scale) | Very Low |
| | | | | | | | | | | Blood Pressure | Positive | Significant pre-post decrease in blood pressure | Very Low |
| | | | | | | | | | | Body mass | No effect | No significant pre-post effect | Very Low |

| | | | | | | | | | | | | |
|---|---|---|---|---|---|---|---|---|---|---|---|---|
| Burgess, Grogan & Burwitz | 2006 | No | RCT | 50 healthy British schoolgirls (mean age = 13.5; 25 dance group, 25 physical education) | UK | Aerobic dance | 6 weeks | 50 min/2x weekly | Self-perception | Positive | Significant group*time effects (Children & Youth Physical Self-Perception Profile) | Moderate |
| | | Other review - Fong Yan et al., 2018 | | | | | | | BMI | | | |
| Daley & Buchanan | 1999 | No | Non-randomized experimental study | 113 females (age range 15-16; 43 aerobic dance group, 70 physical education control) | UK | Aerobic dance | 5 weeks | 1 hour/weekly | Self-perception | Positive | Significant group*time effects (Physical Self Perception Profile) | Low |
| Flores | 1995 | No | RCT | 81 children (age range 10-13; 43 dance group, 38 physical activity control), sex unspecified | USA | Hip-hop, African dance, & aerobic | 12 weeks | 50 min/3x weekly | Timed mile run | No effect | No significant pre-post or intervention vs control differences | Moderate |
| | | | | | | | | | BMI | Positive | Significant pre-post reduction vs control | Moderate |
| Kim & Kim | 2007 | No | RCT | 277 high school and undergraduate students (mean age = 20.6; 45 hip hop dancing group - all female, 84 aerobic exercise group - 40 male/44 female, 64 body conditioning group - 44 male, 20 female, 84 ice skating group - 60 male/24 female) | South Korea | Hip hop | 50 min | Once | Mood | Positive | Significant increase in positive well-being and decrease in psychological distress and fatigue in dance & aerobics groups vs ice skating & body conditioning groups (Subjective Exercise Experiences Scale) | Moderate |
| Leste & Rust | 1990 | No | Non-randomized experimental study | 84 university students (mean age = 19.9; 23 dance group, 16 sport group, 7 music group, 38 mathematics group), sex breakdown unspecified - mixed | UK | Modern | 12 weeks | Unspecified | Anxiety | Positive | Significant benefit of dance intervention vs music/physical exercise/math groups (Spielberger State-Trait Anxiety Inventory) | Low |
| Matthews et al | 2006 | No | Prospective cohort study | 143 girls (age range 8-14; 82 non-elite dancers, 61 controls) | Australia | Ballet | 3 years | Unspecified | Bone mineral density | Positive | Significantly greater bone mineral content in dancers vs controls (total body - DXA) | Low |
| Mavridis et al | 2004 | Other review - Fong Yan et al., 2018 | | | | | | | | | | |

| | | | | | | | | | | | | |
|---|---|---|---|---|---|---|---|---|---|---|---|---|
| Robinson et al | 2003 | | RCT | 61 African American females (age range 8-10, BMI > 50th percentile for age with at least 1 overweight parent/guardian (BMI = 25+) | USA | Hip hop, African dance & step aerobics | 12 weeks | 3x weekly | BMI | No effect | No significant group*time interaction (study underpowered to detect these differences) | Low |
| | | | | | | | | | Waist circumference | No effect | No significant group*time interaction (study underpowered to detect these differences) | Low |
| | | | | | | | | | Insulin | Not reported | | |
| | | | | | | | | | Glucose | Not reported | | |
| | | | | | | | | | Total cholesterol | Not reported | | |
| | | | | | | | | | HDL cholesterol | Not reported | | |
| | | | | | | | | | Triglycerides | Not reported | | |
| | | | | | | | | | Self-esteem | No effect | No significant group*time interaction (study underpowered to detect these differences)(Rosenberg self-esteem scale) | Low |
| Silvestri | 2004 | Excluded - dance with target HR (exercise, not performing arts) | | | | | | | | | | |
| Steinberg et al | 2008 | | Cross-sectional | 1708 young females (age range 8-16; 1482 dancers, 226 non-dancers) | Israel | Ballet and various | Unspecified | Mean 2.4-11.3 hours weekly dance | Musculoskeletal development | No effect | No significant differences between dance & control groups (14 anthropometric measurements) | Low |
| Viscki-Stalec et al | 2007 | No | Non-randomized experimental study | 220 3rd & 4th grade girls (115 aerobic dance group, 105 physical education group; unspecified age) | Croatia | Aerobic dance | 60 high school periods | Unspecified | Core strength | Positive | Significant group* time effect (Trunk Lift Test) | Low |
| | | | | | | | | | Power | Positive | Significant group*time effect (standing long jump) | Low |
| | | | | | | | | | Agility | Positive | Significant group*time effect (side step test) | Low |
| | | Other review - Fong Yan et al., 2018 | | | | | | | % fat mass; Body mass; Range of motion; Sit & reach; Step test | | | |
| ***Clarke et al., 2018*** | | | | | | | | | | | | |

| | | | | | | | | | | | | |
|---|---|---|---|---|---|---|---|---|---|---|---|---|
| Ambegaonkar et al. | 2013 | | Cross-sectional | 33 female dancers and nondancers (18 dancers, mean age = 20; 15 nondancers, mean age = 22.1) | USA | Modern dance | mean 13.1 years dance experience | Unspecified | Balance | Positive | Significantly better balance performance in dancers vs non-dancers (Balance Error Scoring System & SEBT) | Low |
| Ambegaonkar et al. | 2016 | Excluded - single group descriptive analysis of dancers only | | | | | | | | | | |
| Barcellos & Imbiriba | 2002 | Excluded - single group descriptive analysis of dancers only | | | | | | | | | | |
| Batson | 2010 | Excluded - single group descriptive analysis of dancers only | | | | | | | | | | |
| Bronner | 2012 | No | Cross-sectional | 27 dancers (9 expert, 5 male/4 female, mean age = 24.9; 9 advanced, 2 male/7 female, mean age = 19.6; 9 intermediate, 4 male/5 female, mean age = 19.8) | USA | Unspecified | Expert - mean 13.3 years dancing; advanced - mean 11.7 years dancing; intermediate - mean 6.1 years dancing | Unspecified | Balance | Positive | Significantly reduced postural variability in advanced/expert dancers vs intermediate | Low |
| Bruyneel et al. | 2010 | Excluded - single group descriptive analysis of dancers only | | | | | | | | | | |
| Bruyneel et al. | 2010 | Excluded - single group descriptive analysis of dancers only | | | | | | | | | | |
| Caplan & Gibson | 2011 | Excluded - single group descriptive analysis of dancers only | | | | | | | | | | |
| Casabona et al. | 2016 | No | Cross-sectional | 20 female dancers & non-dancers (10 professional dancers, mean age = 23.7; 10 untrained dancers, mean age = 27.6) | Italy | Ballet | Unspecified | Unspecified | Balance | Positive | Significantly better balance in dancers vs untrained, albeit only in 'duck' stance familiar to dancers | Low |
| Clark & Redding | 2012 | Excluded - single group descriptive analysis of dancers only | | | | | | | | | | |
| Cloak et al. | 2010 | Excluded - analysis of intervention study evaluating the impact of vibration training on balance in dancers only | | | | | | | | | | |
| Coutts et al. | 2006 | Excluded - single group descriptive analysis of dancers only | | | | | | | | | | |
| Crotts et al. | 1996 | No | Cross-sectional | 30 university students (mean age 27; 15 dancers; 15 non-dancers from physical therapy department), sex breakdown unspecified - mixed | USA | University dance major (varied) | mean 14.9 years dance experience | Unspecified | Balance | Positive | Significantly better balance in dancers in challenged visual & surface conditions | Low |

| | | | | | | | | | | | |
|---|---|---|---|---|---|---|---|---|---|---|---|
| da Costa et al. | 2013 | Excluded - single group descriptive analysis of dancers only | | | | | | | | | |
| Denardi, Ferraciolo & Rodrigues | 2008 | Excluded - single group descriptive analysis of dancers only | | | | | | | | | |
| Gerbino, Griffin & Zurakowski | 2007 | No | Cross-sectional | 64 female university students (32 dancers - mean age = 20.3; 32 soccer players - mean age = 19.7) | USA | Modern dance & ballet | Unspecified | mean 5.5 hours/day, 5.6 days/week | Balance | Positive | Significantly better balance performance by dancers in 5 of 20 measures | Low |
| Golomer & Dupui | 2000 | No | Cross-sectional | 41 dancers and nondancers (23 dancers - mean age 23.6; 13 female/10 male; 18 untrained - mean age - 21.5; 11 female/7 male) | France | Opera | Unspecified | Unspecified | Balance | Positive | Significantly better balance in dancers vs controls | Low |
| Golomer et al. | 1999 | Excluded - single group descriptive analysis of dancers only | | | | | | | | | |
| Golomer et al. | 1999 | No | Cross-sectional | 46 professional dancers & untrained males (17 professional dancers - mean age 23.8; 29 untrained - mean age ~22.6) | France | Ballet | Unspecified | Unspecified | Balance | Positive | Dancers displayed significantly improved balance in eyes closed tasks vs. untrained | Low |
| Golomer et al. | 2009 | No | Cross-sectional | 15 female dancers & untrained individuals (8 dancers, 7 untrained; mean age = 19) | France | Ballet | Unspecified | Unspecified | Balance | Positive | Significantly better shoulder-hip stabilization in dancers | Low |
| Golomer et al. | 2010 | No | Cross-sectional | 14 professional and untrained female dancers (mean age = 19) | France | Ballet | Minimum 11 years dance training | 20 hours/week from age 8-17 + 1-year professional training (35 hours/week) | Balance | Mixed | Significantly better balance in dancers vs control in 1-leg stance conditions; significantly worse performance by dancers vs control in 'roll equilibrium' conditions | Very Low |
| Golomer, Dupui & Monod | 1997 | No | Cross-sectional | 52 post-pubescent adolescents (31 professional dancers - mean age ~17.8, 15 | France | Ballet | Minimum 1 year professional experience | Unspecified | Balance | Positive | Significantly better balance in dancers vs. untrained cohort (4 | Low |

| | | | | | | | | | | | | |
|---|---|---|---|---|---|---|---|---|---|---|---|---|
| | | | | male/16 female; 21 untrained - mean age 18.6, 10 male/11 female | | | | | | | conditions - 2 eyes open, 2 eyes closed) | |
| Golomer, Gravenhorst & Toussaint | 2009 | Excluded - single group descriptive analysis of dancers only | | | | | | | | | | |
| Guillou, Dupui & Golomer | 2007 | No | Cross-sectional | 36 male dancers, gymnasts, athletes & laypeople (7 dancers, mean age = 18; 9 acrobatic gymnasts, mean age = 19.1; 10 professional soccer players, mean age = 17.1; 10 untrained, mean age = 21.4) | France | Opera | Unspecified | Unspecified | Balance | No effect | No significant differences between dancers & untrained | Low |
| Hopper et al. | 2014 | No | Cross-sectional | 23 dancers of varying experience levels (no gender listed) (9 professional dancers, mean age = 18.8; 6 pre-professional dancers, mean age = 17; 8 recreational dancers, mean age = 20.6) | Australia | Ballet | Professional - mean 12.3 years training; pre-professional - mean 12.7 years training; recreational - mean 16 years training | Professional - mean 20 hours/week; pre-professional - mean 15 hours/week; recreational - mean 3.7 hours/week | Balance | Positive | Significantly better balance after turns and fatiguing activity in professional vs pre-professional & recreational dancers | Low |
| Hugel et al. | 1999 | No | Cross-sectional | 64 professional & nondancers (18 dancers - age range 16-35, 6 male/12 female; 46 nondancers - age range 16-37) | France | Ballet | Mean 10-15 years training | Unspecified | Balance | Positive | Dancers displayed significantly improved balance in eyes open tasks vs. untrained | Low |
| Jarvis, Smith & Kulig | 2014 | No | Cross-sectional | 20 female dancers & non-dancers (10 professional dancers, mean age = 27.1; 10 non-dancers, mean age = 24.8) | USA | Unspecified | Minimum 10 years dance training | Unspecified | Balance | Positive | Significantly reduced variability in intersegmental coordination in dancers vs non-dancers | Low |
| Kiefer et al. | 2011 | Yes (Cincinnati Ballet Company) | Cross-sectional | 56 dancers and non-dancers (28 professional dancers, 10 male/18 female, | USA | Ballet | Unspecified | Unspecified | Balance | Positive | Significantly more consistent ankle hip coordination in | Low |

| | | | | | | | | | | | | |
|---|---|---|---|---|---|---|---|---|---|---|---|---|
| | | | | mean age = 23.6; 28 untrained, 10 male/18 female, mean age = 23.4) | | | | | | | postural corrections in dancers | |
| Kilroy et al. | 2016 | No | Cross-sectional | 14 female university students (age range 18-23; 7 dancers, 7 non-dancers) | USA | Unspecified | Minimum 7 years dance experience | Unspecified | Balance | Positive | Significantly better postural control in dancers vs non-dancers | Low |
| Krityakiarana & Jongkamonwiwat | 2016 | No | Cross-sectional | 50 female university students (25 dancers, mean age = 21.2; 25 non-dancers, mean age 21.2) | Thailand | Thai classical dance | mean 11.4 years dace experience | 4 hours/day, 3-4 days/week | Balance | Positive | Significantly better postural stability in dancers vs. non dancers in the majority of conditions | Low |
| Li et al. | 2014 | Excluded - single group descriptive analysis of dancers only | | | | | | | | | | |
| Lin et al. | 2011 | No | Cross-sectional | 22 university females (mean age = 19.9; 11 uninjured dancers, 11 non-dancers) | Taiwan | Ballet | minimum 7 years training | Unspecified | Balance | No effect | No significant differences in any balance (postural sway) measure between dancers & non-dancers | Low |
| Lin et al. | 2014 | No | Cross-sectional | 18 female dancers (9 super experienced dancers, mean age = 18.2; 9 experienced dancers, mean age = 18.3) | Taiwan | Ballet | novice - 2-5 years ballet training; experienced/ super-experienced (stratified based on maintenance of retire position for min 7 sec with each leg) - at least 6 years ballet training | novice - 1.5 - 3 hours training per week; (super)experienced - at least 3 hours training per week | Balance | Mixed | Experienced dancers had significantly better balance on the non-dominant leg; super experienced dancers had a greater maximum COM-COP distance in AP direction | Low |
| Lin et al. | 2014 | Excluded - single group descriptive analysis of dancers only | | | | | | | | | | |
| Lin, Su & Wu | 2005 | Excluded - single group descriptive analysis of dancers only | | | | | | | | | | |
| Mertz & Docherty | 2012 | Excluded - single group descriptive analysis of dancers only | | | | | | | | | | |
| Morrin & Redding | 2013 | Excluded - single group descriptive analysis of dancers only | | | | | | | | | | |

| | | | | | | | | | | | | | |
|---|---|---|---|---|---|---|---|---|---|---|---|---|---|
| Pappas et al. | 2011 | Excluded - single group descriptive analysis of dancers only | | | | | | | | | | | |
| Perez et al | 2014 | | No | Cross-sectional | 48 female university students (18 dancers, mean age 23.3; 30 non-dancers, mean age 22.2) | Spain | Ballet & contemporary | Minimum 5 years dance training | Unspecified | Balance | Positive | Significantly better balance in dancers vs non-dancers in eyes open conditions | Low |
| Perrin et al. | 2002 | | No | Cross-sectional | 56 professional dancers & nondancers (14 female dancers - mean age 22.1; 42 nondancers - mean age 23.9, 21 male/21 female) | France | Ballet | Mean 10-15 years training | Unspecified | Balance | Mixed | Significantly better balance in dancers vs controls in eyes closed conditions; significantly worse balance in dancers vs controls in eyes open conditions | Low |
| Rein et al. | 2011 | | Yes (Aircast Europe) | Cross-sectional | 90 dancers & non dancers (30 professional dancers - mean age = 27, 20 female/10 male; 30 amateur dancers - mean age = 34, 20 female/10 male; 30 non-dancers, mean age = 31, 15 female/15 male) | Germany | Unspecified | Unspecified | Unspecified | Balance | Mixed | Significantly better balance performance in professional, but not amateur, dancers vs control | Very Low |
| Schmit, Regis & Riley | 2005 | | No | Cross-sectional | 20 university students (10 dancers - mean age 20, 5 female/5 male; 10 varsity track runners - mean age = 19.5, 5 female/5 male) | USA | Ballet | Minimum 5 years dance training | Unspecified | Balance | No effect | No significant differences in balance in dancers vs track athletes | Low |
| Schmitt, Kuni & Sabo | 2005 | | No | Cross-sectional | 82 dancers & non-dancers (42 dancers, 31 female/11 male, mean age = 18.3; 40 untrained, 29 female/11 male, mean age = 19.5) | Germany | Ballet & modern dance | Minimum 7 years | 23-30 hours/weekly | Balance | Positive | Significantly better balance in dancers vs controls | Low |
| Simmons | 2005 | | No | Cross-sectional | 34 female community/university dancers & untrained | USA | Ballet | Mean 10.9 years ballet training | Unspecified | Balance | Mixed | Significantly worse balance control in dancers vs. control in some conditions; | Very Low |

| | | | | | | | | | | | | |
|---|---|---|---|---|---|---|---|---|---|---|---|---|
| | | | | individuals (mean age 21.5) | | | | | | | significantly better balance control in dancers vs controls in others | |
| Simmons | 2005 | No | Cross-sectional | 31 female community/university dancers and untrained individuals (15 dancers - mean age 21.4; 16 untrained - mean age 21.2) | USA | Ballet | mean 10.5 years ballet training | Unspecified | Balance | Positive | Significantly faster and more consistent long-latency balance responses in dancers vs controls; no significant differences between groups for short- or medium latency responses | Low |
| Thullier & Moufti | 2004 | No | Cross-sectional | 12 participants (6 elite ballet dancers, 6 gymnasts; no age or gender specified) | France | Ballet | Unspecified | Unspecified | Balance | Positive | Dancers & gymnasts equally stable | Low |
| Wyon et al. | 2013 | Excluded - single group descriptive analysis of dancers only | | | | | | | | | | |
| Zaferiou, Wilcox & McNitt-Gray | 2016 | Excluded - single group descriptive analysis of dancers only | | | | | | | | | | |
| ***Costa, Ferreira & Felicio, 2013*** | | | | | | | | | | | | |
| Barcellos & Imbiriba | 2002 | Excluded - descriptive analysis of dancers only (no comparison group) | | | | | | | | | | |
| Bruyneel et al | 2010 | Excluded - descriptive analysis of dancers only (no comparison group) | | | | | | | | | | |
| Cheng et al | 2011 | No | Cross-sectional | 51 female adolescents (26 dancers - mean age 17.5; 25 healthy active students - mean age 18.1) | Taiwan | Ballet; Modern | Mean 5.8 years dance experience | 3 hours+/weekly | Balance | Positive | Significantly improved postural stability in single leg standing condition (dominant leg; eyes open & closed) | Low |
| Denardi, Ferraciolo & Rodrigues | 2006 | Excluded - descriptive analysis of dancers only (no comparison group) | | | | | | | | | | |
| Gerbino, Griffin & Zurakowski | 2007 | Other review - Clarke et al., 2018 | | | | | | | | | | |
| Gillou, Dupu & Golomer | 2007 | No | Cross-sectional | 36 right handed & right footed male adults (7 professional dancers – mean age = 18; 9 acrobatic gymnasts – mean age | France | Ballet | Unspecified | Unspecified | Balance | Positive | Significantly reduced roll energy in dancers vs control; significantly greater spectral energy in dancers vs soccer | Low |

| | | | | | | | | | | | | |
|---|---|---|---|---|---|---|---|---|---|---|---|---|
| | | | | = 19.1; 10 professional second division soccer players – mean age = 17.1; 10 inactive control – mean age = 21.$) | | | | | | | players during left leg stance; no other significant differences (unipedal stance on unstable seesaw platform) | |
| Golomer et al | 1999 | Excluded – analysis of dancers only (multiple groups of young/older dancers but analysis of experience unclear) | | | | | | | | | | |
| Golomer et al. | 1999 | Other review - Clarke et al., 2018 | | | | | | | | | | |
| Lin et al. | 2011 | Other review - Clarke et al., 2018 | | | | | | | | | | |
| Lin, Su & Wu | 2005 | Other review - Clarke et al., 2018 | | | | | | | | | | |
| Lobo da Costa | 2012 | Excluded - descriptive analysis of dancers only (no comparison group) | | | | | | | | | | |
| Perrin et al | 2002 | Other review - Clarke et al., 2018 | | | | | | | | | | |
| Rein et al | 2011 | Other review - Clarke et al., 2018 | | | | | | | | | | |
| Schmitt, Kuni & Sabo | 2005 | Other review - Clarke et al., 2018 | | | | | | | | | | |
| Simmons | 2005 | Other review - Clarke et al., 2018 | | | | | | | | | | |
| Thiesen & Sumiya | 2011 | No | Cross-sectional | 15 female dancers (mean age = 13.8; 9 beginner; 6 intermediate) | Brazil | Ballet | Mean 31;84 months experience (beginner; intermediate) | Unspecified | Balance | No effect | No significant differences between beginner & intermediate dancers (standing oscillation speed & plantar arch structure/function) | Low |
| ***Cugusi et al, 2019*** | | | | | | | | | | | | |
| Araneta & Tanori | 2015 | No | Single group experimental study | 13 overweight women with at least 2 metabolic syndrome risk factors (mean age = 52.5) | USA | Zumba | 12 weeks | 60 min/2x weekly | Blood pressure | Positive | Significantly reduced blood pressure pre-post intervention | Very Low |
| | | | | | | | | | Triglycerides | Positive | Significantly reduced triglycerides pre-post intervention | Very Low |
| | | | | | | | | | HDL cholesterol | Unclear | Significantly decreased HDL cholesterol pre-post intervention | Very Low |
| | | | | | | | | | Body mass | No effect | No significant pre-post change | Very Low |

| | | | | | | | | | | | | |
|---|---|---|---|---|---|---|---|---|---|---|---|---|
| | | | | | | | | | Glucose | No effect | No significant pre-post change | Very Low |
| Barene et al | 2014 | Other review - Fong Yan et al., 2018 | | | | | | | | | | |
| Barene et al | 2014 | Other review - Fong Yan et al., 2018 | | | | | | | | | | |
| Cugusi et al | 2016 | No | Single group experimental study | 27 overweight women (mean age = 38.9) | Italy | Zumba | 12 weeks | 50 min/2-3x weekly | Blood pressure | Positive | Significantly reduced blood pressure pre-post Zumba intervention | Very Low |
| | | | | | | | | | % fat mass | Positive | Significantly reduced fat mass pre-post Zumba intervention | Very Low |
| | | | | | | | | | Waist circumference | Positive | Significantly reduced waist circumference pre-post Zumba intervention | Very Low |
| | | | | | | | | | Body mass | Positive | Significantly reduced body mass pre-post Zumba intervention | Very Low |
| | | | | | | | | | BMI | Positive | Significantly reduced BMI pre-post Zumba intervention | Very Low |
| Delextrat et al. | 2016 | No | RCT | 44 healthy, low active women (22 Zumba group - mean age = 26.6; 22 control – mean age = 27.9) | UK | Zumba (home DVD) | 8 weeks | 60 min/3x weekly | VO2 max | Positive | Significant improvement in VO2 max vs control | High |
| | | | | | | | | | % fat mass | No effect | No significant change vs control(DEXA) | High |
| | | | | | | | | | Body mass | No effect | No significant change vs control | High |
| Domene et al | 2016 | No | RCT | 20 overweight and sedentary women (mean age = 34) | UK | Zumba | 8 weeks | 60 min/1-2x weekly | Blood pressure | No effect | No pre-post effect of dance or control | Low |
| | | | | | | | | | Body mass | No effect | No pre-post effect of dance or control | Low |
| | | | | | | | | | % fat mass | No effect | No pre-post effect of dance or control | Low |
| | | | | | | | | | White blood cell count | Negative | Significantly reduced white blood cell count in dance group (pre-post analysis only; no comparison vs control – no significant pre-post difference in control group) | Low |

| | | | | | | | | | | | | |
|---|---|---|---|---|---|---|---|---|---|---|---|---|
| | | | | | | | | | CRP | No effect | No pre-post effect of dance or control | Low |
| | | | | | | | | | IL-6 | Unclear | Significantly reduced IL-6 in dance group (pre-post analysis only; no comparison vs control group – no significant pre-post difference in control group) | Low |
| | | | | | | | | | Total cholesterol to HDL ratio | No effect | No pre-post effect of dance or control | Low |
| | | | | | | | | | VO2 max | Positive | Significantly improved VO2 max in dance group (pre-post analysis only; no comparison vs control group – no significant pre-post difference in control group) | Low |
| | | | | | | | | | BMI | No effect | No pre-post effect of dance or control | Low |
| Donath et al | 2014 | No | RCT | 30 healthy women (mean age = 21) | Switzerland | Zumba | 8 weeks | 60 min/2x weekly | 6-minute walk test | Positive | Significantly increased 6MWT distance in dance vs control group | High |
| | | | | | | | | | BMI | Positive | Significantly reduced BMI in intervention vs control group | High |
| Krishnan et al | 2015 | No | Single group experimental study | 41 obese women (mean age = 49.3) | USA | Zumba | 16 weeks | 60min/3x weekly | Body mass | Positive | Significant pre-post reduction | Very Low |
| | | | | | | | | | BMI | Positive | Significant pre-post reduction | Very Low |
| | | | | | | | | | % body fat | Positive | Significant pre-post reduction (bioelectrical impedance) | Very Low |
| | | | | | | | | | Waist circumference | Positive | Significant pre-post reduction | Very Low |
| | | | | | | | | | Blood pressure | No effect | No significant pre-post change | Very Low |
| | | | | | | | | | Glucose | No effect | No significant pre-post change | Very Low |
| | | | | | | | | | HbA1c | No effect | No significant pre-post change | Very Low |

| | | | | | | | | | | | | | |
|---|---|---|---|---|---|---|---|---|---|---|---|---|---|
| | | | | | | | | | | Insulin | No effect | No significant pre-post change | Very Low |
| | | | | | | | | | | Total cholesterol | No effect | No significant pre-post change | Very Low |
| | | | | | | | | | | HDL cholesterol | No effect | No significant pre-post change | Very Low |
| | | | | | | | | | | Triglycerides | No effect | No significant pre-post change | Very Low |
| | | | | | | | | | | VO2 max | Positive | Significant pre-post increase (estimated from 1 mile walk time) | Very Low |
| Micallef | 2014 | | No | Single group experimental study | 36 overweight women (BMI 25+; mean age = 34.3) | Malta | Zumba | 8 weeks | 60 min/2x weekly | Body mass | Positive | Significant pre-post reduction | Very Low |
| | | | | | | | | | | BMI | Positive | Significant pre-post reduction | Very Low |
| Rossmeissl et al | 2016 | | No | Single group experimental study | 17 overweight women (median age = 54) | Switzerland | Zumba | 12 weeks | 60 min/3x weekly | VO2 max | No effect | No significant pre-post change | Very Low |
| | | | | | | | | | | Body mass | No effect | No significant pre-post change | Very Low |
| | | | | | | | | | | Waist circumference | No effect | No significant pre-post change | Very Low |
| | | | | | | | | | | BMI | No effect | No significant pre-post change | Very Low |
| | | | | | | | | | | Blood pressure | No effect | No significant pre-post change | Very Low |
| | | | | | | | | | | Resting HR | No effect | No significant pre-post change | Very Low |
| | | | | | | | | | | % fat mass | No effect | No significant pre-post change | Very Low |
| ***Fernandez-Arguelles et al., 2015*** | | | | | | | | | | | | | |
| Eyigor et al | 2009 | Other reviews - Liu, Shen & Tsai, 2020; Rodrigues-Krause, Krause & Reischak-Oliveira, 2019 | | | | | | | | | | | |
| Granacher et al | 2012 | Other reviews - Liu, Shen & Tsai, 2020; Rodrigues-Krause, Krause & Reischak-Oliveira, 2019 | | | | | | | | | | | |
| Hui, Chui & Woo | 2009 | Other reviews - Liu, Shen & Tsai, 2020; Rodrigues-Krause, Krause & Reischak-Oliveira, 2019 | | | | | | | | | | | |
| Holmerova et al | 2010 | Other review - Liu, Shen & Tsai, 2020 | | | | | | | | | | | |
| Shigematsu et al | 2002 | Excluded – intensity of dance-based exercise program set at individual‘s lactate threshold (exercise, not performing arts, intervention) | | | | | | | | | | | |
| Sofianidis et al | 2009 | Other reviews - Liu, Shen & Tsai, 2020; Fong Yan et al., 2018 | | | | | | | | | | | |

| | | | | | | | | | | | | | |
|---|---|---|---|---|---|---|---|---|---|---|---|---|---|
| Young, Weeks & Beck | 2007 | Excluded - dance a small part of a broader fitness program (foot stamping, progressively loaded squats & line dancing) | | | | | | | | | | | |
| ***Fong Yan et al., 2018*** | | | | | | | | | | | | | |
| Arzglou et al. | 2013 | Excluded - clinical population (Parkinson's), narratively analyzed outcomes only (postural stability) | | | | | | | | | | | |
| Barene et al. | 2016 | | No | RCT | 72 female hospital employees (mean age = 45; 35 Zumba group, 37 soccer group) | Norway | Zumba | 40 weeks | 1 hour/2x week | Sit & Reach | Meta-analysis | | Moderate |
| | | | | | | | | | | Postural stability | Positive | Significantly greater improvement in postural stability in Zumba vs. soccer groups | Moderate |
| Barene et al. | 2014 | | No | RCT | 72 female hospital employees (mean age = 45; 35 Zumba group, 37 soccer group) | Norway | Zumba | 40 weeks | 1 hour/2x week | Fat mass | Meta-analysis | | Moderate |
| | | | | | | | | | | VO2 max | Meta-analysis | | Moderate |
| | | | | | | | | | | Triglycerides | Meta-analysis | | Moderate |
| | | | | | | | | | | Glucose | No effect | No significant intergroup or pre-post changes | Moderate |
| | | | | | | | | | | Peak ventilation | Meta-analysis | | Moderate |
| | | | | | | | | | | Blood pressure | No effect | No significant intergroup or pre-post changes | Moderate |
| | | | | | | | | | | Respiratory exchange ratio | No effect | No significant intergroup or pre-post changes | Moderate |
| | | | | | | | | | | Total cholesterol | Meta-analysis | | Moderate |
| | | | | | | | | | | Bone mineral density | No effect | No significant intergroup or pre-post changes | Moderate |
| | | | | | | | | | | HDL cholesterol | Meta-analysis | | Moderate |
| | | | | | | | | | | Body mass | Meta-analysis | | Moderate |
| | | | | | | | | | | BMI | Meta-analysis | | Moderate |
| Barene, Krustrup & Holtermann | 2014 | Excluded - unclear outcomes ('questionnaire', but not one listed in review table 3) | | | | | | | | | | | |
| Belardinelli et al. | 2008 | Clinical population, but included in meta-analysis | No | RCT | 88 stable chronic heart failure patients (74 male/14 female; mean age = 59.5; 44 dance group, 44 cardio exercise group) | Italy | Waltz | 8 weeks | 21 min/3x weekly | VO2 max | Meta-analysis | | High |
| | | | | | | | | | | Triglycerides | Meta-analysis | | High |
| | | | | | | | | | | Total cholesterol | Meta-analysis | | High |
| | | | | | | | | | | HDL cholesterol | Meta-analysis | | High |

| | | | | | | | | | | | | | |
|---|---|---|---|---|---|---|---|---|---|---|---|---|---|
| | | Excluded - clinical population (chronic heart failure) | | | | | | | | Glucose; Peak O2 pulse; Respiratory exchange ratio; VE/VCO2; VO2 at anaerobic threshold | | | |
| Burgess, Grogan & Burwitz | 2006 | | No | RCT | 50 healthy British schoolgirls (mean age = 13.5; 25 dance group, 25 physical education) | UK | Aerobic dance | 6 weeks | 50 min/2x weekly | BMI | Meta-analysis | | Moderate |
| Ford et al. | 1989 | | No | Longitudinal cohort study | 88 healthy female university students (mean age = 19.8; 21 aerobic dance group, 17 jogging group, 22 progressive resistance training, 15 swimming group, 13 sport training) | USA | Aerobic dance | 8 weeks | 1 hour/3x weekly | Sit & reach | Meta-analysis | | Low |
| | | | | | | | | | | Sit ups | Positive | Significant pre-post improvement vs control in aerobic dance but not weight training group | Low |
| | | | | | | | | | | Step test | No effect | | Low |
| | | | | | | | | | | Fat mass | Meta-analysis | (skinfold measurements) | Very Low |
| Garber, McKinney & Carleton | 1992 | | No | RCT | 25 university employees (12 male/13 female; mean age = ~37; 14 aerobic dance group, 11 cardio group) | USA | Aerobic dance | 8 weeks | 50 min/3x weekly | VO2 max | Meta-analysis | | Moderate |
| | | | | | | | | | | Peak ventilation | Meta-analysis | | Moderate |
| | | | | | | | | | | Respiratory exchange ratio | No effect | No significant intergroup or pre-post changes | Moderate |
| | | | | | | | | | | Body mass | Meta-analysis | | Moderate |
| | | | | | | | | | | BMI | Meta-analysis | | Moderate |
| Hackney, Kantorovich & Earhart | 2007 | Excluded - clinical population (Parkinson's), narratively analyzed outcomes only (gait velocity, postural stability) | | | | | | | | | | | |
| Hackney et al. | 2007 | Clinical population, but included in meta-analysis | No | RCT | 19 idiopathic Parkinson's disease patients & matched controls (12 male/7 female; mean age = ~71; 9 Latin dance group, 10 progressive resistance training group) | USA | Latin dance | 13 weeks | 1 hour/20 classes over 13 weeks | Timed up and go | Meta-analysis | | High |
| | | | | | | | | | | Berg balance test | Meta-analysis | | High |
| | | Excluded - clinical population (Parkinson's) | | | | | | | | Gait velocity | | | |
| Hackney & Earhart | 2009 | Excluded - clinical population (Parkinson's); narratively analyzed outcomes only (questionnaire) | | | | | | | | | | | |
| Hashimoto et al. | 2015 | Clinical population, but included in | No | RCT | 32 Parkinson's disease patients (15 aerobic dance group - 3 male/12 females, | Japan | Aerobic dance | 12 weeks | 1 hour/weekly | Timed up and go | Meta-analysis | | High |
| | | | | | | | | | | Berg balance test | Meta-analysis | | High |

| | | | | | | | | | | | | |
|---|---|---|---|---|---|---|---|---|---|---|---|---|
| | | meta-analysis | | | mean age = 67.9; 17 progressive resistance training group - 2 male/15 female, mean age = 62.7) | | | | | | | |
| Heffron, Davey & Cochrane | 1997 | | No | Non-randomized experimental cohort study | 29 postmenopausal women (mean age = 58; 14 aerobic dance group, 15 progressive resistance training group) | UK | Aerobic dance | 10 weeks | 1 hour/weekly | Bone mineral density | No effect | No significant intergroup or pre-post changes | Low |
| Janyacharoen et al | 2013 | Other review - Liu, Shen & Tsai, 2020 (included in meta-analysis) | No | RCT | 38 healthy older women (20 folk/traditional dance group - mean age = 64.9; 18 physical education group - mean age = 66.8) | Thailand | Folk/traditional dance | 6 weeks | 40 min/3x weekly | Sit & Reach | Meta-analysis | | High |
| | | Other review - Liu, Shen & Tsai, 2020 | | | | | | | | 6-minute walk test; sit to stand | | | |
| Kalsatou et al. | 2014 | Clinical population, but included in meta-analysis | No | RCT | 34 Greek males with chronic heart failure (mean age = 67; 18 folk/traditional dance group, 16 cardio & progressive resistance training group) | Greece | Greek folk/traditional dance | 8 months | 1 hour/3x weekly | VO2 max | Meta-analysis | | High |
| | | | | | | | | | | Peak ventilation | Meta-analysis | | High |
| | | | | | | | | | | Berg Balance test | Meta-analysis | | High |
| | | | | | | | | | | Body mass | Meta-analysis | | High |
| | | | | | | | | | | BMI | Meta-analysis | | High |
| | | Excluded - clinical population (chronic heart failure) | | | | | | | | Blood pressure; Peak O2 pulse; Sit to stand; VE/VCO2; VO2 at anaerobic threshold | | | |
| Kin Isler, Kosar & Korkusuz | 2001 | Clinical population, but included in meta-analysis | No | RCT | 30 sedentary female college students (mean age = 21.1, 15 aerobic dance group, 15 cardio group) | Turkey | Aerobic dance | 8 weeks | 45 min/3x weekly | Triglycerides | Meta-analysis | | High |
| | | | | | | | | | | Body weight | Meta-analysis | | High |
| | | | | | | | | | | Total cholesterol | Meta-analysis | | High |
| | | | | | | | | | | HDL cholesterol | Meta-analysis | | High |
| Kouli et al | 2009 | | No | Non-randomized experimental study | 57 5th graders (mean age = 10.6; 33 aerobic dance group, 24 physical education group), sex unspecified | Greece | Aerobic dance | 8 weeks | 45 min/3x weekly | Range of motion | Positive | Significantly greater improvement in dance vs. physical education group | Low |
| | | | | | | | | | | Sit ups | Positive | Significantly greater improvement in dance vs. physical education group | Low |

| | | | | | | | | | | | | |
|---|---|---|---|---|---|---|---|---|---|---|---|---|
| | | | | | | | | | 1-mile walk/run | Positive | Significantly greater improvement in dance vs. physical education group | Low |
| | | | | | | | | | Sit & reach (each leg separated) | Positive | Significantly greater improvement in dance vs. physical education group | Low |
| | | | | | | | | | Push ups | Positive | Significantly greater improvement in dance vs. physical education group | Low |
| Mangeri et al. | 2014 | No | Non-randomized experimental study | 100 Type 2 diabetic/obese adults (42 ballroom dance group - mean age = 58.5, 18 male/24 female; 58 cardio group - mean age = 59.4, 34 male/24 female) | Italy | Ballroom dance | 6 months | 2 hours/2x weekly | Blood pressure | No effect | No significant intergroup or pre-post changes | Low |
| | | | | | | | | | Glucose | No effect | No significant intergroup or pre-post changes | Low |
| | | | | | | | | | 6-minute walk test | Positive | Significantly greater improvement in dance vs. cardio group | Low |
| | | | | | | | | | Triglycerides | Meta-analysis | | Low |
| | | | | | | | | | Body mass | Meta-analysis | | Low |
| | | | | | | | | | BMI | Meta-analysis | | Low |
| | | | | | | | | | Waist circumference | Positive | Significant pre-post improvement in dance & cardio groups | Low |
| | | | | | | | | | Total cholesterol | Meta-analysis | | Low |
| | | | | | | | | | HDL cholesterol | Meta-analysis | | Low |
| Mavridis et al. | 2004 | No | Non-randomized experimental study | 40 1st grade children (mean age = 6.6; 21 aerobic dance group, 19 physical education), sex unspecified | Greece | Aerobic dance | 12 weeks | 45 min/3x weekly | Range of motion | Positive | Significantly greater improvement in dance vs. physical education group | Low |
| | | | | | | | | | Sit ups | Positive | Significantly greater improvement in dance vs. physical education group | Low |
| | | | | | | | | | 1-mile walk/run | Positive | Significant pre-post improvement in dance & physical education groups | Low |

| | | | | | | | | | | | | | |
|---|---|---|---|---|---|---|---|---|---|---|---|---|---|
| | | | | | | | | | | Sit & reach (each leg separated) | Positive | Significantly greater improvement in dance vs. physical education group | Low |
| | | | | | | | | | | Push ups | Positive | Significantly greater improvement in dance vs. physical education group | Low |
| | | | | | | | | | | Skinfold thickness | Positive | Significantly greater improvement in dance vs. physical education group | Low |
| McKinley et al. | 2008 | Other review - Liu, Shen & Tsai, 2020; narratively synthesized outcomes only (Gait velocity, sit to stand) | | | | | | | | | | | |
| Milburn & Butts | 1983 | Dance with target HR, but included in meta-analysis | No | Non-randomized experimental study | 34 female college students (15 aerobic dance group - mean age = 21.4; 19 jogging group - mean age = 19) | USA | Aerobic dance | 7 weeks | 30 min/4x weekly | VO2 max | Meta-analysis | | Low |
| | | | | | | | | | | Peak ventilation | Meta-analysis | | Low |
| Rani & Singh | 2013 | | No | RCT | 75 college students (unspecified age & gender; aerobic dance & yoga groups) | India | Aerobic dance | 6 weeks | 30-60 min/3x weekly | Sit & reach | Meta-analysis | | Moderate |
| | | | | | | | | | | Sit ups | Positive | Significantly greater improvement in dance vs. yoga group | Moderate |
| | | | | | | | | | | 1-mile walk/run | Positive | Significantly greater improvement in dance vs. yoga group | Moderate |
| | | | | | | | | | | Skinfold thickness | Positive | Significant pre-post improvement in dance & yoga groups | Moderate |
| Rehfeld et al. | 2017 | Other review - Teixeira-Machado, Arida & de Jesus Mari, 2019, narratively analyzed outcomes only (balance) | | | | | | | | | | | |
| Rios Romenets et al | 2015 | Clinical population, but included in meta-analysis | No | RCT | 33 Parkinson's disease patients (18 Latin dance group - mean age = 63.2, 12 male/6 female; 15 physiotherapy exercises - mean age = 64.3, 7 male/8 female) | Canada | Latin | 12 weeks | 1 hour/2x weekly | Timed up and go | Meta-analysis | | High |
| Shimamoto et al. | 1998 | | No | | | Japan | Aerobic dance | 12 weeks | | VO2 max | Meta-analysis | | Low |

| | | | | | | | | | | | | | |
|---|---|---|---|---|---|---|---|---|---|---|---|---|---|
| | | | | Non-randomized experimental study | 60 moderately obese Japanese women (37 aerobic dance group - mean age = 49.1; 23 cardio group - mean age = 51.7) | | | | 1 hour/2-3x weekly | VO2 at anaerobic threshold | Positive | Significant pre-post improvement in dance & cardio groups | Low |
| | | | | | | | | | | Body mass | Meta-analysis | | Low |
| | | | | | | | | | | BMI | Meta-analysis | | Low |
| | | | | | | | | | | % fat mass | Meta-analysis | | Low |
| | | | | | | | | | | Waist circumference | Positive | Significant pre-post improvement in dance & cardio groups | Low |
| | | | | | | | | | | Skinfold thickness | Positive | Significant pre-post improvement in dance & cardio groups | Low |
| Sofianidis, Dimitriou & Hatzitaki | 2017 | | No | RCT | 24 older adults (7 male/17 female, mean age = 70.7, 12 Latin dance group/12 pilates group) | Greece | Latin | 12 weeks | 1 hour/2x weekly | Postural stability | Positive | Significant pre-post improvement in dance & pilates groups | High |
| Viskic et al | 2007 | | No | Non-randomized experimental study | 220 3rd & 4th grade girls (115 aerobic dance group, 105 physical education group; unspecified age) | Croatia | Aerobic dance | 60 high school periods | Unspecified | Sit & reach | Meta-analysis | | Low |
| | | | | | | | | | | Range of motion | Positive | Significantly greater improvement in dance vs. physical education group | Low |
| | | | | | | | | | | Step test | Positive | Significantly greater improvement in dance vs. physical education group | Low |
| | | | | | | | | | | Body mass | Meta-analysis | | Low |
| | | | | | | | | | | % fat mass | Meta-analysis | | Low |
| Volpe et al. | 2013 | Clinical population, but included in meta-analysis | No | RCT | 24 Parkinson's disease patients (mean age = 63.3; 13 male/11 female; 12 dance group/12 physiotherapy exercise group) | Italy | Folk/traditional dance | 26 weeks | 90 min/weekly | Timed up and go | Meta-analysis | | High |
| | | | | | | | | | | Berg balance test | Meta-analysis | | High |
| ***Hwang & Braun, 2015*** | | | | | | | | | | | | | |
| Alpert et al. | 2009 | Other review - Rodrigues-Krause, Krause & Reischak-Oliveira, 2019 | | | | | | | | | | | |
| Borges et al | 2012 | Other review - Rodrigues-Krause, Krause & Reischak-Oliveira, 2019 | | | | | | | | | | | |

| | | | | | | | | | | | |
|---|---|---|---|---|---|---|---|---|---|---|---|
| Coubard et al | 2011 | Other review - Meng et al., 2020 | | | | | | | | | |
| Eyigor et al. | 2009 | Other reviews - Liu, Shen & Tsai, 2020; Rodrigues-Krause, Krause & Reischak-Oliveira, 2019 | | | | | | | | | |
| Ferrufino et al. | 2011 | No | Non-randomized experimental study | 41 healthy older adults (16 dance group - 16 female/0 male, mean age = 73.7; 25 falls prevention training group - 23 female/2 male, mean age = 72.9) | France | Contemporary | 4.4 months (average) | 1 hour/weekly | Balance | Positive | Significant pre-post improvement in balance performance (superior to falls prevention group) in balance performance (posturography - TechnoConcept platform) | Low |
| Granacher et al | 2012 | Other reviews - Liu, Shen & Tsai, 2020; Rodrigues-Krause, Krause & Reischak-Oliveira, 2019 | | | | | | | | | |
| Hackney et al. | 2013 | Excluded - investigation of clinical population (visual impairment) | | | | | | | | | |
| Holmerova et al | 2010 | Other review - Liu, Shen & Tsai, 2020 meta-analysis; excluded from further narrative analysis due to clinical population (functional impairment) | | | | | | | | | |
| Hui, Chui & Woo | 2008 | Other reviews - Liu, Shen & Tsai, 2020; Rodrigues-Krause, Krause & Reischak-Oliveira, 2019 | | | | | | | | | |
| Kim et al | 2011 | Excluded - Latin dance performed with a target heart rate (exercise, not performing arts, intervention) | | | | | | | | | |
| Krampe | 2013 | Excluded - clinical population (90% of cohort with at least 1 chronic illness) | | | | | | | | | |
| Krampe et al | 2010 | Other review - Rodrigues-Krause, Krause & Reischak-Oliveira, 2019 | | | | | | | | | |
| Marmeleira et al | 2009 | No | RCT | 37 inactive older adults (19 dance group - 4 male/15 female, mean age = 63.6; 18 control group - 5 male/10 female, mean age = 65.3) | Portugal | Creative dance | 12 weeks | 90 min/3x weekly | Proprioception | Positive | Significant improvements in arm positioning vs control; significant pre-post improvements in knee proprioception but not vs control (JPS & kinesthesia - knee & upper limb) | High |
| McKinley et al | 2008 | Other reviews - Liu, Shen & Tsai, 2020; Rodrigues-Krause, Krause & Reischak-Oliveira, 2019 | | | | | | | | | |
| Sofianidis et al | 2009 | Other review - Liu, Shen & Tsai, 2020 | | | | | | | | | |
| Song et al. | 2004 | Excluded - Korean dance intervention with a target heart rate (exercise, not performing arts, initiative) | | | | | | | | | |
| Wallmann | 2008 | Other review - Rodrigues-Krause, Krause & Reischak-Oliveira, 2019 | | | | | | | | | |
| Young, Weeks & Beck | 2007 | Excluded - dance a small part of a broader fitness program (foot stamping, progressively loaded squats & line dancing) | | | | | | | | | |

| | | | | | | | | | | | | |
|---|---|---|---|---|---|---|---|---|---|---|---|---|
| ***Keogh et al., 2009*** | | | | | | | | | | | | |
| Engels et al | 1998 | Excluded - investigation of aerobic dance performed with target HR of 50-70% maximum with participants instructed to monitor HR throughout (exercise, not performing arts, activity)** | | | | | | | | | | |
| Eyigor et al | 2009 | Other reviews - Liu, Shen & Tsai, 2020; Rodrigues-Krause, Krause & Reischak-Oliveira, 2019 | | | | | | | | | | |
| Federici, Bellagamba & Rocchi | 2005 | Other review - Rodrigues-Krause, Krause & Reischak-Oliveira, 2019 | | | | | | | | | | |
| Hackney, Kantorovich & Earhart | 2007 | Excluded - investigation of an intervention designed for clinical population (Parkinson's), but delivered jointly to a healthy/clinical cohort | | | | | | | | | | |
| Holmerova et al | Personal Communi cation | Other review - Liu, Shen & Tsai, 2020 meta-analysis; excluded from further narrative analysis due to clinical population (functional impairment) | | | | | | | | | | |
| Hopkins et al | 1990 | Excluded - investigation of aerobic dance performed with target HR of 100-120 beats per minute with participants instructed to monitor HR throughout (exercise, not performing arts, activity) | | | | | | | | | | |
| Jeon et al | 2005 | Excluded - Korean language article, very similar intervention to Jeon et al 2000 (exercise, not performing arts) | | | | | | | | | | |
| Jeon et al | 2000 | Excluded - Korean dance intervention with a target heart rate (exercise, not performing arts, intervention) | | | | | | | | | | |
| Kim, June & Song | 2003 | Excluded - combined analysis of dance within context of 'health promotion program' including health education classes and counseling sessions | | | | | | | | | | |
| Kudlacek et al | 1997 | No | Single group experimental study | 28 female members of a senior dancing group (mean age 66; 15 osteoporotic, 13 non-osteoporotic) | Austria | Folk, waltz, aerobic | 12 months | Unspecified | Bone density (peripheral and spinal) | Mixed (positive, no effect) | Significant pre-post increase in bone density in osteoporotic participants but not in participants with non-clinical bone density (single photon absorptiometry - X-Ray) | Very Low |
| McKinley et al | 2008 | Other reviews - Liu, Shen & Tsai, 2020; Rodrigues-Krause, Krause & Reischak-Oliveira, 2019 | | | | | | | | | | |
| Shigematsu et al | 2002 | Excluded – intensity of dance-based exercise program set at individual‘s lactate threshold (exercise, not performing arts, intervention) | | | | | | | | | | |
| Sofianidis et al | 2009 | Other reviews - Liu, Shen & Tsai, 2020; Fong Yan et al., 2018 | | | | | | | | | | |
| Song et al | 2004 | Excluded - Korean dance intervention with a target heart rate (exercise, not performing arts, initiative) | | | | | | | | | | |
| Young, Weeks & Beck | 2007 | Excluded - dance a small part of a broader fitness program (foot stamping, progressively loaded squats & line dancing | | | | | | | | | | |
| ***Letton, Thom, & Ward, 2020*** | | | | | | | | | | | | |

| | | | | | | | | | | | | |
|---|---|---|---|---|---|---|---|---|---|---|---|---|
| Annino et al. | 2007 | Excluded - study of whole body vibration in full time professional ballerinas | | | | | | | | | | |
| Candelario-Gorbea et al | 2014 | Excluded - study of PNF stretching in students at a ballet school (mean >3 years training) | | | | | | | | | | |
| da Silva et al | 2015 | Excluded - study of 17-week ballet training intervention in preprofessional ballet dancers (5 years experience) | | | | | | | | | | |
| Hedgepeth | 1987 | No | Cross-sectional (**analysis of the 'pre' data in dancers vs control only; no analysis of intervention results in trained dancers as per exclusion criteria)** | 27 female university students (age range 18-35; 18 ballet students (min 2 years experience); 9 sedentary individuals) | USA | Ballet | 2+ years ballet experience | Unspecified | Strength | No effect | No significant differences between dancers & control (peak knee extension torque - Cybex) | Low |
| | | | | | | | | | Power | No effect | No significant differences between dancers & control ((total work / contraction time) of 30 maximal isokinetic knee extensions - Cybex) | Low |
| | | | | | | | | | Endurance (muscular) | No effect | No significant differences between dancers & control ((total work 1st 6 trials / total work last 6 trials) from 30 maximal isokinetic knee extensions - Cybex) | Low |
| Houston & McGill | 2013 | Excluded - clinical population (Parkinson's) | | | | | | | | | | |
| Khan et al. | 2000 | Excluded - longitudinal study of ballet training in elite ballet dancers | | | | | | | | | | |
| Kirkendall et al. | 1984 | Excluded - longitudinal study of ballet training in professional ballet dancers | | | | | | | | | | |
| Koutedakis & Sharp | 2004 | Excluded – longitudinal study of dance intervention in professional ballerinas | | | | | | | | | | |
| Koutedakis, Cross & Sharp | 1996 | Excluded - longitudinal study of strength training in professional male ballet dancers | | | | | | | | | | |
| Kujala et al | 1997 | No | Cross-sectional (**consideration of pre- and post-intervention results in experienced dancers vs | 35 female adolescents (mean age = 11.9; 18 ballet dancers, 17 non-athletic controls) | Finland | Ballet | 2+ years ballet experience | Mean 7.25 - 11.1 hours weekly training | Flexibility | Positive | Significantly greater hip flexor & hamstring flexibility in dancers vs controls; conclusion that additional lumbar flexibility likely not possible through | Low |

| | | | | | | | | | | | | |
|---|---|---|---|---|---|---|---|---|---|---|---|---|
| | | | controls as cross-sectional outcomes**) | | | | | | | | training (hip flexor, hamstring & lumbar flexibility measured by goniometer) | |
| Liederbach, Gleim & Nicholas | 1992 | Excluded - study of ballet training in professional dancers | | | | | | | | | | |
| Lopez-Ortiz et al | 2016 | Excluded - clinical population (cerebral palsy) | | | | | | | | | | |
| Lopez-Ortiz et al | 2012 | Excluded - clinical population (cerebral palsy) | | | | | | | | | | |
| McGill, Houston & Lee | 2019 | Excluded - clinical population (Parkinson's) | | | | | | | | | | |
| McMillan, Proteau & Lebe | 2019 | Excluded - study of pilates intervention in elite ballet dancers | | | | | | | | | | |
| Micheli et al | 2005 | Excluded - study of ballet training in professional dancers | | | | | | | | | | |
| Moller & Masharawi | 2011 | No | Non-randomized experimental study | 30 1st-3rd grade girls (age range 6-9; 15 ballet group, 15 control group | Israel | Ballet | 6 months | 90 min/2x weekly | Flexibility | Positive | Significantly greater hyper flexibility and hip external rotation in dance vs control groups (hip external rotation (goniometer); Beighton score (hyperflexibility) | Low |
| | | | | | | | | | Body composition | No effect | No significant differences between groups (BMI) | Low |
| | | | | | | | | | Posture | No effect | No significant difference between dance & control groups (thoracic kyphosis; lumbar lordosis) | Low |
| Notarnicola et al. | 2014 | Excluded - study of pedagogical methods in experienced ballet dancers (mean 4+ years experience) | | | | | | | | | | |
| Ozdinc & Turan | 2016 | No | Cross-sectional | 67 young females (mean age ~11; 36 dancers (mean 4+ years dancing experience); 31 non-dancer controls) | Turkey | Ballet | Mean 4.4 years ballet experience | Unspecified | Foot anthropometrics | No effect | No significant differences between dance & control groups on any anthropometric measurement | Low |
| Pigeon et al | 1997 | No | Longitudinal cohort study | 127 young girls (mean age 12.6) | France | Ballet | 5 years | Mean 8.8 hours weekly ballet practice | Growth | Mixed (no effect, negative) | Significantly stunted growth in subgroup of | Very Low |

| | | | | | | | | | | | | | |
|---|---|---|---|---|---|---|---|---|---|---|---|---|---|
| | | | | | | | | | | | | dancers with comorbid insufficient nutrition | |
| | | | | | | | | | | Puberty onset | Negative | Later pubertal onset in dancers vs control | Very Low |
| Scheidler et al. | 2018 | Excluded - clinical population (multiple sclerosis) | | | | | | | | | | | |
| Stalder, Noble & Wilkinson | 1990 | Excluded - study of weight training in experienced ballet dancers | | | | | | | | | | | |
| Tsanaka, Manou & Kellis | 2017 | Excluded - study of pedagogy in college ballet dancers | | | | | | | | | | | |
| ***Liu, Shen & Tsai, 2020*** | | | | | | | | | | | | | |
| Bennett & Hackney | 2017 | | No | RCT | 23 adults with mobility limitations (20 female/3 male; 12 intervention, 11 control; aged 65 or over) | USA | Line dance | 8 weeks | 1 hour/2x weekly | Balance | Positive | Significant improvement in intervention vs control (Berg Balance Scale) | Moderate |
| | | | | | | | | | | Gait speed | Meta-analysis | | Moderate |
| Cepeda et al | 2015 | | No | RCT | 34 female older adults (19 intervention (mean age = 69.1); 15 control (mean age = 71.5)) | Brazil | Ballroom | 8 weeks | 1 hour/3x weekly | Endurance | Meta-analysis | | High |
| | | | | | | | | | | Mobility | Meta-analysis | | High |
| Eyigor et al. | 2009 | | No | RCT | 37 female older adults (19 intervention (mean age 73.5); 18 control (mean age = 71.2)) | Turkey | Turkish folkloric dance | 8 weeks | 1 hour/3x weekly | Balance | Positive | Significant improvement in intervention vs control (Berg Balance Scale) | High |
| | | | | | | | | | | Endurance | Meta-analysis | | High |
| | | | | | | | | | | Mobility | Meta-analysis | | High |
| | | | | | | | | | | General health | Mixed | Significant improvements vs control on a few subscales only (mental health, general health, physical health)(SF-36) | High |
| Granacher et al. | 2012 | | No | RCT | 28 older adults (17 female/11 male; 14 intervention (mean age = 71.6); 14 control (mean age = 68.9)) | Germany | Salsa | 8 weeks | 1 hour/2x weekly | Balance | No effect | No significant difference vs control (center of pressure - dominant leg) | High |
| | | | | | | | | | | Gait speed | Meta-analysis | | High |

| | | | | | | | | | | | | |
|---|---|---|---|---|---|---|---|---|---|---|---|---|
| Hamacher et al. | 2015 | No | RCT | 35 older adults (19 intervention - 11 female, mean age = 67.2; 16 control - 10 female, mean age = 68.5) | Germany | Multiple (Line, Jazz, Square) | 6 months | 90 min/2x weekly | Gait speed | Meta-analysis | | High |
| Holmerova et al. | 2009 | No | RCT | 52 older adults (27 intervention - 25 female/2 male, mean age = 81; 25 control - 21 female/4 male, mean age = 82.8) | Czech Republic | Ballroom | 3 months | 75 min/1x weekly | Flexibility | Meta-analysis | | High |
| | | | | | | | | | Mobility | Meta-analysis | | High |
| Hui, Chui & Woo | 2009 | No | RCT | 97 older adults (52 intervention - 50 female/2 male, mean age = 68; 45 control - 44 female/1 male, mean age = 69.1) | Hong Kong | Low impact aerobic dance | 3 months | 50 min/2x weekly | Endurance | Meta-analysis | | Moderate |
| | | | | | | | | | Flexibility | Meta-analysis | | Moderate |
| | | | | | | | | | Mobility | Meta-analysis | | Moderate |
| | | | | | | | | | General health | Mixed | Significant improvements vs control on a few subscales only (general health, bodily pain)(SF-36) | Moderate |
| Janyacharoen et al. | 2013 | No | RCT | 38 female older adults (20 intervention (mean age = 64.9); 18 control (mean age = 66.8)) | Thailand | Thai dance | 6 weeks | 40 min/3x weekly | Endurance | Meta-analysis | | High |
| | | | | | | | | | Flexibility | Meta-analysis | | High |
| | | | | | | | | | Mobility | Meta-analysis | | High |
| Machacova et al. | 2015 | No | RCT | 52 older adults (27 intervention - 25 female/2 male, mean age = 81; 25 control - 21 female/4 male, mean age = 82.8) | Czech Republic | Ballroom | 3 months | 75 min/1x weekly | Flexibility | **same study as Holmerova et al 2009** | | High |
| | | | | | | | | | Mobility | Meta-analysis | | High |
| McKinley et al. | 2008 | No | RCT | 25 older adults (14 intervention - 11 female/3 male, mean age = 78.1; 11 control - 8 female/3 male, mean age = 74.6) | Canada | Argentine tango dance | 10 weeks | 2 hours/2x weekly | Mobility | Meta-analysis | | High |
| | | | | | | | | | Gait speed | Meta-analysis | | High |
| Merom et al. | 2016 | No | RCT | 530 older adults aged at or over 65 (279 intervention - 231 female/38 male; 251 | Australia | Folk dance | 12 months | 1 hour/2x weekly | Gait speed | Meta-analysis | | Moderate |
| | | | | | | | | | Mobility | Meta-analysis | | Moderate |

| | | | | | | | | | | | | |
|---|---|---|---|---|---|---|---|---|---|---|---|---|
| | | | | control - 217 female/34 control) | | | | | | | | |
| Sofianidis et al. | 2009 | No | RCT | 26 older adults (14 intervention - 13 female/1 male, mean age = 69.2; 12 control - 7 female/5 male, mean age = 72.6) | Greece | Greek traditional dance | 10 weeks | 1 hour/2x weekly | Balance | Positive | Significantly improved balance in intervention group vs control (COP variations) | High |
| Serrano-Guzman et al. | 2016 | No | RCT | 52 female adults (27 intervention (mean age = 69.1); 25 control (mean age = 69.5)) | Spain | Multiple (Flamenco, Sevillanas, ballet) | 8 weeks | 50 min/3x weekly | Mobility | Meta-analysis | | Moderate |
| | | | | | | ***Meng et al., 2020*** | | | | | | |
| Alves | 2013 | No | RCT | 65 healthy older adults (25 dance group - 4 male/21 female, mean age = 69.4; 15 walking group - 15 female, mean age = 66.7; 25 'no contact' control - 1 male/24 female, mean age = 68.4) | Brazil | Ballroom | 4 months | 2 hours/2x weekly | Executive function | Meta-analysis | | High |
| Chen | 2014 | | RCT | 125 older adults (age range 61-82; 65 dance group, 60 control group) | China | Square dancing | 48 weeks | 30-60 min/4-5x weekly | Global cognition | Meta-analysis | | High |
| Coubard et al | 2011 | No | Non-randomized experimental study | 110 healthy older adults (16 dance group - 16 female/0 male, mean age = 74.7; 67 falls prevention training group - 64 female/3 male, mean age = 74.7; 27 Tai Chi group - 24 female/3 male, mean age = 71.5) | France | Contemporary | 5.9 months (mean) | 1 hour/weekly | Executive function | Meta-analysis | | Low |
| Doi et al | 2017 | No | RCT | | Japan | Ballroom | 40 weeks | 1 hour/weekly | Global cognition | Meta-analysis | | High |

| | | | | | | | | | | | | |
|---|---|---|---|---|---|---|---|---|---|---|---|---|
| | | Clinical population, but included in meta-analysis | | | 201 Japanese adults with mild cognitive impairment (67 dance group - 51% female, mean age = 75.7; 67 music training group - 58% female, mean age = 76.2; 67 health education control group - 46% female, mean age = 76) | | | | | Executive function | Meta-analysis | High |
| | | Excluded - clinical population (cognitive impairment) | | | | | | | | Memory | | |
| Hackney et al | 2015 | | No | Non-randomized experimental study | 74 healthy residents of independent living facilities (62 tango group - 14 male/48 female, mean age = 82.3; 12 health education control group - mean age = 84.1, 7 male/5 female) | USA | Tango | 12 weeks | 90 min / 20 classes in 12 weeks | Global cognition | Meta-analysis | Very Low |
| | | | | | | | | | | Executive function | Meta-analysis | Very Low |
| Kattenstroth et al | 2013 | | No | RCT | 35 healthy elderly volunteers (25 dance group - mean age = 68.6, 17 female/8 male; 10 control - mean age = 72.3, 7 female/3 male) | Germany | Agilando (dance program developed for elderly people; no partner necessary) | 24 weeks | 1 hour/weekly | Global cognition | Meta-analysis | Moderate |
| Kim et al | 2011 | Study of Latin dance intervention with target heart rate, but included in meta-analysis | No | Non-randomized experimental study | 38 older adults with metabolic syndrome (26 exercise - 7 male/19 female, mean age = 68.2; 12 control - 2 male/10 female, mean age = 68.2) | South Korea | Latin | 6 months | 1 hour/2x weekly | Global cognition | Meta-analysis | Low |
| | | | | | | | | | | Executive function | Meta-analysis | Low |
| | | Excluded - Latin dance performed with target heart rate (exercise, not performing arts) | | | | | | | | Memory | | |
| Kosmat & Vranic | 2017 | | No | RCT | 24 healthy older adults (12 dance - 6 | Croatia | Waltz | 10 weeks | 45 min/weekly | Executive function | Meta-analysis | High |

| | | | | | | | | | | | | | |
|---|---|---|---|---|---|---|---|---|---|---|---|---|---|
| | | | | | male/6 female, mean age = 80.1; 12 active control - 9 female/3 male, mean age = 79.1) | | | | | Memory | Positive | Significant group*time effect, with significant benefits for memory in dance vs active control (auditory verbal learning test) | High |
| Lazarou et al | 2017 | Clinical population, but included in meta-analysis | No | RCT | 129 older adults with amnesiatic mild cognitive impairment (66 dance group - 53 female/13 male, mean age = 65.9; 63 control group - 48 female/15 male, mean age = 67.9) | Greece | Ballroom (International) | 10 month | 1 hour/2x weekly | Global cognition | Meta-analysis | | High |
| | | | | | | | | | | Executive function | Meta-analysis | | High |
| | | Excluded - clinical population (cognitive impairment) | | | | | | | | Memory | | | |
| Merom et al | 2016 | | No | RCT | 115 healthy older adults (aged 60+; 60 dance group - 73% female; 55 walk group - 80% female) | Australia | Ballroom | 8 months | 1 hour/2x weekly | Executive function | Meta-analysis | | Moderate |
| | | | | | | | | | | Memory | No effect | Significant pre-post effect only, no effect vs control (Rey Auditory Verbal Learning Test; Brief Visuospatial Memory Test) | Moderate |
| Merom et al | 2016 | | No | RCT | 530 older adults aged at or over 65 (279 intervention - 231 female/38 male; 251 control - 217 female/34 control) | Australia | Folk dance | 12 months | 1 hour/2x weekly | Executive function | Meta-analysis | | Moderate |
| Muller et al | 2017 | Excluded by review authors from meta-analysis - 'insufficient intervention' | | | | | | | | Global cognition | | | |
| Zhang, Ni & Liu | 2012 | | No | RCT | 75 older adults (age range 60-70 years; 39 dance group, 37 control group) | China | Square dancing | 48 weeks | 30-60 min/4-5x weekly | Global cognition | Meta-analysis | | High |
| ***Rodrigues-Krause et al., 2016*** | | | | | | | | | | | | | |
| Belardinelli et al | 2008 | Other review - Fong Yan et al., 2018 | | | | | | | | | | | |
| Cruz-Ferreira et al | 2015 | Other review - Rodrigues-Krause, Krause & Reischak-Oliveira, 2019 | | | | | | | | | | | |

| | | | | | | | | | | | | |
|---|---|---|---|---|---|---|---|---|---|---|---|---|
| Engels et al | 1998 | Excluded - dance performed with target HR (exercise, not performing arts intervention) | | | | | | | | | | |
| Hopkins et al | 1990 | Excluded - dance performed with target HR (exercise, not performing arts) | | | | | | | | | | |
| Kalsatou et al | 2014 | Other review - Fong Yan et al., 2018 | | | | | | | | | | |
| Kattenstroth et al | 2013 | Other review - Teixeira-Machado, Arida & de Jesus Mari, 2019 | | | | | | | | | | |
| Kim et al | 2011 | Excluded - dance performed with target HR (exercise, not performing arts) | | | | | | | | | | |
| ***Rodrigues-Krause, Krause, Reischak-Oliveira, 2019*** | | | | | | | | | | | | |
| Alpert et al | 2009 | No | Single group experimental study | 13 older women enrolled in a senior dance class (mean age = 68) | USA | Jazz | 15 weeks | Unspecified | Depression | No effect | No significant pre-post change in depression (Geriatric Depression Scale) | Very Low |
| | | | | | | | | | Balance | Positive | Significant pre-post improvement in balance in intervention vs control groups (Sensory Organization Test) | Very Low |
| | | | | | | | | | Cognition | No effect | No significant pre-post effect of dance on depression (MMSE) | Very Low |
| Belardinelli et al. | 2008 | Excluded - clinical population (heart failure) | | | | | | | | | | |
| Borges et al | 2012 | No | RCT | 75 sedentary elderly subjects (sex unspecified; 39 dance group (mean age = 68); 36 control (mean age = 67.2)) | Brazil | Ballroom | 8 months | 50 min/3x weekly | Functional autonomy | Positive | Significant increase in functional autonomy in dance group vs control (Latin American Group for Maturity protocol) | High |
| | | | | | | | | | Balance | Positive | Significant balance improvement in balance group vs control (postural sway; stabilometer) | High |
| Chen et al | 2013 | Included under 'epidemiologic' studies | | | | | | | | | | |
| Cruz-Ferreira et al | 2015 | No | RCT | 57 older women (32 dance group (mean age = 71.1); 25 | Portugal | Creative Dance | 24 weeks | 50 min/3x weekly | Physical Fitness | Positive | Significantly better fitness improvement in dance group vs control (Senior Fitness Test) | High |

| | | | | | | | | | | | | |
|---|---|---|---|---|---|---|---|---|---|---|---|---|
| | | | | control group (mean age = 72.8)) | | | | | Life Satisfaction | Positive | Significantly greater improvement in life satisfaction in dance vs control groups (Satisfaction with Life Scale) | High |
| Duncan et al | 2014 | Excluded - clinical population (Parkinson's) | | | | | | | | | | |
| Engels et al | 1998 | Excluded - investigation of aerobic dance performed with target HR of 50-70% maximum with participants instructed to monitor HR throughout (exercise, not performing arts, activity) | | | | | | | | | | |
| Eyigor et al. | 2009 | No | RCT | 37 female older adults (19 intervention (mean age 73.5); 18 control (mean age = 71.2)) | Turkey | Turkish folkloric dance | 8 weeks | 1 hour/3x weekly | Depression | No effect | No significant differences in depression in intervention group vs control (GDS) | High |
| | | Other review - Liu, Shen & Tsai, 2020 | | | | | | | Balance, Mobility, Endurance, General Health | | | |
| Federici et al | 2005 | No | RCT | 40 older adults (14 male/26 female; mean age = 63.1) | Italy | Caribbean dance | 3 months | 35-60 min/2x weekly (35 min 1st 2 weeks, then 60 min) | Balance | Positive | Significant improvements vs control on all balance tests (Tinetti test, Romberg test, improved Romberg test, Sit-up-and-go test) | High |
| | | | | | | | | | Smoking | No effect | (bespoke binary questionnaire) | Moderate |
| | | | | | | | | | Alcohol Consumption | No effect | (bespoke binary questionnaire) | Moderate |
| | | | | | | | | | Sexual activity | Positive | Significant improvement in sexual activity in experimental vs control groups (bespoke binary questionnaire) | Moderate |
| | | | | | | | | | Sleep quality | Positive | Significant improvement in sleep quality in experimental vs control group (bespoke binary questionnaire) | Moderate |
| Goertzen et al | 1984 | Excluded - descriptive population survey of physical activity participation | | | | | | | | | | |

| | | | | | | | | | | | | | |
|---|---|---|---|---|---|---|---|---|---|---|---|---|---|
| Granacher et al | 2012 | Yes | No | RCT | 28 older adults (17 female/11 male; 14 intervention (mean age = 71.6); 14 control (mean age = 68.9)) | Germany | Salsa | 8 weeks | 1 hour/2x weekly | Muscular Power (lower leg) | No effect | NO significant group*time effect (maximal countermovement jumps on force platform) | High |
| | | Other review - Liu, Shen & Tsai, 2020 | | | | | | | | Balance, Gait Speed | | | |
| Grant et al | 2002 | Excluded - descriptive analysis of HR & VO2 responses during aerobic dance (included in HR data) | | | | | | | | | | | |
| Hackney et al | 2009 | Excluded - clinical population (Parkinson's) | | | | | | | | | | | |
| Hackney et al | 2007 | Excluded - investigation of an intervention designed for clinical population (Parkinson's), but delivered jointly to a healthy/clinical cohort | | | | | | | | | | | |
| Hackney et al | 2013 | Excluded - clinical population (visual impairment) | | | | | | | | | | | |
| Hackney et al | 2009 | Excluded - clinical population (Parkinson's) | | | | | | | | | | | |
| Holmerova et al | 2010 | Excluded - clinical population (functionally impaired, non-independent) | | | | | | | | | | | |
| Hopkins et al | 1990 | Excluded - investigation of aerobic dance performed with target HR of 100-120 beats per minute with participants instructed to monitor HR throughout (exercise, not performing arts, activity) | | | | | | | | | | | |
| Hui et al | 2009 | | No | RCT | 97 older adults (52 intervention - 50 female/2 male, mean age = 68; 45 control - 44 female/1 male, mean age = 69.1) | Hong Kong | Low impact aerobic dance | 3 months | 50 min/2x weekly | Strength | Positive | Significant improvement vs control of dance intervention (sit and stand test) | Moderate |
| | | | | | | | | | | Balance | No effect | No significant effect of dance intervention vs control (4-test balance scale) | Moderate |
| | | | | | | | | | | Body composition | No effect | No significant effect of dance intervention vs control (BMI & waist to hip ratio) | Moderate |
| | | Other review - Liu, Shen & Tsai, 2020 | | | | | | | | Endurance, Mobility, Flexibility, Quality of Life | | | |
| Janyacharoen et al | 2013 | Other review - Liu, Shen & Tsai, 2020 | | | | | | | | | | | |
| Kalsatou et al | 2014 | Excluded - clinical population (Chronic Heart Failure) | | | | | | | | | | | |
| Kattenstroth et al | 2011 | | No | Cross-sectional | 49 healthy older adults (11 expert competitive dancers – mean age = 71.2, 5 | Germany | Ballroom | Mean 22.1 years competitive | Mean 4.6 hours weekly dance | Independent living | Positive | Significantly higher scores in dance vs control group (Everyday | Low |

| | | | | | | | | | | | | |
|---|---|---|---|---|---|---|---|---|---|---|---|---|
| | | | | female/11 male; 38 sedentary subjects – mean age = 71.7, 30 women/8 men) | | | dance experience | | | | Competence Questionnaire) | |
| | | | | | | | | | Cognitive performance | Positive | Significantly better performance in dance vs control (both groups)(Raven‘s Standard Progressive Matrices; nonverbal geriatric concentration test (AKT)) | Low |
| | | | | | | | | | Reaction time | Positive | Significantly faster reaction times in experimental vs control groups (multiple choice RT) | Low |
| | | | | | | | | | Balance | Positive | Significantly better performance in dance vs control group in Romberg test (eyes open only) and standing turn test (Romberg test; standing turn test)) | Low |
| | | | | | | | | | Mobility | Positive | Significantly better mobility performance in dance vs control groups (timed up and go) | Low |
| | | | | | | | | | Fine motor performance | Positive | Significantly more precise fine motor performance in dance vs control group (computer-based test battery) | Low |
| | | | | | | | | | Tactile performance | Positive | Significantly more sensitivity in dance vs control groups (von Frey filaments; 2-point discrimination) | Low |
| Kattenstroth et al | 2013 | Other review - Teixeira-Machado, Arida & de Jesus Mari, 2019 | | | | | | | | | | |
| Kattenstroth et al | 2010 | No | Cross-sectional | 62 healthy elderly adults (24 dance group - 19 female/5 | Germany | Unspecified ('amateur dancing') | Mean 16.5 years of | Mean 1.3 hours weekly dancing | Independent living | Positive | Significantly higher scores in dance vs control group | Low |

| | | | | | | |
|---|---|---|---|---|---|---|
| | male, mean age = 71.7; 38 control - 30 female/8 male, mean age = 71.7) | amateur dancing | | | (Everyday Competence Questionnaire) | |
| | | | Cognitive performance | Positive | Significantly better performance in dance vs control in general intelligence (RPSM) but not concentration (AKT)(Raven‘s Standard Progressive Matrices; nonverbal geriatric concentration test (AKT)) | Low |
| | | | Reaction time | Positive | Significantly faster reaction times in dance vs control group(multiple choice RT) | Low |
| | | | Balance | Positive | Significantly better performance in dance vs control group in standing turn test; no significant differences in Romberg test (Romberg test; standing turn test)) | Low |
| | | | Mobility | Positive | Significantly better mobility performance in dance vs control group (timed up and go) | Low |
| | | | Fine motor performance | Positive | Significantly more precise fine motor performance in dance vs control group (computer-based test battery) | Low |
| | | | Tactile performance | Positive | Significantly better 2-point discrimination in dance vs control group; no significant difference in von Frey touch threshold (von | Low |

| | | | | | | | | | | | | |
|---|---|---|---|---|---|---|---|---|---|---|---|---|
| | | | | | | | | | | | Frey filaments; 2-point discrimination) | |
| Kim et al | 2011 | Excluded - Latin dance performed with a target heart rate (exercise, not performing arts, intervention) | | | | | | | | | | |
| Kim et al | 2007 | Excluded - combined analysis of dance + nutrition education intervention | | | | | | | | | | |
| Kim et al | 2003 | Excluded - combined analysis of dance within context of 'health promotion program' including health education classes and counseling sessions | | | | | | | | | | |
| Krampe | 2013 | Excluded - clinical population (90% of cohort with at least 1 chronic condition) | | | | | | | | | | |
| Krampe et al | 2014 | Excluded - clinical population (chronic pain) | | | | | | | | | | |
| Krampe et al | 2010 | No | Single group experimental study | 11 elderly adults (7 women/4 men; age unspecified) | USA | Lebed Method | 6 weeks | 45 min/3x weekly | Balance | No effect | No significant pre-post effect (functional reach test) | Very Low |
| | | | | | | | | | Mobility | No effect | No significant pre-post effect (Timed Up & Go) | Very Low |
| Lesser et al | 2016 | No | RCT | 75 post-menopausal obese south east Asian women (no type 2 diabetes or CVD)(26 dance – mean age = 57.7; 23 standard exercise group – mean age = 56.4; 26 control – mean age =57.7) | Canada | Bhangra | 12 weeks | 60 min/3x weekly | Body composition | Positive | Significant reduction in abdominal adipose tissue (total & subcutaneous) in dance vs control group; no significant change in visceral adipose tissue or body fat/lean mass (Visceral adipose tissue (tomography scan); lean body mass/fat mass/% body fat (DEXA)) | High |
| | | | | | | | | | Glucose | No effect | No significant group*time interaction | High |
| | | | | | | | | | Insulin | No effect | No significant group*time interaction | High |
| | | | | | | | | | Aerobic fitness | Positive | Significant improvement in dance group vs control (VO2 peak) | High |
| Lim et al | 2015 | Excluded – Korean dance included as a small portion of a broader exercise program | | | | | | | | | | |
| Marin et al | 2009 | Excluded – dance included as part of broader fitness program (1 day/week dance; 2 days/week toning exercise) | | | | | | | | | | |
| Mavrovouniotis et al | 2008 | No | Non-randomized | 111 healthy older adults (mean age = | Greece | | 60 min | Once | Anxiety | Positive | (State-Trait Anxiety Inventory) | Low |

| | | | | | | | | | | | | |
|---|---|---|---|---|---|---|---|---|---|---|---|---|
| | | | experimental study | 69.8; 75 female/36 male; 76 dance group, 35 control) | | Greek traditional dance | | | Psychological wellbeing | Positive | (Subjective Exercise Experiences Scale (SEES)) | Low |
| | | | | | | | | | Psychological distress | Positive | (Subjective Exercise Experiences Scale (SEES)) | Low |
| | | | | | | | | | Fatigue | Positive | (Subjective Exercise Experiences Scale (SEES)) | Low |
| McKee et al | 2013 | Excluded - clinical population (Parkinson's) | | | | | | | | | | |
| McKinley et al | 2008 | No | RCT | 25 older adults (14 intervention - 11 female/3 male, mean age = 78.1; 11 control - 8 female/3 male, mean age = 74.6) | Canada | Argentine tango dance | 10 weeks | 2 hours/2x weekly | Balance confidence | Positive | Significant improvement in tango vs walk group (Activity specific Balance Confidence Scale (ABC)) | High |
| | | Other review - Liu, Shen & Tsai, 2020 | | | | | | | Gait, Mobility | | | |
| Murrock et al | 2014 | Excluded - clinical population (depression) | | | | | | | | | | |
| Park et al | 2015 | Excluded – Korean dance included as a small portion of a broader exercise program | | | | | | | | | | |
| Rios Romenets et al | 2015 | Excluded - clinical population (Parkinson's) | | | | | | | | | | |
| Shigematsu et al | 2002 | Excluded – intensity of dance-based exercise program set at individual's lactate threshold | | | | | | | | | | |
| Sofianidis et al | 2009 | Other review - Liu, Shen & Tsai, 2020 | | | | | | | | | | |
| Song et al | 2004 | Excluded - Korean dance intervention with a target heart rate (exercise, not performing arts) | | | | | | | | | | |
| Uusi-Rasi et al | 1999 | Excluded - combined analysis of Finnish gymnastics and folk dancing | | | | | | | | | | |
| Verghese | 2006 | No | Cross-sectional | 48 older adults (mean age = 80.4; 16 men/32 women; 24 social dancers/24 non-dancers) | USA | Various ('social' dancing) | Mean 36.5 years social dance experience | Unspecified | General mental status | No effect | No significant differences between dance & control groups (Blessed Information Memory Concentration test) | Low |
| | | | | | | | | | Memory (episodic) | No effect | No significant differences between dance & control groups (Free and Cued Selective Reminding Test) | Low |

| | | | | | | | | | | | | |
|---|---|---|---|---|---|---|---|---|---|---|---|---|
| | | | | | | | | | Executive function | No effect | No significant differences between dance & control groups (Wechsler Adult Intelligence Scale - revised (Digit Span, Digit Symbol, Block Design Tests); Verbal Fluency Test; Trail Making Test | Low |
| | | | | | | | | | Depression | No effect | No significant differences between dance & control groups (Geriatric Depression Scale) | Low |
| | | | | | | | | | Gait | Positive | Significantly greater gait speed, step/stride length & more favorable balance of % swing & stance phase in social dancers vs control (GaitRITE) | Low |
| | | | | | | | | | Balance | Positive | Significantly better performance in standing (physical performance battery, unipedal stance) but not dynamic (chair rise) balance vs control (Physical Performance Battery; unipedal stance; 5x sit to stand) | Low |
| | | | | | | | | | Strength | No effect | No significant differences between dance & control groups (grip strength) | Low |
| Wallmann et al | 2009 | No | Single group experimental study | 12 healthy older women (mean age = 68) | USA | Jazz | 15 weeks | 90 min/weekly | Balance | Positive | Significant pre-post improvement(Sensory Organization Test) | Very Low |
| White et al | 1984 | No | RCT | 96 older Caucasian women (age ~56; 36 walking group, 36 aerobic dancing, 24 control) | USA | Aerobic dance | 6 months | 2-5 dances, 2-4 days/week | Bone Composition | Positive | Significant pre-post increase in bone width & cross-sectional moment of inertia; no significant change in | Low |

| | | | | | | | | | | | | |
|---|---|---|---|---|---|---|---|---|---|---|---|---|
| | | | | | | | | | | | bone mineral content (bone mineral content, bone width, cross-sectional moment of inertia of radius) | |
| | | | | | | | | | Oestrogen | No effect | | Low |
| | | | | | | | | | Strength (arm) | Positive | Significant pre-post improvement vs control (isometric – elbow flexion) | Low |
| Wu et al | 2016 | | No | RCT | 32 sedentary females (mean age 59) | Taiwan | Low impact aerobic dance (no 'fitness parameters' (i.e. target HR, intensity)) | 16 weeks | 60 min/3x weekly | Body composition | Positive | Significant reduction in dance vs control group (% body fat – InBody 720) | High |
| | | | | | | | | | Cholesterol | Positive | Significant increase in HDL in dance vs control group (no change in LDL or total cholesterol) | High |
| | | | | | | | | | Triglycerides | Positive | Significant decrease in triglycerides vs control group | High |
| | | | | | | | | | Glucose | No effect | No significant difference between groups | High |
| | | | | | | | | | Bone density | No effect | No significant difference between groups (DEXA) | High |
| | | | | | | | | | Flexibility | Positive | Significant increases in ankle inversion, eversion and dorsiflexion and knee extension vs control (range of motion – knee, ankle) | High |
| | | | | | | | | | Strength | Positive | Significant increase vs control (peak knee extension torque) | High |
| | | Other review - Liu, Shen & Tsai, 2020 | | | | | | | Falls, Fear of Falling | | | |
| Wu et al | 2011 | | No | Cross-sectional | 38 sedentary and active healthy females (12 sedentary | Taiwan | Low impact aerobic dance | At least 3 years aerobic | At least 150 min/week | Body composition | No effect | No significant differences between dance & control | Low |

| | | | | | | | | | | | | |
|---|---|---|---|---|---|---|---|---|---|---|---|---|
| | | | | | group (<20 mins weekly PA) - mean age = 62; 26 exercise group - mean age = 59.1) | | | dance experience | | | | groups (% body fat - InBody 720) | |
| | | | | | | | | | | Flexibility | Positive | Significantly greater ankle ROM in dance vs control group, no significant differences in knee ROM (range of motion of knee & ankle joints - goniometer) | Low |
| | | | | | | | | | | Bone density | Positive | Significantly greater bone mineral density T-score in dance vs control group (GE bone ultrasonometer) | Low |
| | | | | | | | | | | Strength | Positive | Significantly greater knee extension torque in dance vs control group (knee extension torque) | Low |
| Young et al | 2007 | Excluded - dance a small part of a broader fitness program (foot stamping, progressively loaded squats & line dancing) | | | | | | | | | | | |
| Zhang et al | 2008 | | No | Cross-sectional | 404 older adults (mean age = 61.4; mixed sex - exact breakdown unspecified) | China | Various ('social' dancing) | Mean 6.1 years dancing | Mean 4.8 weekly dancing sessions | Gait speed | Mixed | Significantly faster gait speed vs control in males only (analysis adjusted for BMI, years of education, hypertension & diabetes) | Low |
| | | | | | | | | | | Flexibility | Mixed | Significantly increased flexibility in younger (50-59 year old) & male subjects vs. control only ( (analysis adjusted for BMI, years of education, hypertension & diabetes)(sit & reach test) | Low |
| | | | | | | | | | | Reaction time | Positive | Significantly improved reaction time vs control (analysis adjusted for BMI, years of education, | Low |

| | | | | | | | | | | | | |
|---|---|---|---|---|---|---|---|---|---|---|---|---|
| | | | | | | | | | | | hypertension & diabetes)(leg - Motor Choice Reaction Test) | |
| | | | | | | | | | Balance | Mixed | Significantly improved balance control in older cohort only (analysis adjusted for BMI, years of education, hypertension & diabetes)(postural sway) | Low |
| ***Teixeira-Machado, Arida & de Jesus Mari, 2019*** | | | | | | | | | | | | |
| Baniqued et al. | 2018 | No | RCT | 188 older adults (127 female/61 male; mean age = ~65; minimum Mini-Mental State Examination (MMSE) score = 27; 48 dance group, 42 walk group, 44 walk+ beta-alanine group, 54 stretch-strength-stability) | USA | 'Social' dance sequences (i.e. Contra & English country dancing) | 6 months | 3x/week | VO2 max | No effect | No significant pre-post difference in cardiorespiratory fitness in dance group; group*time interaction - walk & walk+ group generated significantly greater fitness gains | High |
| | | | | | | | | | Executive function | No effect | No significant effect of dance intervention on executive function; group*time interaction demonstrating significant advantages of walk, walk+ & SSS groups vs. dance in executive function improvement(Virginia Cognitive Aging Project fluid reasoning tasks + Switching Task & Spatial Working Memory Task (principal components analysis run to verify relations between tests)) | High |
| | | | | | | | | | Perceptual speed | No effect | No significant effects of any intervention on perceptual speed | High |

| | | | | | | | | | | | | | |
|---|---|---|---|---|---|---|---|---|---|---|---|---|---|
| | | | | | | | | | | | | (Virginia Cognitive Aging Project perceptual speed category) | |
| | | | | | | | | | | Episodic memory | No effect | No significant effects of any intervention on episodic memory(Virginia Cognitive Aging Project episodic memory category) | High |
| | | | | | | | | | | Vocabulary | No effect | No significant effects of any intervention on vocabulary (Virginia Cognitive Aging Project vocabulary category) | High |
| | | | | | | | | | | Fluid intelligence | No effect | No significant difference between groups (Virginia Cognitive Aging Project) | Low |
| | | | | | | | | | | Perceptual speed | No effect | No significant difference between groups (Virginia Cognitive Aging Project) | Low |
| Burzynska et al. | 2017 | | No | Cross-sectional | 40 female university students (mean age = 21.5; 20 dancers, 20 non-dancers) | USA | Unspecified | Mean 12 years dance practice | Mean 14 hours/week | Spatial/relational memory | No effect | No significant difference between groups (Virginia Cognitive Aging Project) | Low |
| | | | | | | | | | | Balance | Positive | significantly better balance in dancers vs non-dancers (eyes closed, 1-legged stance on foam block with dominant leg) | Low |
| | | | | | | | | | | Cognitive structure & function | Positive | Significantly different brain structure & function in dancers vs non-dancers (fMRI) | Low |
| Doi et al. | 2017 | Excluded - study of clinical population (mild cognitive impairment) | | | | | | | | | | | |

| | | | | | | | | | | | | |
|---|---|---|---|---|---|---|---|---|---|---|---|---|
| Ehlers et al. | 2017 | No | RCT | 247 older adults (169 female/78 male; mean age = ~65; minimum Mini-Mental State Examination (MMSE) score = 23; 69 dance group, 54 walk group, 54 walk+ beta-alanine group, 70 stretch-strength-stability) | USA | 'Social' dance sequences (i.e. Contra & English country dancing) | 6 months | 3x/week | Loneliness | Positive | Significant reduction in pre-post loneliness across all 4 groups(UCLA Loneliness Scale) | Moderate |
| | | | | | | | | | Social Support | Positive | Significant improvements in social support across all 4 groups (no inter-group differences)(Social Provisions Scale) | Moderate |
| | | | | | | | | | Stress | Positive | Significant pre-post reduction in stress across all 4 groups (no inter-group differences)(Perceived Stress Scale) | Moderate |
| Kattenstroth et al. | 2013 | No | RCT | 35 healthy elderly volunteers (25 dance group - mean age = 68.6, 17 female/8 male; 10 control - mean age = 72.3, 7 female/3 male) | Germany | Agilando (dance program developed for elderly people; no partner necessary) | 24 weeks | 1 hour/weekly | Lifestyle/quality of life | Positive | Significant pre-post improvement in dance group but not control (Everyday Competence Questionnaire; Fragebogen zur Lebenszufriedenheit (FLZ)) | Low |
| | | | | | | | | | Attention | Positive | Significant pre-post improvement in dance group but not control group (non-verbal geriatric concentration test (AKT); Frankfurt Attention Inventory (FAIR)) | Low |
| | | | | | | | | | Intelligence (nonverbal) | No effect | No pre-post effect of dance on nonverbal intelligence (Non-verbal learning test (NVLT)) | Low |
| | | | | | | | | | Fluid intelligence | No effect | No significant pre-post differences in either dance or control groups (Raven | Low |

| | | | | | | | | | | | | |
|---|---|---|---|---|---|---|---|---|---|---|---|---|
| | | | | | | | | | | | Standard Progressive Matrices (RSPM)) | |
| | | | | | | | | | Reaction time | Positive | Significant pre-post improvement in dance group but not control group | Low |
| | | | | | | | | | Balance | Positive | Significant pre-post improvement in dance group but not control group | Low |
| | | | | | | | | | Fine-motor performance | Positive | Significant pre-post improvement in dance group but not control group (MLS) | Low |
| | | | | | | | | | Tactile performance | Positive | Significant pre-post improvement in dance group but not control group (touch threshold (von Frey filaments), 2-point discrimination threshold, haptic object recognition) | Low |
| | | | | | | | | | Cardiorespiratory fitness | No effect | No significant pre-post differences in dance or control groups (VO2 peak) | Low |
| | | Other review (Meng et al., 2020) | | | | | | | Global cognition (RBANS) | | | |
| Muller et al. | 2017 | No | RCT | 52 healthy older adults (Baseline sex breakdown unspecified; mean age = ~68, 26 dance/26 sport; final analyses completed on only 22 subjects (10 female/12 male; 12 dance, 10 sport)) | Germany | Multiple (line, jazz, rock'n'roll, square dance) | 18 months | 90 min/twice weekly (1st 6 months); 90 min/weekly (last 12 months) | Cardiovascular fitness | No effect | No significant effect of group or time on physical fitness (Physical Working Capacity 130) | Moderate |
| | | | | | | | | | Memory | Positive | Significant effect of time across both groups; No significant group*time effect (Rey Auditory Verbal Learning Test) | Low |
| | | | | | | | | | Attention | Positive | Significant effect of time across both groups; No significant group*time interaction | Low |

| | | | | | | | | | | | | |
|---|---|---|---|---|---|---|---|---|---|---|---|---|
| | | | | | | | | | | | (Test of Attentional Performance (TAP)) | |
| | | | | | | | | | BDNF | Positive | Significant increase in BDNF vs control in dance group at 6-month timepoint (sustained to 18 month timepoint) | Moderate |
| | | | | | | | | | Cognitive structure | Positive | Significant increase in grey matter in dancers vs control | Moderate |
| Rehfeld et al. | 2018 | Excluded - same study as Muller et al., 2017 | | | | | | | | | | |
| Rehfeld et al. | 2017 | No | RCT | 52 healthy older adults (Baseline sex breakdown unspecified; mean age = ~68, 26 dance/26 sport; final analyses completed on only 22 subjects (10 female/12 male; 12 dance, 10 sport)) | Germany | Multiple (line, jazz, rock'n'roll, square dance) | 18 months | 90 min/twice weekly (1st 6 months); 90 min/weekly (last 12 months) | Balance | Positive | Significant group*time effect favoring the dance group (Sensory Organization Test - Balance Master System) | Moderate |
| | | | | | | | | | Hippocampal grey matter volume | Positive | Significant increases in hippocampal grey matter volume in right hippocampus in dance group only (MRI) | Moderate |
| ***Veronese et al., 2017*** | | | | | | | | | | | | |
| da Silva Borges | 2014 | No | RCT | 59 elderly healthy retirement home residents (sex unspecified; dance group - mean age = 68; control group - mean age = 67) | Brazil | Ballroom | 12 weeks | 50 min/3x weekly | Falls | Positive | Significant reduction Significant reduction in falls in dance vs control groups (interviews confirmed with medical records) | High |
| Merom et al. | 2016 | No | RCT | 530 older adults aged at or over 65 (279 intervention - 231 female/38 male; 251 control - 217 female/34 male) | Australia | Folk dance | 12 months | 1 hour/2x weekly | Falls | No effect | (diaries) | High |
| | | | | | | | | | Falls risk | No effect | (PPA) | High |
| Pichierri, Murer & de Bruin | 2012 | No | RCT | 31 retirement home residents (mixed sex - sex breakdown only available for analyzed | Switzerland | Video game dance | 12 weeks | 10-15 minutes/2x weekly (on top of weekly exercise/ | Fear of falling | No effect | No significant differences between dance & control groups (Falls efficacy scale international) | Moderate |

| | | | | | | | | | | | | |
|---|---|---|---|---|---|---|---|---|---|---|---|---|
| | | | | participants; mean age = 86.2) | | | | balance training program) | | | | |
| Ventura et al | 2016 | Excluded - clinical population (Parkinson's) | | | | | | | | | | |
| Volpe et al | 2013 | Excluded - clinical population (Parkinson's) | | | | | | | | | | |
| Wu et al. | 2016 | No | RCT | 32 healthy female older people (16 dance/16 control; mean age = 59) | Taiwan | Low impact aerobic dance (no 'fitness parameters' (i.e. target HR, intensity)) | 16 weeks | 60 min/3x weekly | Fear of falling | Positive | Significantly improved fear of falling in dance group vs control (Modified Falls Efficacy Scale) | High |
| | | | | | | | | | Falls | NO effect | No significant differences in falls in dance vs control group (self-report) | High |

*Table S8. Details of individual studies in included reviews of music participation.*

| Author | Year | In other review / exclusion? | Industry support? | Design | Subjects | Study country | Type of music participation | Length of intervention | Frequency/duration of each session | Outcome | Effect of music participation | Outcome details | GRADE Certainty of Evidence |
|---|---|---|---|---|---|---|---|---|---|---|---|---|---|
| ***Butzlaff, 2000*** | | | | | | | | | | | | | |
| College Board | 1988-1998 (not 1993) | | No | Cross-sectional | 5,732,282 high school seniors, mixed sex | USA | Instrumental or vocal music class | At least 1 course | Not specified | Reading ability (correlation) | Meta-analysis | | Moderate |
| Douglas & Willats | 1994 | | No | Cross-sectional | 78 4th graders (mean age 8), 40 female/38 male | UK | N/A (correlational analysis of rhythm, pitch, reading & spelling) | | | Reading ability (experimental) | Meta-analysis | | Low |
| Engdahl | 1994 | | No | Retrospective cohort study | 598 elementary school students (enrolled as 3rd graders), sex unspecified - mixed | USA | Instrumental music education (orchestra & band) | 3 years | At least 90 minutes, weekly (school weeks) | Reading ability (correlation) | Meta-analysis | | Low |
| Fetzer | 1994 | | No | Longitudinal cohort study | 30 children (unspecified age), sex unspecified | USA | Singing | 5-6 months | Unspecified | Reading ability (experimental) | Meta-analysis | | Low |
| Friedman | 1959 | | No | Longitudinal cohort study | 254 5th & 6th graders, sex unspecified - mixed | USA | Instrumental music education | 8 months | 7-8 hours, weekly | Reading ability (correlation) | Meta-analysis | | Low |
| Groff | 1963 | | No | Longitudinal cohort study | 460 6th graders (278 boys, 182 girls) | USA | Instrumental music education | Unclear | 30 min, 1-2x/week | Reading ability (correlation) | Meta-analysis | | Low |
| Kelly | 1981 | | No | RCT | 42 1st graders (mean age 6.6), sex unspecified - mixed | USA | Orff-Schulwerk music education | 6 months | 30 min, 3x/week | Reading ability (experimental) | Meta-analysis | | Moderate |
| Kvet | 1985 | | No | Retrospective cohort study | 175 6th graders, sex unspecified | USA | Instrumental music education | 8 months | 70 min, weekly | Reading ability (correlation) | Meta-analysis | | Low |
| Lamar | 1989 | | No | Cross-sectional | 140 students (70 1st graders; 70 4th graders), sex unspecified | USA | 'Music education' | Musical experience unclear | 30 min, weekly | Reading ability (correlation) | Meta-analysis | | Low |

| | | | | | | | | | | | | |
|---|---|---|---|---|---|---|---|---|---|---|---|---|
| McCarthy | 1992 | No | Unclear (correlational) | 957 college music majors, age & sex unavailable | USA | Orchestra | At least 2 years college music instruction | Unspecified | Reading ability (correlation) | Meta-analysis | | Low |
| Olanoff & Kirschener | 1969 | No | Unclear (experimental) | 46 students, age & sex unavailable | USA | Music training | Unclear | Unspecified | Reading ability (experimental) | Meta-analysis | | Low |
| Roberts | 1978 | No | Unclear (experimental) | 33 students, age & sex unavailable | USA | 'Note reading, keyboard instrument' | Unclear | Unspecified | Reading ability (experimental) | Meta-analysis | | Low |
| Roskam | 1979 | No | Non-randomized experimental cohort study | 24 learning disabled children (age range 6-9), sex unspecified | USA | Music therapy/educatio n | 3 months | 1 hour, 2x/week | Reading ability (experimental) | Meta-analysis | | Low |
| Weeden | 1971 | No | Non-randomized experimental cohort study | 47 1st & 2nd graders, sex unspecified | USA | Suzuki violin education | 4 months | 2x/week | Reading ability (correlation) | Meta-analysis | | Low |
| ***Clift et al, 2010*** *(**Tables 1 & 2 in review analyzed only; Tables 3 & 4 excluded (clinical populations)**)* | | | | | | | | | | | | |
| Beck et al | 2000 | | Single group experimental study (pre-post analysis) | 42 professional singers (mean age 46.4; mean singing experience 36.6 years; 23 female, 18 male) | USA | Singing | 1.5 (performance) – 2.5 (rehearsal) hours | Once (3 pre-post tests; 2 rehearsal, 1 performance) | Immunoglobulin A | Positive | Significant pre-post increase in IgA after both rehearsals and performance (enhanced immune activity) | Very Low |
| | | | | | | | | | Cortisol | Mixed | Significant decrease in cortisol after rehearsals; significant increase in cortisol after performances | Very Low |
| Clift et al | 2009 | Excluded - qualitative outcomes only | | | | | | | | | | |
| Clift et al | 2010 | Excluded - descriptive, uncontrolled survey-based cross sectional study | | | | | | | | | | |
| Clift et al | 2010 | Excluded - descriptive, uncontrolled survey-based cross sectional study | | | | | | | | | | |
| Cohen | 2009 | Other review - Daykin et al., 2018 | | | | | | | | | | |
| Cohen et al | 2007 | No | Nonrandomized experimental study | 128 English speaking healthy, ambulatory adults older than 64 (mean age = ~79; 60 control group; 68 intervention group), 78-80% female | USA | Singing | 2 periods of 30 weeks over 2 years | Once per week (duration unspecified) | Overall health | No effect | No significant group*time interaction (Self-report - bespoke instrument) | Very Low |
| | | | | | | | | | Health system utilization | Positive | Intervention significantly reduced number of doctor visits in 24 study months in | Very Low |

| | | | | | | | | | | | | |
|---|---|---|---|---|---|---|---|---|---|---|---|---|
| | | | | | | | | | | | intervention vs control group (group*time interaction)(Self-report - bespoke instrument) | |
| | | | | | | | | | Medication usage (over the counter + prescription) | Positive | Intervention significantly impacted reduced usage of over the counter and prescription medication in intervention vs control group (Self-report - bespoke instrument) | Very Low |
| | | | | | | | | | Falls incidence | No effect | No significant group*time effect (Self-report - bespoke instrument) | Very Low |
| | | | | | | | | | Morale | No effect | No significant group*time interaction (Philadelphia Geriatric Center Morale Scale) | Very Low |
| | | | | | | | | | Depression | No effect | NO significant group*time interaction (Geriatric Depression Scale - Short Form) | Very Low |
| | | | | | | | | | Loneliness | No effect | No significant group*time interaction (Loneliness Scale - III) | Very Low |
| | | | | | | | | | Social activity level | Positive | Significantly greater participation in weekly and yearly social activities over time (group*time interaction) in intervention vs control group (Self-report - bespoke instrument) | Very Low |
| Cohen et al | 2007 | Excluded - descriptive, uncontrolled survey-based cross sectional study | | | | | | | | | | |
| Cohen et al | 2006 | Other review - Daykin et al., 2018 | | | | | | | | | | |
| Hillman | 2002 | Excluded - descriptive, uncontrolled survey-based cross sectional study | | | | | | | | | | |
| Houston et al | 1998 | No | Non randomized experimental cohort study | 61 residents of retirement homes | UK | Singing | 4 weeks | 1 hour | Anxiety | Positive | Significantly decreased anxiety vs control in singing group (General | Low |

| | | | | | | | | | | | | |
|---|---|---|---|---|---|---|---|---|---|---|---|---|
| | | | | (mean age 83.7, 44 female, 17 male) | | | | | | | Health Questionnaire (GHQ) & Hospital Anxiety & Depression Scale (HADS)) | |
| | | | | | | | | | Somatic symptoms | No effect | No effect of singing intervention on somatic symptoms (General Health Questionnaire (GHQ)) | Low |
| | | | | | | | | | Depression | Mixed | Significantly decreased depression vs control (Hospital Anxiety and Depression Scale (HADS)); no change in severe depression (General Health Questionnaire (GHQ)) | Low |
| | | | | | | | | | Social functioning | No effect | No effect of singing on social functioning (General Health Questionnaire (GHQ)) | Low |
| Kreutz et al | 2004 | Other review - Fancourt, Ockelford & Belai, 2014 | | | | | | | | | | |
| Kuhn | 2002 | Other review - Fancourt, Ockelford & Belai, 2014 | | | | | | | | | | |
| Lally | 2009 | Excluded – qualitative outcomes only | | | | | | | | | | |
| Latimer | 2008 | Excluded - descriptive, uncontrolled survey-based cross sectional study | | | | | | | | | | |
| Louhivuori, Salminen & Lebaka | 2005 | Excluded - descriptive, uncontrolled survey-based cross sectional study focused on motivations for singing rather than health benefits | | | | | | | | | | |
| Sandgren | 2009 | No | Single group experimental study (pre-post analysis) | 212 amateur/advanced choir members (mean age ~53; 152 women, 60 men) | Finland | Singing | 1 rehearsal (duration unspecified) | Once | Positive affect | Positive | Significantly increased feelings of alertness, happiness & pride following choral rehearsal (bespoke tool completed by the authors) | Very Low |
| Schorr-Lesnick et al | 1985 | No | Cross-sectional | 113 professional musicians (31 string & percussion (mean age 43.5); 48 wind/brass (mean age 47.4); 34 vocalists (mean age | USA | Singing; Wind Instrumentalists | Mean years performing range 18.6-27.5 across groups | Unspecified | Lung function | No effect | No significant differences between percussion/strings, wind/brass or vocalists on any lung function measure (maximum voluntary ventilation; forced expiratory | Low |

| | | | | | | | | | | | | |
|---|---|---|---|---|---|---|---|---|---|---|---|---|
| | | | | 41.3)), 94 male/19 female | | | | | | | volume; forced vital capacity; mean forced expiratory flow during the middle half of forced vital capacity; ratio forced expiratory volume:forced vital capacity; peak expiratory pressure; peak inspiratory pressure) | |
| Steurer et al | 1998 | | No | Cross-sectional | 89 adults (62 professional singers – 30 women (age range 34 – 58), 32 men (age range 39 – 61); 27 noise exposed workers (11 female, 16 male; all under 30 years old without signs of hearing impairment) | Austria | Singing | Mean 24 (male) – 27 (female) years performing experience | Mean weekly singing of 15 (male) – 29 (female) hours | Hearing threshold | Negative | Significantly impaired hearing thresholds in professional singers vs control | Very Low |
| Unwin, Kenny & Davis | 2002 | | No | RCT | 107 adult volunteers with varied singing experience (age range 18 – 73; 23 male, 84 female) | Australia | Singing | 30 minutes | Once | Mood | No effect | No significant effect of singing on mood vs active control (music listening)(POMS) | Moderate |
| Valentine & Evans | 2001 | Other review - Daykin et al., 2018 | | | | | | | | | | | |
| Wise, Hartmann & Fisher | 1992 | | No | Cross-sectional | 98 retirees (49 singing in a retirement village community chorus; 49 not in choir; mean age 64.6), sex unspecified - mixed | USA | Singing | Unspecified (chorus members generally with robust musical histories) | Unspecified | Life satisfaction | No effect | No significant differences between choral & comparison groups (Life Satisfaction Index A) | Very Low |
| | | | | | | | | | | Self-actualization | Unclear | Significantly lower self-actualization in choral vs comparison group, however this is hypothesized to be a surrogate for enhanced social quality of life (healthy dependency) (Personal Orientation Inventory) | Very Low |

| | | | | | | | | | | | | |
|---|---|---|---|---|---|---|---|---|---|---|---|---|
| | | | | | | | | | Social isolation (alienation) | No effect | No significant differences between choral & comparison groups (Middleton's Alienation Scale) | Very Low |
| ***Coffey, Mogilever & Zatorre, 2017*** | | | | | | | | | | | | |
| Baskent & Etienne | 2016 | No | Cross-sectional | 38 adults (mean age 22.2; 24 females, 14 males; 20 musicians, 18 nonmusicians) | Netherlands | Music training | At least 10 years of musical training, beginning before age 7 | Unspecified | Speech in speech | Positive | Significantly improved speech in speech perception in musicians vs nonmusicians (protocol as per Versfeld et al 2000) | Low |
| Bidelman & Weiss | 2014 | No | Cross-sectional | 24 adults (mean age 24.3; 12 musicians, 12 nonmusicians), 16 female/8 male | Canada | Instrumental | Average 13.6 musical training | Unspecified | Auditory processing | Positive | Significantly faster classification of speech and enhanced nervous system responsiveness to speech in musicians vs nonmusicians | Low |
| Boebinger et al. | 2015 | No | Cross-sectional | 50 adults (mean age 27.2; 25 musicians, 25 nonmusicians), sex unspecified | UK | Instrumental | Average 22.7 years of musical training | Practice at least 3x/weekly over the 3 years before testing | Speech in noise (Masked speech perception) | No effect | No significant differences between musicians and nonmusicians (Bamford-Kowal-Bench lists) | Very low |
| | | | | | | | | | Duration discrimination | No effect | No significant differences between musician and nonmusician groups (duration discrimination thresholds test - Grassi & Soranzo adaptive staircase procedure) | Low |
| | | | | | | | | | Pitch discrimination | Positive | Significantly better pitch discrimination performance in musicians vs nonmusicians (frequency discrimination thresholds test - Grassi & Soranzo adaptive staircase procedure) | Low |

| | | | | | | | | | | | | |
|---|---|---|---|---|---|---|---|---|---|---|---|---|
| | | | | | | | | | IQ (nonverbal) | No effect | No significant differences between musicians & nonmusicians (Matrix Reasoning subtest of Wechsler Abbreviated Scale of Intelligence (WASI)) | Low |
| | | | | | | | | | Selective attention | No effect | No significant differences between musicians & nonmusicians (Stroop task) | Low |
| | | | | | | | | | Mental flexibility (Motor speed) | No effect | No significant differences between musicians & nonmusicians (Trail making test) | Low |
| | | | | | | | | | Mental flexibility (Fluid intelligence) | No effect | No significant differences between musicians & nonmusicians (Trail making test) | Low |
| | | | | | | | | | Mental flexibility (Task switching) | No effect | No significant differences between musicians & nonmusicians (Trail making test) | Low |
| | | Other review - Talamini et al., 2017 | | | | | | | Short-term memory | | | |
| | | Other review - Talamini et al., 2017 | | | | | | | Working memory | | | |
| Clayton et al | 2016 | No | Cross-sectional | 34 adults (mean age 23.5; 17 musicians, 17 nonmusicians), sex unspecified | USA | Instrumental | Average 14.4 years of music training | Unclear | IQ (nonverbal) | No effect | No significant differences between musicians & nonmusicians (Wechsler Abbreviated Scale of Intelligence - matrix reasoning) | Low |

| | | | | | | | | | | | | |
|---|---|---|---|---|---|---|---|---|---|---|---|---|
| | | | | | | | | | Executive function (Inhibition control) | No effect | No significant differences between musicians & nonmusicians (color-word interference sub-test of Delis-Kaplan Executive Function System (DKEFS)) | Low |
| | | | | | | | | | Executive function (Cognitive flexibility/goal directed behavior) | No effect | No significant differences between musicians & nonmusicians (design fluency sub-test of the DKEFS) | Low |
| | | | | | | | | | Speech in speech | Positive | Significantly better speech in speech perception in musicians vs nonmusicians(as per Swaminathan et al. 2015) | Low |
| | | | | | | | | | Selective attention (Multiple object tracking) | No effect | No significant differences between musicians & nonmusicians (as per Pylyshyn & Storm, 1988; Drew & Vogel , 2008) | Low |
| | | Other review - Talamini et al., 2017 | | | | | | | Working memory | | | |
| Coffey et al | 2016 | Excluded - no analysis of the impact of musical training/participation on outcomes | | | | | | | | | | |
| Coffey et al | 2016 | No | Cross-sectional | 20 adults (mean age 25.7), 12 female/8 male | Canada | Unspecified | N/A (correlational analysis with musical experience as variable) | N/A | Pitch discrimination | Positive | Positive correlation between hours of musical training and enhanced fine pitch discrimination ability | Very Low |
| | | | | | | | | | Auditory processing | Positive | Positive correlation between hours of musical training and enhanced P2 amplitude | Very Low |
| Du & Zatorre | 2016 | No | Cross-sectional | 30 healthy adults (mean age ~21), sex unspecified | USA | Unspecified (conference abstract) | Unspecified | Unspecified | Speech in noise | Positive | Significantly better speech in noise perception in musicians vs nonmusicians | Low |

| | | | | | | | | | | | | |
|---|---|---|---|---|---|---|---|---|---|---|---|---|
| Fuller et al. | 2014 | No | Cross-sectional | 50 adults (mean age 22.7); 25 musicians, 25 nonmusicians; 14 male/36 female | Netherlands | Instrumental | Average 14.6 years musical training | Unspecified ('regular musical training within the last 3 years') | Speech in noise | Positive | Significantly better speech in noise processing in musicians vs nonmusicians | Low |
| | | | | | | | | | Vocal emotion identification | Positive | Significantly better vocal emotion identification in musicians vs nonmusicians | Low |
| | | | | | | | | | Melodic contour identification | Positive | Significantly better melodic contour identification in musicians vs nonmusicians | Low |
| Lee et al. | 2009 | No | Cross-sectional | 26 adults (10 musicians (mean age 25.8), 11 nonmusicians (mean age 23.5), 5 amateur musicians (mean age 23.2)), 16 female/10 male | USA | Instrumental/ vocal | At least 10 years musical experience beginning before age 7 | Unspecified | Auditory processing | Positive | Significantly more precise neural phase locking to the temporal periodicity of the amplitude modulated envelope (underlies perception of harmony) in musicians vs nonmusicians; significantly better perception of upper harmonic voice (behavioral adaptation) in musicians vs nonmusicians; neural enhancements strongly correlated with years of musical training | Low |
| Musacchia et al. | 2007 | No | Cross-sectional | 29 adults (mean age 25.6; 14 female, 15 male); 16 musicians, 13 controls | USA | Instrumental | At least 10 years musical experience, begun before age 10 | At least 4 hours/week | Auditory processing | Positive | Significantly larger brainstem responses to both speech and music stimuli in auditory and audiovisual conditions in musicians vs nonmusicians; enhanced phase-locking to stimulus periodicity (underlies perception of pitch) in musicians vs nonmusicians | Low |

| | | | | | | | | | | | |
|---|---|---|---|---|---|---|---|---|---|---|---|
| | | | | | | | | | | (correlated with increasing number of musical practice hours) | |
| Musacchia, Strait & Kraus | 2008 | No | Cross-sectional | 26 adults (mean age 25.6; 14 female); 14 musicians, 12 nonmusicians | USA | Instrumental | 10 or more years musical experience, started at or before age 5 | >4 hours weekly practice | Auditory processing (neural timing) | Positive | Significantly faster cortical and subcortical responses to auditory stimuli in musicians vs nonmusicians | Low |
| Oxenham et al. | 2003 | No | Cross-sectional | 24 adults (mean age 24.7); 12 musicians, 12 nonmusicians; sex unspecified | USA | Instrumental | Unspecified | Unspecified | Sound in noise | Positive | Significantly lower threshold values for detecting sound masked in noise in musicians vs nonmusicians | Low |
| | | | | | | | | | Speech in noise (Frequency selectivity) | No effect | No significant differences in frequency selectivity in musicians vs nonmusicians (notched-noise method) | Low |
| Parbery-Clark et al. | 2009 | No | Cross-sectional | 31 right handed English speaking adults (mean age 23; 16 musicians, 15 nonmusicians), 19 female/12 male | USA | Piano & violin | Average 16 years music training | Practice at least 3x/weekly over the 3 years before testing | Speech in Noise | Positive | Significantly better performance in musicians vs non-musicians (Quick Speech in Noise (QuickSIN) & Hearing in Noise Test (HINT)) | Low |
| | | | | | | | | | Working memory | Positive | Significantly better performance in musicians vs non-musicians(Woodcock-Johnson, numbers reversed & auditory working memory tests) | Low |
| | | | | | | | | | Pitch discrimination | Positive | Significantly better performance in musicians vs non-musicians (adaptive staircase method as per Levitt 1971) | Low |
| Parbery-Clark et al. | 2011 | No | Cross-sectional | 37 adults (mean age 50; 18 musicians, 19 nonmusicians); age range 45-65, sex unspecified | USA | Piano & violin | Average 50 years music training | Practice at least 3x/weekly over the 3 | Speech in noise | Positive | Significantly better speech in noise perception in musicians vs nonmusicians (Hearing in Noise Test | Low |

| | | | | | | | | | | | | |
|---|---|---|---|---|---|---|---|---|---|---|---|---|
| | | | | | | | | years before testing | | | (HINT) + Quick Speech in noise test (QuickSIN) + Words in Noise Test (WIN)) | |
| | | | | | | | | | Auditory processing (temporal acuity) | Positive | Significantly better auditory temporal acuity in musicians vs nonmusicians (Backward masking subtest from IHR Multi-center Battery for Auditory Processing) | Low |
| | | Other review - Talmini et al., 2017 | | | | | | | Working memory | | | |
| Parbery-Clark et al. | 2011 | No | Cross-sectional | 31 young adults (mean age 22.4; 16 musicians, 15 nonmusicians), 22 female/9 male | USA | Instrumental | Average 16.4 years musical participation, begun at or before age 7 | Undefined | Speech in noise | Positive | Significantly better speech in noise perception in musicians vs nonmusicians (Hearing in Noise Test (HINT) | Low |
| | | | | | | | | | IQ (nonverbal) | No effect | No significant differences between musicians and nonmusicians (Test of Nonverbal Intelligence (TONI)) | Low |
| Parbery-Clark et al. | 2012 | No | Cross-sectional | 48 middle-aged adults (mean age 56;23 musicians, 25 non-musicians), sex breakdown unspecified - mixed | USA | Instrumental (predominantly piano & violin) | Average 49 years musical experience | Practice at least 3x/weekly | Speech in noise | Positive | Significantly improved speech in noise perception in musicians vs nonmusicians (HINT; Speech subscale of Speech, Spatial and Qualities Questionnaire (self-report); more robust neural encoding of speech in both quiet and noise (neurophysiology) | Low |
| | | | | | | | | | IQ (nonverbal) | No effect | No significant differences between musicians and nonmusicians (Abbreviated Wechsler's Adult Scale | Low |

| | | | | | | | | | | | | |
|---|---|---|---|---|---|---|---|---|---|---|---|---|
| | | | | | | | | | | | of Intelligence matrix reasoning subtest) | |
| Parbery-Clark et al. | 2012 | No | Cross-sectional | 87 adults (50 younger - mean age 23; 37 older - mean age 56); 46 musicians, 41 nonmusicians; 62 female/25 male | USA | Unspecified | Consistent musical training started before age 9 | Engaged in musical activities a minimum of 3 times weekly | Auditory processing | Positive | Musical experience significantly counteracts age-related delays in subcortical response timing to formant transition | Low |
| Parbery-Clark, Skoe & Kraus | 2009 | No | Cross-sectional | 31 adults (mean age 23; 10 females); 16 musicians, 15 nonmusicians | USA | Instrumental | Practiced consistently for at least 10 years prior to study enrollment | Unspecified | Speech in noise | Positive | Significantly improved speech in noise perception in musicians vs nonmusicians (HINT; more robust subcortical representation of the acoustic stimulus in the presence of noise (neurophysiology)) | Low |
| Ruggles et al. | 2014 | No | Cross-sectional | 33 university students (mean age 21.2; 18 female, 15 male); 16 participants with at least 10 years formal music training | USA | Instrumental | At least 10 years of musical training, beginning before age 10 | Currently playing at least 5 hours/week | IQ | Positive | Significantly higher IQ (Wechsler Abbreviated Scale of Intelligence, 2nd edition) in musicians vs nonmusicians | Low |
| | | | | | | | | | Speech in noise (Voiced speech discrimination) | No effect | No significant differences between musicians vs non-musicians in voiced speech discrimination (as per Helfer & Freyman) | Low |
| | | | | | | | | | Speech in noise (Whispered speech discrimination) | No effect | No significant differences between musicians vs non-musicians in whispered speech discrimination (as per Helfer & Freyman) | Low |

| | | | | | | | | | | | | |
|---|---|---|---|---|---|---|---|---|---|---|---|---|
| Ruggles et al. | 2014 | No | Cross-sectional | 26 university students (unspecified age/sex); 14 participants with at least 10 years formal music training | USA | Instrumental | At least 10 years of musical training, beginning before age 10 | Currently playing at least 5 hours/week | Pitch discrimination | Positive | Significantly better pitch discrimination in musicians vs nonmusicians (unclear if this is a validated assessment) | Low |
| | | | | | | | | | Speech in Noise | No effect | No significant differences between musicians and non-musicians on clinical speech in noise assessments (QuickSIN; Adaptive HINT) | Low |
| Slater & Kraus | 2016 | No | Cross-sectional | 38 adult males (mean age 23.3; 21 vocalists, 17 nonmusicians) | USA | Vocal | Average 14.7 years musical experience | Undefined | Speech in noise | No effect | No significant differences between vocalists and nonmusicians in speech in noise perception (QuickSIN) | Low |
| | | | | | | | | | Speech in noise | Positive | Significantly better speech in noise perception in percussionists vs nonmusicians (QuickSIN) | Low |
| | | | | | | | | | Words in noise | No effect | No significant differences between musicians and nonmusicians in words in noise perception (Words in noise test as per Wilson et al. 2007) | Low |
| | | | | | | | | | IQ (nonverbal) | No effect | No significant differences between musicians and nonmusicians in nonverbal IQ (Test of Nonverbal Intelligence (TONI)) | Low |
| Slater et al. | 2015 | No | RCT | 38 elementary school children from disadvantaged backgrounds (mean age 8.2; 17 males, | USA | General music education + (after initial competency demonstrated in | 1 year | 1 hour, twice weekly | Speech in Noise | Positive | Significantly improved speech in noise perception in music group vs control group (Hearing in Noise Test); | Low |

| | | | | | | | | | | | | |
|---|---|---|---|---|---|---|---|---|---|---|---|---|
| | | | | 21 females; 19 music training group; 19 control) | | general classes (up to 1 year)) instrumental music lessons | | | | | **significant group*time main effect, but main significant difference at 2-year timepoint (music group with 2 years training; control group with 1 year music training) | |
| Strait & Kraus | 2011 | No | Cross-sectional | 23 adults (age range 18-35), sex unspecified | USA | Instrumental | Average 16.5 years musical experience | Engaged in musical activities a minimum of 3 times weekly | Auditory processing | Positive | Significantly greater impact of selective auditory attention on prefrontal evoked activity | Low |
| Strait et al. | 2010 | No | Cross-sectional | 33 adults (age range 18-40; 18 musicians, 15 nonmusicians), sex unspecified | USA | Unspecified | At least 10 years musical experience beginning before age 9 | At least 3 hours/week | Auditory working memory | No effect | No significant differences between musicians and nonmusicians (reverse digit task) | Low |
| | | | | | | | | | Auditory processing (attention) | Positive | Significantly better in musicians vs nonmusicians ('similar to the' Test of Attentional Performance) | Low |
| | | | | | | | | | Visual attention | No effect | No significant differences between musicians and nonmusicians ('similar to the' Test of Attentional Performance) | Low |
| | | | | | | | | | Frequency discrimination | Positive | Significantly better in musicians vs nonmusicians (3 down, 1 up procedure as per Amitay et al. 2006) | Low |
| | | | | | | | | | Speech in noise (backward masking) | Positive | Significantly better in musicians vs nonmusicians (3 down, 1 up procedure as per Amitay et al. 2006) | Low |

| | | | | | | | | | | | | |
|---|---|---|---|---|---|---|---|---|---|---|---|---|
| | | | | | | | | | Speech in noise (forward masking) | No effect | No significant differences between musicians and nonmusicians (3 down, 1 up procedure as per Amitay et al. 2006) | Low |
| | | | | | | | | | IQ (nonverbal) | No effect | No significant differences between musicians and nonmusicians (Wechsler Abbreviated Scale of Intelligence matrix reasoning subtest (WASI)) | Low |
| Swaminathan et al. | 2015 | No | Cross-sectional | 24 adults (mean age 21.7 years - 12 musicians, 12 nonmusicians), sex unspecified | USA | Instrumental | At least 10 years music training | Undefined - 'most practiced at least 5 hours per week' | Speech in noise | Positive | Significantly better speech in noise perception in musicians vs. nonmusicians when masking speech panned 15-degress to the right and left; no significant improvement in speech in noise perception when target and masking speech are co-located (procedure as per Kidd et al. 2010) | Very low |
| Tierney, Krizman & Kraus | 2015 | No | Nonrandomized longitudinal cohort study | 68 high schoolers (mean age 14.7), sex unspecified | USA | Instrumental | 3 years | 2.33-3 hours/week | Language skills (Phonological awareness) | Positive | Significantly greater gains in phonological awareness in music vs control group (Comprehensive Test of Phonological Processing) | Very Low |
| | | | | | | | | | Language skills (Phonological memory) | No effect | No significant differences in music vs control group in changes in phonological memory (Comprehensive Test of Phonological Processing) | Very Low |

| | | | | | | | | | | | | |
|---|---|---|---|---|---|---|---|---|---|---|---|---|
| | | | | | | | | | Language skills (Rapid naming) | No effect | No significant differences in music vs control group in changes in rapid naming (Comprehensive Test of Phonological Processing) | Very Low |
| | | | | | | | | | Auditory processing (Subcortical response consistency) | Positive | Significantly delayed waning of subcortical response consistency (characteristically observed between adolescence and young adulthood) in music vs control group | Very Low |
| | | | | | | | | | Auditory processing (cortical onset response) | Positive | Significantly greater increase in cortical onset response (correlate of auditory maturation) in music vs control group | Very Low |
| Varnet et al. | 2015 | No | Cross-sectional | 38 adults (mean age 22.9); 19 musicians, 19 nonmusicians; 12 male/26 female | France | Instrumental | 7-22 years musical practice | Unspecified | Speech in noise | Positive | Significantly lower speech in noise threshold in musicians vs nonmusicians (Levitt 1971 3-down, 1-up staircase procedure) | Low |
| | | | | | | | | | Auditory processing | Positive | Significantly more consistent response to auditory stimuli and greater speed of auditory learning in musicians vs nonmusicians (Auditory Classification Image approach) | Low |
| Zendel & Alain | 2012 | No | Cross-sectional | 163 adults (age range 18-91); 74 musicians, 89 nonmusicians; 86 female, 77 male | Canada | Unspecified | At least 6 years formal music lessons | Unspecified | Speech in noise | Positive | Significant association between musical training hours and superior speech in noise perception (QuickSIN) | Low |

| | | | | | | | | | | | | | |
|---|---|---|---|---|---|---|---|---|---|---|---|---|---|
| | | | | | | | | | | Auditory processing (threshold) | NO effect | NO significant effect of music training on auditory threshold, only age significant (minimum detectable amplitude at various frequencies) | Low |
| | | | | | | | | | | Auditory processing (temporal acuity (gap detection)) | Positive | Significantly less decline in auditory temporal acuity in musicians vs nonmusicians (minimum detectable gap between sounds - 3 down, 1 up as per Levitt 1971) | Low |
| | | | | | | | | | | Pitch discrimination (Mistuned harmonic detection) | Positive | Significantly better, across all ages, in musicians vs nonmusicians (2AFC procedure) | Low |
| Zendel & Alain | 2009 | | No | Cross-sectional | 28 adults (mean age 30.6; 14 musicians, 14 nonmusicians), 15 female/13 male | USA | Unspecified | Unspecified | Unspecified | Pitch discrimination (Two-tone detection) | Positive | Significantly more sensitive detection of two mis-matched tones (harmonically) in musicians vs nonmusicians | Low |
| Zendel et al. | 2015 | | No | Cross-sectional | 26 adults (age range 18-35; 13 musicians, 13 nonmusicians), 14 female/12 male | Canada | Instrumental & vocal | Average 15.5 years musical experience | Average 23.4 hours weekly music practice | Words in noise | Positive | Significantly better words in noise perception in musicians vs nonmusicians in most difficult condition (original test procedure) | Low |
| ***Daykin et al, 2013*** | | | | | | | | | | | | | |
| Anderson & Overy | 2010 | Excluded - Descriptive outcomes only, no statistical analyses of time or intergroup differences | | | | | | | | | | | |
| Baker & Homan | 2007 | Excluded - descriptive outcomes only, no quantitative data or statistical analyses | | | | | | | | | | | |
| Bittman, Dickson & Coddington | 2009 | | Yes (Yamaha & Remo) | RCT (crossover design) | 52 'at risk' youth aged 14-18 (30 females; 22 males) | USA | Percussion & keyboard education | 6 weeks | 1 hour, weekly | Social functioning | No effect | No significant benefit of music intervention vs control on social functioning (Child and Adolescent Functional | Moderate |

| | | | | | | | | | | | | |
|---|---|---|---|---|---|---|---|---|---|---|---|---|
| | | | | | | | | | | | Assessment Scale (CAFAS)) | |
| | | | | | | | | | Psychosocial function | No effect | No significant benefit of music intervention vs control on psychosocial functioning (Adolescent Psychopathology Scale (APS)) | Moderate |
| | | | | | | | | | Anger | No effect | No significant benefit of music intervention vs control on anger (Adolescent Anger Rating Scale (AARS)) | Moderate |
| | | | | | | | | | Depression | Positive | Significant improvement vs. control in depression (Reynolds Adolescent Depression Scale, 2nd edition (RADS 2)) | Moderate |
| de Carlo & Hockman | 2003 | Excluded - analysis of rap music for therapeutic reasons, but no active music participation | | | | | | | | | | |
| de Roeper & Savelsberg | 2009 | Excluded - qualitative outcomes only, no attempt to measure effects | | | | | | | | | | |
| Gann | 2010 | Excluded - unpublished doctoral dissertation, no access available | | | | | | | | | | |
| Kennedy | 1998 | No | RCT | 45 juvenile delinquents or 'at risk' youth aged 7-19 with little/no guitar experience, sex unspecified (n=9 across 5 groups; all groups received 30 mins weekly guitar lessons + 30 min performance training, 15/15 min performance/cognitive training, 30 min cognitive training, 30 min vicarious | USA | Guitar instruction + performance training | 3 months | 1 hour, weekly (30 mins guitar instruction + 30 mins performance training) | Self-esteem | Positive | Significant improvement in self esteem (Rosenberg Self Esteem measure) in guitar instruction + performance (with or without cognitive training) groups vs. other groups with no performance training | High |

| | | | | | | | | | | | | | |
|---|---|---|---|---|---|---|---|---|---|---|---|---|---|
| | | | | | experience no training) | | | | | | | | |
| Lashua | 2005 | Excluded - analysis of 'remixing' intervention for at-risk youth - participants remixed songs using computer software, not ‘active performing arts’ participation | | | | | | | | | | | |
| Lotter | 2003 | Excluded - qualitative outcomes only | | | | | | | | | | | |
| Tyson | 2002 | Excluded - focus on listening and analysis to rap music | | | | | | | | | | | |
| Woodward, Sloth-Nielson & Mathiti | 2008 | Excluded - mostly qualitative outcomes, no analysis or contextualization of the one quantitative outcome (recidivism rate) | | | | | | | | | | | |
| ***Daykin et al., 2018*** *(**Supplementary Table 1. Quantitative Outcomes – included only**)* | | | | | | | | | | | | | |
| Anderson & Overy | 2010 | Excluded - Descriptive outcomes only, no statistical analyses of time or intergroup differences | | | | | | | | | | | |
| Baldari et al. | 2010 | Excluded - music listening during exercise | | | | | | | | | | | |
| Berkiroglu et al. | 2013 | Excluded - clinical population (hypertension) | | | | | | | | | | | |
| Bensimon et al. | 2015 | Excluded - music listening | | | | | | | | | | | |
| Bensimon & Gilboa | 2010 | Excluded - study of 'Musical Presentation' (a therapeutic activity in which participants in a group present themselves using music of their choice) | | | | | | | | | | | |
| Boothby & Robbins | 2011 | Excluded - music listening | | | | | | | | | | | |
| Burns et al. | 2002 | Excluded - music listening | | | | | | | | | | | |
| Campion & Levita | 2014 | Excluded - music listening | | | | | | | | | | | |
| Carissoli et al | 2015 | Excluded - music listening | | | | | | | | | | | |
| Chan et al. | 2010 | Excluded - music listening | | | | | | | | | | | |
| Chan et al. | 2012 | Excluded - music listening | | | | | | | | | | | |
| Chang et al. | 2015 | Excluded - music listening | | | | | | | | | | | |
| Chang et al. | 2008 | Excluded - music listening | | | | | | | | | | | |
| Chen et al. | 2015 | Excluded - music listening | | | | | | | | | | | |
| Cohen et al. | 2006 | | No | Nonrandomized experimental study | 166 English speaking healthy, ambulatory adults older than 64 (mean age = ~79; 76 control group; 90 intervention group), sex unspecified | USA | Singing | 30 weeks | Once per week (duration unspecified) | Overall health | Positive | Significantly improved overall health in intervention vs control group (*statistical comparison only of post-intervention data, no accounting for baseline levels/intergroup analysis of change)(Self-report - bespoke instrument) | Very Low |

| | | | | |
|---|---|---|---|---|
| | Health system utilization | Positive | Significantly reduced number of doctors visits in last 12 months in intervention vs control group (*statistical comparison only of post-intervention data, no accounting for baseline levels/intergroup analysis of change)(Self-report - bespoke instrument) | Very Low |
| | Medication usage (over the counter) | Positive | Significantly reduced usage of over the counter medication in intervention vs control group (*statistical comparison only of post-intervention data, no accounting for baseline levels/intergroup analysis of change)(Self-report - bespoke instrument) | Very Low |
| | Medication usage (prescription) | No effect | No significant difference in prescription medication usage in intervention vs control group (*statistical comparison only of post-intervention data, no accounting for baseline levels/intergroup analysis of change)(Self-report - bespoke instrument) | Very Low |
| | Falls incidence | Positive | Significantly reduced number of falls in intervention vs control group (*statistical comparison only of | Very Low |

| | | | |
|---|---|---|---|
| | | post-intervention data, no accounting for baseline levels/intergroup analysis of change)(Self-report - bespoke instrument) | |
| Other health problems | Positive | Significantly reduced reporting of other health problems in intervention vs control group (*statistical comparison only of post-intervention data, no accounting for baseline levels/intergroup analysis of change)(Self-report - bespoke instrument) | Very Low |
| Morale | Positive | Significantly improved morale in intervention vs control group (*statistical comparison only of post-intervention data, no accounting for baseline levels/intergroup analysis of change)(Self-report - bespoke instrument) | Very Low |
| Depression | No effect | No effect of intervention on depression vs control (statistical analysis of change in depression)(Geriatric Depression Scale - Short Form) | Low |
| Loneliness | Positive | Significantly reduced loneliness in intervention vs control group (appropriate statistical analysis | Low |

| | | | | | | | | | | | | |
|---|---|---|---|---|---|---|---|---|---|---|---|---|
| | | | | | | | | | | | accounting for baseline levels)(Loneliness Scale - III) | |
| | | | | | | | | | Social activity level | Positive | Significantly higher numbers of weekly social activities in intervention vs control group (*statistical comparison only of post-intervention data, no accounting for baseline levels/intergroup analysis of change)(Self-report - bespoke instrument) | Very Low |
| Cohen | 2009 | No | Nonrandomized experimental study | 20 male inmates (mean age 35.5; 10 singers, 10 control) | USA | Singing | 9 weeks (data analyzed only pre-post the final performance) | 90 minutes, weekly | Wellbeing | No effect | No significant acute wellbeing benefit of intervention vs control (Friedman Well-being Scale) | Low |
| | | | | 58 male inmates and community members (10 control - mean age 34.5; 13 'general population inmate singers' - mean age 37.9; 25 'volunteer singers' from surrounding community) | USA | Singing | One performance | Once | Wellbeing | No effect | No significant acute wellbeing benefit of intervention vs control (Friedman Well-being Scale) | Low |
| Cooke et al. | 2010 | Excluded - clinical population (dementia) | | | | | | | | | | |
| Cooke et al. | 2010 | Excluded - clinical population (dementia) | | | | | | | | | | |
| Coulton et al. | 2015 | No | RCT | 258 older adults (inclusion criteria: >/= 60 years old)(mean age 69.2; 214 female, 44 male; 27 control, 131 intervention | UK | Singing | 14 weeks | 90 minutes, weekly | Quality of Life (mental) | Positive | Significant improvement in quality of life vs control at both immediate post (3 month) and 3 month-post (SF12) | High |
| | | | | | | | | | Quality of Life (physical) | No effect | No significant effect of intervention on physical quality of life at either immediate post or 3- | High |

| | | | | | | | | | | | | |
|---|---|---|---|---|---|---|---|---|---|---|---|---|
| | | | | | | | | | | | month FU timepoints(SF12) | |
| | | | | | | | | | Anxiety | Positive | Significantly reduced anxiety in control vs intervention participants at immediate post but not 3-month FU timepoints (Hospital Anxiety and Depression Scale) | High |
| | | | | | | | | | Depression | Positive | Significantly reduced depression in control vs intervention participants at immediate post but not 3-month FU timepoints (Hospital Anxiety and Depression Scale) | High |
| Field et al. | 1997 | Excluded - music listening | | | | | | | | | | |
| Gold et al | 2014 | Excluded - multimodal music therapy approach judged to not closely resemble 'normal' performing arts participation due to prevalence of inactive modalities (i.e. verbal reflection, music listening) | | | | | | | | | | |
| Geutin et al. | 2009 | Excluded - clinical population (dementia) | | | | | | | | | | |
| Gupta & Gupta | 2005 | Excluded - music listening | | | | | | | | | | |
| Haslam et al. | 2014 | Excluded - 'music-based reminiscence' | | | | | | | | | | |
| Kim et al. | 2011 | Excluded - clinical population (stroke) | | | | | | | | | | |
| Koelsch, Offermanns & Franzke | 2010 | No | RCT | 154 adults (81 music group (improvised percussion play-along to recorded music) - mean age 24.4; 73 control group (music listening with rhythmic tapping to beat) - mean age 24.9), 78 females/76 males | Germany | Group drumming | 41 minutes | Once | Depression/ anxiety | Positive | Significant reductions in depression in music vs control groups (POMS) | High |
| | | | | | | | | | Fatigue | Positive | Significant reductions in fatigue in music vs control groups (POMS) | High |
| | | | | | | | | | Vigor | Positive | Significant increase in vigor in music vs control group (POMS) | High |
| | | | | | | | | | Mood (Irritability) | Positive | Significant reduction in irritability in music vs control group (POMS) | High |
| | | | | | | | | | Valence / positive affect | Positive | Significantly increased attractiveness of music playing vs music listening (control) | High |

| | | | | | | | | | | | | |
|---|---|---|---|---|---|---|---|---|---|---|---|---|
| | | | | | | | | | | | activity (Self-assessment manikins (SAMs)) | |
| | | | | | | | | | Arousal | Positive | Significantly increased arousal in music vs control group (Self-assessment manikins (SAMs)) | High |
| | | | | | | | | | Mood (Happiness) | Positive | Significantly increased happiness in music vs control group (Self-assessment manikins (SAMs)) | High |
| | | | | | | | | | Mood (Anger) | Positive | Significantly reduced anger in music vs control group (Self-assessment manikins (SAMs)) | High |
| | | | | | | | | | Mood (Sadness) | Positive | Significantly reduced sadness in music vs control group (Self-assessment manikins (SAMs)) | High |
| | | | | | | | | | Anxiety | Positive | Significantly reduced anxiety in music vs control group (Self-assessment manikins (SAMs)) | High |
| | | | | | | | | | Mood (Disgust) | Positive | Significantly reduced feelings of disgust in music vs control group (Self-assessment manikins (SAMs)) | High |
| Kreutz | 2014 | No | Nonrandomized experimental trial | 21 adults participating in a research choir (age range 18-65; median age ~50; 16 female, 5 male) | Germany | Group singing | 30 minutes | Once | Mood (Positive feelings) | Positive | Significantly increased positive feelings in singing group vs control (ad hoc Likert scale) | Very Low |
| | | | | | | | | | Mood (Negative feelings) | No effect | No significant differences in negative feelings between singing vs control groups (ad hoc Likert scale) | Very Low |

| | | | | | | | | | | | | |
|---|---|---|---|---|---|---|---|---|---|---|---|---|
| | | | | | | | | | Cortisol | No effect | No significant differences in cortisol changes in singing group vs control | Low |
| | | | | | | | | | DHEA | No effect | No significant differences in cortisol changes in singing group vs control | Low |
| | | | | | | | | | Cortisol-DHEA ratio | No effect | No significant differences in cortisol changes in singing group vs control | Low |
| Lee et al. | 2010 | Excluded - music listening | | | | | | | | | | |
| Lord et al. | 2012 | Excluded - clinical population (COPD) | | | | | | | | | | |
| | | | | | | | | | Wellbeing (mental) | No effect | No significant differences between music & control group (Short Warwick-Edinburgh Mental Well-being Scale) | Low |
| | | | | | | | | | Nutrition | NO effect | No significant difference between music & control (Health-Promoting Lifestyle Profile II (HPLPII)) | Low |
| Perkins & Williamon | 2014 | No | Non randomized longitudinal intervention study | 98 older adults (age >/= 50)(mean age 67.9; 30 controls, 32 music-learning (higher socioeconomic status), 36 music-learning (lower socioeconomic status)), 74 female/22 male/2 unspecified | UK | Instrumental music lessons; 'creative music workshops' | 10 weeks | 60-90 mins, weekly | Interpersonal Relations | No effect | No significant difference between music & control (Health-Promoting Lifestyle Profile II (HPLPII)) | Low |
| | | | | | | | | | Spiritual Growth | Positive | Significantly greater spiritual growth in music group (high socioeconomic status ONLY) vs control (Health-Promoting Lifestyle Profile II (HPLPII)) | Low |
| | | | | | | | | | Stress Management | No effect | No significant difference between music & control (Health-Promoting | Low |

| | | | | | | | | | | | | |
|---|---|---|---|---|---|---|---|---|---|---|---|---|
| | | | | | | | | | | | Lifestyle Profile II (HPLPII)) | |
| | | | | | | | | | Health responsibility | No effect | No significant difference between music & control (Health-Promoting Lifestyle Profile II (HPLPII)) | Low |
| Sarkamo et al. | 2014 | Excluded - clinical population (dementia) | | | | | | | | | | |
| Sole et al. | 2010 | Excluded - health outcomes only analyzed as a composite of singing, music therapy & music appreciation groups | | | | | | | | | | |
| Sun & Buys | 2012 | Excluded - clinical population (chronic conditions) | | | | | | | | | | |
| Sun et al. | 2013 | Excluded - clinical population (one or more chronic diseases) | | | | | | | | | | |
| Sole et al. | 2010 | Excluded - health outcomes only analyzed as a composite of singing, music therapy & music appreciation groups | | | | | | | | | | |
| Valentine & Evans | 2001 | | No | Non randomized experimental cohort study | 23 university singers (mean age 21.2), sex unspecified - mixed | UK | Singing | 30 minutes | Once | Blood pressure | No effect | No significant pre-post changes | Very Low |
| | | | | | | | | | Mood (tense arousal) | Positive | Significant pre-post improvement in singing group, although not to the same magnitude as in swimming group (UWIST model adjective checklist (UMACL) | Very Low |
| | | | | | | | | | Mood (energetic arousal) | Positive | Significant pre-post improvement in singing group, although not to the same magnitude as in swimming group (UWIST model adjective checklist (UMACL) | Very Low |
| | | | | | | | | | Mood (hedonic tone) | Positive | Significant pre-post improvement in singing group, although not to the same magnitude as in swimming group (UWIST model adjective checklist (UMACL) | Very Low |
| Warth et al. | 2015 | Excluded - clinical population (cancer) | | | | | | | | | | |

| | | | | | | | | | | | | | |
|---|---|---|---|---|---|---|---|---|---|---|---|---|---|
| Wlodarczyk | 2007 | Excluded - clinical population (hospice) | | | | | | | | | | | |
| Wu | 2002 | Excluded - clinical population (depression) | | | | | | | | | | | |
| ***Eells, 2014*** | | | | | | | | | | | | | |
| Chang et al. | 2008 | Excluded - music listening | | | | | | | | | | | |
| Clift et al. | 2008 | Excluded - review | | | | | | | | | | | |
| Cooke et al. | 2010 | Excluded - clinical population (dementia) | | | | | | | | | | | |
| Guetin et al. | 2009 | Excluded - clinical population (dementia) | | | | | | | | | | | |
| Hays and Minichiello | 2005 | Excluded - qualitative results only | | | | | | | | | | | |
| Hicks-Moore | 2005 | Excluded - music listening | | | | | | | | | | | |
| Houston et al. | 1998 | Other review - Clift et al., 2010 | | | | | | | | | | | |
| Lee, Chan & Mok | 2010 | Excluded - music listening | | | | | | | | | | | |
| Lesta and Petocz | 2006 | Excluded - clinical population (dementia) | | | | | | | | | | | |
| McCaffrey and Freeman | 2003 | Excluded - music listening | | | | | | | | | | | |
| McCaffrey and Locsin | 2006 | Excluded - music listening | | | | | | | | | | | |
| Mok and Wong | 2003 | Excluded - music listening | | | | | | | | | | | |
| Pickles | 2003 | Excluded - qualitative results only | | | | | | | | | | | |
| Skingley and Bungay | 2010 | Excluded - qualitative results only | | | | | | | | | | | |
| Sung et al. | 2010 | Excluded - music listening | | | | | | | | | | | |
| Twiss et al. | 2006 | Excluded - music listening | | | | | | | | | | | |
| ***Fancourt, Ockelford & Belai, 2014*** *(**'active participation' studies only**)* | | | | | | | | | | | | | |
| Bittman et al | 2001 | | Yes (Remo Drums) | RCT | 60 adults (mean age ~30 (precise age unspecified); 31 men, 29 women): musical experience unspecified | USA | Group drumming | 1 hour | Once | Anxiety | No effect | No effect of group drumming vs. control in anxiety (Beck Anxiety Inventory) | Low |
| | | | | | | | | | | Depression | No effect | No effect of group drumming vs. control in depression (Beck Depression Inventory II) | Low |

| | | | | |
|---|---|---|---|---|
| | White blood cell count | No effect | No effect of group drumming vs. control on white blood cell count | Low |
| | Serum cortisol | No effect | No effect of group drumming vs. Control on cortisol | Low |
| | NK cell activity | Positive | Increased NK activity vs. Control at effector:target ratios of 6:1 & 12:1; no significant change in NK activity (vs control) at effector:target ratios of 25:1 & 50:1 | Low |
| | LAK cell activity | Positive | Increased LAK activity vs control in 2 conditions (with IFN-alpha at effector:target of 12:1 & 25:1); No change in LAK activity vs control in 2 conditions (with IL-2 at effector:target of 12:1 and 25:1); **potential confounder →** significant variability in change in baseline IFN-alpha levels pre-post in experimental vs control groups at effector:target 12:1 & 25:1 | Low |
| | IL-2 | No effect | No effect of group drumming vs control on IL-2 (a posteriori measurement) | Low |
| | IFN-alpha | No effect | No effect of group drumming vs control on IFN-alpha (a posteriori measurement) | Low |
| | Plasma DHEA | No effect | No effect of group drumming vs control on plasma DHEA | Low |

| | | | | | | | | | | | | | |
|---|---|---|---|---|---|---|---|---|---|---|---|---|---|
| | | | | | | | | | | DHEA to cortisol ratio | No effect | No effect of group drumming vs control on DHEA to cortisol ratio; *positive touted effect only within drumming group – not significant vs control | Low |
| Cai et al. | 2001 | Excluded - clinical population (cancer) | | | | | | | | | | | |
| Koyama et al. | 2009 | | Yes (Yamaha Corp) | Single group study (uncontrolled) | 27 young adults (mean age 27.9; 19 women, 8 men); no drumming experience | Japan | Group drumming | 1 hour | Once | Tension/anxiety | Positive | Reduction pre-post drumming intervention (POMS subscale) | Very Low |
| | | | | | | | | | | Depression/dejection | Positive | Reduction pre-post drumming intervention (POMS subscale) | Very Low |
| | | | | | | | | | | Anger/hostility | Positive | Reduction pre-post drumming intervention (POMS subscale) | Very Low |
| | | | | | | | | | | Vigor/activity | No effect | No change pre-post drumming intervention (POMS subscale) | Very Low |
| | | | | | | | | | | Fatigue/inertia | Positive | Reduction pre-post drumming intervention (POMS subscale) | Very Low |
| | | | | | | | | | | Confusion/bewilderment | Positive | Reduction pre-post drumming intervention (POMS subscale) | Very Low |
| | | | | | | | | | | Total mood disturbance | Positive | Reduction pre-post drumming intervention (total POMS) | Very Low |
| | | | | | | | | | | Adrenaline | No effect | | Very Low |
| | | | | | | | | | | Noradrenaline | No effect | | Very Low |
| | | | | | | | | | | NK cell activity | No effect | | Very Low |
| | | | | | | | | | | White blood cells | No effect | | Very Low |
| | | | | | | | | | | Neutrophils | No effect | | Very Low |
| | | | | | | | | | | Lymphocytes | No effect | | Very Low |
| | | | | | | | | | | T cells | No effect | | Very Low |
| | | | | | | | | | | B cells | No effect | | Very Low |
| | | | | | | | | | | CD4+ T cells | No effect | | Very Low |
| | | | | | | | | | | CD8+ T cells | No effect | | Very Low |
| | | | | | | | | | | CD4/CD8 ratio | No effect | | Very Low |

| | | | | | | | | | |
|---|---|---|---|---|---|---|---|---|---|
| | | | | | | Naive T cells | No effect | | Very Low |
| | | | | | | Memory T cells | No effect | | Very Low |
| | | | | | | Naive/memory cell ratio | No effect | | Very Low |
| | | | | | | NK cells (count | No effect | | Very Low |
| | | | | | | IFN-alpha production | No effect | | Very Low |
| | | | | | | IL-2 production | No effect | | Very Low |
| | | | | | | IL-4 production | No effect | | Very Low |
| | | | | | | IL-6 production | No effect | | Very Low |
| | | | | | | IL-10 production | No effect | | Very Low |
| | 27 older adults (mean age 70.3; 15 women, 12 men); no drumming experience | Japan | Group drumming | 1 hour | Once | Tension/anxiety | Positive | Reduction pre-post drumming intervention (POMS subscale) | Very Low |
| | | | | | | Depression/ dejection | Positive | Reduction pre-post drumming intervention (POMS subscale) | Very Low |
| | | | | | | Anger/hostility | Positive | Reduction pre-post drumming intervention (POMS subscale) | Very Low |
| | | | | | | Vigor/activity | No effect | No change pre-post drumming intervention (POMS subscale) | Very Low |
| | | | | | | Fatigue/inertia | No effect | No change pre-post drumming intervention (POMS subscale) | Very Low |
| | | | | | | Confusion/ bewilderment | No effect | No change pre-post drumming intervention (POMS subscale) | Very Low |
| | | | | | | Total mood disturbance | Positive | Reduction pre-post drumming intervention (full POMS) | Very Low |
| | | | | | | Adrenaline | No effect | | Very Low |
| | | | | | | Noradrenaline | Positive | Significant pre-post increase | Very Low |
| | | | | | | NK cell activity | No effect | | Very Low |
| | | | | | | White blood cells | No effect | | Very Low |
| | | | | | | Neutrophils | No effect | | Very Low |
| | | | | | | Lymphocytes | Positive | Significant pre-post increase | Very Low |

| | | | | | | | | | | | | |
|---|---|---|---|---|---|---|---|---|---|---|---|---|
| | | | | | | | | | T cells | Positive | Significant pre-post increase | Very Low |
| | | | | | | | | | B cells | No effect | | Very Low |
| | | | | | | | | | CD4+ T cells | Positive | Significant pre-post increase | Very Low |
| | | | | | | | | | CD8+ T cells | No effect | | Very Low |
| | | | | | | | | | CD4/CD8 ratio | No effect | | Very Low |
| | | | | | | | | | Naive T cells | No effect | | Very Low |
| | | | | | | | | | Memory T cells | Positive | Significant pre-post increase | Very Low |
| | | | | | | | | | Naive/memory ratio | No effect | | Very Low |
| | | | | | | | | | NK cells (count | No effect | | Very Low |
| | | | | | | | | | IFN-alpha production | Positive | Significant pre-post increase | Very Low |
| | | | | | | | | | IL-2 production | No effect | | Very Low |
| | | | | | | | | | IL-4 production | No effect | | Very Low |
| | | | | | | | | | IL-6 production | Positive | Significant pre-post increase | Very Low |
| | | | | | | | | | IL-10 production | No effect | | Very Low |
| Kreutz et al. | 2004 | No | Non-randomized intervention study (week 1 – singing; week 2 – listening (control)) | 31 members of an amateur choir (mean age = 56.9, range 29-74), 23 female/8 male | Germany | Singing | 1 hour | Once | Positive affect | Positive | Significant pre-post increase in positive affect in singing group vs control (Positive and Negative Affect Schedule (PANAS)) | Low |
| | | | | | | | | | Negative affect | Positive | Significant pre-post decrease in negative affect vs control (Positive and Negative Affect Schedule (PANAS)) | Low |
| | | | | | | | | | Serum immunoglobulin A / albumin | Positive | Significant pre-post increase in S-IgA/albumin vs control | Low |
| | | | | | | | | | Cortisol | No effect | No significant pre/post effect of singing on cortisol | Low |
| Kuhn | 2002 | No | RCT | 33 undergraduate students (mean age = 20; 5 men, 28 women); 6 | USA | Group music making (singing and drumming) | 30 minutes | Once | Serum Immunoglobulin-A | Positive | Significant increase in S-IgA levels in active music group vs inactive | High |

| | | | | | | | | | | | | |
|---|---|---|---|---|---|---|---|---|---|---|---|---|
| | | | | participants with >1 year formal music training at high school/college level – group allocation unclear) | | | | | | | (listening) & control (no activity) groups | |
| Lane | 1994 | Excluded - clinical population (hospitalized patients) | | | | | | | | | | |
| Lindblad, Hogmark & Theorell | 2007 | No | Non-randomized longitudinal cohort study | 60 5th & 6th graders, age & sex unspecified | Sweden | Music education | 8 months | 1 hour, weekly | Cortisol | No effect | No effect of music education on cortisol levels from saliva samples collected at any timepoint vs control (significant within group change in music group in afternoon sample only) | Low |
| | | | | | | | | | Self-esteem | No effect | No effect of music education on self-esteem (within or between group) ('I think I am' questionnaire) | Low |
| | | | | | | | | | Social anxiety | No effect | No effect of music education on social anxiety (within or between group) (Social Anxiety Scale for Children-Revised (SASC-R) & Social Phobia and Anxiety Inventory-Parental (SPAI-P)) | Low |
| | | | | | | | | | Social functioning | No effect | No effect of music education on self-esteem (within or between group) (Child Behaviour Check List (CBCL), completed by parents) | Low |
| Okada et al | 2009 | Excluded - clinical population (cerebrovascular disease/dementia) | | | | | | | | | | |
| Suzuki et al. | 2005 | Excluded - clinical population (dementia) | | | | | | | | | | |
| ***Gick, 2012*** | | | | | | | | | | | | |

| | | | | | | | | | | | | |
|---|---|---|---|---|---|---|---|---|---|---|---|---|
| Beck et al | 2006 | No | Single group cohort study | 10 conservatory music majors (mean age 20.8; average 10.1 years singing), sex unspecified | USA | Singing | One rehearsal/ performance | Data collections from 4-11 rehearsals/performances over a 10 week period | Immunoglobulin A | Positive | Significant increase in IgA during performance and rehearsals | Very Low |
| Beck et al | 2000 | Other review - Clift et al., 2010 | | | | | | | | | | |
| Bygren et al | 2009 | Excluded – combined analysis of receptive (film, music or art events) & active (choir singing) arts participation | | | | | | | | | | |
| Clift & Hancox | 2001 | Excluded - descriptive, uncontrolled survey-based cross-sectional study | | | | | | | | | | |
| Clift et al | 2007 | Excluded - descriptive, uncontrolled survey-based cross sectional study of active participation | | | | | | | | | | |
| Cohen et al | 2006 | Other review - Daykin et al., 2018 | | | | | | | | | | |
| Fechir et al | 2008 | Excluded – descriptive analysis of the demands and response to singing with incompatible outcomes | | | | | | | | | | |
| Grape et al | 2003 | No | Cross-sectional | 16 singers (8 professionals – mean age 40.2, 4 male/4 female; 8 amateurs – mean age 40.2, 2 male/6 female) | Sweden | Singing | 45 min | Once | Cortisol | No effect | No significant time or group*time interactions | Very Low |
| | | | | | | | | | Heart rate variability | Mixed | Significant group * time interaction – increased HR variability (total power) in professionals, decreased HR variability in amateurs | Very Low |
| | | | | | | | | | TNF-alpha | Mixed | Significant group*time effect; increased TNF-alpha in professionals, but not amateurs | Very Low |
| | | | | | | | | | Positive affect | Positive | Significant increase in energy & feelings of joy and relaxation in both amateurs and professionals; significantly greater feelings of joy in amateurs vs professionals | Very Low |
| Harmat & Theorell | 2010 | Excluded – descriptive analysis of HR responses to music performance only (included in HR data) | | | | | | | | | | |
| Hillman | 2002 | Excluded - descriptive, uncontrolled survey-based cross sectional study | | | | | | | | | | |
| Houston et al | 1998 | Other review - Clift et al., 2010 | | | | | | | | | | |
| Kenny, David & Oates | 2004 | Excluded - descriptive, uncontrolled survey-based cross sectional study of anxiety in professional singers | | | | | | | | | | |
| Pai et al | 2008 | No | Cross-sectional | 107 adults (52 semiprofessional | UK | Singing | Mean 30.8 years singing | Unspecified | Daytime somnolence | No effect | No significant differences between | Low |

| | | | | | | | | | | | | |
|---|---|---|---|---|---|---|---|---|---|---|---|---|
| | | | | singers (mean age 46.3; 20 males, 36 females); 55 non-singers (mean age 43.3; 23 males, 32 females)) | | | | | | | singing & non-singing groups (Epworth Sleepiness Scale) | |
| Silber | 2005 | Excluded – qualitative outcomes only | | | | | | | | | | |
| Skingley & Bungay | 2010 | Excluded – qualitative outcomes only | | | | | | | | | | |
| Unwin, Kenny & Davis | 2002 | Other review - Clift et al., 2010 | | | | | | | | | | |
| Valentine & Evans | 2001 | Other review - Daykin et al., 2018 | | | | | | | | | | |
| Wise, Hartmann & Fisher | 1992 | Other review - Clift et al., 2010 | | | | | | | | | | |
| ***Gordon, Fehd & McCandliss, 2015*** | | | | | | | | | | | | |
| Bolduc & Lefebvre | 2012 | No | Non-randomized experimental study | 54 kindergarten students (mean age = 4.8; 26 language group, 28 music and language group), 27 male/27 female) | Canada | Music education (general) | 10 weeks | 40 min/weekly | Phonological awareness (other) | Meta-analysis | Phonological awareness measure | Low |
| Cogo-Moreira et al. | 2013 | No | RCT | 235 elementary school children with below average reading ability (mean age = 9.15; 114 music education group, 121 control group), 38.3% female | Brazil | Music education (general) | 5 months | 50 min/3x weekly | Reading fluency | Meta-analysis | Test of phonological awareness | Moderate |
| | | | | | | | | | Phonological awareness (other) | Meta-analysis | Accuracy of word reading (custom measure) | Moderate |
| Dege & Schwarzer | 2011 | No | RCT | 41 pre-schoolers, 17 female/22 male; age 5-6 | Germany | Music education (general) | 20 weeks | 10 min, daily | Phonological awareness (other) | Meta-analysis | Total of 4 subtests of Bielefelder Screening | Moderate |
| Gromko | 2005 | No | Non-randomized longitudinal cohort study | 103 kindergarten children (across 2 schools; 43 treatment, 60 control; sex unspecified - mixed) | USA | Music education (general - singing & percussion focus) | 4 months | 30 min, weekly | Reading fluency | Meta-analysis | DIBELS letter naming fluency | Very Low |
| | | | | | | | | | Phonological awareness (other) | Meta-analysis | DIBELS phoneme-segmentation fluency | Very Low |

| | | | | | | | | | | | | |
|---|---|---|---|---|---|---|---|---|---|---|---|---|
| | | | | 29 preschool children (mean age = 4.5), sex breakdown unspecified – mixed | Spain | Music education (general) | 8 weeks | 1 hour/2x weekly | Phonological awareness (rhyming) | Meta-analysis | Rhyme oddity task (custom) | Moderate |
| | | | | | | | | | Phonological awareness (other) | Meta-analysis | Rhyme oddity task (custom) | Moderate |
| Herrera et al | 2011 | No | RCT | 27 preschool children (mean age = 4.5), sex breakdown unspecified – mixed | | | | | Phonological awareness (other) | Meta-analysis | Initial phoneme oddity task (custom) | Moderate |
| | | | | | | | | | Phonological awareness (rhyming) | Meta-analysis | Rhyme oddity task (custom) | Moderate |
| Moreno et al. | 2011 | No | RCT | 60 children (age range 4-6; 30 music group – 12 male/18 female; 30 visual art group – 14 male/16 female) | Canada | Music education (computer administered general music education) | 4 weeks | 2, 1-hour sessions/5x weekly | Phonological awareness (rhyming) | Meta-analysis | Rhyming task from WJ-III | High |
| Moreno et al. | 2009 | No | RCT (pseudo?) | 32 3rd graders (mean age = 8.3), 19 male/13 female | Portugal | Music education (general) | 24 weeks | 75 min/2x weekly | Reading fluency | Meta-analysis | Reading inconsistent words (from Portuguese European reading battery) | Moderate |

| | | | | | | | | | | | | |
|---|---|---|---|---|---|---|---|---|---|---|---|---|
| Moritz et al | 2013 | No | Cross-sectional | 30 kindergarteners (mean age = 5.6), 17 male/13 female | USA | Music education (general) | Unspecified | 45 min/daily | Phonological awareness (rhyming) | Meta-analysis | Rhyming discrimination from Phonological Awareness Test (PAT) | Low |
| | | | | | | | | | Phonological awareness (other) | Meta-analysis | Isolation of initial phonemes from PAT | Low |
| Myant, Armstrong & Healy | 2008 | No | Non-randomized experimental study | 59 children (mean age = 4.3), sex unavailable (no article access) | UK | Music education (general) | No article access | No article access | Phonological awareness (rhyming) | Meta-analysis | Rhyme test from Phonological Assessment Battery (PhAB) | Low |
| | | | | | | | | | Phonological awareness (other) | Meta-analysis | Alliteration test from PhAB | Low |
| Register | 2004 | No | Non-randomized experimental study | 43 Kindergarten students (age 5-7), sex unspecified – mixed | USA | Music education (therapy focus) | 15+ sessions | 30 min/session | Reading fluency | Meta-analysis | Letter-naming fluency from DIBELS | Low |
| | | | | | | | | | Phonological awareness (other) | Meta-analysis | Initial sounds fluency from DIBELS | Low |
| Thomson, Leong & Goswami | 2013 | No | Non randomized experimental study | 21 dyslexic children (mean age = 9.3), sex unspecified | UK | Rhythm intervention | 6 weeks | 30 min/weekly | Reading fluency | Meta-analysis | TOWRE | Low |
| | | | | | | | | | Phonological awareness (rhyming) | Meta-analysis | Rhyme test from Phonological Assessment Battery (PhAB) | Low |
| | | | | | | | | | Phonological awareness (other) | Meta-analysis | Spoonerisms from PhAB | Low |
| Yazejian & Peisner-Feinberg | 2009 | No | Non randomized experimental study | 181 Head Start children (mean age = 4.4), sex breakdown unspecified – mixed | USA | Music & movement intervention | 26 weeks | 30 min/2x weekly | Phonological awareness (rhyming) | Meta-analysis | Rhyming from Early Phonological Awareness Profile (EPAP) | Low |
| | | | | | | | | | Phonological awareness (other) | Meta-analysis | Phoneme deletion from EPAP | Low |
| ***Hetland, 2000*** | | | | | | | | | | | | |
| Bilhartz, Terry & Olson | 2000 | Yes (Kinder-musik Inter-national) | Non randomized experimental cohort study | 66 children (4-5 years old), sex unspecified - mixed | USA | Kindermusic | 30 weeks | 75 min, weekly | Spatial ability | Meta-analysis | | Low |
| Costa-Giomi | 1999 | No | Quasi-randomized longitudinal cohort study | 81 4th graders; age 9 at start of intervention (58 girls, 59 boys) | Canada | Piano lessons | 3 years | 30-45 minutes, weekly (instruction time only) | Spatial reasoning | Meta-analysis | | Very Low |
| Flohr et al | 1998 | No | RCT | 22 children (aged 4-6), sex unspecified | USA | General music education | 7 weeks | 25 minutes, twice weekly | Spatial reasoning | Meta-analysis | | High |

| | | | | | | | | | | | |
|---|---|---|---|---|---|---|---|---|---|---|---|
| Flohr, Miller and Persellin | 1998 | | RCT | 19 children (4-5 year old), sex unspecified | USA | General music education | 10 weeks | 25 minutes, twice weekly | Spatial reasoning, spatial ability | Meta-analysis | High |
| Flohr, Miller and Persellin | 2000 | | RCT | 20 children (age 4), sex unspecified | USA | General music education | 16-18 weeks | 25 minutes, twice weekly | Spatial reasoning | Meta-analysis | High |
| Graziano, Peterson & Shaw | 1999 | Yes (Yamaha Corp) | Non randomized experimental cohort study | 53 2nd graders (age 6-8), sex unspecified | USA | Piano lessons | 14 weeks | 60 minutes, 3x/week | Spatial reasoning | Meta-analysis | Low |
| Gromko & Poorman | 1998 | No | Non randomized experimental cohort study | 30 pre-schoolers (age 3-4), sex unspecified | USA | General music education | 24 weeks | 30 minutes, weekly | Spatial reasoning, spatial ability | Meta-analysis | Low |
| Hurwitz et al | 1975 | No | Retrospective cross sectional analysis | 40 children (age 6-7), 20 male/20 female | USA | Kodaly music instruction | 7 months | 40 minutes, 5x/week | Spatial reasoning, intelligence, spatial ability | Meta-analysis | Very Low |
| Lazco | 1985 | No | Longitudinal cohort study | 301 children (2 groups – 154 3rd graders (age 9-10); 147 6th graders (age 12-13)), sex unspecified | Hungary | Kodaly music instruction | Unspecified | 4-6 classes, weekly | Intelligence | Meta-analysis | Low |
| Mallory & Philbrick | 1995 | Unknow n | Longitudinal cohort study | 44 children (3-5 years old), sex unspecified | USA | General music education | 6 months | 60 min, weekly | Spatial reasoning | Meta-analysis | Low |
| Parente & O'Malley | 1975 | No | Non randomized experimental cohort study | 24 children (5-9 years old), 12 male/12 female | USA | Snare drum | 4 weeks | 60 minutes, 2x/weekly | Spatial ability | Meta-analysis | Low |
| Persellin | 1999 | Unknow n | Longitudinal cohort study | 12 children (5-6 years old), sex unspecified | USA | General music education | 6 weeks | 45 minutes, 3x/week | Spatial reasoning | Meta-analysis | Low |
| Rauscher | 1999 | Unknow n | RCT | 66 children (5-6 years old), sex unspecified | USA | General music education | 8 months | 40 minutes, 1x/week | Spatial reasoning | Meta-analysis | High |
| Rauscher | 1999 | Unknow n | RCT | 87 children (age 3-6), sex unspecified | USA | General music education | Two 6-month periods over 2 years | 10 min, weekly | Spatial reasoning | Meta-analysis | High |
| Rauscher et al | 1994 | Unknow n | Longitudinal cohort study | 33 children (3-4 years old), sex unspecified | USA | Voice & Piano lessons | 8 months | 30 minutes, 3x/week | Spatial reasoning | Meta-analysis | Low |
| Rauscher et al | 1997 | Yes (Yamaha Corp) | Longitudinal cohort study | 78 children (3-4 years old), 42 male/36 female | USA | Voice & Piano lessons | 6-8 months | 10 min, weekly | Spatial reasoning, spatial ability | Meta-analysis | Low |
| Ruascher & Zupan | 1999 | No | Longitudinal cohort study | 48 children (5-7 years old), sex | USA | General music education | Two 8-month periods over 2 years | 20 minutes, 2x/week | Spatial reasoning | Meta-analysis | Low |

| | | | | | | | | | | | | |
|---|---|---|---|---|---|---|---|---|---|---|---|---|
| | | | | breakdown unspecified - mixed | | | | | | | | |
| Taetle | 1999 | No | Longitudinal cohort study | 68 children (5-6 years old), sex unspecified | USA | Orff, singing | 4 months | 30 min, 2x/week | Spatial reasoning, spatial ability | Meta-analysis | | Low |
| Zulauf | 1993 | No | Longitudinal cohort study | 353 adolescents (mean age unspecified), 174 female/179 male | Switzerland | General music education | 3 years | 5x/weekly (school days) | Intelligence, spatial ability | Meta-analysis | | Low |
| ***Jaschke et al., 2013*** | | | | | | | | | | | | |
| Costa-Giomi | 2004 | No | RCT | 117 4th graders; 58 girls/59 boys; age 8-12 | Canada | Piano instruction | 3 years | 30-45 minutes, weekly (instruction time only) | Language processing | No effect | No significant differences in language processing or mathematical achievement (Canadian Achievement Test 2) | Moderate |
| | | | | | | | | | Mathematical ability | No effect | No significant differences in language processing or mathematical achievement (Canadian Achievement Test 2) | Moderate |
| Courey et al. | 2012 | Excluded - not 'music participation', targeted music theory instruction to teach fractions | | | | | | | | | | |
| Dege & Schwarzer | 2011 | Other review - Gordon, Fehd & McCandliss, 2015 | | | | | | | | | | |
| Gromko | 2005 | No | Non-randomized longitudinal cohort study | 103 kindergarten children (across 2 schools; 43 treatment, 60 control; sex unspecified - mixed) | USA | Music education (general - singing & percussion focus) | | 30 min, weekly | Reading ability | No effect | No effect of music training on nonsense-word fluency (Dynamic Indicators of Basic Early Literacy Skills (DIBELS) test) vs. control | Very Low |
| | | Yes - Gordon, Fehd & McCandliss, 2015 | | | | | | | Reading ability | | (Dynamic Indicators of Basic Early Literacy Skills (DIBELS) test - letter naming & phoneme segmentation subtest) | |
| Ho, Cheung and Chan | 2003 | No | Cross-sectional | 90 male right handed participants (average age ~11) | Hong Kong | Classical instrumental education | 1-5 years (music education experience) | 1+ hour, weekly | IQ | No effect | No significant differences in IQ between individuals with and without music | Low |

| | | | | | | | | | | | | |
|---|---|---|---|---|---|---|---|---|---|---|---|---|
| | | | | | | | | | | | training (Hong Kong Weschler Intelligence Scale for Children) | |
| Jentschke, Koelsch & Friederici | 2005 | | No | Cross-sectional | 28 participants (14 with music training experience; 14 without); age 10-12, 15 male/13 female | Germany | Instrumental or singing lessons | Unspecified | Unspecified | Reading ability | No effect | No significant differences between trained & untrained children on verbal scale of WISC-III | Very Low |
| Piro and Ortiz | 2009 | | No | Non-randomized longitudinal cohort study | 103 2nd grade elementary school children (mixed gender) in their 3rd year of piano instruction | USA | Piano instruction | 10 months | 40-45 min, twice weekly | Reading ability | Positive | Improved reading (vocabulary and verbal sequencing subtests of the Meeker Structure of Intellect) in music training group, but not control (no music) group | Very Low |
| Portowitz et al. | 2009 | | No | Non-randomized longitudinal cohort study | 81 'high-risk' elementary schoolchildren, aged 6-12 years: sex unspecified | Israel | Multimodal music education (music appreciation, private instrumental instruction, ensemble performances) | 2 years | 2-3 hours, weekly (split up) | IQ | Positive | Improved general IQ (Raven assessment) in music training group | Low |
| Register | 2001 | | No | Non-randomized longitudinal cohort study | 50 participants (25 control; 25 music training); 61 originally involved but 11 drop outs not analyzed; age 4-5; sex unspecified - mixed | USA | Music therapy sessions' (experimental group focused on teaching/ reinforcing prereading/pre-writing concepts; control group - general 'music therapy') | 30 weeks | 30 min, twice weekly | Writing ability | Positive | Improvements in both music groups in 'prewriting' (Print Awareness Test of Word Identification) | Very Low |
| | | | | | | | | | | Reading ability | Positive | Improvements in both music groups in 'prereading' (The Print Awareness Test for Logos; Print Concepts Checklist) | Very Low |
| Rickard, Bambrick & Gill | 2012 | | No | RCT | 84 7th graders (47 music training group; 37 control (drama lessons); aged 10-13, sex unspecified - mixed | Australia | General music education | 6.5 months | Unspecified | IQ | No effect | No significant differences in music vs. control in non-verbal IQ (Kaufman Brief Intelligence Test) | Low |
| | | | | | | | | | | Writing ability | No effect | No significant differences in music vs. | Low |

| | | | | | | | | | | | | |
|---|---|---|---|---|---|---|---|---|---|---|---|---|
| | | | | | | | | | | | control in verbal IQ (Kaufman Brief Intelligence Test) | |
| Rickard, Bambrick & Gill | 2012 | No | RCT | 69 5th and 6th graders (38 in music group; 31 in control (no intervention)); sex unspecified - mixed | Australia | General music education | 5 months | 1 hour, weekly | Reading ability | Negative | Significantly worse improvement in reading skills (Victorian Education Learning Standards (VELS)) in music intervention vs. control | High |
| | | | | | | | | | Mathematical ability | Positive | Significantly greater improvement in mathematical achievement (VELS) in music group vs. control | High |
| Schellenberg | 2004 | No | RCT | 144 6-year old children (36 keyboard lessons, 36 voice lessons, 36 drama lessons, 36 control (waitlist)); sex unspecified - mixed | Canada | Keyboard or singing lessons | 36 weeks | Unspecified | IQ | Positive | Significantly greater improvement in IQ in music training groups vs. controls (drama & no training)(WISC-III) | High |
| | | | | | | | | | Mathematical ability | No effect | No significant differences between music training & control groups in mathematical achievement (Kaufman Test of Educational Achievement (K-TEA) mathematical applications and computation subscales) | High |
| | | | | | | | | | Reading ability | No effect | No significant differences between music training & control groups in reading achievement (Kaufman Test of Educational Achievement (K-TEA) reading decoding, reading comprehension & spelling subscales) | High |
| Tsang & Conrad | 2011 | No | Cross-sectional | 69 children between the ages of 5 and 9 | Canada | Formal music lessons (21 | Average 1.27 years (range .5 | Unspecified | Cognitive - auditory | Positive | Significantly better performance by music | Very Low |

| | | | | | | | | | | | | |
|---|---|---|---|---|---|---|---|---|---|---|---|---|
| | | | | (43 control; 26 with music training; sex unspecified - mixed) | | piano, 2 violin, 2 voice, 1 voice & cello) | - 5 years) of music training | | | | trained children on phonological skills test (Test of Auditory Analysis Skills) | |
| | | | | | | | | | Reading ability | No effect | No significant differences in trained vs. untrained children in receptive vocabulary (Woodcock Reading Mastery Test) | Very Low |
| | | | | | | | | | Reading ability | No effect | No significant differences between trained and untrained individuals on word identification(Peabody Picture Vocabulary Test) | Very Low |
| ***Moore, 2013*** | | | | | | | | | | | | |
| Ball et al. | 2007 | Excluded - music listening | | | | | | | | | | |
| Baumgartner et al. | 2006 | Excluded - music listening | | | | | | | | | | |
| Bengtsson et al | 2007 | No | Cross-sectional | 11 professional male Swedish concert pianists (mean age = 32) | Sweden | Piano | 40 second trials ('improvise' & 'reproduce') | Once | Emotional Regulation | Unclear | Frontal & temporal association areas activated when musicians improvised; dlPFC implicated in creative aspect of behavior when adapted to satisfy a result | Very Low |
| Berkowitz & Ansari | 2008 | No | Cross-sectional | 12 classically trained undergraduate pianists (mean age = 21.9), 8 female/4 male | USA | Piano | 23.3 minutes | Once | Emotional regulation (Anterior Cingulate Cortex Activation) | Positive | Increased ACC activation during melodic and rhythmic improvisation | Very Low |
| Berns & Moore | 2012 | Excluded - music listening | | | | | | | | | | |
| Berns et al. | 2010 | Excluded - music listening | | | | | | | | | | |
| Blood & Zatorre | 2001 | Excluded - music listening | | | | | | | | | | |
| Brown & Martinez | 2006 | Excluded - music listening | | | | | | | | | | |
| Brown et al | 2004 | Excluded - music listening | | | | | | | | | | |

| | | | | | | | | | | | | |
|---|---|---|---|---|---|---|---|---|---|---|---|---|
| Brown, Martinez & Parsons | 2006 | No | Cross-sectional | 10 amateur musicians (mean age 25 years), 5 male/5 female | USA | Singing | 8.4 minutes | Once | Emotional regulation (Anterior Cingulate Cortex Activation) | Positive | Increased ACC activation during melodic improvisation | Very Low |
| Callan et al | 2006 | No | Cross-sectional | 16 adults with no significant musical experience (mean age = 26), 5 female/11 male | Japan | Singing | Unclear (48 bars) | Once | Emotional regulation (Orbitofrontal cortex activation) | No effect | No significant increase in OFC activation during singing vs speaking (increased activation during listening to singing only) | Very Low |
| Coen et al. | 2009 | Excluded - music listening | | | | | | | | | | |
| de Manzano & Ullen | 2012 | No | Cross-sectional | 18 right handed classical concert pianists (mean age = 39; 1 female, 17 male) | Sweden | Piano | 20 minutes | Once | Emotional regulation (Anterior cingulate cortex & dorsolateral prefrontal cortex activation) | Positive | Increased cortical activation associated with response inhibition and selection but unclear implications for emotional regulation | Very Low |
| Dyck et al. | 2011 | Excluded - music listening | | | | | | | | | | |
| Eldar et al. | 2007 | Excluded - music listening | | | | | | | | | | |
| Flores-Gutierrez et al | 2007 | Excluded - music listening | | | | | | | | | | |
| Ford et al. | 2011 | Excluded - music listening | | | | | | | | | | |
| Foss et al | 2007 | Excluded - music listening | | | | | | | | | | |
| Fujisawa & Cook | 2011 | Excluded - music listening | | | | | | | | | | |
| Green et al. | 2008 | Excluded - music listening | | | | | | | | | | |
| Hugdahl et al. | 1999 | Excluded - music listening | | | | | | | | | | |
| James et al. | 2008 | Excluded - music listening | | | | | | | | | | |
| Jenata | 2009 | Excluded - music listening | | | | | | | | | | |
| Jeffries, Fritz & Braun | 2003 | No | Cross-sectional | 20 adults with no musical experience (mean age = 34), 8 female/12 male | USA | Singing | Unclear | Once | Emotional regulation (dorsolateral prefrontal cortex activation) | Positive | Increased dorsolateral prefrontal cortex activation in singing vs speaking | Very Low |
| Jerde et al. | 2011 | Excluded - music listening | | | | | | | | | | |
| Kleber et al | 2010 | No | Cross-sectional | 49 adults with varied singing experience (10 professional opera | Germany | Singing | 11 min | Once | Emotional regulation (anterior cingulate cortex, | Positive | Increased dlPFC activation associated with increasing singing practice; increased | Very Low |

| | | | | | | | | | | | | |
|---|---|---|---|---|---|---|---|---|---|---|---|---|
| | | | | singers (mean age = 38; 7 females, 3 males); 21 vocal students (mean age 25 years; 14 females, 7 males); 18 laymen (mean age 23.5 years; 6 females, 12 males)) | | | | | ventrolateral prefrontal cortex, amygdala activation) | | activation of ACC in university singers | |
| Kleber et al | 2007 | No | Cross-sectional | 16 classical singers (mean age 31.1; 5 male, 11 female) | Germany | Singing | Unclear | Once | Emotional regulation (dorsolateral prefrontal cortex activation) | Unclear | Increased dlPFC activation during imagined vs overt singing | Very Low |
| Knösche et al | 2005 | Excluded - music listening | | | | | | | | | | |
| Koelsch et al. | 2006 | Excluded - music listening | | | | | | | | | | |
| Koelsch et al. | 2008 | Excluded - music listening | | | | | | | | | | |
| Lee et al. | 2011 | Excluded - music listening | | | | | | | | | | |
| Lerner et al. | 2009 | Excluded - music listening | | | | | | | | | | |
| Limb & Braun | 2008 | No | Cross-sectional | 6 right-handed male professional pianists (mean age = 34.2) | USA | Piano | 34.3 minutes | Once | Emotional regulation (orbital prefrontal cortex, dorsolateral prefrontal cortex & amygdala activation) | Mixed | Increased OFC & amygdala activation during improvisation; however, also decreased activation in dlPFC during improvisation | Very Low |
| Menon & Levitin | 2005 | Excluded - music listening | | | | | | | | | | |
| Mitters-Chiffthaler et al. | 2007 | Excluded - music listening | | | | | | | | | | |
| Mizuno & Sugishita | 2007 | Excluded - music listening | | | | | | | | | | |
| Mutshuler et al. | 2010 | Excluded - music listening | | | | | | | | | | |
| Nakamura et al. | 1999 | Excluded - music listening | | | | | | | | | | |
| Ohnishi et al. | 2001 | Excluded - music listening | | | | | | | | | | |
| Pallesen et al. | 2009 | Excluded - music listening | | | | | | | | | | |
| Pallesen et al. | 2005 | Excluded - music listening | | | | | | | | | | |
| Pallesen et al. | 2010 | Excluded - music listening | | | | | | | | | | |

| | | | | | | | | | | | | | |
|---|---|---|---|---|---|---|---|---|---|---|---|---|---|
| Perry et al. | 1999 | | No | Cross-sectional | 13 adults without substantial musical training (mean age = 24), 6 male/7 female | Canada | Singing | 1 minute | Once | Emotional regulation (anterior cingulate cortex activation) | Positive | Increased ACC activation during singing | Very Low |
| Ruiz, Jabusch & Altenmueller | 2009 | | No | Cross-sectional | 19 healthy professional pianists (mean age = 22), 8 female/11 male | Germany | Piano | 2 hours | Once | Emotional regulation (anterior cingulate cortex activation) | Unclear | Increased ACC activation associated with error monitoring during piano performance, but unclear significance for associations between music performance and emotional regulation | Very Low |
| Satoh et al. | 2001 | Excluded - music listening | | | | | | | | | | | |
| Suda et al. | 2008 | Excluded - music listening | | | | | | | | | | | |
| Thaut et al. | 2009 | Excluded - music listening | | | | | | | | | | | |
| Vogt et al | 2007 | | No | Cross-sectional | 32 healthy adults (16 male, 16 female; 16 non-guitarists - mean age = 26.6; 16 guitarists - mean age = 27.3) | Germany | Guitar | Unclear | Once | Emotional regulation (dorsolateral prefrontal cortex activation) | Positive | Increased activation of lPFC during chord performance in both non-guitarists and guitarists | Very Low |
| Zarate & Zatorre | 2008 | | No | Cross-sectional | 24 healthy adults (mean age = 23; 12 experienced singers, 6 male/6 female; 12 non-musicians, 6 male/6 female) | Canada | Singing | Unclear | Once | Emotional regulation (anterior cingulate cortex activation) | Positive | Increased ACC activation during singing in both singers & non-musicians | Very Low |
| Zarate, Wood & Zatorre | 2010 | | No | Cross-sectional | 9 healthy, experienced singers (mean age = 23), 3 male/6 female | Canada | Singing | Unclear | Once | Emotional regulation (anterior cingulate cortex activation) | Positive | Increased ACC activation during 'compensate tasks' (i.e. singer must compensate for pitch shifts) | Very Low |
| ***Phillips & Becker, 2019*** | | | | | | | | | | | | | |
| Anderson & Gustavson | 2016 | Excluded - study of knitting, not music, intervention | | | | | | | | | | | |
| Bittman et al. | 2003 | | Yes (Yamaha, Remo) | RCT | 125 healthcare workers (Retirement Community; mean age 43.8), 24 male/101 female | USA | Group drumming | 6 weeks | 1 hour, weekly | Burnout (emotional exhaustion) | Positive | Significant pre/post improvement (**1-tailed t-test**) following intervention but no analysis of data | Low |

| | | | | | | | | | | | | |
|---|---|---|---|---|---|---|---|---|---|---|---|---|
| | | | | | | | | | | | vs control (Maslach Burnout Inventory) | |
| | | | | | | | | | Burnout (depersonalization ) | No effect | No significant pre/post differences (Maslach Burnout Inventory) | Low |
| | | | | | | | | | Burnout (personal accomplishment) | Positive | Significant pre/post improvement (**1-tailed t-test**) following intervention but no analysis of data vs control (Maslach Burnout Inventory) | Low |
| | | | | | | | | | Mood | Positive | Significant pre/post decrease in mood disturbance (although no analysis of data vs control)(Profile of Mood States) | Low |
| Bormann et al. | 2006 | No | Single group study | 62 Healthcare workers (mean age = 47.8; 54 female) | USA | 'mantram' singing | 5 weeks | 90 minutes, weekly | Stress | Positive | Significant pre-post reduction in stress (Perceived Stress Scale) | Very Low |
| | | | | | | | | | Anxiety (state) | No effect | No significant pre-post effect on state anxiety (Spielburger State-Trait Anxiety Inventory) | Very Low |
| | | | | | | | | | Anxiety (trait) | Positive | Significant pre-post reduction in trait anxiety(Spielburger State-Trait Anxiety Inventory) | Very Low |
| | | | | | | | | | Anger (state) | No effect | No significant pre-post effect on state anger (Spielburger State-Trait Anger Inventory) | Very Low |
| | | | | | | | | | Anger (trait) | Positive | Significant pre-post reduction in trait anger (Spielburger State-Trait Anger Inventory) | Very Low |
| | | | | | | | | | Quality of life | Positive | Significant pre-post improvement in quality of life (Quality of Life Enjoyment & Satisfaction Short Form) | Very Low |

| | | | | | | | | | | | |
|---|---|---|---|---|---|---|---|---|---|---|---|
| | | | | | | | | | Spiritual Wellbeing | Positive | Significant pre-post improvement in spiritual wellbeing (Total Spiritual Well-Being Scale) | Very Low |
| Hilliard | 2006 | No | Nonrandomized experimental study | 17 hospice workers (age range 28-60; 11 female, 6 male) | USA | Group music-making | 6 weeks | 60 minutes, weekly | Compassion fatigue | No effect | No pre-post effect on compassion fatigue (subscale of Compassion Satisfaction/Fatigue Self-Test for Helpers) | Very Low |
| Ifrach & Miller | 2016 | Excluded - visual art, not music, intervention | | | | | | | | | |
| Italia et al. | 2008 | Excluded - visual art, not music, intervention | | | | | | | | | |
| Karpaviciute & Macijauskiene | 2016 | Excluded - painting, not music, intervention | | | | | | | | | |
| Kravits et al. | 2010 | Excluded - visual art, not music, intervention | | | | | | | | | |
| MacPherson | 2008 | Excluded - storytelling, not music, intervention | | | | | | | | | |
| Potash et al. | 2014 | Excluded - visual art, not music, intervention | | | | | | | | | |
| Rice et al. | 2014 | Excluded - storytelling, not music, intervention | | | | | | | | | |
| Salzano et al. | 2013 | Excluded - visual art, not music, intervention | | | | | | | | | |
| Sands et al. | 2008 | Excluded - 'narrative training', not music, intervention | | | | | | | | | |
| Wlodarcyzyk | 2013 | Excluded - 'songwriting component the main therapeutic intervention'; involves fill in the blank adaptation of 'Fire and Rain' by James Taylor with subsequent lyric analysis; active music participation not primary component | | | | | | | | | |
| ***Raglio et al., 2019*** | | | | | | | | | | | |
| Beck et al. | 2015 | Excluded - guided music imagery, not music participation, intervention | | | | | | | | | |
| Bittman et al. | 2004 | Other review - Yap, Kwan & Ang, 2017 | | | | | | | | | |
| Bae | 2011 | No | RCT (only comparative, not inactive control group) | 20 female graduate music therapy students (mean age = 27.6) | South Korea | Group drumming | 4 weeks | 60 mins, weekly | Job engagement | No effect | No significant pre-post impact of group drumming on job engagement; *effects vs other music group (guided musical imagery) not considered due to unclear relevance to umbrella review aims (Utrecht work and wellbeing survey (UWES)) | Low |
| | | | | | | | | | Anxiety | No effect | No significant pre-post impact of group | Low |

| | | | | |
|---|---|---|---|---|
| | | | drumming on anxiety; *effects vs other music group (guided musical imagery) not considered due to unclear relevance to umbrella review aims (State-trait anxiety inventory (STAI)) | |
| | Mood | No effect | No significant pre-post impact of group drumming on mood disturbance; *effects vs other music group (guided musical imagery) not considered due to unclear relevance to umbrella review aims (Profile of mood states (POMS)) | Low |
| | Perceived self-efficacy | No effect | No significant pre-post impact of group drumming on perceived self-efficacy; *effects vs other music group (guided musical imagery) not considered due to unclear relevance to umbrella review aim (General self-efficacy scale (GSE)) | Low |
| | Positive affect | Positive | Significant pre-post improvement in positive affect; *effects vs other music group (guided musical imagery) not considered due to unclear relevance to umbrella review aims (Positive affect, negative affect schedule (PANAS)) | Low |
| | Negative affect | No effect | No significant pre-post impact of group drumming on negative | Low |

| | | | | | | | | | | | | | |
|---|---|---|---|---|---|---|---|---|---|---|---|---|---|
| | | | | | | | | | | | | affect; *effects vs other music group (guided musical imagery) not considered due to unclear relevance to umbrella review aims (Positive affect, negative affect schedule (PANAS)) | |
| Brooks et al. | 2010 | Excluded - music & imagery, not music participation, intervention | | | | | | | | | | | |
| DuRousseau et al. | 2011 | Excluded - brain music, not music participation, intervention | | | | | | | | | | | |
| Huang & Shih | 2011 | Excluded - music listening | | | | | | | | | | | |
| Lesiuk | 2008 | Excluded - music listening | | | | | | | | | | | |
| Lesiuk | 2010 | Excluded - music listening | | | | | | | | | | | |
| Miskovic et al. | 2008 | Excluded - music listening | | | | | | | | | | | |
| Shih et al. | 2009 | Excluded - music listening | | | | | | | | | | | |
| Shih et al. | 2012 | Excluded - music listening | | | | | | | | | | | |
| Shih et al. | 2016 | Excluded - music listening | | | | | | | | | | | |
| Speer | 2011 | Excluded - music listening | | | | | | | | | | | |
| Wlodarczyk | 2010 | Excluded - 'songwriting component the main therapeutic intervention'; involves fill in the blank adaptation of 'Fire and Rain' by James Taylor with subsequent lyric analysis; active music participation not primary component | | | | | | | | | | | |
| ***Skingley & Vella-Burrows, 2010*** *'Music in everyday life' section only (excluded Table 1 - 'Studies relating to music or singing and people with dementia' and Table 2 - 'Studies relating to music and specific disorders')* | | | | | | | | | | | | | |
| Bungay & Skingley | 2008 | Excluded - qualitative results only | | | | | | | | | | | |
| Cohen et al. | 2006 | Other review - Daykin et al., 2018 | | | | | | | | | | | |
| Hays | 2005 | Excluded - qualitative results only | | | | | | | | | | | |
| Hays & Mihichiello | 2005 | Excluded - qualitative results only | | | | | | | | | | | |
| Pickles | 2003 | Excluded - qualitative results only | | | | | | | | | | | |
| ***Talmini et al, 2017*** | | | | | | | | | | | | | |
| Anaya, Pisoni & Kronenberger | 2016 | | No | Cross-sectional | 48 adults (mean age 22.1; 24 musicians, 24 nonmusicians); sex unspecified | USA | Piano/organ | average 15.5 years of musical training | average 16.3 hours of weekly practice | Short-term memory | Meta-analysis | | Low |
| Bialystok & DePape | 2009 | | No | Cross-sectional | 46 adults (mean age 24.3; 22 musicians, | Canada, UK | Instrumental | Average 16.6 years of | Average 2 hours of | Short-term memory | Meta-analysis | | Low |

| | | | | | | | | | | | |
|---|---|---|---|---|---|---|---|---|---|---|---|
| | | | | 24 nonmusicians); sex unspecified | | | musical training | daily practice | Working memory | Meta-analysis | Low |
| Boebinger et al. | 2015 | No | Cross-sectional | 50 adults (mean age 27.2; 25 musicians, 25 nonmusicians)l sex unspecified | UK | Instrumental | Average 22.7 years of musical training | Practice at least 3x/weekly over the 3 years before testing | Short-term memory | Meta-analysis | Low |
| | | | | | | | | | Working memory | Meta-analysis | Low |
| Brandler & Rammsayer | 2003 | No | Cross-sectional | 70 adults (mean age 28.5, 39 female/31 male; 35 musicians, 35 nonmusicians) | Germany | Instrumental | >14 years of musical training | Unspecified | Long-term memory | Meta-analysis | Low |
| | | | | | | | | | Long-term memory | Meta-analysis | Low |
| Chan, Ho & Cheung | 1998 | No | Cross-sectional | 60 adults (mean age 19.8; 30 musicians, 30 nonmusicians), sex unspecified | Hong Kong | Instrumental | At least 6 years of music training before age 12 | Unspecified | Long-term memory | Meta-analysis | Low |
| | | | | | | | | | Long-term memory | Meta-analysis | Low |
| Clayton et al. | 2016 | No | Cross-sectional | 34 adults (mean age 23.5; 17 musicians, 17 nonmusicians), sex unspecified | USA | Instrumental | average 14.4 years of music training | Unclear | Working memory | Meta-analysis | Low |
| Franklin et al. | 2008 | No | Cross-sectional | 25 adults (mean age 19.7; 12 musicians, 13 nonmusicians), 19 female/6 male | USA | Instrumental | At least 9 years continuous music training, beginning before age 10 | Practice/ performance at least 15 hours/week | Long-term memory | Meta-analysis | Low |
| | | | | 20 adults (mean age 21.6; 11 musicians, 9 nonmusicians), 10 female/10 male | | | | | Working memory | Meta-analysis | Low |
| George & Coch | 2011 | No | Cross-sectional | 32 adults (mean age 20.1; 16 musicians, 16 nonmusicians), 16 female/16 male | USA | Instrumental | Average 12.1 years of music training | Average 6.3 hours of weekly practice | Short-term memory | Meta-analysis | Low |
| | | | | | | | | | Working memory | Meta-analysis | Low |
| | | | | | | | | | Short-term memory | Meta-analysis | Low |
| Hansen, Wallentin & Vuust | 2012 | No | Cross-sectional | 40 adults (mean age 21.1; 20 musicians, 20 nonmusicians), sex breakdown unspecified for included comparison - mixed | Denmark | Instrumental | Average 10.5 years of music training | Average 23.5 hours of weekly practice | Short-term memory | Meta-analysis | Low |
| | | | | | | | | | Working memory | Meta-analysis | Low |
| | | | | | | | | | Short-term memory | Meta-analysis | Low |
| | | | | | | | | | Working memory | Meta-analysis | Low |
| Helmbold, Rammsayer & Altenmueller | 2005 | No | Cross-sectional | 140 adults (mean age 22.5; 70 musicians, 70 | Germany | Instrumental | Average 17 years of music training | Unspecified | Long-term memory | Meta-analysis | Low |

| | | | | | | | | | | | |
|---|---|---|---|---|---|---|---|---|---|---|---|
| | | | | nonmusicians), 74 female/66 male | | | | | Long-term memory | Meta-analysis | Low |
| Huang et al. | 2010 | No | Cross-sectional | 20 adults (mean age 21.5; 10 musicians, 10 nonmusicians), sex unspecified | China | Piano | At least 8 years continuous music training begun before age 9 | Unspecified | Long-term memory | Meta-analysis | Low |
| Jakobson et al. | 2008 | No | Cross-sectional | 36 adults (mean age 19; 15 musicians, 21 nonmusicians), 23 female/13 male | Canada | Piano | Average 11.5 years of music training | Unspecified (11 of 15 in music group 'actively engaged in musical activities') | Long-term memory | Meta-analysis | Low |
| | | | | | | | | | Long-term memory | Meta-analysis | Low |
| Lee, Lu & Ko | 2007 | No | Cross-sectional | 40 adults (mean age 22; 20 musicians, 20 nonmusicians), sex unspecified | Taiwan | Instrumental | Average 14.3 years of music training | Unspecified | Short-term memory | Meta-analysis | Low |
| | | | | | | | | | Working memory | Meta-analysis | Low |
| | | | | | | | | | Working memory | Meta-analysis | Low |
| Monahan, Kendall & Carterette | 1987 | No | Cross-sectional | 22 adults (mean age not provided, introductory psychology students and older); 12 musicians, 10 nonmusicians), sex unspecified | USA | Instrumental | At least 5 years of music training | Unspecified | Short-term memory | Meta-analysis | Low |
| Okhrei, Kutsenko & Makarchuk | 2017 | No | Cross-sectional | 66 adults (mean age 20; 28 musicians, 36 nonmusicians), sex unspecified | Ukraine | Music experience' (may include singing | 10-14 years of music training | Unspecified | Short-term memory | Meta-analysis | Low |
| | | | | | | | | | Short-term memory | Meta-analysis | Low |
| Pallesen et al. | 2010 | No | Cross-sectional | 21 adults (mean age 26.5; 11 musicians, 10 nonmusicians), 14 female/7 male | Finland | Instrumental | Unspecified (graduates of Sibelius Music Academy) | Unspecified | Short-term memory | Meta-analysis | Low |
| | | | | | | | | | Working memory | Meta-analysis | Low |
| Parbery-Clark et al. | 2011 | No | Cross-sectional | 37 adults (mean age 50; 18 musicians, 19 nonmusicians); age range 45-65; sex unspecified | USA | Piano & violin | Average 50 years music training | practice at least 3x/weekly over the 3 years before testing | Working memory | Meta-analysis | Low |
| | 2012 | No | Cross-sectional | 60 adults (mean age 19.5; 30 musicians, | USA | Unspecified | 8+ years music training; | | Short-term memory | Meta-analysis | Low |

| | | | | | | | | | | | |
|---|---|---|---|---|---|---|---|---|---|---|---|
| Ramachandra, Meighan & Gradzki | | | | 30 nonmusicians), sex unspecified | | | 'University music students who began music training before age 10' | Practice at least 4 hours/week | Working memory | Meta-analysis | Low |
| Rodrigues, Loureiro & Caramelli | 2014 | No | Cross-sectional | 76 adults (mean age 32.2; 38 musicians, 38 nonmusicians), 56 male/20 female | Brazil | Instrumental (symphony orchestra musicians) | Average 23 years music training | average 3.2 hours daily practice | Short-term memory | Meta-analysis | Low |
| Schiavio & Timmers | 2016 | No | Cross-sectional | 20 adults (mean age 24.8; 10 musicians, 10 nonmusicians); sex unspecified | UK | Instrumental | Minimum 5 years music training | Unspecified | Long-term memory | Meta-analysis | Very Low |
| Schulze, Dowling & Tillmann | 2012 | No | Cross-sectional | 40 adults (mean age 22.7; 20 musicians, 20 nonmusicians), sex unspecified | France | Instrumental | Average 17.7 years music training | Unspecified | Short-term memory | Meta-analysis | Low |
| | | | | | | | | | Working memory | Meta-analysis | Low |
| Schulze, Mueller & Koelsch | 2011 | No | Cross-sectional | 33 adults (mean age 24.5; 16 musicians, 17 nonmusicians), 18 male/15 female | Germany | Instrumental | Unspecified (began music training at average 6.1 years old) | "Practice for several hours each day" | Short-term memory | Meta-analysis | Low |
| Schulze et al. | 2011 | No | Cross-sectional | 33 adults (mean age 24.5; 16 musicians, 17 nonmusicians), 18 male/15 female | Germany | Instrumental | Unspecified | Unspecified | Working memory | Meta-analysis | Low |
| | | | | | | | | | Working memory | Meta-analysis | Low |
| Suarez, Elangovan & Au | 2015 | No | Cross-sectional | 54 adults (mean age 22.6; 24 musicians, 30 nonmusicians), 11 male/43 female | Singapore | Instrumental | Average 10.4 years music training | Unspecified | Working memory | Meta-analysis | Low |
| | | | | | | | | | Short-term memory | Meta-analysis | Low |
| | | | | | | | | | Short-term memory | Meta-analysis | Low |
| | | | | | | | | | Long-term memory | Meta-analysis | Low |
| Talamini, Carretti, & Grassi | 2016 | No | Cross-sectional | 36 adults (mean age 22.6; 18 musicians, 18 nonmusicians), 22 female/14 male | Italy | Instrumental | Average 12.3 years music training | Average 19.6 hours weekly practice | Short-term memory | Meta-analysis | Low |
| | | | | | | | | | Working memory | Meta-analysis | Low |
| Taylor & Dewhurst | 2017 | No | Cross-sectional | 40 adults (mean age 21.7; 20 musicians, 20 nonmusicians), 22 male/18 female | UK | Instrumental | At least 4 years of music training | Practice at least once pre week | Long-term memory | Meta-analysis | Low |
| Vasuki et al. | 2016 | No | Cross-sectional | 35 adults (mean age 25.8; 17 musicians, | Australia | Instrumental or singing | At least 10 years of music/singing | Unspecified - 'Still actively | Short-term memory | Meta-analysis | Low |
| | | | | | | | | | Working memory | Meta-analysis | Low |

| | | | | | | | | | | | |
|---|---|---|---|---|---|---|---|---|---|---|---|
| | | | | 18 nonmusicians), 10 male/25 female | | | experience beginning before age 9 | practiced music' | | | |
| Weiss et al. | 2014 | No | Cross-sectional | 57 adults (mean age 23.4; 42 musicians, 15 nonmusicians), sex unspecified | Israel | Instrumental or singing | Average 11.9 years music training | Unspecified | Short-term memory | Meta-analysis | Low |
| Zuk et al | 2014 | No | Cross-sectional | 30 adults (mean age 24.8; 15 musicians, 15 nonmusicians), 18 male/12 female | USA | Instrumental | Unclear, likely 15+ years) | Average 21.9 hours weekly practice | Working memory | Meta-analysis | Low |
| ***Vaughn, 2000*** | | | | | | | | | | | |
| Annello | 1972 | No | Cross-sectional | 326 9th-12th graders, 170 male/156 female | USA | Instrumental music education | 1-4 years | Unspecified | Mathematics (correlation) | Meta-analysis | Low |
| Catterall, Chapleau & Iwanaga | 1999 | No | Cross-sectional | 1476 high school seniors, sex unspecified | USA | Instrumental music education | 1-5 years | Unspecified | Mathematics (correlation) | Meta-analysis | Low |
| Ciepluch | 1988 | No | Cross-sectional | 80 9th-12th graders, sex unspecified | USA | Instrumental music education (band) | 1-4 years | Unspecified | Mathematics (correlation) | Meta-analysis | Very Low |
| College Board | 1988-1998 (not 1993) | No | Cross-sectional | 5,732,282 high school seniors, mixed sex | USA | Instrumental or vocal music class | At least 1 course | Not specified | Mathematics (correlation) | Meta-analysis | Moderate |
| Costa-Giomi | 1999 | No | Quasi-randomized longitudinal cohort study | 129 4th graders; age 9 at start of intervention (58 girls, 59 boys) | Canada | Piano lessons | 3 years | 30-45 minutes, weekly (instruction time only) | Mathematics (causation - experimental) | Meta-analysis | Very Low |
| Engdahl | 1994 | No | Retrospective cohort study | 598 elementary school students (enrolled as 3rd graders), sex unspecified | USA | Instrumental music education (orchestra & band) | 3 years | At least 90 minutes, weekly (school weeks) | Mathematics (correlation) | Meta-analysis | Low |
| Friedman | 1959 | No | Longitudinal cohort study | 28 5th & 6th graders, sex unspecified | USA | Instrumental music education | 1-2 years | 7-8 hours, weekly | Mathematics (causation - experimental) | Meta-analysis | Low |
| Graziano, Peterson & Shaw | 1999 | Yes (Yamaha Corp) | Non randomized experimental cohort study | 55 2nd graders, sex unspecified | USA | Piano lessons | 14 weeks | 60 minutes, 3x/week | Mathematics (causation - experimental) | Meta-analysis | Low |

| | | | | | | | | | | | | |
|---|---|---|---|---|---|---|---|---|---|---|---|---|
| Kvet | 1982 | No | Cross-sectional | 350 3rd-6th graders, sex breakdown unspecified - mixed | USA | Instrumental music education | 8 months | 70 min, weekly | Mathematics (correlation) | Meta-analysis | | Low |
| McCarthy | 1992 | | Unclear (correlational) | 1061 college students, age & sex unavailable | USA | Orchestra | At least 2 years college music instruction | Unspecified | Mathematics (correlation) | Meta-analysis | | Low |
| Neufeld | 1986 | No | Non-randomized experimental cohort study | 80 children (kindergarten & pre-k), sex unavailable | USA | Kodaly Music education | 1-2 years | Unspecified | Mathematics (causation - experimental) | Meta-analysis | | Low |
| Weeden | 1971 | No | Non-randomized experimental cohort study | 47 1st & 2nd graders, sex unspecified | USA | Suzuki violin education | 4 months | 2x/week | Mathematics (causation - experimental) | Meta-analysis | | Low |
| Wheeler & Wheeler | 1951 | No | Cross-sectional | 1969 college students, age & sex unspecified | USA | College music majors | At least 2 years college music instruction | Unspecified | Mathematics (correlation) | Meta-analysis | | Low |
| ***Yap, Kwan & Ang, 2017*** | | | | | | | | | | | | |
| Bittman | 2004 | Yes (Remo & Yamaha) | Nonrandomized crossover experimental study | 75 nursing students (mean age 27.5; 64 women, 11 men) | USA | Group drumming | 1 hour | 6 weeks | Mood | Positive | Significant pre-post improvement in overall mood state (POMS), changes persisted for 6 weeks post-intervention | Very Low |
| | | | | | | | | | Emotional exhaustion | Positive | Significant pre-post reduction in emotional exhaustion (Malasch Burnout Inventory), changes persisted 6 weeks post-intervention | Very Low |
| | | | | | | | | | Depersonalization | NO change | No significant pre-post changes in feelings of depersonalization | Very Low |
| | | | | | | | | | Personal accomplishment | No change | No significant pre-post changes in feelings of personal accomplishment | Very Low |
| Dunbar et al | 2012 | No | Nonrandomized experimental study | 32 adults (12 from samba drumming school (mean age 44.2; mean 4.25 years samba drumming | UK | Drumming (samba) | 30 minutes | Once | Positive affect | Positive | Positive effect of samba drumming on positive affect vs control groups(Positive and Negative Affect Schedule (PANAS)) | Low |

| | | | | | | | | | | | | |
|---|---|---|---|---|---|---|---|---|---|---|---|---|
| | | | | experience); 9 employees from city music shop (control 1; mean age 31.1); 11 Master's students (control 2 (watched video of speech), mean age 24.6), 17 male/15 female | | | | | Negative affect | No effect | No effect of samba drumming vs control groups (Positive and Negative Affect Schedule (PANAS)) | Low |
| Ho et al | 2011 | No | Nonrandomized intervention study | 101 5th graders (mean age ~10.5; 54 girls, 47 boys) | USA | Group drumming | 40-45 min | 12 weeks | Social functioning | Positive | Significant improvement in social function in group drumming groups vs control (Teacher's Report Form) | Low |
| Koyama et al. | 2009 | Other review - Fancourt, Ockelford & Belai, 2014 | | | | | | | | | | |
| Mungas & Silverman | 2014 | No | Nonrandomized experimental study | 50 university students (17 experimental group (mean age = 20.8); 33 control group (mean age = 21.5)), 35 female/15 male | USA | Group drumming | 45 min - 1 hour | Once | Drowsiness | Positive | Significantly reduced drowsiness pre-post (Quick Mood Scale) | Very Low |
| | | | | | | | | | Anxiety | Positive | Significantly reduced anxiety pre-post (Quick Mood Scale) | Very Low |
| | | | | | | | | | Depression | Positive | Significantly reduced depression pre-post (Quick Mood Scale) | Very Low |
| | | | | | | | | | Aggression | Positive | Significantly reduced aggression pre-post (Quick Mood Scale) | Very Low |
| | | | | | | | | | Confusion | Positive | Significantly reduced confusion pre-post (Quick Mood Scale) | Very Low |
| | | | | | | | | | Lack of coordination | No effect | NO significant pre-post effect on lack of coordination (Quick Mood Scale) | Very Low |
| Shimzu et al. | 2013 | Excluded - 'music movement therapy' (exercise program incorporating the 'Naruko clapper'; designed as exercise, not music-making) | | | | | | | | | | |
| Smith, Viljoen & McGeachie | 2014 | No | Single group experimental study (pre-post analysis) | 17 young adult novice drummers (mean age 23.4; 13 women, 4 men) | South Africa | Drumming (djembe) | 40 minutes | Once | Blood pressure | No effect | No significant pre-post changes in systolic or diastolic blood pressure | Very Low |
| | | | | 17 middle aged experienced drummers (mean | | | | | Blood pressure | Positive | Significant pre-post decrease in systolic blood pressure; no pre- | Very Low |

| | | | | | | | | | | | | | |
|---|---|---|---|---|---|---|---|---|---|---|---|---|---|
| | | | | | age 53.1; 12 women, 5 men) | | | | | | | post change in diastolic blood pressure | |
| | | | | | 17 young adult novice drummers (mean age 23.4; 13 women, 4 men) | | | | | Stress/anxiety | Positive | Significant pre-post decrease in stress/anxiety (stress anxiety index (SAI)) | Very Low |
| | | | | | 17 middle aged experienced drummers (mean age 53.1; 12 women, 5 men) | | | | | Stress/anxiety | Positive | Significant pre-post decrease in stress/anxiety (stress anxiety index (SAI)) | Very Low |
| Snow & D'Amico | 2010 | Excluded - qualitative results only | | | | | | | | | | | |
| Wachi et al. | 2007 | | Yes (Yamaha Corp) | RCT | 40 healthy males (mean age 38.4) | Japan | Group drumming | 60 minutes | Once | Mood disturbance | Positive | (Profile of Mood States (POMS)) | High |
| | | | | | | | | | | Natural killer cell activity (effector:target 25:1) | Negative | Significant decrease in NK cell activity vs control | High |
| | | | | | | | | | | Natural killer cell activity (effector:target 12:1) | Negative | Significant decrease in NK cell activity pre-post & vs control | High |
| | | | | | | | | | | Natural killer cell activity | Positive | Modulation of NK cell activity towards 'mid-range' pre-post drumming activity (i.e. individuals with low NK cell levels up regulated post-drumming, individuals with high NK cell activity down regulated post-drumming) | High |
| | | | | | | | | | | CD-56+ cells | No effect | No effect of group drumming pre-post or vs control | High |
| | | | | | | | | | | IFN-gamma gene expression (mRNA) | Positive | Significant decrease in IFN-alpha gene expression vs control (anti-inflammatory response) | High |

| | | | | |
|---|---|---|---|---|
| | IL-2 gene expression (mRNA) | NO effect | No effect of group drumming on IL-2 gene expression (pre-post or vs control) | High |
| | IL-6 gene expression (mRNA) | No effect | No effect of group drumming on IL-6 gene expression (pre-post or vs control) | High |
| | IL-10 gene expression (mRNA) | Positive | Significant increase in IL-10 gene expression pre-post group drumming and vs control | High |
| | Leukocyte counts | No effect | No effect of group drumming on leukocyte counts vs control (increased pre-post in both groups) | High |

*Table S9. GRADE Certainty of Evidence ratings for each outcome measure from individual randomized controlled trials (RCTs) of dance participation contained in included reviews.*

| *Author* | *Year* | *Outcome* | ***GRADE Initial quality*** | *Concealed allocation* | *Blinded assessors/ data analysts* | *Low % lost to follow up* | *Intention to treat analysis* | *All outcome result(s) reported* | *Validated outcome measures used* | *Used individual randomization (not cluster randomization)* | *Carryover effects controlled (cross-over trial)* | ***GRADE Bias risk*** | ***GRADE Indirectness*** | ***GRADE up rating*** | ***GRADE Certainty of Evidence*** |
|---|---|---|---|---|---|---|---|---|---|---|---|---|---|---|---|
| ***Burkhardt & Brennan, 2012*** | | | | | | | | | | | | | | | |
| Burgess, Grogan & Burwitz | 2006 | Self-perception | High | Yes | Unspecified | Yes | Yes | Yes | Yes | Yes | No | Serious (immediate transition between dance & exercise groups in previously inactive girls) | None | None | Moderate |
| Flores | 1995 | Timed mile run | High | Yes | Unspecified | Yes | Yes | Yes | Yes | No (classes randomized, not individuals) | N/A | Serious (cluster randomization by classroom) | None | None | Moderate |
| | | BMI | High | Yes | Unspecified | Yes | Yes | Yes | Yes | No (classes randomized, not individuals) | N/A | Serious (cluster randomization by classroom) | None | None | Moderate |
| Kim & Kim | 2007 | Mood | High | Questionable (all female hip-hop dance group, but sex mix in other groups) | N/A (questionnaire) | Yes | Yes | Yes | Yes | Yes | Yes | Serious (questionable blinding of allocation) | None | None | Moderate |
| Robinson et al | 2003 | BMI | High | No (family members allowed to group together) | Yes | Yes | Yes | No (no reporting of analyses of changes in blood lipid/glucose/ insulin markers) | Yes | Yes | N/A | Very serious (family members permitted to circumvent randomization to be in same group as other family members; analysis admittedly underpowered to detect between group differences) | None | None | Low |
| | | Waist circumference | High | No (family members allowed to | Yes | Yes | Yes | No (no reporting of analyses of changes in blood | Yes | Yes | N/A | Very serious (family members permitted to circumvent | None | None | Low |

| | | | | | | | | | | | | | | | |
|---|---|---|---|---|---|---|---|---|---|---|---|---|---|---|---|
| | | | | group together) | | | | lipid/glucose/ insulin markers) | | | | randomization to be in same group as other family members; analysis admittedly underpowered to detect between group differences) | | | |
| | | Self-esteem | High | No (family members allowed to group together) | Yes | Yes | Yes | No (no reporting of analyses of changes in blood lipid/glucose/ insulin markers) | Yes | Yes | N/A | Very serious (family members permitted to circumvent randomization to be in same group as other family members; analysis admittedly underpowered to detect between group differences) | None | None | Low |
| ***Clarke et al., 2018*** | | | | | | | | | | | | | | | |
| ***Costa, Ferreira & Felicio, 2013*** | | | | | | | | | | | | | | | |
| ***Cugusi et al, 2019*** | | | | | | | | | | | | | | | |
| Delextrat et al. | 2016 | VO2 max | High | Yes | Unspecified | Yes | Yes | Yes | Yes | Yes | N/A | Minimal | None | None | High |
| | | % fat mass | High | Yes | Unspecified | Yes | Yes | Yes | Yes | Yes | N/A | Minimal | None | None | High |
| | | Body mass | High | Yes | Unspecified | Yes | Yes | Yes | Yes | Yes | N/A | Minimal | None | None | High |
| Domene et al | 2016 | Blood pressure | High | Yes | Unspecified | Yes | Yes | Yes | No (valid outcomes, but only pre-post analysis of results; no statistical comparison vs control) | Yes | N/A | Very serious (pre-post analyses only; no statistical comparison between dance & control groups – effectively a | None | None | Low |

| | | | | | | | | | | | | | |
|---|---|---|---|---|---|---|---|---|---|---|---|---|---|
| | | | | | | | | | | single group study) | | | |
| Body mass | High | Yes | Unspecified | Yes | Yes | Yes | No (valid outcomes, but only pre-post analysis of results; no statistical comparison vs control) | Yes | N/A | Very serious (pre-post analyses only; no statistical comparison between dance & control groups – effectively a single group study) | None | None | Low |
| % fat mass | High | Yes | Unspecified | Yes | Yes | Yes | No (valid outcomes, but only pre-post analysis of results; no statistical comparison vs control) | Yes | N/A | Very serious (pre-post analyses only; no statistical comparison between dance & control groups – effectively a single group study) | None | None | Low |
| White blood cell count | High | Yes | Unspecified | Yes | Yes | Yes | No (valid outcomes, but only pre-post analysis of results; no statistical comparison vs control) | Yes | N/A | Very serious (pre-post analyses only; no statistical comparison between dance & control groups – effectively a single group study) | None | None | Low |
| CRP | High | Yes | Unspecified | Yes | Yes | Yes | No (valid outcomes, but only pre-post analysis of results; no statistical comparison vs control) | Yes | N/A | Very serious (pre-post analyses only; no statistical comparison between dance & control groups – effectively a single group study) | None | None | Low |
| IL-6 | High | Yes | Unspecified | Yes | Yes | Yes | No (valid outcomes, | Yes | N/A | Very serious (pre-post | None | None | Low |

| | | | | | | | | | | | | | | | |
|---|---|---|---|---|---|---|---|---|---|---|---|---|---|---|---|
| | | | | | | | | but only pre-post analysis of results; no statistical comparison vs control) | | | analyses only; no statistical comparison between dance & control groups – effectively a single group study) | | | |
| | | Total cholesterol to HDL ratio | High | Yes | Unspecified | Yes | Yes | Yes | No (valid outcomes, but only pre-post analysis of results; no statistical comparison vs control) | Yes | N/A | Very serious (pre-post analyses only; no statistical comparison between dance & control groups – effectively a single group study) | None | None | Low |
| | | VO2 max | High | Yes | Unspecified | Yes | Yes | Yes | No (valid outcomes, but only pre-post analysis of results; no statistical comparison vs control) | Yes | N/A | Very serious (pre-post analyses only; no statistical comparison between dance & control groups – effectively a single group study) | None | None | Low |
| | | BMI | High | Yes | Unspecified | Yes | Yes | Yes | No (valid outcomes, but only pre-post analysis of results; no statistical comparison vs control) | Yes | N/A | Very serious (pre-post analyses only; no statistical comparison between dance & control groups – effectively a single group study) | None | None | Low |
| Donath et al | 2014 | 6-minute walk test | High | Yes | Unspecified | Yes | Yes | Yes | Yes | Yes | N/A | Minimal | None | None | High |
| | | BMI | High | Yes | Unspecified | Yes | Yes | Yes | Yes | Yes | N/A | Minimal | None | None | High |
| ***Fernandez-Arguelles et al., 2015*** | | | | | | | | | | | | | | | |

| | | | | | | | | | | | | | | |
|---|---|---|---|---|---|---|---|---|---|---|---|---|---|---|
| ***Fong Yan et al., 2018*** | | | | | | | | | | | | | | |
| Barene et al. | 2016 | Sit & Reach | High | Yes | Unspecified | Yes | Yes | Yes | Yes | No (cluster randomization) | N/A | Serious (cluster randomization + unclear blinding of assessors) | None | None | Moderate |
| | | Postural stability | High | Yes | Unspecified | Yes | Yes | Yes | Yes | No (cluster randomization) | N/A | Serious (cluster randomization + unclear blinding of assessors) | None | None | Moderate |
| Barene et al. | 2014 | Fat mass | High | Yes | Unspecified | Yes | Yes | Yes | Yes | No (cluster randomization) | N/A | Serious (cluster randomization + unclear blinding of assessors) | None | None | Moderate |
| | | VO2 max | High | Yes | Unspecified | Yes | Yes | Yes | Yes | No (cluster randomization) | N/A | Serious (cluster randomization + unclear blinding of assessors) | None | None | Moderate |
| | | Triglycerides | High | Yes | Unspecified | Yes | Yes | Yes | Yes | No (cluster randomization) | N/A | Serious (cluster randomization + unclear blinding of assessors) | None | None | Moderate |
| | | Glucose | High | Yes | Unspecified | Yes | Yes | Yes | Yes | No (cluster randomization) | N/A | Serious (cluster randomization + unclear blinding of assessors) | None | None | Moderate |
| | | Peak ventilation | High | Yes | Unspecified | Yes | Yes | Yes | Yes | No (cluster randomization) | N/A | Serious (cluster randomization + unclear blinding of assessors) | None | None | Moderate |
| | | Blood pressure | High | Yes | Unspecified | Yes | Yes | Yes | Yes | No (cluster randomization) | N/A | Serious (cluster randomization + unclear blinding of assessors) | None | None | Moderate |
| | | Respiratory exchange ratio | High | Yes | Unspecified | Yes | Yes | Yes | Yes | No (cluster randomization) | N/A | Serious (cluster randomization + unclear blinding of assessors) | None | None | Moderate |
| | | Total cholesterol | High | Yes | Unspecified | Yes | Yes | Yes | Yes | No (cluster randomization) | N/A | Serious (cluster randomization + unclear blinding of assessors) | None | None | Moderate |
| | | Bone mineral density | High | Yes | Unspecified | Yes | Yes | Yes | Yes | No (cluster randomization) | N/A | Serious (cluster randomization + | None | None | Moderate |

| | | | | | | | | | | | | | | | |
|---|---|---|---|---|---|---|---|---|---|---|---|---|---|---|---|
| | | | | | | | | | | | | unclear blinding of assessors) | | | |
| | | HDL cholesterol | High | Yes | Unspecified | Yes | Yes | Yes | Yes | No (cluster randomization) | N/A | Serious (cluster randomization + unclear blinding of assessors) | None | None | Moderate |
| | | Body mass | High | Yes | Unspecified | Yes | Yes | Yes | Yes | No (cluster randomization) | N/A | Serious (cluster randomization + unclear blinding of assessors) | None | None | Moderate |
| | | BMI | High | Yes | Unspecified | Yes | Yes | Yes | Yes | No (cluster randomization) | N/A | Serious (cluster randomization + unclear blinding of assessors) | None | None | Moderate |
| Belardinelli et al. | 2008 | VO2 max | High | Yes | Unspecified | Yes | Yes | Yes | Yes | Yes | N/A | Minimal | None | None | High |
| | | Triglycerides | High | Yes | Unspecified | Yes | Yes | Yes | Yes | Yes | N/A | Minimal | None | None | High |
| | | Total cholesterol | High | Yes | Unspecified | Yes | Yes | Yes | Yes | Yes | N/A | Minimal | None | None | High |
| | | HDL cholesterol | High | Yes | Unspecified | Yes | Yes | Yes | Yes | Yes | N/A | Minimal | None | None | High |
| Burgess, Grogan & Burwitz | 2006 | BMI | High | Yes | Unspecified | Yes | Yes | Yes | Yes | Yes | No | Serious (immediate transition between dance & exercise groups in previously inactive girls) | None | None | Moderate |
| Garber, McKinney & Carleton | 1992 | VO2 max | High | Yes | Unspecified | Yes | No (analyses only performed on those with 85%+ adherence) | Yes | Yes | Yes | N/A | Serious (analysis only on participants with >85% adherence) | None | None | Moderate |
| | | Peak ventilation | High | Yes | Unspecified | Yes | No (analyses only performed on those with 85%+ adherence) | Yes | Yes | Yes | N/A | Serious (analysis only on participants with >85% adherence) | None | None | Moderate |
| | | Respiratory exchange ratio | High | Yes | Unspecified | Yes | No (analyses only performed on those with | Yes | Yes | Yes | N/A | Serious (analysis only on participants with >85% adherence) | None | None | Moderate |

| | | | | | | | | | | | | | | | |
|---|---|---|---|---|---|---|---|---|---|---|---|---|---|---|---|
| | | | | | | | 85%+ adherence) | | | | | | | | |
| | | Body mass | High | Yes | Unspecified | Yes | No (analyses only performed on those with 85%+ adherence) | Yes | Yes | Yes | N/A | Serious (analysis only on participants with >85% adherence) | None | None | Moderate |
| | | BMI | High | Yes | Unspecified | Yes | No (analyses only performed on those with 85%+ adherence) | Yes | Yes | Yes | N/A | Serious (analysis only on participants with >85% adherence) | None | None | Moderate |
| Hackney et al. | 2007 | Timed up and go | High | Yes | Yes | Yes | Yes | Yes | Yes | Yes | N/A | Minimal | None | None | High |
| | | Berg balance test | High | Yes | Yes | Yes | Yes | Yes | Yes | Yes | N/A | Minimal | None | None | High |
| Hashimoto et al. | 2015 | Timed up and go | High | Yes | Yes | Yes | Yes | Yes | Yes | Yes | N/A | Minimal | None | None | High |
| | | Berg balance test | High | Yes | Yes | Yes | Yes | Yes | Yes | Yes | N/A | Minimal | None | None | High |
| Janyacharoen et al | 2013 | Sit & Reach | High | Yes | Yes | Yes (4/42) | Yes | Yes | Yes | Yes | N/A | Minimal | None | None | High |
| Kalsatou et al. | 2014 | VO2 max | High | Yes | Yes | Yes | Yes | Yes | Yes | Yes | N/A | Minimal | None | None | High |
| | | Peak ventilation | High | Yes | Yes | Yes | Yes | Yes | Yes | Yes | N/A | Minimal | None | None | High |
| | | Berg Balance test | High | Yes | Yes | Yes | Yes | Yes | Yes | Yes | N/A | Minimal | None | None | High |
| | | Body mass | High | Yes | Yes | Yes | Yes | Yes | Yes | Yes | N/A | Minimal | None | None | High |
| | | BMI | High | Yes | Yes | Yes | Yes | Yes | Yes | Yes | N/A | Minimal | None | None | High |
| Kin Isler, Kosar & Korkusuz | 2001 | Triglycerides | High | Yes | Unspecified | Yes | Yes | Yes | Yes | Yes | N/A | Minimal | None | None | High |
| | | Body weight | High | Yes | Unspecified | Yes | Yes | Yes | Yes | Yes | N/A | Minimal | None | None | High |
| | | Total cholesterol | High | Yes | Unspecified | Yes | Yes | Yes | Yes | Yes | N/A | Minimal | None | None | High |
| | | HDL cholesterol | High | Yes | Unspecified | Yes | Yes | Yes | Yes | Yes | N/A | Minimal | None | None | High |
| Rani & Singh | 2013 | Sit & reach | High | Yes | Unspecified | Unspecified | Unspecified | Yes | Yes | Yes | N/A | Serious (unclear reporting re. % lost to follow up / intent to treat analysis) | None | None | Moderate |

| | | | | | | | | | | | | | | | |
|---|---|---|---|---|---|---|---|---|---|---|---|---|---|---|---|
| | | Sit ups | High | Yes | Unspecified | Unspecified | Unspecified | Yes | Yes | Yes | N/A | Serious (unclear reporting re. % lost to follow up / intent to treat analysis) | None | None | Moderate |
| | | 1-mile walk/run | High | Yes | Unspecified | Unspecified | Unspecified | Yes | Yes | Yes | N/A | Serious (unclear reporting re. % lost to follow up / intent to treat analysis) | None | None | Moderate |
| | | Skinfold thickness | High | Yes | Unspecified | Unspecified | Unspecified | Yes | Yes | Yes | N/A | Serious (unclear reporting re. % lost to follow up / intent to treat analysis) | None | None | Moderate |
| Rios Romenets et al | 2015 | Timed up and go | High | Yes | Unspecified | Yes | Yes | Yes | Yes | Yes | N/A | Minimal | None | None | High |
| Sofianidis, Dimitriou & Hatzitaki | 2017 | Postural stability | High | Yes | Unspecified | Yes | Yes | Yes | Yes | Yes | N/A | Minimal | None | None | High |
| Volpe et al. | 2013 | Timed up and go | High | Yes | Yes | Yes | Yes | Yes | Yes | Yes | N/A | Minimal | None | None | High |
| | | Berg balance test | High | Yes | Yes | Yes | Yes | Yes | Yes | Yes | N/A | Minimal | None | None | High |
| ***Hwang & Braun, 2015*** | | | | | | | | | | | | | | | |
| Marmeleira et al | 2009 | Proprioception | High | Yes | Unspecified | Yes | Yes | Yes | Yes | Yes | N/A | Minimal | None | None | High |
| ***Keogh et al., 2009*** | | | | | | | | | | | | | | | |
| ***Letton, Thom, & Ward, 2020*** | | | | | | | | | | | | | | | |
| ***Liu, Shen & Tsai, 2020*** | | | | | | | | | | | | | | | |
| Bennett & Hackney | 2017 | Balance | High | Yes | No | Yes | Yes | Yes | Yes | No (block randomization) | N/A | Serious (potential selection bias due to block randomization) | None | None | Moderate |
| | | Gait speed | High | Yes | No | Yes | Yes | Yes | Yes | No (block randomization) | N/A | Serious (potential selection bias due | None | None | Moderate |

| | | | | | | | | | | | | | | | |
|---|---|---|---|---|---|---|---|---|---|---|---|---|---|---|---|
| | | | | | | | | | | | | to block randomization) | | | |
| Cepeda et al | 2015 | Endurance | High | Yes | Unspecified | Yes | Yes | Yes | Yes | Yes | N/A | Minimal | None | None | High |
| | | Mobility | High | Yes | Unspecified | Yes | Yes | Yes | Yes | Yes | N/A | Minimal | None | None | High |
| Eyigor et al. | 2009 | Balance | High | Yes | Yes | Yes (3/40 lost to follow up) | Yes | Yes | Yes | Yes | N/A | Minimal | None | None | High |
| | | Endurance | High | Yes | Yes | Yes (3/40 lost to follow up) | Yes | Yes | Yes | Yes | N/A | Minimal | None | None | High |
| | | Mobility | High | Yes | Yes | Yes (3/40 lost to follow up) | Yes | Yes | Yes | Yes | N/A | Minimal | None | None | High |
| | | General health | High | Yes | Yes | Yes (3/40 lost to follow up) | Yes | Yes | Yes | Yes | N/A | Minimal | None | None | High |
| Granacher et al. | 2012 | Balance | High | Yes | Unspecified | Yes | Yes | Yes | Yes | Yes | N/A | Minimal | None | None | High |
| | | Gait speed | High | Yes | Unspecified | Yes | Yes | Yes | Yes | Yes | N/A | Minimal | None | None | High |
| Hamacher et al. | 2015 | Gait speed | High | Yes | Yes | Borderline (13/48) lost to follow up) | Yes | Yes | Yes | Yes | N/A | Minimal | None | None | High |
| Holmerova et al. | 2009 | Flexibility | High | Yes | Yes | Borderline (20/72 lost to follow up) | Yes | Yes | Yes | Yes | N/A | Minimal | None | None | High |
| | | Mobility | High | Yes | Yes | Borderline (20/72 lost to follow up) | Yes | Yes | Yes | Yes | N/A | Minimal | None | None | High |
| Hui, Chui & Woo | 2009 | Endurance | High | Yes | Yes | Yes (9/111) | Yes | Yes | Yes | No (random allocation of 2 groups) | N/A | Serious (group, rather than individual, randomization) | None | None | Moderate |
| | | Flexibility | High | Yes | Yes | Yes (9/111) | Yes | Yes | Yes | No (random allocation of 2 groups) | N/A | Serious (group, rather than individual, randomization) | None | None | Moderate |
| | | Mobility | High | Yes | Yes | Yes (9/111) | Yes | Yes | Yes | No (random allocation of 2 groups) | N/A | Serious (group, rather than individual, randomization) | None | None | Moderate |

| | | | | | | | | | | | | | | |
|---|---|---|---|---|---|---|---|---|---|---|---|---|---|---|
| | | General health | High | Yes | Yes | Yes (9/111) | Yes | Yes | Yes | No (random allocation of 2 groups) | N/A | Serious (group, rather than individual, randomization) | None | None | Moderate |
| Janyacharoen et al. | 2013 | Endurance | High | Yes | Yes | Yes (4/42) | Yes | Yes | Yes | Yes | N/A | Minimal | None | None | High |
| | | Flexibility | High | Yes | Yes | Yes (4/42) | Yes | Yes | Yes | Yes | N/A | Minimal | None | None | High |
| | | Mobility | High | Yes | Yes | Yes (4/42) | Yes | Yes | Yes | Yes | N/A | Minimal | None | None | High |
| Machacova et al. | 2015 | Flexibility | High | Yes | Yes | Borderline (20/72 lost to follow up) | Yes | Yes | Yes | Yes | N/A | Minimal | None | None | High |
| | | Mobility | High | Yes | Yes | Borderline (20/72 lost to follow up) | Yes | Yes | Yes | Yes | N/A | Minimal | None | None | High |
| McKinley et al. | 2008 | Mobility | High | Yes | Yes | Yes | Yes | Yes | Yes | Yes | N/A | Minimal | None | None | High |
| | | Gait speed | High | Yes | Yes | Yes | Yes | Yes | Yes | Yes | N/A | Minimal | None | None | High |
| Merom et al. | 2016 | Gait speed | High | Yes | No | Yes (~80%) | Yes | Yes | Yes | No (cluster randomized by retirement center) | N/A | Serious (cluster randomization + unblinded assessors) | None | None | Moderate |
| | | Mobility | High | Yes | No | Yes (~80%) | Yes | Yes | Yes | No (cluster randomized by retirement center) | N/A | Serious (cluster randomization + unblinded assessors) | None | None | Moderate |
| Sofianidis et al. | 2009 | Balance | High | Yes | Unspecified | Yes | Yes | Yes | Yes | Yes | N/A | Minimal | None | None | High |
| Serrano-Guzman et al. | 2016 | Mobility | High | Yes | Yes | Yes | Yes | Yes | Yes | No (block randomized in groups of 6) | N/A | Serious (block randomization) | None | None | Moderate |
| ***Meng et al., 2020*** | | | | | | | | | | | | | | | |
| Alves | 2013 | Executive function | High | Yes | Unspecified | Yes | Yes | Yes | Yes | Yes | N/A | Minimal | None | None | High |
| Chen | 2014 | Global cognition | High | *Article in Mandarin – rating assigned based on study design* | | | | | | | | | | | High |
| Doi et al | 2017 | Global cognition | High | Yes | Yes | Yes | Yes | Yes | Yes | Yes | N/A | Minimal | None | None | High |
| | | Executive function | High | Yes | Yes | Yes | Yes | Yes | Yes | Yes | N/A | Minimal | None | None | High |
| Kattenstroth et al | 2013 | Global cognition | High | Yes | Unspecified | Yes | Yes | Yes | Yes, but (validated | Yes | N/A | Serious (pre-post analyses of | None | None | Moderate |

| | | | | | | | | | | | | | | | |
|---|---|---|---|---|---|---|---|---|---|---|---|---|---|---|---|
| | | | | | | | | | outcome measures, but only pre-post analysis within groups; no statistical comparisons between control & intervention groups) | | | control & experimental groups only) | | | |
| Kosmat & Vranic | 2017 | Executive function | High | Yes | Unclear | Yes | Yes | Yes | Yes | Yes | N/A | Minimal | None | None | High |
| | | Memory | High | Yes | Unclear | Yes | Yes | Yes | Yes | Yes | N/A | Minimal | None | None | High |
| Lazarou et al | 2017 | Global cognition | High | Yes | Yes | Yes | Yes | Yes | Yes | Yes | N/A | Minimal | None | None | High |
| | | Executive function | High | Yes | Yes | Yes | Yes | Yes | Yes | Yes | N/A | Minimal | None | None | High |
| Merom et al | 2016 | Executive function | High | Yes | No | No (65-70% complete data) | Yes | Yes | Yes | Yes | N/A | Serious (high % lost to follow up; participants lost to follow up had lower baseline scores than other participants) | None | None | Moderate |
| | | Memory | High | Yes | No | No (65-70% complete data) | Yes | Yes | Yes | Yes | N/A | Serious (high % lost to follow up; participants lost to follow up had lower baseline scores than other participants) | None | None | Moderate |
| Merom et al | 2016 | Executive function | High | Yes | No | Yes (~80%) | Yes | Yes | Yes | No (cluster randomized by retirement center) | N/A | Serious (cluster randomization + unblinded assessors) | None | None | Moderate |
| Zhang, Ni & Liu | 2012 | Global cognition | High | *Article in Mandarin – rating assigned based on study design* | | | | | | | | | | | High |

***Rodrigues-Krause et al., 2016***

***Rodrigues-Krause, Krause, Reischak-Oliveira, 2019***

| | | | | | | | | | | | | | | | |
|---|---|---|---|---|---|---|---|---|---|---|---|---|---|---|---|
| Borges et a | 2012 | Functional autonomy | High | Yes | Unspecified | Yes | Yes | Yes | Yes | Yes | N/A | Minimal | None | None | High |
| | | Balance | High | Yes | Unspecified | Yes | Yes | Yes | Yes | Yes | N/A | Minimal | None | None | High |
| Cruz-Ferreira et al | 2015 | Physical Fitness | High | Yes | Unspecified | Yes | Yes | Yes | Yes | Yes | N/A | Minimal | None | None | High |
| | | Life Satisfaction | High | Yes | Unspecified | Yes | Yes | Yes | Yes | Yes | N/A | Minimal | None | None | High |
| Eyigor et al. | 2009 | Depression | High | Yes | Yes | Yes (3/40 lost to follow up) | Yes | Yes | Yes | Yes | N/A | Minimal | None | None | High |
| Federici et al | 2005 | Balance | High | Yes | Unspecified | Yes | Yes | Yes | Yes | Yes | N/A | Minimal | None | None | High |
| | | Smoking | High | Yes | Unspecified | Yes | Yes | Yes | No | Yes | N/A | Serious (unvalidated assessment) | None | None | Moderate |
| | | Alcohol Consumption | High | Yes | Unspecified | Yes | Yes | Yes | No | Yes | N/A | Serious (unvalidated assessment) | None | None | Moderate |
| | | Sexual activity | High | Yes | Unspecified | Yes | Yes | Yes | No | Yes | N/A | Serious (unvalidated assessment) | None | None | Moderate |
| | | Sleep quality | High | Yes | Unspecified | Yes | Yes | Yes | No | Yes | N/A | Serious (unvalidated assessment) | None | None | Moderate |
| Granacher et al | 2012 | Muscular Power (lower leg) | High | Yes | Unspecified | Yes | Yes | Yes | Yes | Yes | N/A | Minimal | None | None | High |
| Hui et al | 2009 | Strength | High | Yes | Yes | Yes (9/111 lost) | Yes | Yes | Yes | No (random allocation of 2 groups) | N/A | Serious (group, rather than individual, randomization) | None | None | Moderate |
| | | Balance | High | Yes | Yes | Yes (9/111 lost) | Yes | Yes | Yes | No (random allocation of 2 groups) | N/A | Serious (group, rather than individual, randomization) | None | None | Moderate |
| | | Body composition | High | Yes | Yes | Yes (9/111 lost) | Yes | Yes | Yes | No (random allocation of 2 groups) | N/A | Serious (group, rather than individual, randomization) | None | None | Moderate |
| Lesser et al | 2016 | Body composition | High | Yes | Yes (blinded assessors, data analysts unclear) | Yes | Yes | Yes | Yes | Yes | N/A | Minimal | None | None | High |
| | | Glucose | High | Yes | Yes (blinded assessors, data | Yes | Yes | Yes | Yes | Yes | N/A | Minimal | None | None | High |

| | | | | | | | | | | | | | | |
|---|---|---|---|---|---|---|---|---|---|---|---|---|---|---|
| | | | | | analysts unclear) | | | | | | | | | |
| | | Insulin | High | Yes | Yes (blinded assessors, data analysts unclear) | Yes | Yes | Yes | Yes | Yes | N/A | Minimal | None | None | High |
| | | Aerobic fitness | High | Yes | Yes (blinded assessors, data analysts unclear) | Yes | Yes | Yes | Yes | Yes | N/A | Minimal | None | None | High |
| McKinley et al | 2008 | Balance confidence | High | Yes | Yes | Yes | Yes | Yes | Yes | Yes | N/A | Minimal | None | None | High |
| Wu et al | 2016 | Body composition | High | Yes | Unspecified | Yes | Yes | Yes | Yes | Yes | N/A | Minimal | None | None | High |
| | | Cholesterol | High | Yes | Unspecified | Yes | Yes | Yes | Yes | Yes | N/A | Minimal | None | None | High |
| | | Triglycerides | High | Yes | Unspecified | Yes | Yes | Yes | Yes | Yes | N/A | Minimal | None | None | High |
| | | Glucose | High | Yes | Unspecified | Yes | Yes | Yes | Yes | Yes | N/A | Minimal | None | None | High |
| | | Bone density | High | Yes | Unspecified | Yes | Yes | Yes | Yes | Yes | N/A | Minimal | None | None | High |
| | | Flexibility | High | Yes | Unspecified | Yes | Yes | Yes | Yes | Yes | N/A | Minimal | None | None | High |
| | | Strength | High | Yes | Unspecified | Yes | Yes | Yes | Yes | Yes | N/A | Minimal | None | None | High |
| ***Teixeira-Machado, Arida & de Jesus Mari, 2019*** | | | | | | | | | | | | | | |
| Baniqued et al. | 2018 | Cardio-respiratory fitness | High | Yes | Unspecified | Yes | No (inclusion criteria revised from MMSE = 23 to MMSE = 27 for this analysis) | Yes | Yes | Yes | N/A | Minimal (higher cognitive performance unlikely to impact certainty of general outcome classifications in umbrella review) | None | None | High |
| | | Executive function | High | Yes | Unspecified | Yes | No (inclusion criteria revised from MMSE = 23 to MMSE = 27 for this analysis) | Yes | Yes | Yes | N/A | Minimal (higher cognitive performance unlikely to impact certainty of general outcome classifications in umbrella review) | None | None | High |
| | | Perceptual speed | High | Yes | Unspecified | Yes | No (inclusion | Yes | Yes | Yes | N/A | Minimal (higher cognitive | None | None | High |

| | | | | | | | | | | | | | | | |
|---|---|---|---|---|---|---|---|---|---|---|---|---|---|---|---|
| | | | | | | | criteria revised from MMSE = 23 to MMSE = 27 for this analysis) | | | | | performance unlikely to impact certainty of general outcome classifications in umbrella review) | | | |
| | | Episodic memory | High | Yes | Unspecified | Yes | No (inclusion criteria revised from MMSE = 23 to MMSE = 27 for this analysis) | Yes | Yes | Yes | N/A | Minimal (higher cognitive performance unlikely to impact certainty of general outcome classifications in umbrella review) | None | None | High |
| | | Vocabulary | High | Yes | Unspecified | Yes | No (inclusion criteria revised from MMSE = 23 to MMSE = 27 for this analysis) | Yes | Yes | Yes | N/A | Minimal (higher cognitive performance unlikely to impact certainty of general outcome classifications in umbrella review) | None | None | High |
| Ehlers et al. | 2017 | Loneliness | High | Yes | Unspecified | Yes | Yes | Yes | Yes (validated measure, although no inactive control; comparison vs exercise only) | Yes | N/A | Serious (no inactive control reported) | None | None | Moderate |
| | | Social Support | High | Yes | Unspecified | Yes | Yes | Yes | Yes (validated measure, although no inactive control; comparison vs exercise only) | Yes | N/A | Serious (no inactive control reported) | None | None | Moderate |

| | | | | | | | | | | | | | | |
|---|---|---|---|---|---|---|---|---|---|---|---|---|---|---|
| | | Stress | High | Yes | Unspecified | Yes | Yes | Yes | Yes (validated measure, although no inactive control; comparison vs exercise only) | Yes | N/A | Serious (no inactive control reported) | None | None | Moderate |
| | | Lifestyle/ quality of life | High | Yes | Unspecified | Yes | Yes | Yes | Yes, but (validated outcome measures, but only pre-post analysis within groups; no comparisons between control & intervention groups) | Yes | N/A | Very Serious (pre-post analyses of control & experimental groups only, effectively a single group study) | None | None | Low |
| Kattenstroth et al. | 2013 | Attention | High | Yes | Unspecified | Yes | Yes | Yes | Yes, but (validated outcome measures, but only pre-post analysis within groups; no comparisons between control & intervention groups) | Yes | N/A | Very Serious (pre-post analyses of control & experimental groups only, effectively a single group study) | None | None | Low |
| | | Intelligence (nonverbal) | High | Yes | Unspecified | Yes | Yes | Yes | Yes, but (validated outcome measures, but only pre-post analysis within groups; no comparisons | Yes | N/A | Very Serious (pre-post analyses of control & experimental groups only, effectively a single group study) | None | None | Low |

| | | | | | | | | | | | | | | |
|---|---|---|---|---|---|---|---|---|---|---|---|---|---|---|
| | | | | | | | | between control & intervention groups) | | | | | | |
| | Fluid intelligence | High | Yes | Unspecified | Yes | Yes | Yes | Yes, but (validated outcome measures, but only pre-post analysis within groups; no comparisons between control & intervention groups) | Yes | N/A | Very Serious (pre-post analyses of control & experimental groups only, effectively a single group study) | None | None | Low |
| | Reaction time | High | Yes | Unspecified | Yes | Yes | Yes | Yes, but (validated outcome measures, but only pre-post analysis within groups; no comparisons between control & intervention groups) | Yes | N/A | Very Serious (pre-post analyses of control & experimental groups only, effectively a single group study) | None | None | Low |
| | Balance | High | Yes | Unspecified | Yes | Yes | Yes | Yes, but (validated outcome measures, but only pre-post analysis within groups; no comparisons between control & intervention groups) | Yes | N/A | Very Serious (pre-post analyses of control & experimental groups only, effectively a single group study) | None | None | Low |

| | | | | | | | | | | | | | | | |
|---|---|---|---|---|---|---|---|---|---|---|---|---|---|---|---|
| | | Fine-motor performance | High | Yes | Unspecified | Yes | Yes | Yes | Yes, but (validated outcome measures, but only pre-post analysis within groups; no comparisons between control & intervention groups) | Yes | N/A | Very Serious (pre-post analyses of control & experimental groups only, effectively a single group study) | None | None | Low |
| | | Tactile performance | High | Yes | Unspecified | Yes | Yes | Yes | Yes, but (validated outcome measures, but only pre-post analysis within groups; no comparisons between control & intervention groups) | Yes | N/A | Very Serious (pre-post analyses of control & experimental groups only, effectively a single group study) | None | None | Low |
| | | Cardio-respiratory fitness | High | Yes | Unspecified | Yes | Yes | Yes | Yes, but (validated outcome measures, but only pre-post analysis within groups; no comparisons between control & intervention groups) | Yes | N/A | Very Serious (pre-post analyses of control & experimental groups only, effectively a single group study) | None | None | Low |
| Muller et al. | 2017 | Cardiovascular fitness | High | Yes | Unspecified | No (52 enrolled, final analysis on 22 subjects) | No (analyses only performed on those with | Yes | Yes | Yes | N/A | Serious (high % lost to follow up, no intention-to-treat analysis) | None | None | Moderate |

| | | | | | | | | | | | | | | | |
|---|---|---|---|---|---|---|---|---|---|---|---|---|---|---|---|
| | | | | | | | 70%+ participation) | | | | | | | | |
| | | Memory | High | Yes | Unspecified | No (52 enrolled, final analysis on 22 subjects) | No (analyses only performed on those with 70%+ participation) | No (some neuropsychological tests excluded from reporting for unspecified rationale) | Yes | Yes | N/A | Very serious (high % lost to follow up, incomplete outcome reporting, no intention-to-treat analysis) | None | None | Low |
| | | Attention | High | Yes | Unspecified | No (52 enrolled, final analysis on 22 subjects) | No (analyses only performed on those with 70%+ participation) | No (some neuropsychological tests excluded from reporting for unspecified rationale) | Yes | Yes | N/A | Very serious (high % lost to follow up, incomplete outcome reporting, no intention-to-treat analysis) | None | None | Low |
| | | BDNF | High | Yes | Unspecified | No (52 enrolled, final analysis on 22 subjects) | No (analyses only performed on those with 70%+ participation) | Yes | Yes | Yes | N/A | Serious (high % lost to follow up, no intention-to-treat analysis) | None | None | Moderate |
| | | Cognitive structure | High | Yes | Unspecified | No (52 enrolled, final analysis on 22 subjects) | No (analyses only performed on those with 70%+ participation) | Yes | Yes | Yes | N/A | Serious (high % lost to follow up, no intention-to-treat analysis) | None | None | Moderate |
| Rehfeld et al. | 2017 | Balance | High | Yes | Unspecified | No (52 enrolled, final analysis on 26 subjects) | No (analyses only performed on those with 70%+ participation) | Yes | Yes | Yes | N/A | Serious (high % lost to follow up, no intention-to-treat analysis) | None | None | Moderate |
| | | Hippocampal grey matter volume | High | Yes | Unspecified | No (52 enrolled, final analysis on 26 subjects) | No (analyses only performed on those with 70%+ participation) | Yes | Yes | Yes | N/A | Serious (high % lost to follow up, no intention-to-treat analysis) | None | None | Moderate |
| ***Veronese et al., 2017*** | | | | | | | | | | | | | | | |

| | | | | | | | | | | | | | | | |
|---|---|---|---|---|---|---|---|---|---|---|---|---|---|---|---|
| da Silva Borges | 2014 | Falls | High | Yes | Unspecified | Yes | Yes | Yes | Yes | Yes | N/A | Minimal | None | None | High |
| Pichierri, Murer & de Bruin | 2012 | Fear of falling | High | Yes | No | Borderline (9/31 lost) | No (analysis only performed on individuals who completed >75% of intervention) | Yes | Yes | Yes | N/A | Serious (no intention to treat analysis + moderate % lost to follow up) | None | None | Moderate |
| Merom et al. | 2016 | Falls | High | Yes | No | Yes (~80%) | Yes | Yes | Yes | No (cluster randomized by retirement center) | N/A | Minimal | None | None | High |
| | | Falls risk | High | Yes | No | Yes (~80%) | Yes | Yes | Yes | No (cluster randomized by retirement center) | N/A | Minimal | None | None | High |
| Wu et al. | 2016 | Fear of falling | High | Yes | Unspecified | Yes | Yes | Yes | Yes | Yes | N/A | Minimal | None | None | High |
| | | Falls | High | Yes | Unspecified | Yes | Yes | Yes | Yes | Yes | N/A | Minimal | None | None | High |

*Table S10. GRADE Certainty of Evidence ratings for each outcome measure from individual non-randomized studies of <u>dance</u> participation contained in included reviews.*

| *Author* | *Year* | *Outcome* | ***GRADE Initial quality*** | *Eligibility criteria developed and appropriately applied* | *Exposed/unexposed sourced from a common population?* | *Appropriate & rigorous measurement of exposure & outcome* | *Identical surveillance for outcome in exposed & unexposed* | *Known confounders adequately measured, controlled & adjusted for?* | *Follow up period complete* | ***GRADE Bias risk*** | ***GRADE Indirectness*** | ***GRADE up rating*** | ***GRADE Certainty of Evidence*** |
|---|---|---|---|---|---|---|---|---|---|---|---|---|---|
| | | | | | | ***Burkhardt & Brennan, 2012*** | | | | | | | |
| Adiputra et al | 1996 | VO2 max | Low | Yes | Yes | Yes | Yes | Yes | Yes | Minimal | None | None | Low |
| Bennell et al | 2000 | Bone mineral density | Low | Yes | Yes | Yes | N/A (cross-sectional) | Yes | N/A (cross-sectional) | Minimal | None | None | Low |
| Blackman et al | 1988 | VO2 max | Low | Yes | Yes | No (pre-post analyses and comparison of post-test values only; no analysis of change in parameters in dance vs control groups) | Yes | Yes | Yes | Serious (pre-post analyses & post-test comparisons between dance & control groups only) | None | None | Very Low |
| | | Sit and reach | Low | Yes | Yes | No (pre-post analyses and comparison of post-test values only; no analysis of change in parameters in dance vs control groups) | Yes | Yes | Yes | Serious (pre-post analyses & post-test comparisons between dance & control groups only) | None | None | Very Low |
| | | Strength | Low | Yes | Yes | No (pre-post analyses and comparison of post-test values only; no analysis of change in parameters in dance vs control groups) | Yes | Yes | Yes | Serious (pre-post analyses & post-test comparisons between dance & control groups only) | None | None | Very Low |
| | | % body fat | Low | Yes | Yes | No (pre-post analyses and comparison of post-test values only; no analysis of change in parameters in dance vs control groups) | Yes | Yes | Yes | Serious (pre-post analyses & post-test comparisons between dance & control groups only) | None | None | Very Low |
| | | Self esteem | Low | Yes | Yes | No (pre-post analyses and comparison of post-test values only; no analysis of change in parameters in dance vs control groups) | Yes | Yes | Yes | Serious (pre-post analyses & post-test comparisons between dance & control groups only) | None | None | Very Low |
| | | Body satisfaction (Cathexis) | Low | Yes | Yes | No (pre-post analyses and comparison of post-test values only; no analysis of change in parameters in dance vs control groups) | Yes | Yes | Yes | Serious (pre-post analyses & post-test comparisons between dance & control groups only) | None | None | Very Low |
| | | Blood Pressure | Low | Yes | Yes | No (pre-post analyses and comparison of post-test values only; no analysis of | Yes | Yes | Yes | Serious (pre-post analyses & post-test comparisons between | None | None | Very Low |

| | | | | | | | | | | | | | |
|---|---|---|---|---|---|---|---|---|---|---|---|---|---|
| | | | | | | change in parameters in dance vs control groups) | | | | dance & control groups only) | | | |
| | | Body mass | Low | Yes | Yes | No (pre-post analyses and comparison of post-test values only; no analysis of change in parameters in dance vs control groups) | Yes | Yes | Yes | Serious (pre-post analyses & post-test comparisons between dance & control groups only) | None | None | Very Low |
| Daley & Buchanan | 1999 | Self-perception | Low | Yes | Yes | Yes | Yes | Yes | Yes | Minimal | None | None | Low |
| Leste & Rust | 1990 | Anxiety | Low | Yes | Yes | Yes | Yes | Yes | Yes | Minimal | None | None | Low |
| Matthews et al | 2006 | Bone mineral density | Low | Yes | Yes | Yes | Yes | Yes | Yes | Minimal | None | None | Low |
| Steinberg et al | 2008 | Musculoskeletal development | Low | Yes | Yes | Yes | N/A (cross-sectional) | Yes | N/A (cross-sectional) | Minimal | None | None | Low |
| Viscki-Stalec et al | 2007 | Core strength | Low | Yes | Yes | Yes | Yes | Yes | Yes | Minimal | None | None | Low |
| | | Power | Low | Yes | Yes | Yes | Yes | Yes | Yes | Minimal | None | None | Low |
| | | Agility | Low | Yes | Yes | Yes | Yes | Yes | Yes | Minimal | None | None | Low |
| ***Clarke et al., 2018*** | | | | | | | | | | | | | |
| Ambegaonkar et al. | 2013 | Balance | Low | | | | *Rating extracted from review* | | | | | | Low |
| Bronner | 2012 | Balance | Low | | | | *Rating extracted from review* | | | | | | Low |
| Casabona et al. | 2016 | Balance | Low | | | | *Rating extracted from review* | | | | | | Low |
| Crotts et al. | 1996 | Balance | Low | | | | *Rating extracted from review* | | | | | | Low |
| Gerbino, Griffin & Zurakowski | 2007 | Balance | Low | | | | *Rating extracted from review* | | | | | | Low |
| Golomer & Dupui | 2000 | Balance | Low | | | | *Rating extracted from review* | | | | | | Low |
| Golomer et al. | 1999 | Balance | Low | | | | *Rating extracted from review* | | | | | | Low |
| Golomer et al. | 2009 | Balance | Low | | | | *Rating extracted from review* | | | | | | Low |
| Golomer et al. | 2010 | Balance | Low | | | | *Rating extracted from review* | | | | | | Low |
| Golomer, Dupui & Monod | 1997 | Balance | Low | | | | *Rating extracted from review* | | | | | | Low |
| Guillou, Dupui & Golomer | 2007 | Balance | Low | | | | *Rating extracted from review* | | | | | | Low |
| Hopper et al. | 2014 | Balance | Low | | | | *Rating extracted from review* | | | | | | Low |
| Hugel et al. | 1999 | Balance | Low | | | | *Rating extracted from review* | | | | | | Low |
| Jarvis, Smith & Kulig | 2014 | Balance | Low | | | | *Rating extracted from review* | | | | | | Low |
| Kiefer et al. | 2011 | Balance | Low | | | | *Rating extracted from review* | | | | | | Low |

| | | | | | | | | | | | | | |
|---|---|---|---|---|---|---|---|---|---|---|---|---|---|
| Kilroy et al. | 2016 | Balance | Low | | | | *Rating extracted from review* | | | | | | Low |
| Krityakiarana & Jongkamonwiwat | 2016 | Balance | Low | | | | *Rating extracted from review* | | | | | | Low |
| Lin et al. | 2011 | Balance | Low | | | | *Rating extracted from review* | | | | | | Low |
| Lin et al. | 2014 | Balance | Low | | | | *Rating extracted from review* | | | | | | Low |
| Perez et al | 2014 | Balance | Low | | | | *Rating extracted from review* | | | | | | Low |
| Perrin et al. | 2002 | Balance | Low | | | | *Rating extracted from review* | | | | | | Low |
| Rein et al. | 2011 | Balance | Low | | | | *Rating extracted from review* | | | | | | Low |
| Schmit, Regis & Riley | 2005 | Balance | Low | | | | *Rating extracted from review* | | | | | | Low |
| Schmitt, Kuni & Sabo | 2005 | Balance | Low | | | | *Rating extracted from review* | | | | | | Low |
| Simmons | 2005 | Balance | Low | | | | *Rating extracted from review* | | | | | | Low |
| Simmons | 2005 | Balance | Low | | | | *Rating extracted from review* | | | | | | Low |
| Thullier & Moufti | 2004 | Balance | Low | | | | *Rating extracted from review* | | | | | | Low |
| ***Costa, Ferreira & Felicio, 2013*** | | | | | | | | | | | | | |
| Cheng et al | 2011 | Balance | Low | Yes | Yes | Yes | N/A (cross sectional) | Yes | N/A (cross sectional) | Minimal | None | None | Low |
| Gillou, Dupu & Golomer | 2007 | Balance | Low | Yes | Yes | Yes | N/A (cross sectional) | Yes | N/A (cross sectional) | Minimal | None | None | Low |
| Thiesen & Sumiya | 2011 | Balance | Low | Yes | Yes | Yes | N/A (cross sectional) | Yes | N/A (cross sectional) | Minimal | None | None | Low |
| ***Cugusi et al., 2019*** | | | | | | | | | | | | | |
| Araneta & Tanori | 2015 | Blood pressure | Low | Yes | N/A (single group study; no control group) | No (single group uncontrolled study) | N/A (single group study; no control group) | Unclear | Yes | Serious (single group study) | None | None | Very Low |
| | | Triglycerides | Low | Yes | N/A (single group study; no control group) | No (single group uncontrolled study) | N/A (single group study; no control group) | Unclear | Yes | Serious (single group study) | None | None | Very Low |
| | | HDL cholesterol | Low | Yes | N/A (single group study; no control group) | No (single group uncontrolled study) | N/A (single group study; no control group) | Unclear | Yes | Serious (single group study) | None | None | Very Low |
| | | Body mass | Low | Yes | N/A (single group study; no control group) | No (single group uncontrolled study) | N/A (single group study; | Unclear | Yes | Serious (single group study) | None | None | Very Low |

| | | | | | | | | | | | | | |
|---|---|---|---|---|---|---|---|---|---|---|---|---|---|
| | | | | | | | no control group) | | | | | | |
| | | Glucose | Low | Yes | N/A (single group study; no control group) | No (single group uncontrolled study) | N/A (single group study; no control group) | Unclear | Yes | Serious (single group study) | None | None | Very Low |
| Cugusi et al | 2016 | Blood pressure | Low | Yes | N/A (single group study; no control group) | No (single group uncontrolled study) | N/A (single group study; no control group) | Unclear | Yes | Serious (single group study) | None | None | Very Low |
| | | % fat mass | Low | Yes | N/A (single group study; no control group) | No (single group uncontrolled study) | N/A (single group study; no control group) | Unclear | Yes | Serious (single group study) | None | None | Very Low |
| | | Waist circumference | Low | Yes | N/A (single group study; no control group) | No (single group uncontrolled study) | N/A (single group study; no control group) | Unclear | Yes | Serious (single group study) | None | None | Very Low |
| | | Body mass | Low | Yes | N/A (single group study; no control group) | No (single group uncontrolled study) | N/A (single group study; no control group) | Unclear | Yes | Serious (single group study) | None | None | Very Low |
| | | BMI | Low | Yes | N/A (single group study; no control group) | No (single group uncontrolled study) | N/A (single group study; no control group) | Unclear | Yes | Serious (single group study) | None | None | Very Low |
| Krishnan et al | 2015 | Body mass | Low | Yes | N/A (single group study; no control group) | Yes | N/A (single group study; no control group) | Yes | Somewhat (>30% lost to follow up) | Very serious (single group uncontrolled study, high % lost to follow up) | None | None | Very Low |
| | | BMI | Low | Yes | N/A (single group study; no control group) | Yes | N/A (single group study; no control group) | Yes | Somewhat (>30% lost to follow up) | Very serious (single group uncontrolled study, high % lost to follow up) | None | None | Very Low |
| | | % body fat | Low | Yes | N/A (single group study; no control group) | No (bioelectric impedance notably unreliable) | N/A (single group study; no control group) | Yes | Somewhat (>30% lost to follow up) | Very serious (single group uncontrolled study, high % lost to follow up, unreliable outcomes) | None | None | Very Low |
| | | Waist circumference | Low | Yes | N/A (single group study; no control group) | Yes | N/A (single group study; no control group) | Yes | Somewhat (>30% lost to follow up) | Very serious (single group uncontrolled study, high % lost to follow up) | None | None | Very Low |

| | | | | | | | | | | | | | |
|---|---|---|---|---|---|---|---|---|---|---|---|---|---|
| | | Blood pressure | Low | Yes | N/A (single group study; no control group) | Yes | N/A (single group study; no control group) | Yes | Somewhat (>30% lost to follow up) | Very serious (single group uncontrolled study, high % lost to follow up) | None | None | Very Low |
| | | Glucose | Low | Yes | N/A (single group study; no control group) | Yes | N/A (single group study; no control group) | Yes | Somewhat (>30% lost to follow up) | Very serious (single group uncontrolled study, high % lost to follow up) | None | None | Very Low |
| | | HbA1c | Low | Yes | N/A (single group study; no control group) | Yes | N/A (single group study; no control group) | Yes | Somewhat (>30% lost to follow up) | Very serious (single group uncontrolled study, high % lost to follow up) | None | None | Very Low |
| | | Insulin | Low | Yes | N/A (single group study; no control group) | Yes | N/A (single group study; no control group) | Yes | Somewhat (>30% lost to follow up) | Very serious (single group uncontrolled study, high % lost to follow up) | None | None | Very Low |
| | | Total cholesterol | Low | Yes | N/A (single group study; no control group) | Yes | N/A (single group study; no control group) | Yes | Somewhat (>30% lost to follow up) | Very serious (single group uncontrolled study, high % lost to follow up) | None | None | Very Low |
| | | HDL cholesterol | Low | Yes | N/A (single group study; no control group) | Yes | N/A (single group study; no control group) | Yes | Somewhat (>30% lost to follow up) | Very serious (single group uncontrolled study, high % lost to follow up) | None | None | Very Low |
| | | Triglycerides | Low | Yes | N/A (single group study; no control group) | Yes | N/A (single group study; no control group) | Yes | Somewhat (>30% lost to follow up) | Very serious (single group uncontrolled study, high % lost to follow up) | None | None | Very Low |
| | | VO2 max | Low | Yes | N/A (single group study; no control group) | No (VO2 max based on estimate of time to completion and HR from 1-mile walk test) | N/A (single group study; no control group) | Yes | Somewhat (>30% lost to follow up) | Very serious (single group uncontrolled study, high % lost to follow up, unreliable outcomes) | None | None | Very Low |
| Micallef | 2014 | Body mass | Low | Yes | N/A (single group study; no control group) | No (single group uncontrolled study) | N/A (single group study; no control group) | No | Yes | Serious (single group study, no apparent consideration for apparent confounding variables (i.e. age)) | None | None | Very Low |
| | | BMI | Low | Yes | N/A (single group study; no control group) | No (single group uncontrolled study) | N/A (single group study; no control group) | No | Yes | Serious (single group study, no apparent consideration for apparent confounding variables (i.e. age)) | None | None | Very Low |

| | | | | | | | | | | | | | |
|---|---|---|---|---|---|---|---|---|---|---|---|---|---|
| Rossmeissl et al | 2016 | VO2 max | Low | Yes | N/A (single group study; no control group) | Yes | N/A (single group study; no control group) | Yes | Yes | Serious (single group uncontrolled study) | None | None | Very Low |
| | | Body mass | Low | Yes | N/A (single group study; no control group) | Yes | N/A (single group study; no control group) | Yes | Yes | Serious (single group uncontrolled study) | None | None | Very Low |
| | | Waist circumference | Low | Yes | N/A (single group study; no control group) | Yes | N/A (single group study; no control group) | Yes | Yes | Serious (single group uncontrolled study) | None | None | Very Low |
| | | BMI | Low | Yes | N/A (single group study; no control group) | Yes | N/A (single group study; no control group) | Yes | Yes | Serious (single group uncontrolled study) | None | None | Very Low |
| | | Blood pressure | Low | Yes | N/A (single group study; no control group) | Yes | N/A (single group study; no control group) | Yes | Yes | Serious (single group uncontrolled study) | None | None | Very Low |
| | | Resting HR | Low | Yes | N/A (single group study; no control group) | Yes | N/A (single group study; no control group) | Yes | Yes | Serious (single group uncontrolled study) | None | None | Very Low |
| | | % fat mass | Low | Yes | N/A (single group study; no control group) | Yes | N/A (single group study; no control group) | Yes | Yes | Serious (single group uncontrolled study) | None | None | Very Low |
| ***Fernandez-Arguelles et al., 2015*** | | | | | | | | | | | | | |
| ***Fong Yan et al., 2018*** | | | | | | | | | | | | | |
| Ford et al. | 1989 | Sit & reach | Low | Yes | Yes | Yes | Yes | Yes | Yes | Minimal | None | None | Low |
| | | Sit ups | Low | Yes | Yes | Yes | Yes | Yes | Yes | Minimal | None | None | Low |
| | | Step test | Low | Yes | Yes | Yes | Yes | Yes | Yes | Minimal | None | None | Low |
| | | Fat mass | Low | Yes | Yes | No (skin calipers not gold standard for % body fat measurement; combining with DXA measurements in meta-analysis questionable) | Yes | Yes | Yes | Serious (skin caliper measurements for % body fat) | None | None | Very Low |
| Heffron, Davey & Cochrane | 1997 | Bone mineral density | Low | Yes | Yes | Yes | Yes | Yes | Yes | Minimal | None | None | Low |

| | | | | | | | | | | | | |
|---|---|---|---|---|---|---|---|---|---|---|---|---|
| Kouli et al | 2009 | Range of motion | Low | Yes | Yes | Yes | Yes | Yes | Yes | Minimal | None | None | Low |
| | | Sit ups | Low | Yes | Yes | Yes | Yes | Yes | Yes | Minimal | None | None | Low |
| | | 1-mile walk/run | Low | Yes | Yes | Yes | Yes | Yes | Yes | Minimal | None | None | Low |
| | | Sit & reach (each leg separated) | Low | Yes | Yes | Yes | Yes | Yes | Yes | Minimal | None | None | Low |
| | | Push ups | Low | Yes | Yes | Yes | Yes | Yes | Yes | Minimal | None | None | Low |
| Mangeri et al. | 2014 | Blood pressure | Low | Yes | Yes | Yes | Yes | Yes | Yes | Minimal | None | None | Low |
| | | Glucose | Low | Yes | Yes | Yes | Yes | Yes | Yes | Minimal | None | None | Low |
| | | 6-minute walk test | Low | Yes | Yes | Yes | Yes | Yes | Yes | Minimal | None | None | Low |
| | | Triglycerides | Low | Yes | Yes | Yes | Yes | Yes | Yes | Minimal | None | None | Low |
| | | Body mass | Low | Yes | Yes | Yes | Yes | Yes | Yes | Minimal | None | None | Low |
| | | BMI | Low | Yes | Yes | Yes | Yes | Yes | Yes | Minimal | None | None | Low |
| | | Waist circumference | Low | Yes | Yes | Yes | Yes | Yes | Yes | Minimal | None | None | Low |
| | | Total cholesterol | Low | Yes | Yes | Yes | Yes | Yes | Yes | Minimal | None | None | Low |
| | | HDL cholesterol | Low | Yes | Yes | Yes | Yes | Yes | Yes | Minimal | None | None | Low |
| Mavridis et al. | 2004 | Range of motion | Low | Yes | Yes | Yes | Yes | Yes | Yes | Minimal | None | None | Low |
| | | Sit ups | Low | Yes | Yes | Yes | Yes | Yes | Yes | Minimal | None | None | Low |
| | | 1-mile walk/run | Low | Yes | Yes | Yes | Yes | Yes | Yes | Minimal | None | None | Low |
| | | Sit & reach (each leg separated) | Low | Yes | Yes | Yes | Yes | Yes | Yes | Minimal | None | None | Low |
| | | Push ups | Low | Yes | Yes | Yes | Yes | Yes | Yes | Minimal | None | None | Low |
| | | Skinfold thickness | Low | Yes | Yes | Yes | Yes | Yes | Yes | Minimal | None | None | Low |
| Milburn & Butts | 1983 | VO2 max | Low | Yes | Yes | Yes | Yes | Yes | Yes | Minimal | None | None | Low |
| | | Peak ventilation | Low | Yes | Yes | Yes | Yes | Yes | Yes | Minimal | None | None | Low |
| Shimamoto et al. | 1998 | VO2 max | Low | Yes | Yes | Yes | Yes | Yes | Yes | Minimal | None | None | Low |
| | | VO2 at anaerobic threshold | Low | Yes | Yes | Yes | Yes | Yes | Yes | Minimal | None | None | Low |
| | | Body mass | Low | Yes | Yes | Yes | Yes | Yes | Yes | Minimal | None | None | Low |
| | | BMI | Low | Yes | Yes | Yes | Yes | Yes | Yes | Minimal | None | None | Low |
| | | % fat mass | Low | Yes | Yes | Yes | Yes | Yes | Yes | Minimal | None | None | Low |
| | | Waist circumference | Low | Yes | Yes | Yes | Yes | Yes | Yes | Minimal | None | None | Low |

| | | | | | | | | | | | | |
|---|---|---|---|---|---|---|---|---|---|---|---|---|
| | | Skinfold thickness | Low | Yes | Yes | Yes | Yes | Yes | Yes | Minimal | None | None | Low |
| Viskic et al | 2007 | Sit & reach | Low | Yes | Yes | Yes | Yes | Yes | Yes | Minimal | None | None | Low |
| | | Range of motion | Low | Yes | Yes | Yes | Yes | Yes | Yes | Minimal | None | None | Low |
| | | Step test | Low | Yes | Yes | Yes | Yes | Yes | Yes | Minimal | None | None | Low |
| | | Body mass | Low | Yes | Yes | Yes | Yes | Yes | Yes | Minimal | None | None | Low |
| | | % fat mass | Low | Yes | Yes | Yes | Yes | Yes | Yes | Minimal | None | None | Low |
| ***Hwang & Braun, 2015*** | | | | | | | | | | | | |
| Ferrufino et al. | 2011 | Balance | Low | Yes | Yes | Yes | Yes | Yes | Yes | Minimal | None | None | Low |
| ***Keogh et al., 2009*** | | | | | | | | | | | | |
| Kudlacek et al | 1997 | Bone density (peripheral and spinal) | Low | Yes | Yes | No (single group uncontrolled study) | Yes | Yes | Yes | Serious (single group uncontrolled study | None | None | Very Low |
| ***Letton, Thom, & Ward, 2020*** | | | | | | | | | | | | |
| Hedgepeth | 1987 | Strength | Low | Yes | Yes | Yes | N/A (cross sectional) | Yes | N/A (cross-sectional) | Minimal | None | None | Low |
| | | Power | Low | Yes | Yes | Yes | N/A (cross sectional) | Yes | N/A (cross-sectional) | Minimal | None | None | Low |
| | | Endurance (muscular) | Low | Yes | Yes | Yes | N/A (cross sectional) | Yes | N/A (cross-sectional) | Minimal | None | None | Low |
| Kujala et al | 1997 | Flexibility | Low | Yes | Yes | Yes | N/A (cross sectional) | Yes | N/A (cross-sectional) | Minimal | None | None | Low |
| Moller & Masharawi | 2011 | Flexibility | Low | Yes | Yes | Yes | Yes | Yes | Yes | Minimal | None | None | Low |
| | | Body composition | Low | Yes | Yes | Yes | Yes | Yes | Yes | Minimal | None | None | Low |
| | | Posture | Low | Yes | Yes | Yes | Yes | Yes | Yes | Minimal | None | None | Low |
| Ozdinc & Turan | 2016 | Foot anthropometrics | Low | Yes | Yes | Yes | N/A (cross sectional) | Yes | N/A (cross-sectional) | Minimal | None | None | Low |
| Pigeon et al | 1997 | Growth | Low | Yes | Yes | Yes | No (prospective for dancers; retrospective for controls) | Yes | Yes | Serious (prospective data collection for dancers; retrospective data collection for controls | None | None | Very Low |
| | | Puberty onset | Low | Yes | Yes | Yes | No (prospective for dancers; | Yes | Yes | Serious (prospective data collection for dancers; retrospective | None | None | Very Low |

| | | | | | | | | | | | | | |
|---|---|---|---|---|---|---|---|---|---|---|---|---|---|
| | | | | | | | retrospective for controls) | | | data collection for controls | | | |
| ***Liu, Shen & Tsai, 2020*** | | | | | | | | | | | | | |
| ***Meng et al., 2020*** | | | | | | | | | | | | | |
| Coubard et al | 2011 | Executive function | Low | Yes | Yes | Yes | Yes | Yes | Yes | Minimal | None | None | Low |
| Hackney et al | 2015 | Global cognition | Low | Yes | Yes | Yes | Yes | Yes | No (high % (40%) not included in final analysis due to noncompliance (<100% compliance)) | Serious (high % lost to follow up) | Non | None | Very Low |
| | | Executive function | | Yes | Yes | Yes | Yes | Yes | No (high % (40%) not included in final analysis due to noncompliance (<100% compliance)) | Serious (high % lost to follow up) | Non | None | Very Low |
| Kim et al | 2011 | Global cognition | Low | Yes | Yes | Yes | Yes | Yes | Yes | Minimal | None | None | Low |
| | | Executive function | Low | Yes | Yes | Yes | Yes | Yes | Yes | Minimal | None | None | Low |
| ***Rodrigues-Krause et al., 2016*** | | | | | | | | | | | | | |
| ***Rodrigues-Krause, Krause & Reischak-Oliveira, 2019*** | | | | | | | | | | | | | |
| Alpert et al | 2009 | Depression | Low | Yes | N/A (single group study; no control group) | Yes | N/A (single group study; no control group) | Yes | Yes | Serious (single group uncontrolled study) | None | None | Very Low |
| | | Balance | Low | Yes | N/A (single group study; no control group) | Yes | N/A (single group study; no control group) | Yes | Yes | Serious (single group uncontrolled study) | None | None | Very Low |
| | | Cognition | Low | Yes | N/A (single group study; no control group) | Yes | N/A (single group study; | Yes | Yes | Serious (single group uncontrolled study) | None | None | Very Low |

| | | | | | | | | | | | | | | |
|---|---|---|---|---|---|---|---|---|---|---|---|---|---|---|
| | | | | | | | | no control group) | | | | | | |
| Kattenstroth et al | 2011 | Independent living | Low | Yes | Unclear (both elderly populations but unclear where sourced) | Yes | | N/A (cross-sectional) | Yes | N/A (cross-sectional) | Minimal | None | None | Low |
| | | Cognitive performance | Low | Yes | Unclear (both elderly populations but unclear where sourced) | Yes | | N/A (cross-sectional) | Yes | N/A (cross-sectional) | Minimal | None | None | Low |
| | | Reaction time | Low | Yes | Unclear (both elderly populations but unclear where sourced) | Yes | | N/A (cross-sectional) | Yes | N/A (cross-sectional) | Minimal | None | None | Low |
| | | Balance | Low | Yes | Unclear (both elderly populations but unclear where sourced) | Yes | | N/A (cross-sectional) | Yes | N/A (cross-sectional) | Minimal | None | None | Low |
| | | Mobility | Low | Yes | Unclear (both elderly populations but unclear where sourced) | Yes | | N/A (cross-sectional) | Yes | N/A (cross-sectional) | Minimal | None | None | Low |
| | | Fine motor performance | Low | Yes | Unclear (both elderly populations but unclear where sourced) | Yes | | N/A (cross-sectional) | Yes | N/A (cross-sectional) | Minimal | None | None | Low |
| | | Tactile performance | Low | Yes | Unclear (both elderly populations but unclear where sourced) | Yes | | N/A (cross-sectional) | Yes | N/A (cross-sectional) | Minimal | None | None | Low |
| Kattenstroth et al | 2010 | Independent living | Low | Yes | Unclear (both elderly populations but unclear where sourced) | Yes | | N/A (cross-sectional) | Yes | N/A (cross-sectional) | Minimal | None | None | Low |
| | | Cognitive performance | Low | Yes | Unclear (both elderly populations but unclear where sourced) | Yes | | N/A (cross-sectional) | Yes | N/A (cross-sectional) | Minimal | None | None | Low |
| | | Reaction time | Low | Yes | Unclear (both elderly populations but unclear where sourced) | Yes | | N/A (cross-sectional) | Yes | N/A (cross-sectional) | Minimal | None | None | Low |
| | | Balance | Low | Yes | Unclear (both elderly populations | Yes | | N/A (cross-sectional) | Yes | N/A (cross-sectional) | Minimal | None | None | Low |

| | | | | | | | | | | | | | |
|---|---|---|---|---|---|---|---|---|---|---|---|---|---|
| | | | | | but unclear where sourced) | | | | | | | | |
| | | Mobility | Low | Yes | Unclear (both elderly populations but unclear where sourced) | Yes | N/A (cross-sectional) | Yes | N/A (cross-sectional) | Minimal | None | None | Low |
| | | Fine motor performance | Low | Yes | Unclear (both elderly populations but unclear where sourced) | Yes | N/A (cross-sectional) | Yes | N/A (cross-sectional) | Minimal | None | None | Low |
| | | Tactile performance | Low | Yes | Unclear (both elderly populations but unclear where sourced) | Yes | N/A (cross-sectional) | Yes | N/A (cross-sectional) | Minimal | None | None | Low |
| Krampe et al | 2010 | Balance | Low | Yes | N/A (single group study; no control group) | Yes | N/A (single group study; no control group) | Yes | Yes | Serious (single group uncontrolled study) | None | None | Very Low |
| | | Mobility | Low | Yes | N/A (single group study; no control group) | Yes | N/A (single group study; no control group) | Yes | Yes | Serious (single group uncontrolled study) | None | None | Very Low |
| Mavrovouniotis et al | 2008 | Anxiety | Low | Yes | Yes (self-report questionnaires, unclear blinding of analysis) | Yes | Yes | Yes | Yes | Minimal | None | None | Low |
| | | Psychological wellbeing | Low | Yes | Yes (self-report questionnaires, unclear blinding of analysis) | Yes | Yes | Yes | Yes | Minimal | None | None | Low |
| | | Psychological distress | Low | Yes | Yes (self-report questionnaires, unclear blinding of analysis) | Yes | Yes | Yes | Yes | Minimal | None | None | Low |
| | | Fatigue | Low | Yes | Yes (self-report questionnaires, unclear blinding of analysis) | Yes | Yes | Yes | Yes | Minimal | None | None | Low |
| Verghese | 2006 | General mental status | Low | Yes | Yes | Yes | N/A (cross-sectional) | Yes | N/A (cross-sectional) | Minimal | None | None | Low |
| | | Memory (episodic) | Low | Yes | Yes | Yes | N/A (cross-sectional) | Yes | N/A (cross-sectional) | Minimal | None | None | Low |
| | | Executive function | Low | Yes | Yes | Yes | N/A (cross-sectional) | Yes | N/A (cross-sectional) | Minimal | None | None | Low |

| | | | | | | | | | | | | | |
|---|---|---|---|---|---|---|---|---|---|---|---|---|---|
| | | Depression | Low | Yes | Yes | Yes | N/A (cross-sectional) | Yes | N/A (cross-sectional) | Minimal | None | None | Low |
| | | Gait | Low | Yes | Yes | Yes | N/A (cross-sectional) | Yes | N/A (cross-sectional) | Minimal | None | None | Low |
| | | Balance | Low | Yes | Yes | Yes | N/A (cross-sectional) | Yes | N/A (cross-sectional) | Minimal | None | None | Low |
| | | Strength | Low | Yes | Yes | Yes | N/A (cross-sectional) | Yes | N/A (cross-sectional) | Minimal | None | None | Low |
| Wallmann et al | 2009 | Balance | Low | Yes | N/A (single group study) | Yes | N/A (single group study; no control group) | Yes | Yes | Serious (single group uncontrolled study) | None | None | Very Low |
| Wu et al | 2011 | Body composition | Low | Yes | Unclear | Yes | N/A (cross sectional) | Yes | N/A (cross sectional) | Minimal | None | None | Low |
| | | Flexibility | Low | Yes | Unclear | Yes | N/A (cross sectional) | Yes | N/A (cross sectional) | Minimal | None | None | Low |
| | | Bone density | Low | Yes | Unclear | Yes | N/A (cross sectional) | Yes | N/A (cross sectional) | Minimal | None | None | Low |
| | | Strength | Low | Yes | Unclear | Yes | N/A (cross sectional) | Yes | N/A (cross sectional) | Minimal | None | None | Low |
| Zhang et al | 2008 | Gait speed | Low | Yes | Yes | Yes | N/A (cross-sectional) | Yes | N/A (cross-sectional) | Minimal | None | None | Low |
| | | Flexibility | Low | Yes | Yes | Yes | N/A (cross-sectional) | Yes | N/A (cross-sectional) | Minimal | None | None | Low |
| | | Reaction time | Low | Yes | Yes | Yes | N/A (cross-sectional) | Yes | N/A (cross-sectional) | Minimal | None | None | Low |
| | | Balance | Low | Yes | Yes | Yes | N/A (cross-sectional) | Yes | N/A (cross-sectional) | Minimal | None | None | Low |
| ***Teixeira-Machado, Arida & de Jesus Mari, 2019*** | | | | | | | | | | | | | |
| Burzynska et al. | 2017 | Fluid intelligence | Low | Yes | Yes | Yes | N/A (cross-sectional) | Yes | N/A (cross-sectional) | Minimal | None | None | Low |
| | | Perceptual speed | Low | Yes | Yes | Yes | N/A (cross-sectional) | Yes | N/A (cross-sectional) | Minimal | None | None | Low |
| | | Spatial/relational memory | Low | Yes | Yes | Yes | N/A (cross-sectional) | Yes | N/A (cross-sectional) | Minimal | None | None | Low |
| | | Balance | Low | Yes | Yes | Yes | N/A (cross-sectional) | Yes | N/A (cross-sectional) | Minimal | None | None | Low |
| | | Cognitive structure & function | Low | Yes | Yes | Yes | N/A (cross-sectional) | Yes | N/A (cross-sectional) | Minimal | None | None | Low |
| ***Veronese et al., 2017*** | | | | | | | | | | | | | |

*Table S11. GRADE Certainty of Evidence ratings for each outcome measure from individual randomized controlled trials (RCTs) of music participation contained in included reviews.*

| *Author* | *Year* | *Outcome* | ***GRADE Initial quality*** | *Allocation concealed* | *Blinded assessors/ data analysts?* | *Low % lost to follow up?* | *Intention to treat analysis?* | *All outcome result(s) reported?* | *Validated outcome measures used* | *Used individual randomization (not cluster randomization)* | *Carryover effects controlled for (cross-over trial)?* | ***GRADE Bias risk*** | ***GRADE Indirectness*** | ***GRADE up rating*** | ***GRADE Certainty of Evidence*** |
|---|---|---|---|---|---|---|---|---|---|---|---|---|---|---|---|
| ***Butzlaff, 2000*** | | | | | | | | | | | | | | | |
| Kelly | 1981 | Reading ability (experimental) | High | Yes | No | Yes | Yes | Yes | Yes | Yes | N/A | Serious (assessments performed by classroom teachers who were likely aware of group allocations) | None | None | Moderate |
| ***Clift et al., 2010*** | | | | | | | | | | | | | | | |
| Unwin, Kenny & Davis | 2002 | Mood | High | Yes | N/A (self-report instrument); unspecified blinding for data analysis | Yes | Yes | Yes | Yes | Yes | Yes | Serious (active music listening control group) | None | None | Moderate |
| | | Mood | High | Yes | N/A (self-report instrument); unspecified blinding for data analysis | Yes | Yes | Yes | Yes | Yes | Yes | Serious (active music listening control group) | None | None | Moderate |
| ***Coffey, Mogilever & Zatorre, 2017*** | | | | | | | | | | | | | | | |
| Slater et al. | 2015 | Speech in Noise | High | Yes | Unclear (likely not, but randomized administration of tasks during speech in noise testing) | No (>50%; 80 initially enrolled, 38 available for analysis) | No (selective analysis of those who participated in whole intervention) | Yes | Yes | Yes | N/a | Very serious (high % lost to follow up + selective analysis of active participants + likely no blinding of assessors/data analysts) | None | None | Low |

| | | | | | | | | | | | | | | | |
|---|---|---|---|---|---|---|---|---|---|---|---|---|---|---|---|
| ***Daykin et al, 2013*** | | | | | | | | | | | | | | | |
| Bittman, Dickson & Coddington | 2009 | Social functioning | High | Yes | No | Yes | Unclear (8 participants excluded for missing 2 or more sessions due to 'illness, discharge, or physician appointments) | Yes | Yes | Yes | Yes | Serious (unblinded data collection & analysis + questionable intent to treat analysis) | None | None | Moderate |
| | | Psychosocial function | High | YEs | No (N/A for assessors; self-report | Yes | Unclear (8 participants excluded for missing 2 or more sessions due to 'illness, discharge, or physician appointments) | Yes | Yes | Yes | Yes | Serious (unblinded data analysis + questionable intent to treat analysis) | None | None | Moderate |
| | | Anger | High | Yes | No (N/A for assessors; self-report | Yes | Unclear (8 participants excluded for missing 2 or more sessions due to 'illness, discharge, or physician appointments) | Yes | Yes | Yes | Yes | Serious (unblinded data analysis + questionable intent to treat analysis) | None | None | Moderate |
| | | Depression | High | Yes | No (N/A for assessors; self-report | Yes | Unclear (8 participants excluded for missing 2 or more sessions due to 'illness, discharge, or physician appointments) | Yes | Yes | Yes | Yes | Serious (unblinded data analysis + questionable intent to treat analysis) | None | None | Moderate |
| Kennedy | 1998 | Self-esteem | High | Yes | No (self-report measure) | Yes (0%) | Yes | Yes | Yes | Yes | N/A | Minimal | None | None | High |
| ***Daykin et al., 2018*** | | | | | | | | | | | | | | | |

| | | | | | | | | | | | | | | | |
|---|---|---|---|---|---|---|---|---|---|---|---|---|---|---|---|
| Coulton et al. | 2015 | Quality of Life (mental) | High | Yes | N/A; unspecified (self report questionnaires; unspecified blinding of analysts) | Yes (20% lost to follow up) | Yes | Yes | Yes | Yes | N/A | Minimal | None | None | High |
| | | Quality of Life (physical) | High | Yes | N/A; unspecified (self report questionnaires; unspecified blinding of analysts) | Yes (20% lost to follow up) | Yes | Yes | Yes | Yes | N/A | Minimal | None | None | High |
| | | Anxiety | High | Yes | N/A; unspecified (self report questionnaires; unspecified blinding of analysts) | Yes (20% lost to follow up) | Yes | Yes | Yes | Yes | N/A | Minimal | None | None | High |
| | | Depression | High | Yes | N/A; unspecified (self report questionnaires; unspecified blinding of analysts) | Yes (20% lost to follow up) | Yes | Yes | Yes | Yes | N/A | Minimal | None | None | High |
| Koelsch, Offermanns & Franzke | 2010 | Depression/ anxiety | High | Yes | N/A; unspecified (self report questionnaires; unspecified blinding of analysts) | N/A (experiment completed in a single day) | Yes | Yes | Yes | Yes | N/A | Minimal | None | None | High |
| | | Fatigue | High | Yes | N/A; unspecified (self report questionnaires; unspecified blinding of analysts) | N/A (experiment completed in a single day) | Yes | Yes | Yes | Yes | N/A | Minimal | None | None | High |
| | | Vigor | High | Yes | N/A; unspecified | N/A (experiment | Yes | Yes | Yes | Yes | N/A | Minimal | None | None | High |

| | | | | | | | | | | | | | |
|---|---|---|---|---|---|---|---|---|---|---|---|---|---|
| | | | (self report questionnaires; unspecified blinding of assessors) | completed in a single day) | | | | | | | | | |
| Mood (Irritability) | High | Yes | N/A; unspecified (self report questionnaires; unspecified blinding of analysts) | N/A (experiment completed in a single day) | Yes | Yes | Yes | Yes | N/A | Minimal | None | None | High |
| Valence / positive affect | High | Yes | N/A; unspecified (self report questionnaires; unspecified blinding of analysts) | N/A (experiment completed in a single day) | Yes | Yes | Yes | Yes | N/A | Minimal | None | None | High |
| Arousal | High | Yes | N/A; unspecified (self report questionnaires; unspecified blinding of analysts) | N/A (experiment completed in a single day) | Yes | Yes | Yes | Yes | N/A | Minimal | None | None | High |
| Mood (Happiness) | High | Yes | N/A; unspecified (self report questionnaires; unspecified blinding of analysts) | N/A (experiment completed in a single day) | Yes | Yes | Yes | Yes | N/A | Minimal | None | None | High |
| Mood (Anger) | High | Yes | N/A; unspecified (self report questionnaires; unspecified blinding of analysts) | N/A (experiment completed in a single day) | Yes | Yes | Yes | Yes | N/A | Minimal | None | None | High |
| Mood (Sadness) | High | Yes | N/A; unspecified (self report questionnaire | N/A (experiment completed in a single day) | Yes | Yes | Yes | Yes | N/A | Minimal | None | None | High |

| | | | | | | | | | | | | | | | |
|---|---|---|---|---|---|---|---|---|---|---|---|---|---|---|---|
| | | | | | s; unspecified blinding of assessors) | | | | | | | | | | |
| | | Anxiety | High | Yes | N/A; unspecified (self report questionnaires; unspecified blinding of analysts) | N/A (experiment completed in a single day) | Yes | Yes | Yes | Yes | N/A | Minimal | None | None | High |
| | | Mood (Disgust) | High | Yes | N/A; unspecified (self report questionnaires; unspecified blinding of analysts) | N/A (experiment completed in a single day) | Yes | Yes | Yes | Yes | N/A | Minimal | None | None | High |
| ***Eells, 2014*** | | | | | | | | | | | | | | | |
| ***Fancourt, Ockelford & Belai, 2014*** | | | | | | | | | | | | | | | |
| Bittman et al | 2001 | Anxiety | High | Yes | Not specified (analysis likely not blinded) | N/A (single visit intervention | No | No (outcomes from 1 of 5 drumming groups 'with the largest effect on NK cell activity' reported only; additional 50 control participants with unreported data and unclear utility) | Yes | Yes | N/A | Very serious (unblinded assessment & analysis + substantial amounts of selective outcome reporting) | None | None | Low |
| | | Depression | High | Yes | Not specified (analysis likely not blinded) | N/A (single visit intervention | No | No (outcomes from 1 of 5 drumming groups 'with the largest effect on NK cell activity' reported only; | Yes | Yes | N/A | Very serious (unblinded assessment & analysis + substantial amounts of selective | None | None | Low |

| | | | | | | | | | | | | | |
|---|---|---|---|---|---|---|---|---|---|---|---|---|---|
| | | | | | | additional 50 control participants with unreported data and unclear utility) | | | | outcome reporting) | | | |
| White blood cell count | High | Yes | Not specified (analysis likely not blinded) | N/A (single visit intervention | No | No (outcomes from 1 of 5 drumming groups ‘with the largest effect on NK cell activity‘ reported only; additional 50 control participants with unreported data and unclear utility) | Yes | Yes | N/A | Very serious (unblinded assessment & analysis + substantial amounts of selective outcome reporting) | None | None | Low |
| Serum cortisol | High | Yes | Not specified (analysis likely not blinded) | N/A (single visit intervention | No | No (outcomes from 1 of 5 drumming groups ‘with the largest effect on NK cell activity‘ reported only; additional 50 control participants with unreported data and unclear utility) | Yes | Yes | N/A | Very serious (unblinded assessment & analysis + substantial amounts of selective outcome reporting) | None | None | Low |
| NK cell activity | High | Yes | Not specified (analysis likely not blinded) | N/A (single visit intervention | No | No (outcomes from 1 of 5 drumming groups ‘with the largest effect on NK cell activity‘ reported only; additional 50 control | Yes | Yes | N/A | Very serious (unblinded assessment & analysis + substantial amounts of selective outcome reporting) | None | None | Low |

| | | | | | | | | | | | | | |
|---|---|---|---|---|---|---|---|---|---|---|---|---|---|
| | | | | | | participants with unreported data and unclear utility) | | | | | | | |
| LAK cell activity | High | Yes | Not specified (analysis likely not blinded) | N/A (single visit intervention | No | No (outcomes from 1 of 5 drumming groups ‘with the largest effect on NK cell activity‘ reported only; additional 50 control participants with unreported data and unclear utility) | Yes | Yes | N/A | Very serious (unblinded assessment & analysis + substantial amounts of selective outcome reporting) | None | None | Low |
| IL-2 | High | Yes | Not specified (analysis likely not blinded) | N/A (single visit intervention | No | No (outcomes from 1 of 5 drumming groups ‘with the largest effect on NK cell activity‘ reported only; additional 50 control participants with unreported data and unclear utility) | Yes | Yes | N/A | Very serious (unblinded assessment & analysis + substantial amounts of selective outcome reporting) | None | None | Low |
| IFN-alpha | High | Yes | Not specified (analysis likely not blinded) | N/A (single visit intervention | No | No (outcomes from 1 of 5 drumming groups ‘with the largest effect on NK cell activity‘ reported only; additional 50 control participants with unreported | Yes | Yes | N/A | Very serious (unblinded assessment & analysis + substantial amounts of selective outcome reporting) | None | None | Low |

| | | | | | | | | | | | | | | |
|---|---|---|---|---|---|---|---|---|---|---|---|---|---|---|
| | | | | | | | | data and unclear utility) | | | | | | |
| | | Plasma DHEA | High | Yes | Not specified (analysis likely not blinded) | N/A (single visit intervention | No | No (outcomes from 1 of 5 drumming groups 'with the largest effect on NK cell activity' reported only; additional 50 control participants with unreported data and unclear utility) | Yes | Yes | N/A | Very serious (unblinded assessment & analysis + substantial amounts of selective outcome reporting) | None | None | Low |
| | | DHEA to cortisol ratio | High | Yes | Not specified (analysis likely not blinded) | N/A (single visit intervention | No | No (outcomes from 1 of 5 drumming groups 'with the largest effect on NK cell activity' reported only; additional 50 control participants with unreported data and unclear utility) | Yes | Yes | N/A | Very serious (unblinded assessment & analysis + substantial amounts of selective outcome reporting) | None | None | Low |
| Kuhn | 2002 | Serum Immunoglobulin-A | High | Yes | Unclear | Yes | Yes | Yes | Yes | Yes | N/A | Minimal (unclear blinding of laboratory analysis of salivary samples unlikely to have major impact) | None | None | High |

***Gick, 2012***

| | | | | | | | | | | | | | | | |
|---|---|---|---|---|---|---|---|---|---|---|---|---|---|---|---|
| ***Gordon, Fehd & McCandliss, 2015*** | | | | | | | | | | | | | | | |
| Cogo-Moreira et al. | 2013 | Reading fluency | High | Yes | Somewhat (intended, but group status often revealed) | Yes (only 13/235 lost to follow up) | Yes | Yes | Yes | No (cluster randomized by school) | N/A | Serious (cluster randomized, plus variable blinding of assessors) | None | None | Moderate |
| | | Phonological awareness (other) | High | Yes | Somewhat (intended, but group status often revealed) | Yes (only 13/235 lost to follow up) | Yes | Yes | Yes | No (cluster randomized by school) | N/A | Serious (cluster randomized, plus variable blinding of assessors) | None | None | Moderate |
| Dege & Schwarzer | 2011 | Phonological awareness (other) | High | Yes | Not specified (likely not blinded) | Yes | Yes | Yes | Yes | Yes | N/A | Serious (unblinded assessors) | None | None | Moderate |
| Herrera et al | 2011 | Phonological awareness (rhyming) | High | Unclear | Unspecified | Yes | Yes | Yes | Yes | Unclear | Yes (6 month break) | Serious (unclear randomization procedures) | None | None | Moderate |
| | | Phonological awareness (other) | High | Unclear | Unspecified | Yes | Yes | Yes | Yes | Unclear | Yes (6 month break) | Serious (unclear randomization procedures) | None | None | Moderate |
| | | Phonological awareness (other) | High | Unclear | Unspecified | Yes | Yes | Yes | Yes | Unclear | Yes (6 month break) | Serious (unclear randomization procedures) | None | None | Moderate |
| | | Phonological awareness (rhyming) | High | Unclear | Unspecified | Yes | Yes | Yes | Yes | Unclear | Yes (6 month break) | Serious (unclear randomization procedures) | None | None | Moderate |
| Moreno et al. | 2011 | Phonological awareness (rhyming) | High | Yes | Yes | Yes | No (5 participants excluded for exhibiting floor or ceiling effects on visual/auditory learning tests at baseline) | Yes | Yes | Yes | N/A | Minimal | None | None | High |
| Moreno et al. | 2009 | Reading fluency | High | Unclear ('pseudo-random-ization') | Unspecified | Yes | Yes | Yes | Yes | Unclear | N/A | Serious (unclear randomization procedures) | None | None | Moderate |

| | | | | | | | | | | | | | | | |
|---|---|---|---|---|---|---|---|---|---|---|---|---|---|---|---|
| ***Hetland, 2000*** | | | | | | | | | | | | | | | |
| Flohr et al | 1998 | Spatial reasoning | High | Yes | Unclear | Yes | Yes | Yes | Yes | Yes | N/A | Minimal | None | None | High |
| Flohr, Miller and Persellin | 1998 | Spatial reasoning, spatial ability | High | Yes | Unclear | Yes | Yes | Yes | Yes | Yes | N/A | Minimal | None | None | High |
| Flohr, Miller and Persellin | 2000 | Spatial reasoning | High | Yes | Unclear | Yes | Yes | Yes | Yes | Yes | N/A | Minimal | None | None | High |
| Rauscher | 1999 | Spatial reasoning | High | Yes | Unclear | Yes | Yes | Yes | Yes | Yes | N/A | Minimal | None | None | High |
| Rauscher | 1999 | Spatial reasoning | High | Yes | Unclear | Yes | Yes | Yes | Yes | Yes | N/A | Minimal | None | None | High |
| ***Jaschke et al., 2013*** | | | | | | | | | | | | | | | |
| Costa-Giomi | 2004 | Language processing | High | Yes | Not specified (analysis likely not blinded) | No (117 enrolled; 81 available for analysis) | Yes | Yes | Yes | Yes | N/A | Serious (unblinded analysis + high % lost to follow up) | None | None | Moderate |
| | | Mathematical ability | High | Yes | Not specified (analysis likely not blinded) | No (117 enrolled; 80 available for analysis) | Yes | Yes | Yes | Yes | N/A | Serious (unblinded analysis + high % lost to follow up) | None | None | Moderate |
| Dege & Schwarzer | 2011 | Reading ability | High | Yes | Not specified (likely not blinded) | Yes | Yes | Yes | Yes | Yes | N/A | Serious (unblinded assessors) | None | None | Moderate |
| Rickard, Bambrick & Gill | 2012 | IQ | High | No (students had some input into their allocated group) | Not specified | Yes | Yes | Yes | Yes | Yes | N/A | Very serious (participants involved in randomization process + unblinded assessment & analysis of outcomes) | None | None | Low |
| | | Writing ability | High | No (students had some input into their | Not specified | Yes | Yes | Yes | Yes | Yes | N/A | Very serious (participants involved in randomization process + unblinded | None | None | Low |

| | | | | | | | | | | | | | | | |
|---|---|---|---|---|---|---|---|---|---|---|---|---|---|---|---|
| | | | | allocated group) | | | | | | | | assessment & analysis of outcomes) | | | |
| Rickard, Bambrick & Gill | 2012 | Reading ability | High | Yes | Not specified | Yes | Yes | Yes | Yes | Yes | N/A | Minimal (administration and distribution of VELS results by Victorian State Government (external to study)) | None | None | High |
| | | Mathematical ability | High | Yes | Not specified | Yes | Yes | Yes | Yes | Yes | N/A | Minimal (administration and distribution of VELS results by Victorian State Government (external to study)) | None | None | High |
| Schellen-berg | 2004 | IQ | High | Yes | Yes | Yes | Yes | Yes | Yes | Yes | N/A | Minimal | None | None | High |
| | | Mathematical ability | High | Yes | Yes | Yes | Yes | Yes | Yes | Yes | N/A | Minimal | None | None | High |
| | | Reading ability | High | Yes | Yes | Yes | Yes | Yes | Yes | Yes | N/A | Minimal | None | None | High |
| ***Moore, 2013*** | | | | | | | | | | | | | | | |
| ***Phillips & Becker, 2019*** | | | | | | | | | | | | | | | |
| Bittman et al. | 2003 | Burnout (emotional exhaustion) | High | No (random selection of subjects, but assign-ment based on schedules) | N/A; unspecified (self-report assessments, but no stated blinding of statistical analyses) | Yes | Yes | Yes | Yes* (validated measures but results not rigorously analyzed (1-tailed t-test, separate analysis of pre/post effects of music & control groups) | Yes | Yes | Very serious (no rigorous statistical analysis - 1-tailed t-test, separate pre/post analysis of music & control groups only) | None | None | Low |

| | | | | | | | | | | | | | | | |
|---|---|---|---|---|---|---|---|---|---|---|---|---|---|---|---|
| | | Burnout (depersonal-ization) | High | No (random selection of subjects, but assign-ment based on schedules) | N/A; unspecified (self-report assessments, but no stated blinding of statistical analyses) | Yes | Yes | Yes | Yes* (validated measures but results not rigorously analyzed (1-tailed t-test, separate analysis of pre/post effects of music & control groups) | Yes | Yes | Very serious (no rigorous statistical analysis - 1-tailed t-test, separate pre/post analysis of music & control groups only) | None | None | Low |
| | | Burnout (personal accomplish-ment) | High | No (random selection of subjects, but assignmen t based on schedules) | N/A; unspecified (self-report assessments, but no stated blinding of statistical analyses) | Yes | Yes | Yes | Yes* (validated measures but results not rigorously analyzed (1-tailed t-test, separate analysis of pre/post effects of music & control groups) | Yes | Yes | Very serious (no rigorous statistical analysis - 1-tailed t-test, separate pre/post analysis of music & control groups only) | None | None | Low |
| | | Mood | High | No (random selection of subjects, but assignmen t based on schedules) | N/A; unspecified (self-report assessments, but no stated blinding of statistical analyses) | Yes | Yes | Yes | Yes* (validated measures but results not rigorously analyzed (1-tailed t-test, separate analysis of pre/post effects of music & control groups) | Yes | Yes | Very serious (no rigorous statistical analysis - 1-tailed t-test, separate pre/post analysis of music & control groups only) | None | None | Low |
| | | | | | | | | ***Raglio et al., 2019*** | | | | | | | |
| Bae | 2011 | Job engagement | High | Yes-ish (random allocation 'after being matched with the available days and | Not specified (unlikely) | Yes | No (10 of initial 30 participants excluded from analysis because of 'time conflicts and | Yes | Yes | Yes | N/A | Very serious (potentially biased randomization; not intention to treat analysis; comparative (rather than inactive) | None | None | Low |

| | | | | | | | | | | | | | |
|---|---|---|---|---|---|---|---|---|---|---|---|---|---|
| | | | times'; potentially not truly unbiased randomiza tion) | | | inconsistent participation') | | | | | control group meaning that only pre-post results considered for purposes of this review) | | | |
| | Anxiety | High | High | Yes-ish (random allocation 'after being matched with the available days and times'; potentially not truly unbiased randomizatio n) | Not specified (unlikely) | Yes | No (10 of initial 30 participants excluded from analysis because of 'time conflicts and inconsistent participation') | Yes | Yes | Yes | Very serious (potentially biased randomization; not intention to treat analysis; comparative (rather than inactive) control group meaning that only pre-post results considered for purposes of this review) | None | None | Low |
| | Mood | High | High | Yes-ish (random allocation 'after being matched with the available days and times'; potentially not truly unbiased randomizatio n) | Not specified (unlikely) | Yes | No (10 of initial 30 participants excluded from analysis because of 'time conflicts and inconsistent participation') | Yes | Yes | Yes | Very serious (potentially biased randomization; not intention to treat analysis; comparative (rather than inactive) control group meaning that only pre-post results considered for purposes of this review) | None | None | Low |
| | Perceived self-efficacy | High | High | Yes-ish (random allocation 'after being matched with | Not specified (unlikely) | Yes | No (10 of initial 30 participants excluded from analysis because of 'time | Yes | Yes | Yes | Very serious (potentially biased randomization; not intention to | None | None | Low |

| | | | | | | | | | | | | | |
|---|---|---|---|---|---|---|---|---|---|---|---|---|---|
| | | | the available days and times'; potentially not truly unbiased randomizatio n) | | | conflicts and inconsistent participation') | | | | treat analysis; comparative (rather than inactive) control group meaning that only pre-post results considered for purposes of this review) | | | |
| Positive affect | High | High | Yes-ish (random allocation 'after being matched with the available days and times'; potentially not truly unbiased randomizatio n) | Not specified (unlikely) | Yes | No (10 of initial 30 participants excluded from analysis because of 'time conflicts and inconsistent participation') | Yes | Yes | Yes | Very serious (potentially biased randomization; not intention to treat analysis; comparative (rather than inactive) control group meaning that only pre-post results considered for purposes of this review) | None | None | Low |
| Negative affect | High | High | Yes-ish (random allocation 'after being matched with the available days and times'; potentially not truly unbiased randomizatio n) | Not specified (unlikely) | Yes | No (10 of initial 30 participants excluded from analysis because of 'time conflicts and inconsistent participation') | Yes | Yes | Yes | Very serious (potentially biased randomization; not intention to treat analysis; comparative (rather than inactive) control group meaning that only pre-post results considered for purposes of this review) | None | None | Low |

| | | | | | | | | | | | | | | |
|---|---|---|---|---|---|---|---|---|---|---|---|---|---|---|
| ***Skingley & Vella-Burrows, 2010*** | | | | | | | | | | | | | | |
| ***Talmini et al, 2017*** | | | | | | | | | | | | | | |
| ***Vaughn, 2000*** | | | | | | | | | | | | | | |
| ***Yap, Kwan & Ang, 2017*** | | | | | | | | | | | | | | |
| Wachi et al. | 2007 | Mood disturbance | High | Yes | Not specified | Yes | Yes (1 of 40) | Yes | Yes | Yes | Yes | Minimal | None | None | High |
| | | Natural killer cell activity (effector:target 25:1) | High | Yes | Not specified | Yes | Yes (1 of 40) | Yes | Yes | Yes | Yes | Minimal | None | None | High |
| | | Natural killer cell activity (effector:target 12:1) | High | Yes | Not specified | Yes | Yes (1 of 40) | Yes | Yes | Yes | Yes | Minimal | None | None | High |
| | | Natural killer cell activity/ regulation | High | Yes | Not specified | Yes | Yes (1 of 40) | Yes | Yes | Yes | Yes | Minimal | None | None | High |
| | | CD-56+ cells | High | Yes | Not specified | Yes | Yes (1 of 40) | Yes | Yes | Yes | Yes | Minimal | None | None | High |
| | | IFN-gamma gene expression (mRNA) | High | Yes | Not specified | Yes | Yes (1 of 40) | Yes | Yes | Yes | Yes | Minimal | None | None | High |
| | | IL-2 gene expression (mRNA) | High | Yes | Not specified | Yes | Yes (1 of 40) | Yes | Yes | Yes | Yes | Minimal | None | None | High |
| | | IL-6 gene expression (mRNA) | High | Yes | Not specified | Yes | Yes (1 of 40) | Yes | Yes | Yes | Yes | Minimal | None | None | High |
| | | IL-10 gene expression (mRNA) | High | Yes | Not specified | Yes | Yes (1 of 40) | Yes | Yes | Yes | Yes | Minimal | None | None | High |
| | | Leukocyte counts | High | Yes | Not specified | Yes | Yes (1 of 40) | Yes | Yes | Yes | Yes | Minimal | None | None | High |

*Table S12. GRADE Certainty of Evidence ratings for each outcome measure from individual non-randomized studies of music participation contained in included reviews.*

| *Author* | *Year* | *Outcome* | ***GRADE Initial quality*** | *Eligibility criteria developed and appropriately applied* | *Exposed/ unexposed sourced from a common population* | *Appropriate & rigorous measurement of exposure & outcome* | *Identical surveillance for outcome in exposed & unexposed* | *Known confounders adequately measured, controlled & adjusted for* | *Follow up period complete* | ***GRADE Bias risk*** | ***GRADE Indirectness*** | ***GRADE up rating*** | ***GRADE Certainty of Evidence*** |
|---|---|---|---|---|---|---|---|---|---|---|---|---|---|
| | | | | | | ***Butzlaff, 2000*** | | | | | | | |
| College Board | 1988-1998 (not 1993) | Reading ability (correlation) | Low | Yes | Yes | Yes | N/A (cross sectional study) | Yes | N/A (cross sectional) | Minimal | None | '+1 massive sample size with consistent results across years | Moderate |
| Douglas & Willats | 1994 | Reading ability (experimental) | Low | Yes | N/A (no control group; correlational analyses only) | Yes | N/A (cross sectional study) | Yes | N/A (cross sectional) | Minimal | None | None | Low |
| Engdahl | 1994 | Reading ability (correlation) | Low | Yes | Yes | Yes | Yes | Yes | Yes | Minimal | None | None | Low |
| Fetzer | 1994 | Reading ability (experimental) | Low | Yes | Yes | Yes | Yes | Uncertain | Yes | Minimal | None | None | Low |
| Friedman | 1959 | Reading ability (correlation) | Low | Yes | Yes | Yes | Yes | Yes | Yes | Minimal | None | None | Low |
| Groff | 1963 | Reading ability (correlation) | Low | Yes | Yes | Yes | Yes | Unclear | Yes | Minimal | None | None | Low |
| Kvet | 1985 | Reading ability (correlation) | Low | Yes | Yes | Yes | Yes | Yes | Yes | Minimal | None | None | Low |
| Lamar | 1989 | Reading ability (correlation) | Low | Yes | N/A (no control group; correlational analyses only) | Yes | N/A (cross sectional study) | Yes | Yes | Minimal | None | None | Low |
| McCarthy | 1992 | Reading ability (correlation) | Low | Article access unavailable - risk of bias extrapolated from review data | | | | | | | None | None | Low |
| Olanoff & Kirschener | 1969 | Reading ability (experimental) | Low | Article access unavailable - risk of bias extrapolated from review data | | | | | | | None | None | Low |
| Roberts | 1978 | Reading ability (experimental) | Low | Article access unavailable - risk of bias extrapolated from review data | | | | | | | None | None | Low |
| Roskam | 1979 | Reading ability (experimental) | Low | Yes | Yes | Yes | Yes | Yes | Yes | Minimal | None | None | Low |
| Weeden | 1971 | Reading ability (correlation) | Low | Yes | Yes | Yes | Yes | Yes | Yes | Minimal | None | None | Low |
| | | | | | | ***Clift et al, 2010*** | | | | | | | |
| Beck et al. | 2000 | Immunoglobulin A | Low | Yes | N/A (single group analysis) | Yes | N/A (single group study) | Yes | N/A (analysis of acute effects of singing | Serious (single group | None | None | Very Low |

| | | | | | | | | | | | | | |
|---|---|---|---|---|---|---|---|---|---|---|---|---|---|
| | | | | | | | | | rehearsals and performances) | uncontrolled study) | | | |
| | | Cortisol | Low | Yes | N/A (single group analysis) | Yes | N/A (single group study) | Yes | N/A (analysis of acute effects of singing rehearsals and performances) | Serious (single group uncontrolled study) | None | None | Very Low |
| Cohen et al | 2007 | Overall health | Low | Yes | Yes | No (unvalidated, bespoke instrument) | Yes | Yes | No (analysis of less than 50% of enrolled patients included in analysis - only those with available 12 month & 24 month data) | Very serious (unvalidated questionnaire + highly incomplete follow up) | None | None | Very Low |
| | | Health system utilization | Low | Yes | Yes | No (unvalidated, bespoke instrument) | Yes | Yes | No (analysis of less than 50% of enrolled patients included in analysis - only those with available 12 month & 24 month data) | Very serious (unvalidated questionnaire + highly incomplete follow up) | None | None | Very Low |
| | | Medication usage (over the counter + prescription) | Low | Yes | Yes | No (unvalidated, bespoke instrument) | Yes | Yes | No (analysis of less than 50% of enrolled patients included in analysis - only those with available 12 month & 24 month data) | Very serious (unvalidated questionnaire + highly incomplete follow up) | None | None | Very Low |
| | | Falls incidence | Low | Yes | Yes | No (unvalidated, bespoke instrument) | Yes | Yes | No (analysis of less than 50% of enrolled patients included in analysis - only those with | Very serious (unvalidated questionnaire + highly incomplete follow up) | None | None | Very Low |

| | | | | | | | | | | | | | |
|---|---|---|---|---|---|---|---|---|---|---|---|---|---|
| | | | | | | | | | available 12 month & 24 month data) | | | | |
| | | Morale | Low | Yes | Yes | Yes | Yes | Yes | No (analysis of less than 50% of enrolled patients included in analysis - only those with available 12 month & 24 month data) | Serious (incomplete follow up) | None | None | Very Low |
| | | Depression | Low | Yes | Yes | Yes | Yes | Yes | No (analysis of less than 50% of enrolled patients included in analysis - only those with available 12 month & 24 month data) | Serious (incomplete follow up) | None | None | Very Low |
| | | Loneliness | Low | Yes | Yes | Yes | Yes | Yes | No (analysis of less than 50% of enrolled patients included in analysis - only those with available 12 month & 24 month data) | Serious (incomplete follow up) | None | None | Very Low |
| | | Social activity level | Low | Yes | Yes | No (unvalidated, bespoke instrument) | Yes | Yes | No (analysis of less than 50% of enrolled patients included in analysis - only those with available 12 month & 24 month data) | Very serious (unvalidated questionnaire + highly incomplete follow up) | None | None | Very Low |
| Houston et al | 1998 | Anxiety | Low | Yes | Yes | Yes | Yes | Yes | Yes | Minimal | None | None | Low |

| | | | | | | | | | | | | |
|---|---|---|---|---|---|---|---|---|---|---|---|---|
| | | Somatic symptoms | Low | Yes | Yes | Yes | Yes | Yes | Yes | Minimal | None | None | Low |
| | | Depression | Low | Yes | Yes | Yes | Yes | Yes | Yes | Minimal | None | None | Low |
| | | Social functioning | Low | Yes | Yes | Yes | Yes | Yes | Yes | Minimal | None | None | Low |
| | | Life satisfaction | Low | Yes | No (choral members sourced from a retirement home vs comparison group randomly selected from the broader community) | No (some members of comparison group still musically active) | N/A (cross sectional study) | Yes | N/A (cross sectional | Serious (authors acknowledge that comparison group is not a true control group & likely biases results) | None | None | Very Low |
| Wise, Hartmann & Fisher | 1992 | Self-actualization | Low | Yes | No (choral members sourced from a retirement home vs comparison group randomly selected from the broader community) | No (some members of comparison group still musically active) | N/A (cross sectional study) | Yes | N/A (cross sectional | Serious (authors acknowledge that comparison group is not a true control group & likely biases results) | None | None | Very Low |
| | | Social isolation (alienation) | Low | Yes | No (choral members sourced from a retirement home vs comparison group randomly selected from the broader community) | No (some members of comparison group still musically active) | N/A (cross sectional study) | Yes | N/A (cross sectional | Serious (authors acknowledge that comparison group is not a true control group & likely biases results) | None | None | Very Low |
| ***Coffey, Mogilever & Zatorre, 2017*** | | | | | | | | | | | | |
| Baskent & Etienne | 2016 | Speech in speech | Low | Yes | Unclear (matched subjects but source population unclear | Yes | N/A (cross sectional study) | Yes | N/A (cross sectional study | Minimal (source population unclear, but subjects well matched & intergroup education difference statistically controlled) | None | None | Low |
| Bidelman & Weiss | 2014 | Auditory processing | Low | Yes | Unclear | Yes | N/A (cross sectional) | Yes | N/A (cross sectional) | Minimal (unclear source populations but likely both university + robust | None | None | Low |

| | | | | | | | | | | | | | |
|---|---|---|---|---|---|---|---|---|---|---|---|---|---|
| | | | | | | | | | | neurophysiologic testing) | | | |
| Boebinger et al. | 2015 | Speech in noise (Masked speech perception) | Low | Yes | Yes | Yes | N/A (cross sectional study) | No (analysis is underpowered to detect possible significant advantage for musicians | N/A (cross sectional study | Serious (underpowered) | None | None | Very low |
| | | Duration discrimination | Low | Yes | Yes | Yes | N/A (cross sectional study) | Yes | N/A (cross sectional study | Minimal | None | None | Low |
| | | Pitch discrimination | Low | Yes | Yes | Yes | N/A (cross sectional study) | Yes | N/A (cross sectional study | Minimal | None | None | Low |
| | | IQ (nonverbal) | Low | Yes | Yes | Yes | N/A (cross sectional study) | Yes | N/A (cross sectional study | Minimal | None | None | Low |
| | | Selective attention | Low | Yes | Yes | Yes | N/A (cross sectional study) | Yes | N/A (cross sectional study | Minimal | None | None | Low |
| | | Mental flexibility (Motor speed) | Low | Yes | Yes | Yes | N/A (cross sectional study) | Yes | N/A (cross sectional study | Minimal | None | None | Low |
| | | Mental flexibility (Fluid intelligence) | Low | Yes | Yes | Yes | N/A (cross sectional study) | Yes | N/A (cross sectional study | Minimal | None | None | Low |
| | | Mental flexibility (Task switching) | Low | Yes | Yes | Yes | N/A (cross sectional study) | Yes | N/A (cross sectional study | Minimal | None | None | Low |
| Clayton et al | 2016 | IQ (nonverbal) | Low | Yes | Unclear (likely that subjects sourced from university population but not explicitly stated) | Yes | N/A (cross sectional study) | Yes | N/A (cross sectional study | Minimal | None | None | Low |
| | | Executive function (Inhibition control) | Low | Yes | Unclear (likely that subjects sourced from university population but not explicitly stated) | Yes | N/A (cross sectional study) | Yes | N/A (cross sectional study | Minimal | None | None | Low |
| | | Executive function (Cognitive flexibility/goal directed behavior) | Low | Yes | Unclear (likely that subjects sourced from university population but not explicitly stated) | Yes | N/A (cross sectional study) | Yes | N/A (cross sectional study | Minimal | None | None | Low |
| | | Speech in speech | Low | Yes | Unclear (likely that subjects sourced from university population but not explicitly stated) | Yes | N/A (cross sectional study) | Yes | N/A (cross sectional study | Minimal | None | None | Low |

| | | | | | | | | | | | | | |
|---|---|---|---|---|---|---|---|---|---|---|---|---|---|
| | | Selective attention (Multiple object tracking) | Low | Yes | Unclear (likely that subjects sourced from university population but not explicitly stated) | Yes | N/A (cross sectional study) | Yes | N/A (cross sectional study | Minimal | None | None | Low |
| Coffey et al | 2016 | Pitch discrimination | Low | Yes | N/A | Yes | N/A (cross sectional) | No (very small sample of musicians (n=12) for correlational analyses) | N/A (cross sectional | Serious (small sample size undermines confidence in replicability of correlation results) | None | None | Very Low |
| | | Auditory processing | Low | Yes | N/A | Yes | N/A (cross sectional) | No (very small sample of musicians (n=12) for correlational analyses) | N/A (cross sectional | Serious (small sample size undermines confidence in replicability of correlation results) | None | None | Very Low |
| Du & Zatorre | 2016 | Speech in noise | Low | Unclear - conference abstract summary only available, risk of bias extrapolated from review data | | | | | | Unknown | None | None | Low |
| Fuller et al. | 2014 | Speech in noise | Low | Yes | Yes | Yes | N/A (cross sectional) | Yes | N/A (cross sectional | Minimal | None | None | Low |
| | | Vocal emotion identification | Low | Yes | Yes | Yes | N/A (cross sectional) | Yes | N/A (cross sectional | Minimal | None | None | Low |
| | | Melodic contour identification | Low | Yes | Yes | Yes | N/A (cross sectional) | Yes | N/A (cross sectional | Minimal | None | None | Low |
| Lee et al. | 2009 | Auditory processing | Low | Yes | Unclear | Yes | N/A (cross sectional) | Yes | N/A (cross sectional) | Minimal (unclear source populations but likely both university + robust testing) | None | None | Low |
| Musacchia et al. | 2007 | Auditory processing | Low | Yes | Yes | Yes | N/A (cross sectional) | Yes | N/A (cross sectional | Minimal | None | None | Low |
| Musacchia, Strait & Kraus | 2008 | Auditory processing (neural timing) | Low | Yes | Unclear | Yes | N/A (cross sectional) | Yes | N/A (cross sectional) | Minimal (unclear source populations but likely both university + robust testing) | None | None | Low |
| Oxenham et al. | 2003 | Sound in noise | Low | Yes | Unclear | Yes | N/A (cross sectional) | Yes | N/A (cross sectional) | Minimal (unclear source populations but likely both | None | None | Low |

| | | | | | | | | | | | | | |
|---|---|---|---|---|---|---|---|---|---|---|---|---|---|
| | | | | | | | | | | university + robust testing) | | | |
| | | Speech in noise (Frequency selectivity) | Low | Yes | Unclear | Yes | N/A (cross sectional) | Yes | N/A (cross sectional) | Minimal (unclear source populations but likely both university + robust testing) | None | None | Low |
| Parbery-Clark et al. | 2009 | Speech in Noise | Low | Yes | Unclear (likely that subjects sourced from university population but not explicitly stated) | Yes | N/A (cross sectional study) | Yes | N/A (cross sectional study | Minimal | None | None | Low |
| | | Working memory | Low | Yes | Unclear (likely that subjects sourced from university population but not explicitly stated) | Yes | N/A (cross sectional study) | Yes | N/A (cross sectional study | Minimal | None | None | Low |
| | | Pitch discrimination | Low | Yes | Unclear (likely that subjects sourced from university population but not explicitly stated) | Yes | N/A (cross sectional study) | Yes | N/A (cross sectional study | Minimal | None | None | Low |
| Parbery-Clark et al. | 2011 | Speech in noise | Low | Yes | Unclear (musician/nonmusician subjects well matched but source population unclear) | Yes | N/A (cross sectional) | Yes | N/A (cross sectional study | Minimal | None | None | Low |
| | | Auditory processing (temporal acuity) | Low | Yes | Unclear (musician/nonmusician subjects well matched but source population unclear) | Yes | N/A (cross sectional) | Yes | N/A (cross sectional study | Minimal | None | None | Low |
| Parbery-Clark et al. | 2011 | Speech in noise | Low | Yes | Unclear (source population(s) not specified | Yes | N/A (cross sectional) | Yes | N/A (cross sectional) | Minimal (unclear source populations but substantial controlling of potential confounders) | None | None | Low |
| | | IQ (nonverbal) | Low | Yes | Unclear (source population(s) not specified | Yes | N/A (cross sectional) | Yes | N/A (cross sectional) | Minimal (unclear source populations but substantial | None | None | Low |

| | | | | | | | | | | | | |
|---|---|---|---|---|---|---|---|---|---|---|---|---|
| | | | | | | | | | controlling of potential confounders) | | | |
| Parbery-Clark et al. | 2012 | Speech in noise | Low | Yes | Unclear | Yes | N/A (cross sectional) | Yes | N/A (cross sectional) | Minimal (unclear source populations but likely both university + robust testing) | None | None | Low |
| | | IQ (nonverbal) | Low | Yes | Unclear | Yes | N/A (cross sectional) | Yes | N/A (cross sectional) | Minimal (unclear source populations but likely both university + robust testing) | None | None | Low |
| Parbery-Clark et al. | 2012 | Auditory processing | Low | Yes | No (by design) | Yes | N/A (cross sectional) | Yes | N/A (cross sectional | Minimal | None | None | Low |
| Parbery-Clark, Skoe & Kraus | 2009 | Speech in noise | Low | Yes | Unclear | Yes | N/A (cross sectional) | Yes | N/A (cross sectional) | Minimal (unclear source populations but likely both university + robust testing) | None | None | Low |
| Ruggles et al. | 2014 | IQ | Low | Yes | Yes | Yes | N/A (cross sectional study) | Yes | N/A (cross sectional study | Minimal | None | None | Low |
| | | Speech in noise (Voiced speech discrimination) | Low | Yes | Yes | Yes | N/A (cross sectional study) | Yes | N/A (cross sectional study | Minimal | None | None | Low |
| | | Speech in noise (Whispered speech discrimination) | Low | Yes | Yes | Yes | N/A (cross sectional study) | Yes | N/A (cross sectional study | Minimal | None | None | Low |
| Ruggles et al. | 2014 | Pitch discrimination | Low | Yes | Yes | Unclear (unclear if validated pitch discrimination assessment used) | N/A (cross sectional study) | Yes | N/A (cross sectional study | Minimal (unclear about validation of methodology, but methodology considered and robust) | None | None | Low |
| | | Speech in Noise | Low | Yes | Yes | Yes | N/A (cross sectional study) | Yes | N/A (cross sectional study | Minimal | None | None | Low |
| Slater & Kraus | 2016 | Speech in noise | Low | Yes | Unclear (source population(s) not specified) | Yes | N/A (cross sectional) | Yes | N/A (cross sectional) | Minimal (unclear source populations but substantial controlling of | None | None | Low |

| | | | | | | | | | | | | |
|---|---|---|---|---|---|---|---|---|---|---|---|---|
| | | | | | | | | | potential confounders) | | | |
| | | Speech in noise | Low | Yes | Unclear (source population(s) not specified) | Yes | N/A (cross sectional) | Yes | N/A (cross sectional) | Minimal (unclear source populations but substantial controlling of potential confounders) | None | None | Low |
| | | Words in noise | Low | Yes | Unclear (source population(s) not specified) | Yes | N/A (cross sectional) | Yes | N/A (cross sectional) | Minimal (unclear source populations but substantial controlling of potential confounders) | None | None | Low |
| | | IQ (nonverbal) | Low | Yes | Unclear (source population(s) not specified) | Yes | N/A (cross sectional) | Yes | N/A (cross sectional) | Minimal (unclear source populations but substantial controlling of potential confounders) | None | None | Low |
| Strait & Kraus | 2011 | Auditory processing | Low | Yes | Unclear | Yes | N/A (cross sectional) | Yes | N/A (cross sectional | Minimal (unclear source populations but likely both university + robust testing) | None | None | Low |
| Strait et al. | 2010 | Auditory working memory | Low | Yes | Unclear | Yes | N/A (cross sectional) | Yes | N/A (cross sectional) | Minimal (unclear source populations but likely both university + robust testing) | None | None | Low |
| | | Auditory processing (attention) | Low | Yes | Unclear | Yes | N/A (cross sectional) | Yes | N/A (cross sectional) | Minimal (unclear source populations but likely both university + robust testing) | None | None | Low |
| | | Visual attention | Low | Yes | Unclear | Yes | N/A (cross sectional) | Yes | N/A (cross sectional) | Minimal (unclear source populations but | None | None | Low |

| | | | | | | | | | | | | |
|---|---|---|---|---|---|---|---|---|---|---|---|---|
| | | | | | | | | | likely both university + robust testing) | | | |
| | | Frequency discrimination | Low | Yes | Unclear | Yes | N/A (cross sectional) | Yes | N/A (cross sectional) | Minimal (unclear source populations but likely both university + robust testing) | None | None | Low |
| | | Speech in noise (backward masking) | Low | Yes | Unclear | Yes | N/A (cross sectional) | Yes | N/A (cross sectional) | Minimal (unclear source populations but likely both university + robust testing) | None | None | Low |
| | | Speech in noise (forward masking) | Low | Yes | Unclear | Yes | N/A (cross sectional) | Yes | N/A (cross sectional) | Minimal (unclear source populations but likely both university + robust testing) | None | None | Low |
| | | IQ (nonverbal) | Low | Yes | Unclear | Yes | N/A (cross sectional) | Yes | N/A (cross sectional) | Minimal (unclear source populations but likely both university + robust testing) | None | None | Low |
| Swaminathan et al. | 2015 | Speech in noise | Low | Yes | Unclear | Yes | N/A (cross sectional) | No reported controlling or adjustment for confounders other than 'no known neurological conditions' | N/A (cross sectional) | Serious (unclear if musician & nonmusician cohorts sourced from similar population + minimal adjustment for potential confounders (i.e. education)) | None | None | Very low |
| Tierney, Krizman & Kraus | 2015 | Language skills (Phonological awareness) | Low | Yes | Yes | Yes | Yes | Yes | High % lost to follow up (68 enrolled; 40 analyzed) | Serious | None | None | Very Low |

| | | | | | | | | | | | | | |
|---|---|---|---|---|---|---|---|---|---|---|---|---|---|
| | | Language skills (Phonological memory) | Low | Yes | Yes | Yes | Yes | Yes | High % lost to follow up (68 enrolled; 40 analyzed) | Serious | None | None | Very Low |
| | | Language skills (Rapid naming) | Low | Yes | Yes | Yes | Yes | Yes | High % lost to follow up (68 enrolled; 40 analyzed) | Serious | None | None | Very Low |
| | | Auditory processing (Subcortical response consistency) | Low | Yes | Yes | Yes | Yes | Yes | High % lost to follow up (68 enrolled; 40 analyzed) | Serious | None | None | Very Low |
| | | Auditory processing (cortical onset response) | Low | Yes | Yes | Yes | Yes | Yes | High % lost to follow up (68 enrolled; 40 analyzed) | Serious | None | None | Very Low |
| Varnet et al. | 2015 | Speech in noise | Low | Yes | Yes | Yes | N/A (cross sectional) | Yes | N/A (cross sectional | Minimal | None | None | Low |
| | | Auditory processing | Low | Yes | Yes | Yes | N/A (cross sectional) | Yes | N/A (cross sectional | Minimal | None | None | Low |
| Zendel & Alain | 2012 | Speech in noise | Low | Yes | Unclear | Yes | N/A (cross sectional) | Yes | N/A (cross sectional) | Minimal (unclear source populations but likely both university + robust testing) | None | None | Low |
| | | Auditory processing (threshold) | Low | Yes | Unclear | Yes | N/A (cross sectional) | Yes | N/A (cross sectional) | Minimal (unclear source populations but likely both university + robust testing) | None | None | Low |
| | | Auditory processing (temporal acuity (gap detection)) | Low | Yes | Unclear | Yes | N/A (cross sectional) | Yes | N/A (cross sectional) | Minimal (unclear source populations but likely both university + robust testing) | None | None | Low |
| | | Pitch discrimination (Mistuned harmonic detection) | Low | Yes | Unclear | Yes | N/A (cross sectional) | Yes | N/A (cross sectional) | Minimal (unclear source populations but likely both university + robust testing) | None | None | Low |

| | | | | | | | | | | | | |
|---|---|---|---|---|---|---|---|---|---|---|---|---|
| Zendel & Alain | 2009 | Pitch discrimination (Two-tone detection) | Low | Yes | Unclear | Yes | N/A (cross sectional) | Yes | N/A (cross sectional) | Minimal (unclear source populations but likely both university + robust testing) | None | None | Low |
| Zendel et al. | 2015 | Words in noise | Low | Yes | Yes | Yes | N/A (cross sectional) | Yes | N/A (cross sectional) | Minimal (words in noise test procedure appears to be original but based reasonably in precedent) | None | None | Low |
| ***Daykin et al, 2013*** | | | | | | | | | | | | | |
| ***Daykin et al, 2018*** | | | | | | | | | | | | | |
| Cohen et al. | 2006 | Overall health | Low | Yes | Yes | No (unvalidated measure) | Yes | No (statistical analysis of post-intervention data only; no analysis of change from baseline levels vs control) | Yes | Very serious (unvalidated measure + non-rigorous statistical analysis) | None | None | Very Low |
| | | Health system utilization | Low | Yes | Yes | Yes (self-report of health system utilization shown to be valid) | Yes | No (statistical analysis of post-intervention data only; no analysis of change from baseline levels vs control) | Yes | Serious (non-rigorous statistical analysis) | None | None | Very Low |
| | | Medication usage (over the counter) | Low | Yes | Yes | No (unvalidated measure) | Yes | No (statistical analysis of post-intervention data only; no analysis of change from baseline levels vs control) | Yes | Very serious (unvalidated measure + non-rigorous statistical analysis) | None | None | Very Low |
| | | Medication usage (prescription) | Low | Yes | Yes | No (unvalidated measure) | Yes | No (statistical analysis of post-intervention | Yes | Very serious (unvalidated measure + non- | None | None | Very Low |

| | | | | | | | | | | | | | |
|---|---|---|---|---|---|---|---|---|---|---|---|---|---|
| | | | | | | | | data only; no analysis of change from baseline levels vs control) | | rigorous statistical analysis) | | | |
| | | Falls incidence | Low | Yes | Yes | No (unvalidated measure) | Yes | No (statistical analysis of post-intervention data only; no analysis of change from baseline levels vs control) | Yes | Very serious (unvalidated measure + non-rigorous statistical analysis) | None | None | Very Low |
| | | Other health problems | Low | Yes | Yes | No (unvalidated measure) | Yes | No (statistical analysis of post-intervention data only; no analysis of change from baseline levels vs control) | Yes | Very serious (unvalidated measure + non-rigorous statistical analysis) | None | None | Very Low |
| | | Morale | Low | Yes | Yes | Yes | Yes | No (statistical analysis of post-intervention data only; no analysis of change from baseline levels vs control) | Yes | Serious (non-rigorous statistical analysis) | None | None | Very Low |
| | | Depression | Low | Yes | Yes | Yes | Yes | Yes | Yes | Minimal | None | None | Low |
| | | Loneliness | Low | Yes | Yes | Yes | Yes | Yes | Yes | Minimal | None | None | Low |
| | | Social activity level | Low | Yes | Yes | No (unvalidated measure) | Yes | No (statistical analysis of post-intervention data only; no analysis of change from baseline levels vs control) | Yes | Very serious (unvalidated measure + non-rigorous statistical analysis) | None | None | Very Low |
| Cohen | 2009 | Wellbeing | Low | Yes | Yes | Yes | Yes | Yes | N/A (analysis of acute wellbeing effects of a | Minimal | None | None | Low |

| | | | | | | | | | | | | |
|---|---|---|---|---|---|---|---|---|---|---|---|---|
| | | | | | | | | single performance | | | | |
| | | Wellbeing | Low | Yes | Yes | Yes | Yes | Yes | N/A (analysis of acute wellbeing effects of a single performance | Minimal | None | None | Low |
| Kreutz | 2014 | Mood (Positive feelings) | Low | Yes | Yes | No (unvalidated measure of positive & negative feelings) | Yes | Yes | N/A (experiment compares acute effects of 2 separate 30 minute sessions) | Serious (use of unvalidated measure) | None | None | Very Low |
| | | Mood (Negative feelings) | Low | Yes | Yes | No (unvalidated measure of positive & negative feelings) | Yes | Yes | N/A (experiment compares acute effects of 2 separate 30 minute sessions) | Serious (use of unvalidated measure) | None | None | Very Low |
| | | Cortisol | Low | Yes | Yes | Yes | Yes | Yes | N/A (experiment compares acute effects of 2 separate 30 minute sessions) | Minimal | None | None | Low |
| | | DHEA | Low | Yes | Yes | Yes | Yes | Yes | N/A (experiment compares acute effects of 2 separate 30 minute sessions) | Minimal | None | None | Low |
| | | Cortisol-DHEA ratio | Low | Yes | Yes | Yes | Yes | Yes | N/A (experiment compares acute effects of 2 separate 30 minute sessions) | Minimal | None | None | Low |
| | 2014 | Wellbeing (mental) | Low | Yes | Yes | Yes | Yes | Yes | Yes | Minimal | None | None | Low |

| | | | | | | | | | | | | |
|---|---|---|---|---|---|---|---|---|---|---|---|---|
| Perkins & Williamon | | Nutrition | Low | Yes | Yes | Yes | Yes | Yes | Yes | Minimal | None | None | Low |
| | | Interpersonal Relations | Low | Yes | Yes | Yes | Yes | Yes | Yes | Minimal | None | None | Low |
| | | Spiritual Growth | Low | Yes | Yes | Yes | Yes | Yes | Yes | Minimal | None | None | Low |
| | | Stress Management | Low | Yes | Yes | Yes | Yes | Yes | Yes | Minimal | None | None | Low |
| | | Health responsibility | Low | Yes | Yes | Yes | Yes | Yes | Yes | Minimal | None | None | Low |
| Valentine & Evans | 2001 | Blood pressure | Low | Yes | Yes | No (no robust comparisons with control group) | Yes | Yes | N/A (experiment conducted over 1 day) | Serious (uncontrolled study | None | None | Very Low |
| | | Mood (tense arousal) | Low | Yes | Yes | No (no robust comparisons with control group) | Yes | Yes | N/A (experiment conducted over 1 day) | Serious (uncontrolled study | None | None | Very Low |
| | | Mood (energetic arousal) | Low | Yes | Yes | No (no robust comparisons with control group) | Yes | Yes | N/A (experiment conducted over 1 day) | Serious (uncontrolled study | None | None | Very Low |
| | | Mood (hedonic tone) | Low | Yes | Yes | No (no robust comparisons with control group) | Yes | Yes | N/A (experiment conducted over 1 day) | Serious (uncontrolled study | None | None | Very Low |
| ***Eells, 2014*** | | | | | | | | | | | | |
| ***Fancourt, Ockelford & Belai, 2014*** | | | | | | | | | | | | |
| Koyama et al. | 2009 | Tension/anxiety | Low | Yes | N/A (no control group) | Yes | N/A (no control group) | Yes | N/A (single visit intervention study) | Serious (uncontrolled study) | None | None | Very Low |
| | | Depression/dejection | Low | Yes | N/A (no control group) | Yes | N/A (no control group) | Yes | N/A (single visit intervention study) | Serious (uncontrolled study) | None | None | Very Low |
| | | Anger/hostility | Low | Yes | N/A (no control group) | Yes | N/A (no control group) | Yes | N/A (single visit intervention study) | Serious (uncontrolled study) | None | None | Very Low |
| | | Vigor/activity | Low | Yes | N/A (no control group) | Yes | N/A (no control group) | Yes | N/A (single visit intervention study) | Serious (uncontrolled study) | None | None | Very Low |

| | | | | | | | | | | | |
|---|---|---|---|---|---|---|---|---|---|---|---|
| | Fatigue/inertia | Low | Yes | N/A (no control group) | Yes | N/A (no control group) | Yes | N/A (single visit intervention study) | Serious (uncontrolled study) | None | None | Very Low |
| | Confusion/ bewilderment | Low | Yes | N/A (no control group) | Yes | N/A (no control group) | Yes | N/A (single visit intervention study) | Serious (uncontrolled study) | None | None | Very Low |
| | Total mood disturbance | Low | Yes | N/A (no control group) | Yes | N/A (no control group) | Yes | N/A (single visit intervention study) | Serious (uncontrolled study) | None | None | Very Low |
| | Adrenaline | Low | Yes | N/A (no control group) | Yes | N/A (no control group) | Yes | N/A (single visit intervention study) | Serious (uncontrolled study) | None | None | Very Low |
| | Noradrenaline | Low | Yes | N/A (no control group) | Yes | N/A (no control group) | Yes | N/A (single visit intervention study) | Serious (uncontrolled study) | None | None | Very Low |
| | NK cell activity | Low | Yes | N/A (no control group) | Yes | N/A (no control group) | Yes | N/A (single visit intervention study) | Serious (uncontrolled study) | None | None | Very Low |
| | White blood cells | Low | Yes | N/A (no control group) | Yes | N/A (no control group) | Yes | N/A (single visit intervention study) | Serious (uncontrolled study) | None | None | Very Low |
| | Neutrophils | Low | Yes | N/A (no control group) | Yes | N/A (no control group) | Yes | N/A (single visit intervention study) | Serious (uncontrolled study) | None | None | Very Low |
| | Lymphocytes | Low | Yes | N/A (no control group) | Yes | N/A (no control group) | Yes | N/A (single visit intervention study) | Serious (uncontrolled study) | None | None | Very Low |
| | T cells | Low | Yes | N/A (no control group) | Yes | N/A (no control group) | Yes | N/A (single visit intervention study) | Serious (uncontrolled study) | None | None | Very Low |
| | B cells | Low | Yes | N/A (no control group) | Yes | N/A (no control group) | Yes | N/A (single visit intervention study) | Serious (uncontrolled study) | None | None | Very Low |

| | | | | | | | | | | | | |
|---|---|---|---|---|---|---|---|---|---|---|---|---|
| | CD4+ T cells | Low | Yes | N/A (no control group) | Yes | N/A (no control group) | Yes | N/A (single visit intervention study) | Serious (uncontrolled study) | None | None | Very Low |
| | CD8+ T cells | Low | Yes | N/A (no control group) | Yes | N/A (no control group) | Yes | N/A (single visit intervention study) | Serious (uncontrolled study) | None | None | Very Low |
| | CD4/CD8 ratio | Low | Yes | N/A (no control group) | Yes | N/A (no control group) | Yes | N/A (single visit intervention study) | Serious (uncontrolled study) | None | None | Very Low |
| | Naive T cells | Low | Yes | N/A (no control group) | Yes | N/A (no control group) | Yes | N/A (single visit intervention study) | Serious (uncontrolled study) | None | None | Very Low |
| | Memory T cells | Low | Yes | N/A (no control group) | Yes | N/A (no control group) | Yes | N/A (single visit intervention study) | Serious (uncontrolled study) | None | None | Very Low |
| | Naive/memory ratio | Low | Yes | N/A (no control group) | Yes | N/A (no control group) | Yes | N/A (single visit intervention study) | Serious (uncontrolled study) | None | None | Very Low |
| | NK cells (count | Low | Yes | N/A (no control group) | Yes | N/A (no control group) | Yes | N/A (single visit intervention study) | Serious (uncontrolled study) | None | None | Very Low |
| | IFN-alpha production | Low | Yes | N/A (no control group) | Yes | N/A (no control group) | Yes | N/A (single visit intervention study) | Serious (uncontrolled study) | None | None | Very Low |
| | IL-2 production | Low | Yes | N/A (no control group) | Yes | N/A (no control group) | Yes | N/A (single visit intervention study) | Serious (uncontrolled study) | None | None | Very Low |
| | IL-4 production | Low | Yes | N/A (no control group) | Yes | N/A (no control group) | Yes | N/A (single visit intervention study) | Serious (uncontrolled study) | None | None | Very Low |
| | IL-6 production | Low | Yes | N/A (no control group) | Yes | N/A (no control group) | Yes | N/A (single visit intervention study) | Serious (uncontrolled study) | None | None | Very Low |

| | | | | | | | | | | | | | |
|---|---|---|---|---|---|---|---|---|---|---|---|---|---|
| | | IL-10 production | Low | Yes | N/A (no control group) | Yes | N/A (no control group) | Yes | N/A (single visit intervention study) | Serious (uncontrolled study) | None | None | Very Low |
| | *Young cohort* ***above****; Old cohort* ***below*** | Tension/anxiety | Low | Yes | N/A (no control group) | Yes | N/A (no control group) | Yes | N/A (single visit intervention study) | Serious (uncontrolled study) | None | None | Very Low |
| | | Depression/dejection | Low | Yes | N/A (no control group) | Yes | N/A (no control group) | Yes | N/A (single visit intervention study) | Serious (uncontrolled study) | None | None | Very Low |
| | | Anger/hostility | Low | Yes | N/A (no control group) | Yes | N/A (no control group) | Yes | N/A (single visit intervention study) | Serious (uncontrolled study) | None | None | Very Low |
| | | Vigor/activity | Low | Yes | N/A (no control group) | Yes | N/A (no control group) | Yes | N/A (single visit intervention study) | Serious (uncontrolled study) | None | None | Very Low |
| | | Fatigue/inertia | Low | Yes | N/A (no control group) | Yes | N/A (no control group) | Yes | N/A (single visit intervention study) | Serious (uncontrolled study) | None | None | Very Low |
| | | Confusion/ bewilderment | Low | Yes | N/A (no control group) | Yes | N/A (no control group) | Yes | N/A (single visit intervention study) | Serious (uncontrolled study) | None | None | Very Low |
| | | Total mood disturbance | Low | Yes | N/A (no control group) | Yes | N/A (no control group) | Yes | N/A (single visit intervention study) | Serious (uncontrolled study) | None | None | Very Low |
| | | Adrenaline | Low | Yes | N/A (no control group) | Yes | N/A (no control group) | Yes | N/A (single visit intervention study) | Serious (uncontrolled study) | None | None | Very Low |
| | | Noradrenaline | Low | Yes | N/A (no control group) | Yes | N/A (no control group) | Yes | N/A (single visit intervention study) | Serious (uncontrolled study) | None | None | Very Low |

| | | | | | | | | | | | | |
|---|---|---|---|---|---|---|---|---|---|---|---|---|
| | NK cell activity | Low | Yes | N/A (no control group) | Yes | N/A (no control group) | Yes | N/A (single visit intervention study) | Serious (uncontrolled study) | None | None | Very Low |
| | White blood cells | Low | Yes | N/A (no control group) | Yes | N/A (no control group) | Yes | N/A (single visit intervention study) | Serious (uncontrolled study) | None | None | Very Low |
| | Neutrophils | Low | Yes | N/A (no control group) | Yes | N/A (no control group) | Yes | N/A (single visit intervention study) | Serious (uncontrolled study) | None | None | Very Low |
| | Lymphocytes | Low | Yes | N/A (no control group) | Yes | N/A (no control group) | Yes | N/A (single visit intervention study) | Serious (uncontrolled study) | None | None | Very Low |
| | T cells | Low | Yes | N/A (no control group) | Yes | N/A (no control group) | Yes | N/A (single visit intervention study) | Serious (uncontrolled study) | None | None | Very Low |
| | B cells | Low | Yes | N/A (no control group) | Yes | N/A (no control group) | Yes | N/A (single visit intervention study) | Serious (uncontrolled study) | None | None | Very Low |
| | CD4+ T cells | Low | Yes | N/A (no control group) | Yes | N/A (no control group) | Yes | N/A (single visit intervention study) | Serious (uncontrolled study) | None | None | Very Low |
| | CD8+ T cells | Low | Yes | N/A (no control group) | Yes | N/A (no control group) | Yes | N/A (single visit intervention study) | Serious (uncontrolled study) | None | None | Very Low |
| | CD4/CD8 ratio | Low | Yes | N/A (no control group) | Yes | N/A (no control group) | Yes | N/A (single visit intervention study) | Serious (uncontrolled study) | None | None | Very Low |
| | Naive T cells | Low | Yes | N/A (no control group) | Yes | N/A (no control group) | Yes | N/A (single visit intervention study) | Serious (uncontrolled study) | None | None | Very Low |
| | Memory T cells | Low | Yes | N/A (no control group) | Yes | N/A (no control group) | Yes | N/A (single visit intervention study) | Serious (uncontrolled study) | None | None | Very Low |

| | | | | | | | | | | | | | |
|---|---|---|---|---|---|---|---|---|---|---|---|---|---|
| | | Naive/memory ratio | Low | Yes | N/A (no control group) | Yes | N/A (no control group) | Yes | N/A (single visit intervention study) | Serious (uncontrolled study) | None | None | Very Low |
| | | NK cells (count | Low | Yes | N/A (no control group) | Yes | N/A (no control group) | Yes | N/A (single visit intervention study) | Serious (uncontrolled study) | None | None | Very Low |
| | | IFN-alpha production | Low | Yes | N/A (no control group) | Yes | N/A (no control group) | Yes | N/A (single visit intervention study) | Serious (uncontrolled study) | None | None | Very Low |
| | | IL-2 production | Low | Yes | N/A (no control group) | Yes | N/A (no control group) | Yes | N/A (single visit intervention study) | Serious (uncontrolled study) | None | None | Very Low |
| | | IL-4 production | Low | Yes | N/A (no control group) | Yes | N/A (no control group) | Yes | N/A (single visit intervention study) | Serious (uncontrolled study) | None | None | Very Low |
| | | IL-6 production | Low | Yes | N/A (no control group) | Yes | N/A (no control group) | Yes | N/A (single visit intervention study) | Serious (uncontrolled study) | None | None | Very Low |
| | | IL-10 production | Low | Yes | N/A (no control group) | Yes | N/A (no control group) | Yes | N/A (single visit intervention study) | Serious (uncontrolled study) | None | None | Very Low |
| Kreutz et al. | 2004 | Positive affect | Low | Yes | Yes | Yes | N/A (experimental intervention study conducted over 2 visits) | Yes | N/A (experimental intervention study conducted over 2 visits) | Minimal | None | None | Low |
| | | Negative affect | Low | Yes | Yes | Yes | N/A (experimental intervention study conducted over 2 visits) | Yes | N/A (experimental intervention study conducted over 2 visits) | Minimal | None | None | Low |
| | | Serum immunoglobulin A/albumin | Low | Yes | Yes | Yes | N/A (experimental intervention study conducted over 2 visits) | Yes | N/A (experimental intervention study conducted over 2 visits) | Minimal | None | None | Low |

| | | | | | | | | | | | | | |
|---|---|---|---|---|---|---|---|---|---|---|---|---|---|
| | | Cortisol | Low | Yes | Yes | Yes | N/A (experimental intervention study conducted over 2 visits) | Yes | N/A (experimental intervention study conducted over 2 visits) | Minimal | None | None | Low |
| Lindblad, Hogmark & Theorell | 2007 | Cortisol | Low | Yes | Yes | Yes | Yes | Yes | Yes | Minimal | None | None | Low |
| | | Self-esteem | Low | Yes | Yes | Yes | Yes | Yes | Yes | Minimal | None | None | Low |
| | | Social anxiety | Low | Yes | Yes | Yes | Yes | Yes | Yes | Minimal | None | None | Low |
| | | Social functioning | Low | Yes | Yes | Yes | Yes | Yes | Yes | Minimal | None | None | Low |
| ***Gick, 2012*** | | | | | | | | | | | | | |
| Pai et al | 2008 | Daytime somnolence | Low | Yes | Yes | Yes | N/A (cross sectional study) | Yes | N/A (cross sectional | Minimal | None | None | Low |
| Grape et al | 2003 | Cortisol | Low | Yes | Yes | No (no true control group; comparison of amateur & professional singers only) | N/A (cross sectional study) | Yes | N/A (cross sectional study) | Serious (no non-singing control group; impact of singing vs no singing unclear) | None | None | Very Low |
| | | Heart rate variability | Low | Yes | Yes | No (no true control group; comparison of amateur & professional singers only) | N/A (cross sectional study) | Yes | N/A (cross sectional study) | Serious (no non-singing control group; impact of singing vs no singing unclear) | None | None | Very Low |
| | | TNF-alpha | Low | Yes | Yes | No (no true control group; comparison of amateur & professional singers only) | N/A (cross sectional study) | Yes | N/A (cross sectional study) | Serious (no non-singing control group; impact of singing vs no singing unclear) | None | None | Very Low |
| | | Positive affect | Low | Yes | Yes | No (no true control group; comparison of amateur & professional singers only) | N/A (cross sectional study) | Yes | N/A (cross sectional study) | Serious (no non-singing control group; impact of singing vs no singing unclear) | None | None | Very Low |
| Beck et al | 2006 | Immunoglobulin A | Low | Yes | N/A (no control group) | No (self-collection of saliva samples by musicians (non expert collection)) | N/A (no control group) | Yes | N/A (series of acute pre/post analyses) | Very serious (single group study + self-collection of saliva samples by musicians) | None | None | Very Low |

| | | | | | | | | | | | | | |
|---|---|---|---|---|---|---|---|---|---|---|---|---|---|
| | | Cortisol | Low | Yes | N/A (no control group) | No (self-collection of saliva samples by musicians (non expert collection)) | N/A (no control group) | Yes | N/A (series of acute pre/post analyses) | Very serious (single group study + self-collection of saliva samples by musicians) | None | None | Very Low |
| ***Gordon, Fehd & McCandliss, 2015*** | | | | | | | | | | | | | |
| Bolduc & Lefebvre | 2012 | Phonological awareness (other) | Low | Yes | Yes | Yes | Yes | Yes | Yes | Minimal | None | None | Low |
| Gromko | 2005 | Reading fluency | Low | Yes | No (exposure & control groups sourced from two different schools with significant socioeconomic differences) | Yes | Yes | No (baseline differences in socioeconomic status not controlled for in analysis) | Yes | Serious (comparison of two different groups without statistical adjustment) | None | None | Very Low |
| | | Phonological awareness (other) | Low | Yes | No (exposure & control groups sourced from two different schools, with significant socioeconomic differences) | Yes | Yes | No (baseline differences in socioeconomic status not controlled for in analysis) | Yes | Serious (comparison of two different groups without statistical adjustment) | None | None | Very Low |
| Moritz et al | 2013 | Phonological awareness (rhyming) | Low | Yes | Yes | Yes | N/A (cross sectional study) | Yes | N/A (cross-sectional study) | Minimal | None | None | Low |
| | | Phonological awareness (other) | Low | Yes | Yes | Yes | N/A (cross sectional study) | Yes | N/A (cross-sectional study) | Minimal | None | None | Low |
| Myant, Armstrong & Healy | 2008 | Phonological awareness (rhyming) | Low | Article access unavailable - risk of bias extrapolated from review data | | | | | | Minimal | None | None | Low |
| | | Phonological awareness (other) | Low | Article access unavailable - risk of bias extrapolated from review data | | | | | | Minimal | None | None | Low |
| Register | 2004 | Reading fluency | Low | Yes | Yes | Yes | Yes | Yes | Yes | Minimal | None | None | Low |
| | | Phonological awareness (other) | Low | Yes | Yes | Yes | Yes | Yes | Yes | Minimal | None | None | Low |
| Thomson, Leong & Goswami | 2013 | Reading fluency | Low | Yes | Yes | Yes | Yes | Yes | Yes | Minimal | None | None | Low |
| | | Phonological awareness (rhyming) | Low | Yes | Yes | Yes | Yes | Yes | Yes | Minimal | None | None | Low |
| | | Phonological awareness (other) | Low | Yes | Yes | Yes | Yes | Yes | Yes | Minimal | None | None | Low |
| | 2009 | Phonological awareness (rhyming) | Low | Yes | Yes | Yes | Yes | Yes | Yes | Minimal | None | None | Low |

| | | | | | | | | | | | | |
|---|---|---|---|---|---|---|---|---|---|---|---|---|
| Yazejian & Peisner-Feinberg | | Phonological awareness (other) | Low | Yes | Yes | Yes | Yes | Yes | Yes | Minimal | None | None | Low |
| ***Hetland, 2000*** | | | | | | | | | | | | | |
| Bilhartz, Terry & Olson | 2000 | Spatial ability | Low | Yes | Yes | Yes | Yes | Yes | yes | Minimal | None | None | Low |
| Costa-Giomi | 1999 | Spatial reasoning | Low | Yes | Yes | Yes | Yes | Yes | No (78 analyzed out of 117 enrolled) | Serious (analysis of only 78 of 117 enrolled children) | None | None | Very Low |
| Graziano, Peterson & Shaw | 1999 | Spatial reasoning | Low | Yes | Yes | Yes | Yes | Yes | Yes | Minimal | None | None | Low |
| Gromko & Poorman | 1998 | Spatial reasoning, spatial ability | Low | Yes | Yes | Yes | Yes | Yes | Yes | Minimal | None | None | Low |
| Hurwitz et al | 1975 | Spatial reasoning, intelligence, spatial ability | Low | Yes | Yes | Yes | No (7 months surveillance for music group, but unclear surveillance of control group activities over same period) | Yes | N/A (cross sectional | Serious (unclear activities of control group over 7 months that music group received music training) | None | None | Very Low |
| Lazco | 1985 | Intelligence | Low | Yes | Yes | Yes | Yes | Yes | Yes | Minimal | None | None | Low |
| Mallory & Philbrick | 1995 | Spatial reasoning | Low | Yes | Yes | Yes | Yes | Yes | Yes | Minimal | None | None | Low |
| Parente & O'Malley | 1975 | Spatial ability | Low | Yes | Yes | Yes | Yes | Yes | Yes | Minimal | None | None | Low |
| Persellin | 1999 | Spatial reasoning | Low | Yes | Yes | Yes | Yes | Yes | Yes | Minimal | None | None | Low |
| Rauscher et al | 1994 | Spatial reasoning | Low | Yes | Yes | Yes | Yes | Yes | Yes | Minimal | None | None | Low |
| Rauscher et al | 1997 | Spatial reasoning, spatial ability | Low | Yes | Yes | Yes | Yes | Yes | Somewhat (34 of 111 withdrew from study, but evenly distributed amongst experimental groups) | Minimal | None | None | Low |
| Ruascher & Zupan | 1999 | Spatial reasoning | Low | Yes | Yes | Yes | Yes | Yes | Yes | Minimal | None | None | Low |

| | | | | | | | | | | | | | |
|---|---|---|---|---|---|---|---|---|---|---|---|---|---|
| Taetle | 1999 | Spatial reasoning, spatial ability | Low | Yes | Yes | Yes | Yes | Yes | Yes | Minimal | None | None | Low |
| Zulauf | 1993 | Intelligence, spatial ability | Low | Yes | Yes | Yes | Yes | Yes | Yes | Minimal | None | None | Low |
| ***Jaschke et al., 2013*** | | | | | | | | | | | | | |
| Gromko | 2005 | Reading ability | Low | Yes | No (exposure & control groups sourced from two different schools, with significant socioeconomic differences) | Yes | Yes | No (baseline differences in socioeconomic status not controlled for in analysis) | Yes | Serious (comparison of two different groups without statistical adjustment) | None | None | Very Low |
| Ho, Cheung and Chan | 2003 | IQ | Low | Yes | Yes | Yes | N/A (cross-sectional study) | Yes | N/A (cross-sectional) | Minimal | None | None | Low |
| Jentschke, Koelsch & Friederici | 2005 | Reading ability | Low | Yes | No (populations sourced from different public schools - music school vs. general public schools) | Yes | N/A (cross-sectional study) | No (no adjustment for any baseline differences between trained & untrained | N/A (cross-sectional) | Serious (no adjustment for potential confounding & trained/untrained cohorts sourced from two different locations | None | None | Very Low |
| Piro and Ortiz | 2009 | Reading ability | Low | Yes | No (exposure & control groups sourced from two different schools, with significant differences in reading scores & median income) | Yes | Yes | No (baseline differences in socioeconomic status not controlled for in analysis) | Yes | Serious (comparison of two different groups without statistical adjustment) | None | None | Very Low |
| Portowitz et al. | 2009 | IQ | Low | Yes | Yes (different centers of same institution with similar eligibility criteria) | Yes | Yes | Yes | Yes | Minimal | None | None | Low |
| Register | 2001 | Writing ability | Low | Yes | Yes | Yes | Yes | No identification or adjustment for any potential confounding variables | Not for all participants (11 of 61 lost to follow up) | Serious (no adjusting for confounding + analysis only of participants with >80% attendance) | None | None | Very Low |
| | | Reading ability | Low | Yes | Yes | Yes | Yes | No identification or adjustment for any potential | Not for all participants (11 of 61 lost to follow up) | Serious (no adjusting for confounding + analysis only of | None | None | Very Low |

| | | | | | | | | | | | | | |
|---|---|---|---|---|---|---|---|---|---|---|---|---|---|
| | | | | | | | | confounding variables | | participants with >80% attendance) | | | |
| | | Cognitive - auditory | Low | Yes | Yes | Yes | N/A (cross-sectional study) | No (between group socioeconomic differences noted but not controlled for) | N/A (cross-sectional) | Serious (comparison of two different groups without statistical adjustment) | None | None | Very Low |
| Tsang & Conrad | 2011 | Reading ability | Low | Yes | Yes | Yes | N/A (cross-sectional study) | No (between group socioeconomic differences noted but not controlled for) | N/A (cross-sectional) | Serious (comparison of two different groups without statistical adjustment) | None | None | Very Low |
| | | Reading ability | Low | Yes | Yes | Yes | N/A (cross-sectional study) | No (between group socioeconomic differences noted but not controlled for) | N/A (cross-sectional) | Serious (comparison of two different groups without statistical adjustment) | None | None | Very Low |
| ***Moore, 2013*** | | | | | | | | | | | | | |
| Bengtsson et al | 2007 | Emotional Regulation | Low | Yes | N/A (single group analysis) | Yes | N/A (single group study) | Yes | N/A (single cross-sectional assessment) | Minimal | Serious (emotional regulation measured through activation in proposed relevant brain regions) | None | Very Low |
| Berkowitz & Ansari | 2008 | Emotional regulation (Anterior Cingulate Cortex Activation) | Low | Yes | N/A (single group analysis) | Yes | N/A (single group study) | Yes | N/A (single cross-sectional assessment) | Minimal | Serious (as above) | None | Very Low |
| Brown, Martinez & Parsons | 2006 | Emotional regulation (Anterior Cingulate Cortex Activation) | Low | Yes | N/A (single group analysis) | Yes | N/A (single group study) | Yes | N/A (single cross-sectional assessment) | Minimal | Serious (as above) | None | Very Low |
| Callan et al | 2006 | Emotional regulation (Orbitofrontal cortex activation) | Low | Yes | N/A (single group analysis) | Yes | N/A (single group study) | Yes | N/A (single cross-sectional assessment) | Minimal | Serious (as above) | None | Very Low |

| | | | | | | | | | | | | | |
|---|---|---|---|---|---|---|---|---|---|---|---|---|---|
| de Manzano & Ullen | 2012 | Emotional regulation (Anterior cingulate cortex & dorsolateral prefrontal cortex activation) | Low | Yes | N/A (single group analysis) | Yes | N/A (single group study) | Yes | N/A (single cross-sectional assessment) | Minimal | Serious (as above) | None | Very Low |
| Jeffries, Fritz & Braun | 2003 | Emotional regulation (dorsolateral prefrontal cortex activation) | Low | Yes | N/A (single group analysis) | Yes | N/A (single group study) | Yes | N/A (single cross-sectional assessment) | Minimal | Serious (as above) | None | Very Low |
| Kleber et al | 2010 | Emotional regulation (anterior cingulate cortex, ventrolateral prefrontal cortex, amygdala activation) | Low | Yes | N/A (single group analysis) | Yes | N/A (single group study) | Yes | N/A (single cross-sectional assessment) | Minimal | Serious (as above) | None | Very Low |
| Kleber et al | 2007 | Emotional regulation (dorsolateral prefrontal cortex activation) | Low | Yes | N/A (single group analysis) | Yes | N/A (single group study) | Yes | N/A (single cross-sectional assessment) | Minimal | Serious (as above) | None | Very Low |
| Limb & Braun | 2008 | Emotional regulation (orbital prefrontal cortex, dorsolateral prefrontal cortex & amygdala activation) | Low | Yes | N/A (single group analysis) | Yes | N/A (single group study) | Yes | N/A (single cross-sectional assessment) | Minimal | Serious (as above) | None | Very Low |
| Perry et al. | 1999 | Emotional regulation (anterior cingulate cortex activation) | Low | Yes | N/A (single group analysis) | Yes | N/A (single group study) | Yes | N/A (single cross-sectional assessment) | Minimal | Serious (as above) | None | Very Low |
| Ruiz, Jabusch & Altenmueller | 2009 | Emotional regulation (anterior cingulate cortex activation) | Low | Yes | N/A (single group analysis) | Yes | N/A (single group study) | Yes | N/A (single cross-sectional assessment) | Minimal | Serious (as above) | None | Very Low |
| Vogt et al | 2007 | Emotional regulation (dorsolateral prefrontal cortex activation) | Low | Yes | N/A (single group analysis) | Yes | N/A (single group study) | Yes | N/A (single cross-sectional assessment) | Minimal | Serious (as above) | None | Very Low |
| Zarate & Zatorre | 2008 | Emotional regulation (anterior | Low | Yes | N/A (single group analysis) | Yes | N/A (single group study) | Yes | N/A (single cross-sectional assessment) | Minimal | Serious (as above) | None | Very Low |

| | | | | | | | | | | | | | |
|---|---|---|---|---|---|---|---|---|---|---|---|---|---|
| | | cingulate cortex activation) | | | | | | | | | | | |
| Zarate, Wood & Zatorre | 2010 | Emotional regulation (anterior cingulate cortex activation) | Low | Yes | N/A (single group analysis) | Yes | N/A (single group study) | Yes | N/A (single cross-sectional assessment) | Minimal | Serious (as above) | None | Very Low |
| ***Phillips & Becker, 2019*** | | | | | | | | | | | | | |
| Bormann et al. | 2006 | Stress | Low | Yes | N/A (single group study; no control group) | Yes | N/A (single group study) | Yes | Yes | Serious (single group uncontrolled study) | None | None | Very Low |
| | | Anxiety (state) | Low | Yes | N/A (single group study; no control group) | Yes | N/A (single group study) | Yes | Yes | Serious (single group uncontrolled study) | None | None | Very Low |
| | | Anxiety (trait) | Low | Yes | N/A (single group study; no control group) | Yes | N/A (single group study) | Yes | Yes | Serious (single group uncontrolled study) | None | None | Very Low |
| | | Anger (state) | Low | Yes | N/A (single group study; no control group) | Yes | N/A (single group study) | Yes | Yes | Serious (single group uncontrolled study) | None | None | Very Low |
| | | Anger (trait) | Low | Yes | N/A (single group study; no control group) | Yes | N/A (single group study) | Yes | Yes | Serious (single group uncontrolled study) | None | None | Very Low |
| | | Quality of life | Low | Yes | N/A (single group study; no control group) | Yes | N/A (single group study) | Yes | Yes | Serious (single group uncontrolled study) | None | None | Very Low |
| | | Spiritual Wellbeing | Low | Yes | N/A (single group study; no control group) | Yes | N/A (single group study) | Yes | Yes | Serious (single group uncontrolled study) | None | None | Very Low |
| Hilliard | 2006 | Compassion fatigue | Low | Yes | N/A (uncontrolled study with 2 experimental groups | Yes | N/A (uncontrolled study with 2 experimental groups | No (uncontrolled study with 2 experimental groups) | Yes | Serious (uncontrolled study) | None | None | Very Low |
| ***Raglio et al., 2019*** | | | | | | | | | | | | | |

| | | | | | | | | | | | | | |
|---|---|---|---|---|---|---|---|---|---|---|---|---|---|
| ***Skingley & Vella-Burrows, 2010*** | | | | | | | | | | | | | |
| ***Talmini et al, 2017*** | | | | | | | | | | | | | |
| Anaya, Pisoni & Kronenberger | 2016 | Short-term memory | Low | Yes | Yes | Yes | N/A (cross-sectional) | Yes | N/A (cross-sectional study) | Minimal | None | None | Low |
| Bialystok & DePape | 2009 | Short-term memory | Low | Yes | Yes | Yes | N/A (cross-sectional) | Yes | N/A (cross-sectional study) | Minimal | None | None | Low |
| | | Working memory | Low | Yes | Yes | Yes | N/A (cross-sectional) | Yes | N/A (cross-sectional study) | Minimal | None | None | Low |
| Boebinger et al. | 2015 | Short-term memory | Low | Yes | Yes | Yes | N/A (cross-sectional) | Yes | N/A (cross-sectional study) | Minimal | None | None | Low |
| | | Working memory | Low | Yes | Yes | Yes | N/A (cross-sectional) | Yes | N/A (cross-sectional study) | Minimal | None | None | Low |
| Brandler & Rammsayer | 2003 | Long-term memory | Low | Yes | Yes | Yes | N/A (cross-sectional) | Yes | N/A (cross-sectional study) | Minimal | None | None | Low |
| | | Long-term memory | Low | Yes | Yes | Yes | N/A (cross-sectional) | Yes | N/A (cross-sectional study) | Minimal | None | None | Low |
| Chan, Ho & Cheung | 1998 | Long-term memory | Low | Yes | Yes | Yes | N/A (cross-sectional) | Yes | N/A (cross-sectional study) | Minimal | None | None | Low |
| | | Long-term memory | Low | Yes | Yes | Yes | N/A (cross-sectional) | Yes | N/A (cross-sectional study) | Minimal | None | None | Low |
| Clayton et al. | 2016 | Working memory | Low | Yes | Unclear, but probably (groups mostly similarly aged in young 20s; musicians sourced from Boston University, likely nonmusicians as well) | Yes | N/A (cross-sectional) | Yes | N/A (cross-sectional study) | Minimal | None | None | Low |
| Franklin et al. | 2008 | Long-term memory | Low | Yes | Yes | Yes | N/A (cross-sectional) | Yes | N/A (cross-sectional study) | Minimal | None | None | Low |
| | | Working memory | Low | Yes | Yes | Yes | N/A (cross-sectional) | Yes | N/A (cross-sectional study) | Minimal | None | None | Low |
| George & Coch | 2011 | Short-term memory | Low | Yes | Yes | Yes | N/A (cross-sectional) | Yes | N/A (cross-sectional study) | Minimal | None | None | Low |
| | | Working memory | Low | Yes | Yes | Yes | N/A (cross-sectional) | Yes | N/A (cross-sectional study) | Minimal | None | None | Low |
| | | Short-term memory | Low | Yes | Yes | Yes | N/A (cross-sectional) | Yes | N/A (cross-sectional study) | Minimal | None | None | Low |
| | 2012 | Short-term memory | Low | Yes | Yes | Yes | N/A (cross-sectional) | Yes | N/A (cross-sectional study) | Minimal | None | None | Low |

| | | | | | | | | | | | | | |
|---|---|---|---|---|---|---|---|---|---|---|---|---|---|
| Hansen, Wallentin & Vuust | | Working memory | Low | Yes | Yes | Yes | N/A (cross-sectional) | Yes | N/A (cross-sectional study) | Minimal | None | None | Low |
| | | Short-term memory | Low | Yes | Yes | Yes | N/A (cross-sectional) | Yes | N/A (cross-sectional study) | Minimal | None | None | Low |
| | | Working memory | Low | Yes | Yes | Yes | N/A (cross-sectional) | Yes | N/A (cross-sectional study) | Minimal | None | None | Low |
| Helmbold, Rammsayer & Altenmueller | 2005 | Long-term memory | Low | Yes | Yes | Yes | N/A (cross-sectional) | Yes | N/A (cross-sectional study) | Minimal | None | None | Low |
| | | Long-term memory | Low | Yes | Yes | Yes | N/A (cross-sectional) | Yes | N/A (cross-sectional study) | Minimal | None | None | Low |
| Huang et al. | 2010 | Long-term memory | Low | Yes | Yes | Yes | N/A (cross-sectional) | Yes | N/A (cross-sectional study) | Minimal | None | None | Low |
| Jakobson et al. | 2008 | Long-term memory | Low | Yes | Yes | Yes | N/A (cross-sectional) | Yes | N/A (cross-sectional study) | Minimal | None | None | Low |
| | | Long-term memory | Low | Yes | Yes | Yes | N/A (cross-sectional) | Yes | N/A (cross-sectional study) | Minimal | None | None | Low |
| Lee, Lu & Ko | 2007 | Short-term memory | Low | Yes | Yes | Yes | N/A (cross-sectional) | Yes | N/A (cross-sectional study) | Minimal | None | None | Low |
| | | Working memory | Low | Yes | Yes | Yes | N/A (cross-sectional) | Yes | N/A (cross-sectional study) | Minimal | None | None | Low |
| | | Working memory | Low | Yes | Yes | Yes | N/A (cross-sectional) | Yes | N/A (cross-sectional study) | Minimal | None | None | Low |
| Monahan, Kendall & Carterette | 1987 | Short-term memory | Low | Yes | Yes | Yes | N/A (cross-sectional study) | Yes | N/A (cross-sectional study) | Minimal | None | None | Low |
| Okhrei, Kutsenko & Makarchuk | 2017 | Short-term memory | Low | Yes | Yes | Yes | N/A (cross-sectional study) | Yes | N/A (cross-sectional study) | Minimal | None | None | Low |
| | | Short-term memory | Low | Yes | Yes | Yes | N/A (cross-sectional study) | Yes | N/A (cross-sectional study) | Minimal | None | None | Low |
| Pallesen et al. | 2010 | Short-term memory | Low | Yes | Yes | Yes | N/A (cross-sectional study) | Yes | N/A (cross-sectional study) | Minimal | None | None | Low |
| | | Working memory | Low | Yes | Yes | Yes | N/A (cross-sectional study) | Yes | N/A (cross-sectional study) | Minimal | None | None | Low |
| Parbery-Clark et al. | 2011 | Working memory | Low | Yes | Yes | Yes | N/A (cross-sectional study) | Yes | N/A (cross-sectional study) | Minimal | None | None | Low |
| Ramachandra, Meighan & Gradzki | 2012 | Short-term memory | Low | Yes | Yes | Yes | N/A (cross-sectional study) | Yes | N/A (cross-sectional study) | Minimal | None | None | Low |
| | | Working memory | Low | Yes | Yes | Yes | N/A (cross-sectional study) | Yes | N/A (cross-sectional study) | Minimal | None | None | Low |
| Rodrigues, Loureiro & Caramelli | 2014 | Short-term memory | Low | Yes | Yes | Yes | N/A (cross-sectional study) | Yes | N/A (cross-sectional study) | Minimal | None | None | Low |
| Schiavio & Timmers | 2016 | Long-term memory | Low | Yes | Unclear | Yes | N/A (cross-sectional study) | Unclear | N/A (cross-sectional study) | Serious (population | None | None | Very Low |

| | | | | | | | | | | | | | |
|---|---|---|---|---|---|---|---|---|---|---|---|---|---|
| | | | | | | | | | | details and rigor of adjustment for confounding unclear | | | |
| Schulze, Dowling & Tillmann | 2012 | Short-term memory | Low | Yes | Yes | Yes | N/A (cross-sectional study) | Yes | N/A (cross-sectional study) | Minimal | None | None | Low |
| | | Working memory | Low | Yes | Yes | Yes | N/A (cross-sectional study) | Yes | N/A (cross-sectional study) | Minimal | None | None | Low |
| Schulze, Mueller & Koelsch | 2011 | Short-term memory | Low | Yes | Unclear, but probably both groups sourced from university cohorts | Yes | N/A (cross-sectional study) | Yes | N/A (cross-sectional study) | Minimal | None | None | Low |
| Schulze et al. | 2011 | Working memory | Low | Yes | Unclear, but probably both groups sourced from university cohorts | Yes | N/A (cross-sectional study) | Yes | N/A (cross-sectional study) | Minimal | None | None | Low |
| | | Working memory | Low | Yes | Unclear, but probably both groups sourced from university cohorts | Yes | N/A (cross-sectional study) | Yes | N/A (cross-sectional study) | Minimal | None | None | Low |
| Suarez, Elangovan & Au | 2015 | Working memory | Low | Yes | Yes | Yes | N/A (cross-sectional study) | Yes | N/A (cross-sectional study) | Minimal | None | None | Low |
| | | Short-term memory | Low | Yes | Yes | Yes | N/A (cross-sectional study) | Yes | N/A (cross-sectional study) | Minimal | None | None | Low |
| | | Short-term memory | Low | Yes | Yes | Yes | N/A (cross-sectional study) | Yes | N/A (cross-sectional study) | Minimal | None | None | Low |
| | | Long-term memory | Low | Yes | Yes | Yes | N/A (cross-sectional study) | Yes | N/A (cross-sectional study) | Minimal | None | None | Low |
| Talamini, Carretti, & Grassi | 2016 | Short-term memory | Low | Yes | Unclear, but probably both groups sourced from university cohorts | Yes | N/A (cross-sectional study) | Yes | N/A (cross-sectional study) | Minimal | None | None | Low |
| | | Working memory | Low | Yes | Unclear, but probably both groups sourced from university cohorts | Yes | N/A (cross-sectional study) | Yes | N/A (cross-sectional study) | Minimal | None | None | Low |
| Taylor & Dewhurst | 2017 | Long-term memory | Low | Yes | Yes | Yes | N/A (cross-sectional study) | Yes | N/A (cross-sectional study) | Minimal | None | None | Low |
| Vasuki et al. | 2016 | Short-term memory | Low | Yes | Yes | Yes | N/A (cross-sectional study) | Yes | N/A (cross-sectional study) | Minimal | None | None | Low |
| | | Working memory | Low | Yes | Yes | Yes | N/A (cross-sectional study) | Yes | N/A (cross-sectional study) | Minimal | None | None | Low |
| Weiss et al. | 2014 | Short-term memory | Low | Yes | Yes | Yes | N/A (cross-sectional study) | Yes | N/A (cross-sectional study) | Minimal | None | None | Low |

| | | | | | | | | | | | | | |
|---|---|---|---|---|---|---|---|---|---|---|---|---|---|
| Zuk et al | 2014 | Working memory | Low | Yes | Yes | Yes | N/A (cross-sectional study) | Yes | N/A (cross-sectional study) | Minimal | None | None | Low |
| ***Vaughn, 2000*** | | | | | | | | | | | | | |
| Annello | 1972 | Mathematics (correlation) | Low | Yes | Yes | Yes | N/A (cross sectional study) | Yes | N/A (cross sectional) | Minimal | None | None | Low |
| Catterall, Chapleau & Iwanaga | 1999 | Mathematics (correlation) | Low | Yes | Yes | Yes | N/A (cross sectional study) | Unclear (generally unclear description of statistical methods) | N/A (cross sectional) | Minimal | None | None | Low |
| Ciepluch | 1988 | Mathematics (correlation) | Low | Yes | N/A (single group study of correlations) | Yes | N/A (cross sectional study) | No (test of correlations between instrumental sightreading achievement & mathematical achievement - not a test of participation vs. math achievement; analysis confounding in the context of the rest of the meta-analysis) | N/A (cross sectional) | Serious (confounding analysis in context of rest of meta-analysis) | None | None | Very Low |
| College Board | 1988-1998 (not 1993) | Mathematics (correlation) | Low | Yes | Yes | Yes | N/A (cross sectional study) | Yes | N/A (cross sectional) | Minimal | None | '+1 massive sample size with consistent results across years | Moderate |
| Costa-Giomi | 1999 | Mathematics (causation - experimental) | Low | Yes | Yes | Yes | Yes | Yes | No (78 analyzed out of 117 enrolled) | Serious (analysis of only 78 of 117 enrolled children) | None | None | Very Low |
| Engdahl | 1994 | Mathematics (correlation) | Low | Yes | Yes | Yes | Yes | Yes | Yes | Minimal | None | None | Low |
| Friedman | 1959 | Mathematics (causation - experimental) | Low | Yes | Yes | Yes | Yes | Yes | Yes | Minimal | None | None | Low |

| | | | | | | | | | | | | |
|---|---|---|---|---|---|---|---|---|---|---|---|---|
| Graziano, Peterson & Shaw | 1999 | Mathematics (causation - experimental) | Low | Yes | Yes | Yes | Yes | Yes | Yes | Minimal | None | None | Low |
| Kvet | 1982 | Mathematics (correlation) | Low | Yes | Yes | Yes | Yes | Yes | Yes | Minimal | None | None | Low |
| McCarthy | 1992 | Mathematics (correlation) | Low | Yes | Yes | Yes | Yes | Yes | Yes | Minimal | None | None | Low |
| Neufeld | 1986 | Mathematics (causation - experimental) | Low | Yes | Yes | Yes | Yes | Yes | Yes | Minimal | None | None | Low |
| Weeden | 1971 | Mathematics (causation - experimental) | Low | Yes | Yes | Yes | Yes | Yes | Yes | Minimal | None | None | Low |
| Wheeler & Wheeler | 1951 | Mathematics (correlation) | Low | Yes | Yes | Yes | N/A (cross sectional study) | Yes | N/A (cross sectional) | Minimal | None | None | Low |
| ***Yap, Kwan & Ang, 2017*** | | | | | | | | | | | | | |
| Bittman | 2004 | Mood | Low | Yes | Yes | No (no direct statistical comparisons between pre-post effects in intervention vs control groups) | Yes | Yes | Yes | Serious (results not rigorously analyzed; single group study based on the way it's analyzed) | None | None | Very Low |
| | | Emotional exhaustion | Low | Yes | Yes | No (no direct statistical comparisons between pre-post effects in intervention vs control groups) | Yes | Yes | Yes | Serious (results not rigorously analyzed; single group study based on the way it's analyzed) | None | None | Very Low |
| | | Depersonalization | Low | Yes | Yes | No (no direct statistical comparisons between pre-post effects in intervention vs control groups) | Yes | Yes | Yes | Serious (results not rigorously analyzed; single group study based on the way it's analyzed) | None | None | Very Low |
| | | Personal accomplishment | Low | Yes | Yes | No (no direct statistical comparisons between pre-post effects in | Yes | Yes | Yes | Serious (results not rigorously analyzed; single group study based on the way it's analyzed) | None | None | Very Low |

| | | | | | | | | | | | | | |
|---|---|---|---|---|---|---|---|---|---|---|---|---|---|
| | | | | | | intervention vs control groups) | | | | | | | |
| Dunbar et al | 2012 | Positive affect | Low | Yes | No (by design) | Yes | Yes | Yes | N/A (experiment all conducted in 1 day) | Minimal | None | None | Low |
| | | Negative affect | Low | Yes | No (by design) | Yes | Yes | Yes | N/A (experiment all conducted in 1 day) | Minimal | None | None | Low |
| Ho et al | 2011 | Social functioning | Low | Yes | Yes | Yes | Yes | Yes | Yes | Minimal | None | None | Low |
| Mungas & Silverman | 2014 | Drowsiness | Low | Yes | No (significant differences in musical experience between groups) | Yes | Yes | No (substantial gender difference between groups not accounted for) | N/A (experiment conducted in single day) | Serious (no controlling for gender confound + significantly greater musical experience in intervention vs control group) | None | None | Very Low |
| | | Anxiety | Low | Yes | No (significant differences in musical experience between groups) | Yes | Yes | No (substantial gender difference between groups not accounted for) | N/A (experiment conducted in single day) | Serious (no controlling for gender confound + significantly greater musical experience in intervention vs control group) | None | None | Very Low |
| | | Depression | Low | Yes | No (significant differences in musical experience between groups) | Yes | Yes | No (substantial gender difference between groups not accounted for) | N/A (experiment conducted in single day) | Serious (no controlling for gender confound + significantly greater musical experience in intervention vs control group) | None | None | Very Low |
| | | Aggression | Low | Yes | No (significant differences in musical experience between groups) | Yes | Yes | No (substantial gender difference between groups not accounted for) | N/A (experiment conducted in single day) | Serious (no controlling for gender confound + significantly greater musical experience in intervention vs control group) | None | None | Very Low |

| | | | | | | | | | | | | |
|---|---|---|---|---|---|---|---|---|---|---|---|---|
| | | Confusion | Low | Yes | No (significant differences in musical experience between groups) | Yes | Yes | No (substantial gender difference between groups not accounted for) | N/A (experiment conducted in single day) | Serious (no controlling for gender confound + significantly greater musical experience in intervention vs control group) | None | None | Very Low |
| | | Lack of coordination | Low | Yes | No (significant differences in musical experience between groups) | Yes | Yes | No (substantial gender difference between groups not accounted for) | N/A (experiment conducted in single day) | Serious (no controlling for gender confound + significantly greater musical experience in intervention vs control group) | None | None | Very Low |
| Smith, Viljoen & McGeachie | 2014 | Blood pressure | Low | Yes | N/A (single group study) | Yes | N/A (single group study) | Yes (pre-post design) | N/A (experiment all conducted in 1 day) | Serious (uncontrolled study) | None | None | Very Low |
| | | Blood pressure | Low | Yes | N/A (single group study) | Yes | N/A (single group study) | Yes (pre-post design) | N/A (experiment all conducted in 1 day) | Serious (uncontrolled study) | None | None | Very Low |
| | | Stress/anxiety | Low | Yes | N/A (single group study) | Yes | N/A (single group study) | Yes (pre-post design) | N/A (experiment all conducted in 1 day) | Serious (uncontrolled study) | None | None | Very Low |
| | | Stress/anxiety | Low | Yes | N/A (single group study) | Yes | N/A (single group study) | Yes (pre-post design) | N/A (experiment all conducted in 1 day) | Serious (uncontrolled study) | None | None | Very Low |

*Table S13. Review articles excluded after full-text review, with reasons.*

| Author | Year | Title | Journal | Music/Dance | Review type | Reason for exclusion |
|---|---|---|---|---|---|---|
| Alain et al. | 2014 | Turning down the noise: the benefit of musical training on the aging auditory brain | Hearing Research | Music | Narrative | No reproducible search methodology / evidence of comprehensive synthesis |
| Angioi et al | 2009 | Fitness in Contemporary Dance: A Systematic Review | International Journal of Sports Medicine | Dance | Narrative | Focus on studies describing the physical fitness of dancers & evaluating the effect of supplementary exercise programs on dancers |
| Antoniadou, Michaelidis, Tsara | 2012 | Lung function in wind instrument players | Pneumon | Music | Narrative | No reproducible search methodology / evidence of comprehensive synthesis |
| Bashwiner & Bacon | 2019 | Musical creativity and the motor system | Current Opinion in Behavioral Sciences | Music | Narrative | No reproducible search methodology / evidence of comprehensive synthesis |
| Bonde, Juel, & Ekholm | 2018 | Associations between music and health-related outcomes in adult non-musicians, amateur musicians and professional musicians - Results from a nationwide Danish study | Nordic Journal of Music Therapy | Music | Narrative | No reproducible search methodology / evidence of comprehensive synthesis |
| Chobert & Besson | 2013 | Musical expertise and second language learning | Brain Sciences | Music | Narrative | No reproducible search methodology / evidence of comprehensive synthesis |
| Croom | 2015 | Music practice and participation for psychological well-being: A review of how music influences positive emotion, engagement, relationships, meaning, and accomplishment | Musicae Scientiae | Music | Narrative | No reproducible search methodology / evidence of comprehensive synthesis |
| Dawson | 2014 | Benefits of music training are widespread and lifelong: a bibliographic review of their non-musical effects | Medical Problems of Performing Artists | Music | Narrative | Focus not on comprehensively reviewing the benefits of music participation; largely non-reproducible search methodology |
| Dawson | 2011 | How and why musicians are different from nonmusicians: A bibliographic review | Medical Problems of Performing Artists | Music | Narrative | Focus not on comprehensively reviewing the benefits of music participation; largely non-reproducible search methodology |
| Ellis & Thayer | 2010 | Music and autonomic nervous system (DYS) function | Music Perception | Music | Narrative | No reproducible search methodology / evidence of comprehensive synthesis |
| Fernandez | 2018 | Music and brain development | Pediatric Annals | Music | Narrative | No reproducible search methodology / evidence of comprehensive synthesis |
| Herholz & Zatorre | 2012 | Musical Training as a Framework for Brain Plasticity: Behavior, Function, and Structure | Neuron | Music | Narrative | No reproducible search methodology / evidence of comprehensive synthesis |
| Huotilainen & Tervaniemi | 2018 | Planning music-based amelioration and training in infancy and childhood based on neural evidence | Annals of the New York Academy of Sciences | Music | Narrative | No reproducible search methodology / evidence of comprehensive synthesis |
| Jordan | 2019 | When I'm 64: A review of instrumental music-making and brain health in later life | Experimental Gerontology | Music | Narrative | No reproducible search methodology / evidence of comprehensive synthesis |
| Karpati et al | 2015 | Dance and the brain: a review | Annals of the New York Academy of Sciences | Dance | Narrative | No reproducible search methodology / evidence of comprehensive synthesis |

| | | | | | | |
|---|---|---|---|---|---|---|
| Kraus & Chandrasekaran | 2010 | Music training for the development of auditory skills | Nature reviews | Music | Narrative (Perspective) | No reproducible search methodology / evidence of comprehensive synthesis |
| Marks | 2016 | Narrative review of dance-based exercise and its specific impact on depressive symptoms in older adults | AIMS Medical Science | Dance | Narrative | Focus on dance as a clinical therapeutic |
| Miendlarzewska | 2014 | How musical training affects cognitive development: Rhythm, reward and other modulating variables | Frontiers in Neuroscience | Music | Narrative | No reproducible search methodology / evidence of comprehensive synthesis |
| Pantev & Herholz | 2011 | Plasticity of the human auditory cortex related to musical training | Neuroscience and Biobehavioral Reviews | Music | Narrative | No reproducible search methodology / evidence of comprehensive synthesis |
| Perrone-Capano, Volpicelli, di Porzio | 2017 | Biological bases of human musicality | Reviews in the Neurosciences | Music | Narrative | No reproducible search methodology / evidence of comprehensive synthesis |
| Perrot & Collet | 2014 | Function and plasticity of the medial olivocochlear system in musicians: A review | Hearing Research | Music | Narrative | No reproducible search methodology / evidence of comprehensive synthesis |
| Sarkamo | 2018 | Cognitive, emotional, and neural benefits of musical leisure activities in aging and neurological rehabilitation: A critical review | Annals of Physical and Rehabilitation Medicine | Music | Narrative | No reproducible search methodology / evidence of comprehensive synthesis |
| Schellenberg | 2001 | Music and nonmusical abilities | Annals of the New York Academy of Sciences | Music | Narrative | No reproducible search methodology / evidence of comprehensive synthesis |
| Standley | 1996 | A meta-analysis on the effects of music as reinforcement for education/therapy objectives | Journal of Research in Music Education | Music | Meta-analysis | Meta-analysis of contingent music (i.e. music lessons as a reward for achieving a certain outcome) |
| Strassel et al | 2011 | A systematic review of the evidence for the effectiveness of dance therapy | Alternative Therapies | Dance | Narrative | Focus on dance as a clinical therapeutic |
| Sutcliffe, Du, Ruffman | 2020 | Music Making and Neuropsychological Aging: A Review | Neuroscience and Biobehavioral Reviews | Music | Narrative | No reproducible search methodology / evidence of comprehensive synthesis |
| Twitchett, Koutedakis & Wyon | 2009 | Psychological fitness and professional classical ballet performance: A brief review | Journal of Strength and Conditioning Research | Dance | Narrative | No reproducible search methodology / evidence of comprehensive synthesis |
| Wan & Schlaug | 2010 | Music making as a tool for promoting brain plasticity across the life span | The Neuroscientist | Music | Narrative | No reproducible search methodology / evidence of comprehensive synthesis |
| Wolff | 1978 | Nonmusical Outcomes of Music Education: A Review of the literature | Bulletin of the Council for Research in Music Education | Music | Narrative | No reproducible search methodology / evidence of comprehensive synthesis |
| Zalatel, Gabrilio & Peric | 2013 | The training effects of dance aerobics: A review with an emphasis on the perspectives of investigations | Collegium Antropologicum | Dance | Narrative | No listing of included studies + unintelligible citations |
| Zimmerman & Lahav | 2012 | The multisensory brain and its ability to learn music | Annals of the New York Academy of Sciences | Music | Narrative | No reproducible search methodology / evidence of comprehensive synthesis |
| Zuk & Gaab | 2018 | Evaluating predisposition and training in shaping the musician's brain: The need for a developmental perspective | Annals of the New York Academy of Sciences | Music | Narrative | No reproducible search methodology / evidence of comprehensive synthesis |

*Table S14. Details of included studies evaluating the impact of performing arts participation on non-communicable disease risk.*

| Author | Year | Design | Subjects | Study Country | Music/ Dance | Type of participation | Definition of participation | Outcome | Effect of participation | Outcome details | GRADE certainty of evidence |
|---|---|---|---|---|---|---|---|---|---|---|---|
| Balbag, Pederson & Gatz | 2014 | Case-control | 157 pairs of Swedish twins at least 65 years old and discordant for dementia or cognitive impairment and with available data re. musical participation, mixed sex | Sweden | Music | Instrumental | Questionnaire regarding experience playing a musical instrument: played new and/or familiar music, frequency of playing, if still playing at time of assessment or stopped at a specific age; 'musicians' classified as those who played new and/or familiar music frequently and/or occasionally at the time of assessment OR if they stopped playing within 5 years of dementia onset | Dementia or cognitive impairment | Positive | Cognitive assessment at time of questionnaire + historical analysis of medical records | Low |
| Bygren, Konlaan, & Johansson | 1996 | Longitudinal cohort | 12,675 Swedish citizens aged 16-74 years; mixed sex | Sweden | Music | Instrumental or vocal | Survey (recall period unclear) – how often participants 'played music or sang in a choir' – dichotomized into 'now and then' or less frequently than 'now and then' | All cause mortality | No effect | 7-8 year follow up | Very Low |
| Chen et al. | 2013 | Cross-sectional | 15,514 individuals (6,952 men; 8.562 women) aged 50-70 years from the Dongfeng-Tongji Cohort (retirees of the Dongfeng Motor Corporation) in Shiyan, China; participants excluded if history of cancer, diabetes, CVD or stroke or any missing data | China | Dance | Unspecified | Interview re. frequency and average duration of a range of physical activities (undefined recall period); average METs from ACSM compendium of physical activity used to approximate dance energy expenditure (5 METs per hour), then multiplied by frequency and duration = total MET hours per week | Metabolic syndrome | Positive | Metabolic syndrome defined using updated National Cholesterol Education Program/Adult treatment Panel III criteria for Asian Americans | Very Low |
| Ekholm & Bonde | 2018 | Cross-sectional | 14,265 Danish adults (16 or older), mixed sex | Denmark | Music | Instrumental or vocal | Postal questionnaire: 'have you ever voluntarily sung in a choir/band or played a musical instrument in a band, orchestra or musical ensemble?'(yes, sing or play now; yes, previously; no); if yes, 'have you ever sung or played as a professional musician?' (same answer choices) | Asthma | No effect | | Very Low |
| | | | | | | | | Diabetes | No effect | | Very Low |
| | | | | | | | | Hypertension | No effect | | Very Low |
| | | | | | | | | Osteoarthritis | No effect | | Very Low |
| | | | | | | | | Rheumatoid Arthritis | No effect | | Very Low |
| | | | | | | | | Tinnitus | Negative | | Very Low |
| Hughes et al. | 2010 | Longitudinal cohort | 942 older adults in Pennsylvania, mixed sex | USA | Music | Instrumental | Survey – yes/no 'engagement in musical instruments as a hobby'; time spent in hobbies overall calculated, but not for individual hobbies | Dementia | No effect | Average 6 year follow up | Very Low |
| Konlaan, Bygren, Johansson | 2000 | Longitudinal cohort | 10,609 Swedish citizens aged 25-74, mixed sex | Sweden | Music | Instrumental or vocal | Survey (recall period unclear) – how often participants 'played music or sang in a choir' – dichotomized into 'now and then' or less frequently than 'now and then' | All-cause mortality | Positive | 14 year follow up | Very Low |

| | | | | | | | | | | | |
|---|---|---|---|---|---|---|---|---|---|---|---|
| Lin et al. | 2019 | Cross-sectional | 18,424 Taiwan Biobank participants – aged 30-70 with no history of cancer and unrelated to other biobank subjects, mixed sex | Taiwan | Dance | International standard | Survey of physical activity (undefined recall period); ‘regular participation’ defined as weekly dancing at least 30 min/3x weekly | Obesity metrics | Positive | Surveyed metrics: BMI, body fat %, waist circumference, hip circumference, waist-to-hip ratio | Very Low |
| Merom, Ding, Stamatakis | 2016 | Longitudinal cohort | 48,390 British/Scottish residents; Aged >/=40 and without baseline cardiovascular disease; n=3169 reported recreational dancing; mixed | UK | Dance | Unspecified | Health Survey for England + Scottish Health Survey (4 week recall); Interviewer-administered questionnaire re. dance participation & intensity (moderate intensity defined as ‘effort usually enough to make you out of breath or sweaty’) | Cardiovascular Disease Mortality | Mixed (positive – moderate intensity walking; no effect – light intensity dance) | 3-17 year follow up | Very Low |
| Vergehese et al. | 2003 | Longitudinal cohort | 469 community-dwellers without baseline dementia & aged 75-85 (Bronx Aging Study, mixed sex); participants also excluded with severe hearing or visual impairment, previous diagnosis of idiopathic Parkinson’s, liver disease, alcoholism or known terminal illness | USA | Dance & Music | Unspecified & instrumental | Interview (undefined recall period) re. frequency of 6 cognitive activities (including ‘playing musical instruments’) and 11 physical activities (including ‘dancing’); Participation frequency scored as: 7 (daily); 4 (several days per week); 1 (once weekly); 0 (monthly, occasionally, or never); summed scores from 6 cognitive activities = ‘cognitive activity score’ (out of 42); summed scores from 11 physical activities = ‘physical activity score’ (out of 77) | Dementia diagnosis | Positive | Median 5.1 year follow up | Low |

*Table S15. GRADE Certainty of Evidence ratings of each outcome measure of included studies evaluating the impact of performing arts participation on non-communicable disease risk.*

| **Author** | **Year** | **Outcome** | **Grade Initial Quality** | *Eligibility criteria developed and appropriately applied* | *Exposed/ unexposed sourced from a common population?* | *Appropriate & rigorous measurement of exposure & outcome* | *Identical surveillance for outcome in exposed & unexposed* | *Known confounders adequately measured, controlled & adjusted for* | *Follow up period complete?* | ***GRADE Bias risk*** | ***GRADE Indirectness*** | ***GRADE up rating*** | ***GRADE Certainty of Evidence*** |
|---|---|---|---|---|---|---|---|---|---|---|---|---|---|
| Balbag, Pederson & Gatz | 2014 | Dementia or cognitive impairment | Low | Yes | Yes | Yes | Yes | Yes | Yes | Minimal | None | None | Low |
| Bygren, Konlaan, & Johansson | 1996 | All cause mortality | Low | Yes | Yes | No (unvalidated/relatively insensitive measure of performing arts participation) | Yes | Yes | Yes | Serious (questionable methods & sensitivity of performing arts questionnaire) | None | None | Very Low |
| Chen et al. | 2013 | Metabolic syndrome | Low | Yes | Yes | No (undefined recall period + use of standardized conversions from ACSM for MET equivalents for all dance modes) | N/A (cross-sectional) | Yes | N/A (cross-sectional) | Serious (unvalidated measure + indirect conversion to METs) | None | None | Very Low |
| Ekholm & Bonde | 2018 | Asthma | Low | Yes | Yes | No (self-reported incidence of non-communicable diseases; insensitive measure of music participation | N/A (cross-sectional) | Yes | N/A (cross-sectional) | Serious (self-reported incidence of non-communicable diseases; insensitive measure of music participation) | None | None | Very Low |
| | | Diabetes | Low | Yes | Yes | No (self-reported incidence of non-communicable diseases; insensitive measure of music participation | N/A (cross-sectional) | Yes | N/A (cross-sectional) | Serious (self-reported incidence of non-communicable diseases; insensitive measure of music participation) | None | None | Very Low |
| | | Hypertension | Low | Yes | Yes | No (self-reported incidence of non-communicable diseases; insensitive measure of music participation | N/A (cross-sectional) | Yes | N/A (cross-sectional) | Serious (self-reported incidence of non-communicable diseases; insensitive measure of music participation) | None | None | Very Low |
| | | Osteoarthritis | Low | Yes | Yes | No (self-reported incidence of non-communicable diseases; insensitive measure of music participation | N/A (cross-sectional) | Yes | N/A (cross-sectional) | Serious (self-reported incidence of non-communicable diseases; | None | None | Very Low |

| | | | | | | | | | | | | |
|---|---|---|---|---|---|---|---|---|---|---|---|---|
| | | | | | | | | | insensitive measure of music participation) | | | |
| | | Rheumatoid Arthritis | Low | Yes | Yes | No (self-reported incidence of non-communicable diseases; insensitive measure of music participation | N/A (cross-sectional) | Yes | N/A (cross-sectional) | Serious (self-reported incidence of non-communicable diseases; insensitive measure of music participation) | None | None | Very Low |
| | | Tinnitus | low | Yes | Yes | No (self-reported incidence of non-communicable diseases; insensitive measure of music participation | N/A (cross-sectional) | Yes | N/A (cross-sectional) | Serious (self-reported incidence of non-communicable diseases; insensitive measure of music participation) | None | None | Very Low |
| Hughes et al. | 2010 | Dementia | Low | Yes | Yes | No (binary yes/no definition of playing musical instruments lacks sensitivity) | Yes | Yes | Yes | Serious (binary yes/no classification of playing a musical instrument lacks sensitivity | None | None | Very Low |
| Konlaan, Bygren, Johansson | 2000 | All-cause mortality | Low | Yes | Yes | No (unvalidated/relatively insensitive measure of performing arts participation) | Yes | Yes | Yes | Serious (questionable methods & sensitivity of performing arts questionnaire) | None | None | Very Low |
| Lin et al. | 2019 | Obesity metrics | Low | Yes | Yes | No (dance defined as 'exercise'; undefined recall period) | N/A (cross-sectional) | Yes | N/A (cross-sectional) | Serious (dance defined as 'exercise'; undefined recall period) | None | None | Very Low |
| Merom, Ding, Stamatakis | 2016 | Cardiovascular Disease Mortality | Low | Yes | Yes | No (unvalidated measure of dance participation) | Yes | Yes | Yes | Serious (unvalidated measure of dance participation) | None | None | Very Low |
| Vergehese et al. | 2003 | Dementia diagnosis | Low | Yes | Yes | Yes (unvalidated instrument, but good sensitivity in questions about frequency & analysis) | Yes | Yes | Yes | Minimal | None | None | Low |

*Table S16. Studies of non-communicable disease risk excluded after full-text review, with reasons.*

| Author | Year | Title | Journal | Reason for Exclusion |
|---|---|---|---|---|
| Bonde, Juel & Ekholm | 2018 | Associations between music and health-related outcomes in adult non-musicians, amateur musicians and professional musicians – Results from a nationwide Danish study | Nordic Journal of Music Therapy | No data on the impact of music participation on noncommunicable diseases or mortality; included as review article |
| Chomistek et al | 2012 | Vigorous-intensity leisure-time physical activity and risk of major chronic disease in men | Medicine and Science in Sports and Exercise | No examination of performing arts participation |
| Cuypers et al. | 2011 | Patterns of receptive and creative cultural activities and their association with perceived health, anxiety, depression and satisfaction with life among adults in the HUNT study, Norway | Journal of Epidemiology and Community Health | No examination of noncommunicable disease risk |
| Danneberg et al | 1989 | Leisure time physical activity in the Framingham Offspring study. Description, seasonal variation, and risk factor correlates | American Journal of Epidemiology | Excluded, no individual analysis of PA modes |
| Darweesh et al | 2018 | Professional occupation and the risk of Parkinson's disease | European Journal of Neurology | All arts occupations (visual & performing) grouped together |
| Davies, Knuiman & Rosenberg | 2016 | The art of being mentally healthy: A study to quantify the relationship between recreational arts engagement and Mental well-being in the general population | BMC Public Health | Relationships only examined between all arts engagement (participation & attendance at a range of art forms) and mental well-being; no specific analyses of music/dance participation |
| Ekholm, Bonde & Juel | 2016 | Music and Public Health – An empirical study of the use of music in the daily life of the adult Danish citizens and the health implications of musical participation | Arts & Health | No data on the impact of music participation on noncommunicable diseases or mortality |
| Ekholm, Juel & Bonde | 2016 | Associations between daily musicking and health: Results from a nationwide survey in Denmark | Scandinavian Journal of Public Health | No data on the impact of music participation on noncommunicable diseases or mortality |
| Hyyppa & Maki | 2001 | Individual-level relationships between social capital and self-rated health in a bilingual community | Preventive Medicine | Correlations only reported between singing in a choir and self-rated health |
| Hyyppa et al | 2007 | Individual-level measures of social capital as predictors of all-cause and cardiovascular mortality: A population-based prospective study of men and women in Finland | European Journal of Epidemiology | Music participation only analyzed as a group, not stand alone, variable |
| Lee et al. | 2010 | Comparisons of leisure-time physical activity and cardiorespiratory fitness as predictors of all-cause mortality in men and women | British Journal of Sports Medicine | No independent examination of performing arts participation – only analyzed as grouped 'physical activity' variable |
| Merom et al. | 2014 | Swimming and other sporting activities and the rate of falls in older men: Longitudinal findings from the concord health and ageing in men project | American Journal of Epidemiology | No independent examination of performing arts participation – only analyzed as grouped 'moderate activity' variable |
| Moisan, Meyer, Gingras | 1991 | Leisure physical activity and age at menarche | Medicine and Science in Sports and Exercise | Dancing only presented as grouped variable with gymnastics, figure skating, synchronized swimming, diving |
| Peeters et al. | 2010 | Which types of activities are associated with risk of recurrent falling in older persons | The Journals of Gerontology | No examination of performing arts participation |
| Tanasescu et al. | 2002 | Exercise type and intensity in relation to coronary heart disease in men | JAMA | Performing arts participation not analyzed |
| Vaisto et al | 2014 | Physical activity and sedentary behavior in relation to cardiometabolic risk in children: Cross-sectional findings from the Physical Activity and Nutrition in Children (PANIC) Study | International Journal of Behavioral Nutrition and Physical Activity | Examination of general cardiometabolic factors, not noncommunicable disease risk (i.e. metabolic syndrome) |

*Table S17. Details of all included mean heart rate data from included studies of <u>dance</u> participation.*

| | Dance style | Type of performance | Mean Heart Rate (% max) | Authors | Year |
|---|---|---|---|---|---|
| Very Light (<57% max) | Active Video Game Dance (DDR) | Laboratory (30 min) | 50.5% | Sell, Lillie & Taylor | 2008 |
| | Active Video Game Dancing (Dance Central) | Laboratory (unspecified duration; 3 songs | 52.4% | Bronner et al. | 2016 |
| | Active Video Game Dancing (DDR) | Laboratory (30 min) | 54.5% | Rauber et al. | 2018 |
| | Active Video Game Dance (DDR) | Laboratory (15 min - level 1) | 54.6% | Graf et al. | 2009 |
| | Active Video Game Dancing (Dance Central) | Laboratory (unspecified duration; 3 songs | 54.8% | Bronner et al. | 2016 |
| | Modern Dance | Rehearsal (unspecified duration) | 56.4% | Wyon et al. | 2004 |
| | Active Video Game Dance (JustDance) | Laboratory (3 songs - unspecified duration) | 56.2 - 57.8% | Jin | 2015 |
| Light (57-63% max heart rate) | Active Video Game Dance (Kinect Dance Central) | Laboratory (13 min) | 57.1% | Bronner, Pinsker & Noah | 2013 |
| | Active Video Game Dance (JustDance) | Laboratory (10 min) | 57.7% | Marks, Rispen & Calara | 2015 |
| | Active Video Game Dance (JustDance) | Laboratory (3 songs - unspecified duration) | 56.2 - 57.8% | Jin | 2015 |
| | Modern | Class (90 min) | 58.1% | Wyon et al. | 2002 |
| | Active Video Game Dancing (Dance Central) | Laboratory (unspecified duration; 3 songs | 58.2% | Bronner et al. | 2016 |
| | Modern Dance | Rehearsal (unspecified duration) | 58.5% | Wyon et al. | 2004 |
| | Waltz | Class (5 min) | 59.0% | Nelson et al. | 2011 |
| | Fox trot | Class (5 min) | 59.2% | Nelson et al. | 2011 |
| | Ballet | Class (90 min) | 59.4% | Sanders et al. | 2019 |
| | Rhumba | Class (4 min) | 59.5% | Nelson et al. | 2011 |
| | Active Video Game Dance (using DDR-style mat) | Laboratory (10 min - level 1) | 59.5% | Fawkner et al | 2010 |
| | Salsa | Class (60 min) | 59.6% | Emerenziani et al | 2013 |
| | Active Video Game Dancing (Dance Central) | Laboratory (15 min) | 59.7% | Smallwood et al. | 2012 |
| | Tango | Class (5 min) | 59.8% | Nelson et al. | 2011 |
| | Active Video Game Dance (Wii Just Dance 3) | Laboratory (21 min) | 60.3% | Bronner, Pinsker & Noah | 2013 |
| | Active Video Game Dance | Laboratory (4 min) | 60.6% | Oh et al. | 2017 |
| | Merengue | Class (4 min | 60.9% | Nelson et al. | 2011 |
| | Modern Dance | Class (unspecified duration) | 61.1% | Wyon et al. | 2004 |
| | Active Video Game Dancing (Dance Central) | Laboratory (unspecified duration; 3 songs | 61.2% | Bronner et al. | 2016 |
| | Ballet | Class (24 minutes - barre exercises) | 61.4% | Cohen et al | 1982 |
| | Modern | Class (90 min) | 61.6% | Wyon et al. | 2002 |
| | Modern Dance | Class (unspecified duration) | 61.6% | Wyon et al. | 2004 |
| | Mixed ('Dancing Classrooms') | Class (50 - 80 min) | 61.6% | Pennington & Nelson | 2020 |
| | Active Video Game Dance (DDR) | Laboratory (15 min - level 2) | 61.6% | Graf et al. | 2009 |
| | Active Video Game Dance (DDR) | Laboratory (30 min) | 62.1% | Kraft et al. | 2011 |
| | Modern Dance | Class (90 min) | 62.4% | Sanders et al. | 2019 |

| | | | | | |
|---|---|---|---|---|---|
| | Active Video Game Dance (using DDR-style mat) | Laboratory (10 min - level 2) | 63.1% | Fawkner et al | 2010 |
| **Moderate (64-76% max heart rate)** | Fun Dance | Class (6 min) | 64.0% | Nelson et al. | 2011 |
| | Active Video Game Dance (DDR) | Laboratory (12 min) | 64.1% | Unnithan, Houser & Fernhall | 2006 |
| | Mixture - 'specially designed to develop functional and fitness elements that tend to naturally deteriorate with the aging process' | Class (60 minute) | 64.7% | Rodrigues-Krause et al. | 2018 |
| | Salsa | Class (60 min - 'rueda de casino' style) | 64.7% | Emerenziani et al | 2013 |
| | Salsa | Nightclub (60 min) | 65.3% | Emerenziani et al | 2013 |
| | Salsa | Nightclub (60 min) | 65.7% | Emerenziani et al | 2013 |
| | Line dancing | Class (~90 min) | 65.8% | Zan, Hannon & Carson | 2009 |
| | Hip-hop - 'New style' | Laboratory (unspecified duration - routine 1) | 65.8% | Wyon et al. | 2018 |
| | Maori haka | Laboratory (5 min) | 66.6% | Zhu et al | 2018 |
| | Active Video Game Dance (Dance game that comes with Xbox 360) | Laboratory (8 min) | 66.6% | Canabrava et al. | 2018 |
| | Tongan | Laboratory (5 min) | 66.7% | Zhu et al | 2018 |
| | Salsa | Class (60 min) | 67.4% | Emerenziani et al | 2013 |
| | Active Video Game Dance (Wii DDR) | Laboratory (9 min) | 67.6% | Bronner, Pinsker & Noah | 2013 |
| | Latin | Laboratory (15 min) | 67.9% | Domene & Easton | 2014 |
| | Active Video Game Dance (using DDR-style mat) | Laboratory (10 min - level 3) | 68.0% | Fawkner et al | 2010 |
| | Zumba (video instructor) | Laboratory (60 min) | 68.0% | Schneekloth & Brown | 2018 |
| | Active Video Game Dance (JustDance - Xbox Kinect) | Laboratory (60 min) | 68.2% | Eason et al. | 2016 |
| | Maori poi balls | Laboratory (5 min) | 68.3% | Zhu et al | 2018 |
| | Modern Dance | Dress rehearsal (unspecified duration) | 68.9% | Wyon et al. | 2004 |
| | Disco | Party (6 min) | 69.4% | Leger | 1982 |
| | Modern | Class (90 min) | 69.4% | Wyon et al. | 2002 |
| | Modern Dance | Dress rehearsal (unspecified duration) | 69.9% | Wyon et al. | 2004 |
| | Disco | Party (6 min) | 70.3% | Leger | 1982 |
| | Active Video Game Dance (DDR) | Laboratory (10 min - maximum possible difficulty for each individual) | 70.3% | Tan et al. | 2002 |
| | Ballet | Class (24 minutes - barre exercises) | 70.4% | Cohen et al | 1982 |
| | Active Video Game Dance (JustDance) | Laboratory (15 min - Playstation) | 70.4% | Powers et al. | 2016 |
| | Active Video Game Dance (JustDance) | Laboratory (15 min - Wii) | 70.4% | Powers et al. | 2016 |
| | Aerobic Dance | Laboratory (3 min - level 1 | 70.8% | Thomsen & Ballor | 1991 |
| | Active Video Game Dance | Laboratory (20 min) | 71.1% | Cakir-Atabek, Aygun & Dokumaci | 2020 |
| | Ballet | Performance (unspecified duration) | 71.2% | Seliger et al | 1970 |
| | Active Video Game Dance (Zumba Fitness Rush) | Laboratory (50 min - multiplayer mode) | 71.2% | Giancotti et al | 2018 |

| | | | | | |
|---|---|---|---|---|---|
| | Pole Dancing | Class (min) | 71.3% | Nicholas et al. | 2019 |
| | Swing | Class (5 min) | 71.6% | Nelson et al. | 2011 |
| | Aerobic Dance | Laboratory (3 min - level 1 | 71.7% | Thomsen & Ballor | 1991 |
| | Active Video Game Dance (Dance Central 3) | Laboratory (15 min | 71.7% | Aygun, Dokumaci & Cakir | 2018 |
| | Ballet | Class (7 minutes - center floor exercises) | 72.0% | Cohen et al | 1982 |
| | Hawaiian Hula | Laboratory (5 min) | 72.3% | Zhu et al | 2018 |
| | Unspecified | Dance sequence (4 min - timepoint 3) | 72.5% | Beck, Wyon & Redding | 2018 |
| | Salsa | Class (60 min - 'rueda de casino' style) | 72.6% | Emerenziani et al | 2013 |
| | Active Video Game Dance (JustDance) | Laboratory (15 min - Xbox Kinect) | 72.6% | Powers et al. | 2016 |
| | Aerobic dance | Laboratory (15 min - low impact) | 72.8% | Rousanoglou & Boudolos | 2005 |
| | Active Video Game Dance (Kinect Just Dance 3) | Laboratory (21 min) | 73.0% | Bronner, Pinsker & Noah | 2013 |
| | Dance Fitness Class | Laboratory (60 min) | 73.1% | Eason et al. | 2016 |
| | Fijian | Laboratory (5 min) | 73.7% | Zhu et al | 2018 |
| | Active Video Game Dance (Zumba Fitness Rush) | Laboratory (50 min - single player mode) | 73.9% | Giancotti et al | 2018 |
| | Unspecified | Dance sequence (4 min - timepoint 2) | 74.0% | Beck, Wyon & Redding | 2018 |
| | Unspecified | Dance sequence (4 min - timepoint 3) | 74.3% | Beck, Wyon & Redding | 2018 |
| | Zumba - DVD | Home workout (60 min) | 74.5% | Delextrat & Neupert | 2016 |
| | Ballet | Class (30 minute) | 75.6% | Rodrigues-Krause et al. | 2014 |
| | Zumba | Class (60 min) | 75.9% | Strejcova et al | 2013 |
| | Aerobic bench stepping | Class (60 minute) | 76.2% | Rixon, Rehor & Bemben | 2006 |
| | Samoan sasa | Laboratory (5 min) | 76.3% | Zhu et al | 2018 |
| | Ballet - Plies | Laboratory (3 min) | 76.3% | Rodrigues-Krause | 2014 |
| | Unspecified | Dance sequence (4 min - timepoint 1) | 76.4% | Beck, Wyon & Redding | 2018 |
| **Vigorous (77-95% max heart rate)** | Ballet - Tendus | Laboratory (2 min) | 76.7% | Rodrigues-Krause | 2014 |
| | Highland Dance | Class (77 min) | 77.1% | Bailie, Wyon & Head | 2007 |
| | Zumba - class | Class (60 min) | 77.7% | Delextrat & Neupert | 2016 |
| | Unspecified | Dance sequence (4 min - timepoint 2) | 78.2% | Beck, Wyon & Redding | 2018 |
| | Ballet - Grand Battement | Laboratory (1 min) | 78.3% | Rodrigues-Krause | 2014 |
| | Musical Theater (dance + singing) | Laboratory (4 min) | 78.5% | Stephens & Wyon | 2020 |
| | Aerobic Dance | Laboratory (3 min - level 1 | 78.8% | Thomsen & Ballor | 1991 |
| | Hula (low intensity) | Laboratory (12-16 min) | 78.8% | Usgawa et al | 2014 |
| | Samoan slap | Laboratory (5 min) | 78.8% | Zhu et al | 2018 |
| | Step Aerobic Dance | Class (20 min - aerobic portion) | 79.0% | Forte et al. | 2001 |
| | Unspecified | Dance sequence (4 min - timepoint 1) | 79.1% | Beck, Wyon & Redding | 2018 |

| | | | | |
|---|---|---|---|---|
| Aerobic Dance | Laboratory (3 min - level 2) | 79.5% | Thomsen & Ballor | 1991 |
| Musical Theater (dance only) | Laboratory (4 min) | 80.0% | Stephens & Wyon | 2020 |
| Sardinian folk dance (ballu sardu) | Laboratory (14 min) | 80.2% | Cugusi et al. | 2015 |
| Aerobic dance | Class (18 min) | 80.3% | Grant et al | 2002 |
| Ballet | Class (7 minutes - center floor exercises) | 80.4% | Cohen et al | 1982 |
| Ballet - Rond de Jambes | Laboratory (3 min) | 80.8% | Rodrigues-Krause | 2014 |
| Aerobic dancing | Laboratory (10 min) | 80.8% | Carroll, Otto & Wygand | 1991 |
| Zumba (human instructor) | Class (60 min) | 81.0% | Schneekloth & Brown | 2018 |
| Sports Dancing | Laboratory (10 min) | 81.1% | Uspuriene & Cepulenas | 2012 |
| Aerobic dance - march | Laboratory (6 min) | 81.2% | Darby, Browder & Reeves | 1995 |
| Hip-hop - 'New style' | Laboratory (unspecified duration - routine 2) | 81.3% | Wyon et al. | 2018 |
| Aerobic Dance | Class (30 min) | 81.5% | Sekulic, Rausavljevic & Zenic | 2006 |
| Aerobic dance (low impact, vigorous arms) | Laboratory (3 min) | 81.6% | Schaeffer-Gerschutz, Darby & Browder | 2000 |
| Aerobic dance (low impact, no arm involvement) | Laboratory (3 min) | 81.6% | Schaeffer-Gerschutz, Darby & Browder | 2000 |
| Active Video Game Dance (StepMania) | Laboratory (9 min) | 81.8% | Bronner, Pinsker & Noah | 2013 |
| Ballet | Performance (unspecified duration) | 82.1% | Seliger et al | 1970 |
| Tahitian | Laboratory (5 min) | 82.2% | Zhu et al | 2018 |
| Aerobic dancing | Laboratory (10 min - 6 in bench) | 82.4% | Grier et al. | 2002 |
| Active Video Game Dance (Dance Central 3) | Laboratory (15 min) | 82.6% | Aygun, Dokumaci & Cakir | 2018 |
| Aerobic dance | Laboratory (15 min - high impact) | 82.6% | Rousanoglou & Boudolos | 2005 |
| Sports Dancing | Laboratory (10 min) | 82.7% | Uspuriene & Cepulenas | 2012 |
| Ballet - Jetes | Laboratory (2 min) | 83.4% | Rodrigues-Krause | 2014 |
| Active Video Game Dance (DDR) | Laboratory (30 min) | 83.4% | Sell, Lillie & Taylor | 2008 |
| Aerobic Dance | Class (30 min) | 84.1% | Sekulic, Rausavljevic & Zenic | 2006 |
| Aerobic Dance | Laboratory (3 min - level 2) | 84.2% | Thomsen & Ballor | 1991 |
| Aerobic Dance | Class (40 min) | 84.3-85.9% | Rockefeller & Burke | 1979 |
| Aerobic bench stepping | Laboratory (20 min) | 84.7% | Lowe et al | 2010 |
| Aerobic Dance | Class (55 min) | 84.7% | De Angelis et al. | 1998 |
| Step Aerobic Dance | Class (30 min - central portion (no warm up / cool down)) | 85.4% | La Torre et al. | 2005 |
| Ballet - Fondus | Laboratory (3 min) | 85.9% | Rodrigues-Krause | 2014 |
| Aerobic dance - jumping jack | Laboratory (6 min) | 85.9% | Darby, Browder & Reeves | 1995 |
| Aerobic (high & low impact) | Laboratory (25 min - 5 min warmup, 15 min session, 5 min cool down) | 86.1% | Thompson et al. | 1991 |
| Aerobic dance - jog | Laboratory (6 min) | 86.4% | Darby, Browder & Reeves | 1995 |
| Aerobic Dance | Laboratory (3 min - level 3) | 86.5% | Thomsen & Ballor | 1991 |

| | | | | |
|---|---|---|---|---|
| Aerobic dance (low impact) | Class (40 min) | 86.5% | Tarrant & McNaughton | 1997 |
| Aerobic dance (low impact) | Laboratory (9 min) | 86.7% | Bronstein et al | 1990 |
| Ballet - Temps Leves | Laboratory (1 min) | 87.0% | Rodrigues-Krause | 2014 |
| Musical Theater (dance only) | Laboratory (4 min) | 87.0% | Stephens & Wyon | 2020 |
| Polish folk dancing | Laboratory (8 min) | 87.6% | Maciejczyk & Fec | 2013 |
| Highland Dance | Rehearsal (unspecified duration) | 87.6% | Bailie, Wyon & Head | 2007 |
| Aerobic dancing | Laboratory (10 min - 8 in bench) | 87.7% | Grier et al. | 2002 |
| Aerobic dance - power jack | Laboratory (6 min) | 88.0% | Darby, Browder & Reeves | 1995 |
| Ballroom - 'Latin sequence' | Laboratory (unspecified duration - 'competition simulation') | 88.1% | Blanksby & Reidy | 1988 |
| Aerobic Dance | Laboratory (24 min) | 88.2% | Schaeffer et al. | 1995 |
| Tap dance | Laboratory (36 min) | 88.5% | Oliveira et al. | 2010 |
| Aerobic Dance | Laboratory (3 min - level 3) | 88.6% | Thomsen & Ballor | 1991 |
| Ballet - Grant Adage | Laboratory (2 min) | 88.7% | Rodrigues-Krause | 2014 |
| Ballroom - 'Modern sequence' | Laboratory (unspecified duration - 'competition simulation') | 89.1% | Blanksby & Reidy | 1988 |
| Musical Theater (dance + singing) | Laboratory (4 min) | 89.1% | Stephens & Wyon | 2020 |
| Aerobic dance (high impact, vigorous arms) | Laboratory (3 min) | 90.0% | Schaeffer-Gerschutz, Darby & Browder | 2000 |
| Ballroom - 'Modern sequence' | Laboratory (unspecified duration - 'competition simulation') | 90.2% | Blanksby & Reidy | 1988 |
| Ballet | Rehearsal (30 minute) | 90.6% | Rodrigues-Krause et al. | 2014 |
| Sports Dancing (standard) | Laboratory (10 min) | 90.6% | Liiv et al. | 2014 |
| Sport Dancing (standard) | Laboratory (10 min) | 90.8% | Bria et al. | 2011 |
| Aerobic Dance | Laboratory (3 min - level 2) | 91.3% | Thomsen & Ballor | 1991 |
| Swedish folk dance (hambo) | Laboratory (4 min) | 91.4% | Wigaeus & Kilbom | 1980 |
| Hula (high intensity) | Laboratory (12-16 min) | 91.5% | Usgawa et al | 2014 |
| Sports Dancing (standard) | Laboratory (10 min) | 91.8% | Liiv et al. | 2014 |
| Sports Dancing (Ten Dance) | Laboratory (10 min) | 91.9% | Liiv et al. | 2014 |
| Tinikling (traditional Filipino dance) | Laboratory (5 min) | 92.2% | Heil et al. | 2019 |
| Ballroom - 'Latin sequence' | Laboratory (unspecified duration - 'competition simulation') | 92.3% | Blanksby & Reidy | 1988 |
| Sports Dancing (Ten Dance) | Laboratory (10 min) | 92.5% | Liiv et al. | 2014 |
| Sport Dancing (standard) | Laboratory (10 min) | 92.6% | Bria et al. | 2011 |
| Hip-hop - Breakdancing | Laboratory (unspecified duration - set 2) | 92.7% | Wyon et al. | 2018 |
| Polish folk dancing | Laboratory (8 min) | 93.1% | Maciejczyk & Fec | 2013 |
| Aerobic dance (high impact, no arm involvement) | Laboratory (3 min) | 93.1% | Schaeffer-Gerschutz, Darby & Browder | 2000 |
| Hip-hop - Breakdancing | Laboratory (unspecified duration - set 1) | 93.2% | Wyon et al. | 2018 |
| Aerobic Dance | Laboratory (3 min - level 3) | 94.2% | Thomsen & Ballor | 1991 |

| | | | | |
|---|---|---|---|---|
| Sports Dancing (Latin) | Laboratory (10 min) | 94.7% | Liiv et al. | 2014 |
| Swedish folk dance (hambo) | Laboratory (4 min) | 94.8% | Wigaeus & Kilbom | 1980 |
| Sports Dancing | Laboratory (10 min) | 95.0% | Pilch et al. | 2017 |
| Sport Dancing (Latin) | Laboratory (10 min) | 95.3% | Bria et al. | 2011 |
| Sport Dancing (Latin) | Laboratory (10 min) | 95.5% | Bria et al. | 2011 |
| Highland Dance | Performance (unspecified duration) | 99.0% | Bailie, Wyon & Head | 2007 |
| Sports Dancing (Latin) | Laboratory (10 min) | 99.4% | Liiv et al. | 2014 |

*Table S18. Details of all included mean heart rate data from included studies of music participation.*

| | Instrument/singing | Type of performance | Mean heart rate (% max) | Author | Year |
|---|---|---|---|---|---|
| **Very Light (<57% max heart rate)** | Trumpet | Seated trumpet exercises | 42.3-44.5% | Robertson, Federoff & Eisensmith | 2010 |
| | Trumpet | Seated trumpet exercises (1 min each) | 47.2-53.5% | Robertson, Federoff & Eisensmith | 2010 |
| | Strings (n=23) | Rehearsal | 47.7% | Inesta et al | 2008 |
| | Varied instruments in orchestra | Concert 2 - rehearsal | 49.2% | Mulcahy et al. | 1990 |
| | Stringed instrument | Playing in experimental conditions for 60 mins | 49.4% | Horswill, Kien & Zipf | 1995 |
| | Marching band - light instruments (<5 pounds) | Rehearsal (75 min) | 49.7-50.6% | Strand & Sommer | 2005 |
| | Piano (n=10) | Rehearsal | 49.8% | Inesta et al | 2008 |
| | Varied instruments in orchestra | Concert 1 - rehearsal | 50.7% | Mulcahy et al. | 1990 |
| | Varied instruments in orchestra | Rehearsal ('private session') | 51.1% | Studer et al. | 2014 |
| | Singing/flute | Rehearsal | 51.6% | Harmat & Theorell | 2010 |
| | Band' instruments - i.e. guitar/bass/drums/vocals/keys/violin/trumpet/saxophone | Rehearsal | 52.1% | Vellers, Irwin & Lightfoot | 2015 |
| | Winds (n=25) | Rehearsal | 53.2% | Inesta et al | 2008 |
| | Singing/flute | Rehearsal | 54.2% | Harmat & Theorell | 2010 |
| | Varied instruments in orchestra | Concert 1 - performance | 54.3% | Mulcahy et al. | 1990 |
| | Percussion - drum corps (battery & front ensemble) | Rehearsal - 96 BPM | 54.4% | Dye & Barry | 2016 |
| | Marching band - heavy instruments (5 pounds or greater) | Rehearsal (75 min) | 54.6-55.5% | Strand & Sommer | 2005 |
| | Varied instruments in orchestra | Concert 2 - performance | 55.2% | Mulcahy et al. | 1990 |
| | Classical Indian Music (n=2) | Performance | 56.7% | Inesta et al | 2008 |
| **Light (57-63% max heart rate)** | Strings (n=23) | Performance | 58.2% | Inesta et al | 2008 |
| | Band' instruments - i.e. guitar/bass/drums/vocals/keys/violin/trumpet/saxophone | Performance | 59.1% | Vellers, Irwin & Lightfoot | 2015 |
| | Percussion (n=2 | Performance | 59.3% | Inesta et al | 2008 |
| | Percussion - drum corps (battery & front ensemble) | Rehearsal - 176 BPM | 60.5% | Dye & Barry | 2016 |
| | Clarinet | 1st Movement - Repertoire performance (Schumann, Phantasiestucke - 3 movements of increasing intensity) | 60.6% | Hahnengress & Boning | 2010 |
| | Winds (n=25) | Performance | 62.3% | Inesta et al | 2008 |
| | Operetta | Performance | 62.5% | Seliger et al | 1970 |
| | Clarinet | 2nd Movement - Repertoire performance (Schumann, Phantasiestucke - 3 movements of increasing intensity) | 62.7% | Hahnengress & Boning | 2010 |
| | Trumpet | Laboratory (etude - undefined length) | 61.0-67.7% | Hunsaker LA | 1994 |
| **Moderate (64-76% max heart rate)** | Singing/flute | Performance | 64.0% | Harmat & Theorell | 2010 |
| | Varied instruments in orchestra | Performance ('public session') | 65.8% | Studer et al. | 2014 |
| | Clarinet | 3rd Movement - Repertoire performance (Schumann, Phantasiestucke - 3 movements of increasing intensity) | 66.9% | Hahnengress & Boning | 2010 |

| | | | | | |
|---|---|---|---|---|---|
| | Trumpet | Laboratory (etude - undefined length) | 61.0-67.7% | Hunsaker LA | 1994 |
| | Opera | Performance | 68.6% | Seliger et al | 1970 |
| | Singing/flute | Performance | 68.7% | Harmat & Theorell | 2010 |
| | Opera | Performance | 70.2% | Seliger et al | 1970 |
| | Bagpipes | Laboratory (5 min) | 70.2-80.4% | Barr et al | 2000 |
| | Conductor (opera) | Performance | 72.3% | Seliger et al | 1970 |
| | Piano (n=10) | Performance | 73.7% | Inesta et al | 2008 |
| | Marching band - average of wind, percussion & flag carrier | Parade performance (132 steps/min; 50 min) | 75.4% | Erdmann et al | 2003 |
| | Drum set | 8 metal songs in lab | 76.7% | Romero et al. | 2016 |
| **Vigorous (77-95% max heart rate)** | Operetta | Performance | 77.4% | Seliger et al | 1970 |
| | Musical theater (singing & dance) | Laboratory (4 min) | 78.5% | Stephens & Wyon | 2020 |
| | Bagpipes | Laboratory (5 min) | 70.2-80.4% | Barr et al | 2000 |
| | Drum set | Concert (mean duration 38.6 min) | 87.8% | De La Rue et al. | 2013 |
| | Musical theater (singing & dance) | Laboratory (4 min) | 89.1% | Stephens & Wyon | 2020 |

*Table S19. Studies of the physical demands of performing arts participation excluded after full-text review, with reasons.*

| Author | Year | Title | Journal | Music/Dance | Reason for Exclusion |
|---|---|---|---|---|---|
| Adams | 2009 | High School Physical Education Students' Heart Rates During Different Activities | The Journal of Physical Education, Recreation & Dance | Dance | No reporting of mean HR during dance performance |
| Altamirano-Diaz et al | 2019 | Are active video games effective in eliciting at least moderate intensity physical activity in children and do children enjoy playing them? | Canadian Journal of Cardiology | Dance | No reporting of HR during dance performance |
| Angioi et al | 2009 | Fitness in contemporary dance: A systematic review | International Journal of Sports Medicine | Dance | Review article, no reporting of raw HR data |
| Bailey & McInnis | 2011 | Energy cost of exergaming: a comparison of the energy cost of 6 forms of exergaming | Archives of Pediatrics & Adolescent Medicine | Dance | No reporting of mean HR during dance performance |
| Bell & Bassey | 1994 | A comparison of the relation between oxygen uptake and heart rate during different styles of aerobic dance and a traditional step test in women | European Journal of Applied Physiology | Dance | No reporting of mean HR, only peak HR |
| Blessing et al. | 1987 | The physiologic effects of eight weeks of aerobic dance with and without hand-held weights | American Journal of Sports Medicine | Dance | No reporting of mean HR during dance performance |
| Blessing, Tucker & Williford | 1987 | Training factors and physical fitness among aerobic dance instructors. | Perceptual & Motor Skills | Dance | Incompatible data, reporting only of %HR max using experimentally determined max HR |
| Blyth & Goslin | 1985 | Cardiorespiratory responses to "aerobic dance" | Journal of Sports Medicine & Physical Fitness | Dance | No article access |
| Cain et al. | 2015 | Physical activity in youth dance classes | Pediatrics | Dance | No reporting of mean HR during dance performance |
| Campos et al. | 2020 | Evaluation of the exercise intensity generated by active video gaming in patients with cystic fibrosis and healthy individuals | Journal of Cystic Fibrosis | Dance | No reporting of mean HR during dance performance |
| Cheng & Zheng | 1997 | Study on the training load of aerobic dance in men's individual event | Journal of Guangzhou Physical Education Institute | Dance | Article in Mandarin |
| Chernozub et al. | 2018 | The influence of dance and power fitness loads on the body morphometric parameters and peculiarities of adaptive-compensatory reactions of organism of young women | Journal of Physical Education & Sport | Dance | No reporting of mean HR during dance performance |
| Chren, Spanik & Kyselovicova | 2010 | Blood lactate concentration of ballroom dancers according to the length of their routines | Acta Facultatis Educationis Physicae Universitatis Comenianae | Dance | No reporting of HR during dance performance |
| Cohen | 1984 | Dance - aerobic and anaerobic | Journal of Physical Education, Recreation & Dance | Dance | Review article, no reporting of raw HR data |
| Cohen, Segal & McArdle | 1982 | Heart rate response to ballet stage performance | Physician and Sportsmedicine | Dance | No synthesis of HR data, reporting of selected individual data only |
| Cossette et al | 2008 | Chest wall dynamics and muscle recruitment during professional flute playing | Respiratory Physiology and Neurobiology | Music | No reporting of HR during music performance |
| Dahlstrom | 1997 | Physical effort during dance training: a comparison between teachers and students | Journal of Dance Medicine & Science | Dance | No reporting of mean HR during dance performance |
| Dahlstrom et al | 1996 | Physical fitness and physical effort in dancers: a comparison of four major dance styles | Impulse: International Journal of Dance Science, Medicine & Education | Dance | No reporting of mean HR during dance performance |
| Dahlstrom et al. | 1990 | Discrepancy between estimated energy intake and requirement in female dancers | Clinical Physiology | Dance | No reporting of mean HR during dance performance |
| D'Angelo | 1994 | The effect of foot articulation on exercise intensity during aerobic dance | Thesis | Dance | Study of isolated individual aerobic dance movements (march stepping) only |

| | | | | | |
|---|---|---|---|---|---|
| Domene et al. | 2014 | Physiological and perceptual responses to Latin partnered social dance | Human Movement Science | Dance | No reporting of mean HR during dance performance |
| Domene et al. | 2016 | Salsa dance and Zumba fitness: Acute responses during community-based classes | Journal of Sport and Health Science | Dance | Incompatible data, reporting only of %HRmax & %HRR using laboratory measured HR max values |
| D'Ottavio et al. | 2016 | Energy Expenditure in Professional DanceSport | Journal of Dance Medicine & Science | Dance | HR only collected during the last 30 seconds of each dance |
| Drinkwater & Klopper | 2010 | Quantifying the physical demands of a musical performance and their effects on performance quality | Medical Problems of Performing Artists | Music | No reporting of HR during music performance |
| Duan, Zhang & Shao-jun | 2010 | A Research on the Heart Rates' Variation during the Second Set Performance of the Northern Lion Dance | Journal of Beijing Sport University | Dance | Article in Mandarin |
| Elliot, Morton & Johnston | 1991 | Biomechanical and physiological responses to modes of locomotion used in aerobic dance | Australian Journal of Science and Medicine in Sport | Dance | No examination of dancing, only 'aerobic dance locomotion' (walking & jogging) |
| Engels, Bowen & Wirth | 1995 | Routine use of external weights during a low-impact aerobic dance conditioning program: Training benefit | Sports Medicine, Training & Rehabilitation | Dance | No reporting of mean HR during dance performance |
| Evardsen, Ingjer & Bo | 2011 | Fit women are not able to use the whole aerobic capacity during aerobic dance | Journal of Strength & Conditioning Research | Dance | No reporting of mean HR during dance performance |
| Froberg et al. | 2017 | Levels of physical activity during physical education lessons in Sweden | Acta Paediatrica | Dance | No reporting of mean HR during dance performance |
| Froemel, Oscheutz & Schaller | 1999 | Heart rate and energy costs during dancing (country dancing) by elderly women | Sportonomics | Dance | No reporting of mean HR during dance performance |
| Grant et al. | 1998 | A comparison of physiological responses and rating of perceived exertion between high-impact and low-impact aerobic dance sessions | European Journal of Applied Physiology and Occupational Physiology | Dance | Reporting of % HR max calculated using actual HR max (not predicted) only, no raw HR values reported |
| Guidetti et al. | 2008 | Energy cost and energy sources of a ballet dance exercise in female adolescents with different technical ability | European Journal of Applied Physiology | Dance | No reporting of mean HR during dance performance |
| Guidetti et al. | 2007 | Exercise intensities during a ballet lesson in female adolescents with different technical ability | International Journal of Sports Medicine | Dance | No reporting of raw HR data (only statistical analyses of HR data) |
| Hausken & Dyrstad | 2016 | Using heart rate monitors to assess energy expenditure in four training types | Gazzetta Medica Italiana Archivio per le Scienze Mediche | Dance | Incompatible data, reporting only of %HR max using experimentally determined max HR |
| Hawley, Williams & Hurley | 1990 | Physiological and psychological response to aerobic dance classes | New Zealand Journal of Sports Medicine | Dance | No reporting of mean HR during dance performance |
| Helin | 1988 | Activation in professional ballet dancers | Physiology & Behavior | Dance | No reporting of HR during dance performance |
| Hirsch et al | 2003 | Energy balance and physical activity patterns in university ballet dancers | Journal of Dance Medicine & Science | Dance | No article access + appears to focus on total daily EE (not dance-specific) |
| Hollis et al. | 2017 | A systematic review and meta-analysis of moderate-to-vigorous physical activity levels in secondary school physical education lessons | International Journal of Behavioral Nutrition and Physical Activity | Dance | Review article, no reporting of raw HR data |
| Hunsaker & Ramsey | 1998 | Cardiac dynamics in marching band trumpet players | Medical Problems of Performing Artists | Music | Reporting of maximal HR data only during performance |
| Ipsen | 1994 | Cardiovascular and body composition responses to aerobic dance training of varying frequencies and total program lengths | Thesis | Dance | No reporting of mean HR during dance performance |
| Jakubec et al | 2008 | Changes in heart rate variability after a six month long aerobic dance or step-dance program in women 40-65 years old: The influence of different degrees of adherence, intensity and initial levels | Acta Universitatis Palackianae Olomucensis. Gymnica | Dance | No reporting of mean HR during dance performance |
| Jeffries, Wallace & Coutts | 2017 | Quantifying Training Loads in Contemporary Dance | International Journal of Sports Physiology and Performance | Dance | No reporting of mean HR during dance performance |
| Jette & Inglis | 1975 | Energy cost of square dancing | Journal of Applied Physiology | Dance | No reporting of mean HR during dance performance |

| | | | | | |
|---|---|---|---|---|---|
| Kirkendall & Calabrese | 1983 | Physiological aspects of dance | Clinics in Sports Medicine | Dance | No reporting of mean HR during dance performance |
| Kozai et al. | 2020 | Workload Intensity and Rest Periods in Professional Ballet: Connotations for Injury | International Journal of Sports Medicine | Dance | No reporting of mean HR during dance performance |
| Lankford et al. | 2014 | The Energy Expenditure of Recreational Ballroom Dance | International Journal of Exercise Science | Dance | No reporting of mean HR during dance performance |
| Larger & Ledoux | 1996 | Dimsdale JE, Nelesen RA. French-horn hypertension. N EnglJ Med 1995; 333: 326-27. | Lancet | Music | Letter to the editor only describing heart response of one note at a time |
| Liebana et al. | 2018 | Muscle Activation in the Main Muscle Groups of the Lower Limbs in High-Level Dancesport Athletes | Medical Problems of Performing Artists | Dance | No reporting of mean HR during dance performance |
| Lira et al. | 2018 | Acute Caffeine Supplementation Does Not Change Sweat Rate and Blood Pressure in Ballet Dancers: A Double-Blind and Placebo-Controlled Study | Journal of Dance Medicine & Science | Dance | No reporting of HR during dance performance |
| Lyons et al | 2011 | Energy expenditure and enjoyment during video game play: differences by game type | Medicine and Science in Sports and Exercise | Dance | No reporting of mean HR during dance performance |
| Lyons et al. | 2014 | Engagement, enjoyment and energy expenditure during active video game play | Health Psychology | Dance | No reporting of mean HR during dance performance |
| Massidda et al | 2011 | Energy expenditure during competitive Latin American dancing simulation | Medical Problems of Performing Artists | Dance | No reporting of mean HR during dance performance |
| Matthe et al. | 2019 | Central retinal venous pressure is higher than intraocular pressure during amateur trumpet playing | Graefe's Archive for Clinical and Experimental Opthalmology | Music | No reporting of HR during music performance |
| Mavrovouniotis et al | 2008 | Greek traditional dances and quality of old people’s life | Journal of Body and Movement Therapy | Dance | No reporting of mean HR during dance performance |
| McDonough et al | 2018 | Comparison of college students' energy expenditure, physical activity, and enjoyment during exergaming and traditional exercise | Journal of Clinical Medicine | Dance | No reporting of mean HR during dance performance |
| Mengelkock, Highsmith & Morris | 2014 | Comparison of the Metabolic Demands of Dance Performance Using Three Mobility Devices for a Dancer with Spinal Cord Injury and an Able-Bodied Dancer | Medical Problems of Performing Artists | Dance | Analysis of dance in 'clinical situations' (different types of mobility devices) only |
| Metcalf | 1983 | Telemetered heart rate responses in adult men and women aerobic dance instructors | Sports Medicine Digest | Dance | No article access |
| Metcalf et al. | 1981 | ECG effects of aerobic dance: A study of five exercise-conditioned young women | Postgraduate Medicine | Dance | No reporting of mean HR during dance performance |
| Milburn & Butts | 1983 | A comparison of the training responses to aerobic dance and jogging in college females | Medicine and Science in Sports and Exercise | Dance | Dancing with a 'target heart rate' |
| Miranda et al | 2017 | Energy Cost of Active and Sedentary Music Video Games: Drum and Handheld Gaming vs. Walking and Sitting | International Journal of Exercise Science | Music | No reporting of HR during music performance |
| Morgan | 1985 | A comparison of the energy requirements of selected dance programs | Physical Educator | Dance | Review article, no reporting of raw HR data |
| Nelson et al. | 1988 | Cardiac frequency and caloric cost of aerobic dancing in young women | Research Quarterly for Exercise and Sport | Dance | No reporting of mean HR during dance performance |
| Noah et al. | 2011 | Vigorous Energy Expenditure with a Dance Exer-game | Journal of Exercise Physiology Online | Dance | No reporting of mean HR during dance performance |
| Novak, Magill & Schutte | 1978 | Maximal oxygen intake and body composition of female dancers | European Journal of Applied Physiology and Occupational Physiology | Dance | No reporting of mean HR during dance performance |
| Papatzikis & Papatziki | 2016 | Investigating heart rate and rhythm changes in an infant's music education course: A case study | Psychology of Music | Music | No reporting of HR during music performance |
| Parker et al. | 1989 | Failure of target heart rate to accurately monitor intensity during aerobic dance | Medicine and Science in Sports and Exercise | Dance | Aerobic dance performed based on 'target HR |

| | | | | | |
|---|---|---|---|---|---|
| Perry et al. | 1988 | A comparison of training responses to interval versus continuous aerobic dance | The Journal of Sports Medicine and Physical Fitness | Dance | Article unable to be accessed, insufficient information in abstract |
| Raymond et al | 2005 | Biofeedback and dance performance: a preliminary investigation | Applied Psychophysiology and Biofeedback | Dance | No reporting of mean HR, only HR variability |
| Raynor, Cardoso & Bond | 2016 | Effect of exposure to greater active videogame variety on time spent in moderate- to vigorous-intensity physical activity | Physiology and Behavior | Dance | No reporting of mean HR during dance performance |
| Redding et al. | 2009 | The development of a high intensity dance performance fitness test | Journal of Dance Medicine & Science | Dance | Study of a dance-based fitness test, not dance performance |
| Redding et al. | 2004 | Validity of using heart rate as a predictor of oxygen consumption in dance | Journal of Dance Medicine & Science | Dance | No reporting of mean HR during dance performance |
| Rensing, Schemmann & Zalpour | 2018 | Musculoskeletal Demands in Violin and Viola Playing: A Literature Review | Medical Problems of Performing Artists | Music | No reporting of HR during music performance |
| Rice, Gurchiek & McBride | 2018 | Physiological and Biomechanical Responses to an Acute Bout of High Kicking in Dancers | Journal of Strength & Conditioning Research | Dance | Analysis of fitness, not dance, routine |
| Rimmer, Jay & Plowman | 1994 | Physiological characteristics of trained dancers and intensity level of ballet class and rehearsal | Impulse: International Journal of Dance Science, Medicine & Education | Dance | No reporting of mean HR during dance performance |
| Rinne, Miilunpalo & Heinonen | 2007 | Evaluation of required motor abilities in commonly practiced exercise modes and potential training effects among adults | Journal of Physical Activity & Health | Dance | No reporting of mean HR during dance performance |
| Rokka, Mavridis & Kouli | 2010 | The impact of exercise intensity on mood state of participants in dance aerobics programs | Studies in Physical Culture & Tourism | Dance | No reporting of mean HR during dance performance |
| Roopchand-Martin et al. | 2015 | A pilot study using the XBOX Kinect for exercise conditioning in sedentary female university students | Technology and Healthcare | Dance | No reporting of mean HR during dance performance |
| Schantz & Astrand | 1984 | Physiological characteristics of classical ballet | Medicine and Science in Sports and Exercise | Dance | No reporting of mean HR during dance performance |
| Scharff-Olson, Williford & Smith | 1992 | The heart rate VO2 relationship of aerobic dance: A comparison of target heart rate methods | Journal of Sports Medicine & Physical Fitness | Dance | No reporting of mean HR during dance performance |
| Shi et al | 2001 | Preliminary study of physiological characteristics of dragon dance | Journal of Beijing Sport University | Dance | Article in Mandarin |
| Shimzu et al. | 2012 | Catecholamine responses to 6h extended daily practice of marching band activities as a performing art | Journal of Science and Medicine in Sport | Music | No reporting of HR during music performance |
| Silva et al. | 2012 | Analysis of Acute Cardiovascular Responses in Experienced Practitioners of Capoeira: A Brazilian Art Form | Journal of Exercise Physiology Online | Dance | No reporting of HR during dance performance |
| Silva et al. | 2013 | Heart Rate Responses During and After the Practice of Capoeira: A Brazilian Art Form -- Part II | Journal of Exercise Physiology Online | Dance | No reporting of mean HR during dance performance |
| Skarsem Reitlo et al. | 2017 | Exercise patterns in older adults randomized to moderate-or high-intensity training: The Generation 100 study | European Geriatric Medicine | Dance | No reporting of mean HR during dance performance |
| Surgenor & Wyon | 2019 | Measuring Training Load in Dance: The Construct Validity of Session-RPE | Medical Problems of Performing Artists | Dance | No reporting of mean HR during dance performance |
| Taskin & Vardar Yagli | 2019 | An evaluation of physical activity level, respiratory muscle endurance and pain relationship in professional dancers | Turkish Journal of Physiotherapy and Rehabilitation | Dance | No reporting of mean HR during dance performance |
| Taylor et al | 2011 | Activity-promoting gaming systems in exercise and rehabilitation | Journal of Rehabilitation Research and Development | Dance | No reporting of HR during dance performance |
| Trost, Drovandi & Pfeiffer | 2016 | Developmental Trends in the Energy Cost of Physical Activities Performed by Youth | Journal of Physical Activity & Health | Dance | No reporting of mean HR during dance performance |
| Tucker, Faulkner & Horvath | 1971 | Electrocardiography and lung function in brass instrument players | Archives of Environmental Health | Music | HR data only for brief arpeggios |

| | | | | | |
|---|---|---|---|---|---|
| Vaczi et al. | 2016 | Ballroom dancing is more intensive for the female partners due to their unique hold technique | Physiology International | Dance | No reporting of mean HR, only peak HR |
| Verhoeven et al | 2015 | Energy Expenditure During Xbox Kinect Play in Early Adolescents: The Relationship with Player Mode and Game Enjoyment | Games for Health Journal | Dance | No reporting of mean HR during dance performance |
| Vogelaar | 1998 | Castrati in western art music, Part 2: Selection and musical training: Physical and psychosocial implications | Medical Problems of Performing Artists | Music | No reporting of HR during music performance |
| Wanke et al. | 2020 | Work related cardiovascular load in professional dance teachers-a pilot study | Journal of Occupational Medicine and Toxicology | Dance | Incompatible data, reporting only of %HR max using experimentally determined max HR |
| Wasley et al. | 2012 | Influence of fitness and physical activity on cardiovascular reactivity to musical performance | Work | Music | No reporting of HR during music performance |
| Williford, Scharff-Olson & Blessing | 1989 | The physiological effects of aerobic dance. A review | Sports Medicine | Dance | No reporting of mean HR during dance performance |
| Wyon | 2005 | Cardiorespiratory Training for Dancers | Journal of Dance Medicine & Science | Dance | Review article, no reporting of raw HR data |
| Yang | 2013 | Samba fitness efficacy research based on analysis of body compositions changes | Journal of Chemical and Pharmaceutical Research | Dance | No reporting of mean HR during dance performance |
| Zanchini & Malaguti | 2014 | Energy requirements in top-level DanceSport athletes | Journal of Human Sport & Exercise | Dance | No reporting of mean HR during dance performance |

### *REFERENCE LIST*

**Included reviews – Dance**

Burkhardt, J. and C. Brennan (2012). "The effects of recreational dance interventions on the health and well-being of children and young people: a systematic review." Arts & Health **4**(2): 148-161.

Clarke, F., et al. (2018). "Balance in theatrical dance performance: a systematic review." Medical Problems of Performing Artists **33**(4): 275-285.

Cugusi, L., et al. (2019). "Zumba Fitness and Women's Cardiovascular Health: A SYSTEMATIC REVIEW." Journal of cardiopulmonary rehabilitation and prevention **39**(3): 153-160.

da Silveira Costa, M. S., et al. (2013). "Static and dynamic balance in ballet dancers: a literature review." Fisioter. Pesqui **20**: 299-305.

Fernández-Argüelles, E. L., et al. (2015). "Effects of dancing on the risk of falling related factors of healthy older adults: a systematic review." Archives of gerontology and geriatrics **60**(1): 1-8.

Hwang, P. W.-N. and K. L. Braun (2015). "The effectiveness of dance interventions to improve older adults' health: a systematic literature review." Alternative therapies in health and medicine **21**(5): 64.

Keogh, J. W., et al. (2009). "Physical benefits of dancing for healthy older adults: a review." Journal of aging and physical activity **17**(4): 479-500.

Letton, M. E., et al. (2020). "The Effectiveness of Classical Ballet Training on Health-Related Outcomes: A Systematic Review." Journal of Physical Activity and Health **17**(5): 566-574.

Liu, X., et al. (2020). "Dance intervention effects on physical function in healthy older adults: a systematic review and meta-analysis." Aging clinical and experimental research: 1-11.

Meng, X., et al. (2020). "Effects of dance intervention on global cognition, executive function and memory of older adults: a meta-analysis and systematic review." Aging clinical and experimental research **32**(1): 7-19.

Rodrigues-Krause, J., et al. (2016). "Effects of dance interventions on cardiovascular risk with ageing: systematic review and meta-analysis." Complementary Therapies in Medicine **29**: 16-28.

Rodrigues-Krause, J., et al. (2019). "Dancing for Healthy Aging: Functional and Metabolic Perspectives." Alternative Therapies in Health & Medicine **25**(1).

Teixeira-Machado, L., et al. (2019). "Dance for neuroplasticity: A descriptive systematic review." Neuroscience & Biobehavioral Reviews **96**: 232-240.

Veronese, N., et al. (2017). "Dance movement therapy and falls prevention." Maturitas **102**: 1-5.

Yan, A. F., et al. (2018). "The effectiveness of dance interventions on physical health outcomes compared to other forms of physical activity: a systematic review and meta-analysis." Sports Medicine **48**(4): 933-951.

**Included Reviews – Music**

Butzlaff, R. (2000). "Can music be used to teach reading?" Journal of aesthetic education **34**(3/4): 167-178.

Clift, S., et al. (2010). "Group singing, wellbeing and health: A systematic mapping of research evidence." Unesco Observatory **2**(1).

Coffey, E. B., et al. (2017). "Speech-in-noise perception in musicians: A review." Hearing research **352**: 49-69.

Daykin, N., et al. (2013). "Music making for health, well-being and behaviour change in youth justice settings: a systematic review." Health promotion international **28**(2): 197-210.

Daykin, N., et al. (2018). "What works for wellbeing? A systematic review of wellbeing outcomes for music and singing in adults." Perspectives in Public Health **138**(1): 39-46.

Eells, K. (2014). "The use of music and singing to help manage anxiety in older adults." Mental Health Practice **17**(5).

Fancourt, D., et al. (2014). "The psychoneuroimmunological effects of music: A systematic review and a new model." Brain, behavior, and immunity **36**: 15-26.

Gick, M. L. (2011). "Singing, health and well-being: A health psychologist's review." Psychomusicology: Music, Mind and Brain **21**(1-2): 176.

Gordon, R. L., et al. (2015). "Does music training enhance literacy skills? A meta-analysis." Frontiers in psychology **6**: 1777.

Hetland, L. (2000). "Learning to make music enhances spatial reasoning." Journal of aesthetic education **34**(3/4): 179-238.

Jaschke, A. C., et al. (2013). "Music education and its effect on intellectual abilities in children: a systematic review." Reviews in the Neurosciences **24**(6): 665-675.

Moore, K. S. (2013). "A systematic review on the neural effects of music on emotion regulation: implications for music therapy practice." Journal of music therapy **50**(3): 198-242.

Phillips, C. S. and H. Becker (2019). "Systematic Review: Expressive arts interventions to address psychosocial stress in healthcare workers." Journal of advanced nursing **75**(11): 2285-2298.

Raglio, A., et al. (2019). "Music in the workplace: A narrative literature review of intervention studies." Journal of Complementary and Integrative Medicine **1**(ahead-of-print).

Skingley, A. and T. Vella-Burrows (2010). "Therapeutic effects of music and singing for older people." Nursing standard **24**(19).

Talamini, F., et al. (2017). "Musicians have better memory than nonmusicians: A meta-analysis." PloS one **12**(10): e0186773.

Vaughn, K. (2000). "Music and mathematics: Modest support for the oft-claimed relationship." Journal of aesthetic education **34**(3/4): 149-166.

Yap, A. F., et al. (2017). "A systematic review on the effects of active participation in rhythm-centred music making on different aspects of health." European Journal of Integrative Medicine **9**: 44-49.

**Included Individual Studies** *(from included reviews)* **– Dance**

Adiputra, N., et al. (1996). "Balinese dance exercises improve the maximum aerobic capacity." Journal of human ergology **25**(1): 25.

Alpert, P. T., et al. (2009). "The effect of modified jazz dance on balance, cognition, and mood in older adults." Journal of the American Academy of Nurse Practitioners **21**(2): 108-115.

Alves, H. (2013). Dancing and the aging brain: the effects of a 4-month ballroom dance intervention on the cognition of healthy older adults, University of Illinois at Urbana-Champaign.

Ambegaonkar, J. P., et al. (2013). "Balance comparisons between female dancers and active nondancers." Research quarterly for exercise and sport **84**(1): 24-29.

Araneta, M. and D. Tanori (2014). "Benefits of Zumba Fitness® among sedentary adults with components of the metabolic syndrome: a pilot study." The Journal of sports medicine and physical fitness **55**(10): 1227-1233.

Baniqued, P. L., et al. (2018). "Brain network modularity predicts exercise-related executive function gains in older adults." Frontiers in aging neuroscience **9**: 426.

Barene, S., et al. (2016). "Effects on muscle strength, maximal jump height, flexibility and postural sway after soccer and Zumba exercise among female hospital employees: a 9-month randomised controlled trial." Journal of sports sciences **34**(19): 1849-1858.

Barene, S., et al. (2014). "Soccer and Zumba as health-promoting activities among female hospital employees: a 40-weeks cluster randomised intervention study." Journal of sports sciences **32**(16): 1539-1549.

Barene, S., et al. (2014). "Effects of the workplace health promotion activities soccer and Zumba on muscle pain, work ability and perceived physical exertion among female hospital employees." PloS one **9**(12): e115059.

Belardinelli, R., et al. (2008). "Waltz dancing in patients with chronic heart failure: new form of exercise training." Circulation: Heart Failure **1**(2): 107-114.

Bennell, K., et al. (2000). "Activity-associated differences in bone mineral are evident before puberty: a cross-sectional study of 130 female novice dancers and controls." Pediatric Exercise Science **12**(4): 371-381.

Bennett, C. G. and M. E. Hackney (2018). "Effects of line dancing on physical function and perceived limitation in older adults with self-reported mobility limitations." Disability and rehabilitation **40**(11): 1259-1265.

Blackman, L., et al. (1988). "The effects of dance team participation on female adolescent physical fitness and self-concept." Adolescence **23**(90): 437.

Bronner, S. (2012). "Differences in segmental coordination and postural control in a multi-joint dance movement: developpe arabesque." Journal of dance medicine & science **16**(1): 26-35.

Burgess, G., et al. (2006). "Effects of a 6-week aerobic dance intervention on body image and physical self-perceptions in adolescent girls." Body image **3**(1): 57-66.

Burzynska, A. Z., et al. (2017). "The dancing brain: structural and functional signatures of expert dance training." Frontiers in human neuroscience **11**: 566.

Casabona, A., et al. (2016). "Specificity of foot configuration during bipedal stance in ballet dancers." Gait & posture **46**: 91-97.

Cepeda, C. C., et al. (2015). "Effect of an eight-week ballroom dancing program on muscle architecture in older adult females." Journal of aging and physical activity **23**(4): 607-612.

Chen, Z. (2014). "The Effect of square dance movement intervention on cognitive function in the elderly." Chinese Rural Health Service Administration **34**: 879-881.

Coubard, O. A., et al. (2011). "Practice of contemporary dance improves cognitive flexibility in aging." Frontiers in aging neuroscience **3**: 13.

Crotts, D., et al. (1996). "Balance abilities of professional dancers on select balance tests." Journal of Orthopaedic & Sports Physical Therapy **23**(1): 12-17.

Cruz-Ferreira, A., et al. (2015). "Creative dance improves physical fitness and life satisfaction in older women." Research on aging **37**(8): 837-855.

Cugusi, L., et al. (2015). "Cardiovascular effects, body composition, quality of life and pain after a Zumba fitness program in Italian overweight women." The Journal of sports medicine and physical fitness **56**(3): 328-335.

da Silva Borges, E. G., et al. (2012). "The effect of ballroom dance on balance and functional autonomy among the isolated elderly." Archives of gerontology and geriatrics **55**(2): 492-496.

da Silva Borges, E. G., et al. (2014). "Postural balance and falls in elderly nursing home residents enrolled in a ballroom dancing program." Archives of gerontology and geriatrics **59**(2): 312-316.

Daley, A. J. and J. Buchanan (1999). "Aerobic dance and physical self-perceptions in female adolescents: Some implications for physical education." Research quarterly for exercise and sport **70**(2): 196-200.

Delextrat, A. A., et al. (2016). "An 8-week exercise intervention based on Zumba improves aerobic fitness and psychological well-being in healthy women." Journal of Physical Activity and Health **13**(2): 131-139.

Doi, T., et al. (2017). "Effects of cognitive leisure activity on cognition in mild cognitive impairment: results of a randomized controlled trial." Journal of the American Medical Directors Association **18**(8): 686-691.

Domene, P. A., et al. (2016). "The health-enhancing efficacy of Zumba® fitness: An 8-week randomised controlled study." Journal of sports sciences **34**(15): 1396-1404.

Donath, L., et al. (2014). "The effects of Zumba training on cardiovascular and neuromuscular function in female college students." European Journal of Sport Science **14**(6): 569-577.

Ehlers, D. K., et al. (2017). "Regional brain volumes moderate, but do not mediate, the effects of group-based exercise training on reductions in loneliness in older adults." Frontiers in aging neuroscience **9**: 110.

Eyigor, S., et al. (2009). "A randomized controlled trial of Turkish folklore dance on the physical performance, balance, depression and quality of life in older women." Archives of gerontology and geriatrics **48**(1): 84-88.

Federici, A., et al. (2005). "Does dance-based training improve balance in adult and young old subjects? A pilot randomized controlled trial." Aging clinical and experimental research **17**(5): 385-389.

Ferrufino, L., et al. (2011). "Practice of contemporary dance promotes stochastic postural control in aging." Frontiers in human neuroscience **5**: 169.

Flores, R. (1995). "Dance for health: improving fitness in African American and Hispanic adolescents." Public health reports **110**(2): 189.

Ford Jr, H. T., et al. (1989). "Effects of selected physical activities on health-related fitness and psychological well-being." Psychological Reports **64**(1): 203-208.

Garber, C. E., et al. (1992). "Is aerobic dance an effective alternative to walk-jog exercise training?" The Journal of sports medicine and physical fitness **32**(2): 136-141.

Gerbino, P. G., et al. (2007). "Comparison of standing balance between female collegiate dancers and soccer players." Gait & posture **26**(4): 501-507.

Goertzen, D., et al. (1984). "The functional capacity and physical activity levels of women over 60 years of age." Journal of sports medicine and physical fitness **24**(1): 30-36.

Golomer, E. and P. Dupui (2000). "Spectral analysis of adult dancers' sways: sex and interaction vision-proprioception." International Journal of Neuroscience **105**(1-4): 15-26.

Golomer, E., et al. (1997). "Sex-linked differences in equilibrium reactions among adolescents performing complex sensorimotor tasks." Journal of Physiology-Paris **91**(2): 49-55.

Golomer, E., et al. (1999). "The contribution of vision in dynamic spontaneous sways of male classical dancers according to student or professional level." Journal of Physiology-Paris **93**(3): 233-237.

Golomer, E., et al. (2010). "Right hemisphere in visual regulation of complex equilibrium: the female ballet dancers' experience." Neurological Research **32**(4): 409-415.

Golomer, E., et al. (2009). "Spontaneous whole body rotations and classical dance expertise: How shoulder–hip coordination influences supporting leg displacements." Journal of Electromyography and Kinesiology **19**(2): 314-321.

Granacher, U., et al. (2012). "Effects of a salsa dance training on balance and strength performance in older adults." Gerontology **58**(4): 305-312.

Grant, S., et al. (2002). "A comparison of physiological responses and rating of perceived exertion in two modes of aerobic exercise in men and women over 50 years of age." British Journal of Sports Medicine **36**(4): 276-281.

Guillou, E., et al. (2007). "Dynamic balance sensory motor control and symmetrical or asymmetrical equilibrium training." Clinical Neurophysiology **118**(2): 317-324.

Hackney, M. E., et al. (2015). "Adapted Tango improves mobility, motor–cognitive function, and gait but not cognition in older adults in independent living." Journal of the American Geriatrics Society **63**(10): 2105-2113.

Hackney, M. E., et al. (2007). "Effects of tango on functional mobility in Parkinson's disease: a preliminary study." Journal of neurologic physical therapy **31**(4): 173-179.

Hamacher, D., et al. (2015). "The effect of a six-month dancing program on motor-cognitive dual-task performance in older adults." Journal of aging and physical activity **23**(4): 647-652.

Hashimoto, H., et al. (2015). "Effects of dance on motor functions, cognitive functions, and mental symptoms of Parkinson's disease: a quasi-randomized pilot trial." Complementary Therapies in Medicine **23**(2): 210-219.

Hedgepeth, S. L. C. (1987). The effects of ballet training on isokinetic strength, power, and muscular endurance, Virginia Tech.

Heffron, M., et al. (1997). "Weight-training and bone mass in post-menopausal women." Sports Excercise and Injury **3**(3): 143-149.

Holmerová, I., et al. (2010). "Effect of the exercise dance for seniors (EXDASE) program on lower-body functioning among institutionalized older adults." Journal of aging and health **22**(1): 106-119.

Hopper, D. M., et al. (2014). "The effects of vestibular stimulation and fatigue on postural control in classical ballet dancers." Journal of dance medicine & science **18**(2): 67-73.

Houston, S. and A. McGill (2013). "A mixed-methods study into ballet for people living with Parkinson's." Arts & Health **5**(2): 103-119.

Hugel, F., et al. (1999). "Postural control of ballet dancers: a specific use of visual input for artistic purposes." International journal of sports medicine **20**(02): 86-92.

Hui, E., et al. (2009). "Effects of dance on physical and psychological well-being in older persons." Archives of gerontology and geriatrics **49**(1): e45-e50.

Isler, A. K., et al. (2001). "Effects of step aerobics and aerobic dancing on serum lipids and lipoproteins." Journal of sports medicine and physical fitness **41**(3): 380-385.

Janyacharoen, T., et al. (2013). "Physical performance in recently aged adults after 6 weeks traditional Thai dance: a randomized controlled trial." Clinical interventions in aging **8**: 855.

Jarvis, D. N., et al. (2014). "Trunk coordination in dancers and nondancers." Journal of applied biomechanics **30**(4): 547-554.

Kaltsatou, A. C., et al. (2014). "Functional and psychosocial effects of either a traditional dancing or a formal exercising training program in patients with chronic heart failure: a comparative randomized controlled study." Clinical Rehabilitation **28**(2): 128-138.

Kattenstroth, J.-C., et al. (2013). "Six months of dance intervention enhances postural, sensorimotor, and cognitive performance in elderly without affecting cardio-respiratory functions." Frontiers in aging neuroscience **5**: 5.

Kattenstroth, J.-C., et al. (2011). "Balance, sensorimotor, and cognitive performance in long-year expert senior ballroom dancers." Journal of Aging Research **2011**.

Kattenstroth, J.-C., et al. (2010). "Superior sensory, motor, and cognitive performance in elderly individuals with multi-year dancing activities." Frontiers in aging neuroscience **2**: 31.

Kiefer, A. W., et al. (2011). "Multi-segmental postural coordination in professional ballet dancers." Gait & posture **34**(1): 76-80.

Kilroy, E. A., et al. (2016). "The effect of single-leg stance on dancer and control group static balance." International Journal of Exercise Science **9**(2): 110.

Kim, S. and J. Kim (2007). "Mood after various brief exercise and sport modes: aerobics, hip-hop dancing, ice skating, and body conditioning." Perceptual and motor skills **104**(3_suppl): 1265-1270.

Kim, S.-H., et al. (2011). "Effect of dance exercise on cognitive function in elderly patients with metabolic syndrome: a pilot study." Journal of sports science & Medicine **10**(4): 671.

Kosmat, H. and A. Vranic (2017). "The efficacy of a dance intervention as cognitive training for the old-old." Journal of aging and physical activity **25**(1): 32-40.

Kouli, O., et al. (2009). "The effects of an aerobic program on health-related fitness and intrinsic motivation in elementary school pupils." Studies in Physical Culture & Tourism **16**(3): 301-306.

Krampe, J., et al. (2010). "Dance-based therapy in a program of all-inclusive care for the elderly: an integrative approach to decrease fall risk." Nursing administration quarterly **34**(2): 156-161.

Krishnan, S., et al. (2015). "Zumba® dance improves health in overweight/obese or type 2 diabetic women." American journal of health behavior **39**(1): 109-120.

Krityakiarana, W. and N. Jongkamonwiwat (2016). "Comparison of balance performance between Thai classical dancers and non-dancers." Journal of dance medicine & science **20**(2): 72-78.

Kudlacek, S., et al. (1997). "The impact of a senior dancing program on spinal and peripheral bone mass1." American journal of physical medicine & rehabilitation **76**(6): 477-481.

Kujala, U., et al. (1997). "Training does not increase maximal lumbar extension in healthy adolescents." Clinical biomechanics **12**(3): 181-184.

Lazarou, I., et al. (2017). "International ballroom dancing against neurodegeneration: a randomized controlled trial in Greek community-dwelling elders with mild cognitive impairment." American Journal of Alzheimer's Disease & Other Dementias® **32**(8): 489-499.

Lesser, I. A., et al. (2016). "Effectiveness of exercise on visceral adipose tissue in older South Asian Women." Med Sci Sports Exerc **48**(7): 1371-1378.

Lesté, A. and J. Rust (1990). "Effects of dance on anxiety." American Journal of Dance Therapy **12**(1): 19-25.

Lin, C.-F., et al. (2011). "Comparison of postural stability between injured and uninjured ballet dancers." The American journal of sports medicine **39**(6): 1324-1331.

Lin, C.-W., et al. (2014). "A comparison of ballet dancers with different level of experience in performing single-leg stance on retiré position." Motor control **18**(2): 199-212.

López-Ortiz, C., et al. (2016). "Pilot study of a targeted dance class for physical rehabilitation in children with cerebral palsy." SAGE open medicine **4**: 2050312116670926.

Machacova, K., et al. (2017). "Dance as prevention of late life functional decline among nursing home residents." Journal of Applied Gerontology **36**(12): 1453-1470.

Mangeri, F., et al. (2014). "A standard ballroom and Latin dance program to improve fitness and adherence to physical activity in individuals with type 2 diabetes and in obesity." Diabetology & metabolic syndrome **6**(1): 74.

Marmeleira, J., et al. (2009). "Creative dance can enhance proprioception in older adults." Journal of sports medicine and physical fitness **49**(4): 480-485.

Matthews, B., et al. (2006). "Dancing for bone health: a 3-year longitudinal study of bone mineral accrual across puberty in female non-elite dancers and controls." Osteoporosis International **17**(7): 1043-1054.

Mavridis, G., et al. (2004). "The effect of a health-related aerobic dance program on elementary school children." Journal of Human Movement Studies **47**(4): 337-349.

Mavrovouniotis, F. H., et al. (2010). "Greek traditional dances and quality of old people's life." Journal of Bodywork and movement therapies **14**(3): 209-218.

McKinley, P., et al. (2008). "Effect of a community-based Argentine tango dance program on functional balance and confidence in older adults." Journal of aging and physical activity **16**(4): 435-453.

Merom, D., et al. (2016). "Cognitive benefits of social dancing and walking in old age: the dancing mind randomized controlled trial." Frontiers in aging neuroscience **8**: 26.

Merom, D., et al. (2016). "Social dancing and incidence of falls in older adults: a cluster randomised controlled trial." PLoS medicine **13**(8): e1002112.

Micallef, C. (2014). "The effectiveness of an 8-week Zumba programme for weight reduction in a group of Maltese overweight and obese women." Sport Sciences for Health **10**(3): 211-217.

Milburn, S. and N. K. Butts (1983). "A comparison of the training responses to aerobic dance and jogging in college females." Medicine and Science in Sports and Exercise **15**(6): 510-513.

Moller, A. and Y. Masharawi (2011). "The effect of first ballet classes in the community on various postural parameters in young girls." Physical therapy in sport **12**(4): 188-193.

Müller, P., et al. (2017). "Evolution of neuroplasticity in response to physical activity in old age: the case for dancing." Frontiers in aging neuroscience **9**: 56.

Ozdinc, S. A. and F. N. Turan (2016). "Effects of ballet training of children in Turkey on foot anthropometric measurements and medial longitudinal arc development." J. Pak. Med. Assoc **66**(7): 869-874.

Park, J.-H., et al. (2015). "Effects of a 12-week healthy-life exercise program on oxidized low-density lipoprotein cholesterol and carotid intima-media thickness in obese elderly women." Journal of physical therapy science **27**(5): 1435-1439.

Pérez, R. M., et al. (2014). "Visual availability, balance performance and movement complexity in dancers." Gait & posture **40**(4): 556-560.

Perrin, P., et al. (2002). "Judo, better than dance, develops sensorimotor adaptabilities involved in balance control." Gait & posture **15**(2): 187-194.

Pichierri, G., et al. (2012). "A cognitive-motor intervention using a dance video game to enhance foot placement accuracy and gait under dual task conditions in older adults: a randomized controlled trial." BMC geriatrics **12**(1): 74.

Pigeon, P., et al. (1997). "Intensive dance practice: repercussions on growth and puberty." The American journal of sports medicine **25**(2): 243-247.

Rani, S. and S. P. Sing (2013). "Effect of selected yogic practices and aerobic dance on health related physical fitness variables among Nicobari women students." Int J Phys Educ Fitness Sports **2**(4): 76-79.

Rehfeld, K., et al. (2017). "Dancing or fitness sport? The effects of two training programs on hippocampal plasticity and balance abilities in healthy seniors." Frontiers in human neuroscience **11**: 305.

Rein, S., et al. (2011). "Postural control and functional ankle stability in professional and amateur dancers." Clinical Neurophysiology **122**(8): 1602-1610.

Romenets, S. R., et al. (2015). "Tango for treatment of motor and non-motor manifestations in Parkinson's disease: a randomized control study." Complementary Therapies in Medicine **23**(2): 175-184.

Rossmeissl, A., et al. (2016). "ZumBeat: Evaluation of a Zumba dance intervention in postmenopausal overweight women." Sports **4**(1): 5.

Schmit, J. M., et al. (2005). "Dynamic patterns of postural sway in ballet dancers and track athletes." Experimental Brain Research **163**(3): 370-378.

Schmitt, H., et al. (2005). "Influence of professional dance training on peak torque and proprioception at the ankle." Clinical Journal of Sport Medicine **15**(5): 331-339.

Serrano-Guzmán, M., et al. (2016). "Effectiveness of a flamenco and sevillanas program to enhance mobility, balance, physical activity, blood pressure, body mass, and quality of life in postmenopausal women living in the community in Spain: a randomized clinical trial." Menopause **23**(9): 965-973.

Shimamoto, H., et al. (1998). "Low impact aerobic dance as a useful exercise mode for reducing body mass in mildly obese middle-aged women." Applied Human Science **17**(3): 109-114.

Simmons, R. W. (2005). "Neuromuscular responses of trained ballet dancers to postural perturbations." International Journal of Neuroscience **115**(8): 1193-1203.

Simmons, R. W. (2005). "Sensory organization determinants of postural stability in trained ballet dancers." International Journal of Neuroscience **115**(1): 87-97.

Sofianidis, G., et al. (2017). "A comparative study of the effects of pilates and Latin dance on static and dynamic balance in older adults." Journal of aging and physical activity **25**(3): 412-419.

Sofianidis, G., et al. (2009). "Effect of a 10-week traditional dance program on static and dynamic balance control in elderly adults." Journal of aging and physical activity **17**(2): 167-180.

Stalder, M. A., et al. (1990). "The effects of supplemental weight training for ballet dancers." The Journal of Strength & Conditioning Research **4**(3): 95-102.

Štalec, J., et al. (2007). "The impact of dance-aerobics training on the morpho-motor status in female high-schoolers." Collegium antropologicum **31**(1): 259-266.

Steinberg, N., et al. (2008). "Growth and development of female dancers aged 8–16 years." American Journal of Human Biology: The Official Journal of the Human Biology Association **20**(3): 299-307.

Thiesen, T. and A. Sumiya (2011). "Equilíbrio e arco plantar no balé clássico." Conscientiae Saúde **10**(1): 138-142.

Thullier, F. and H. Moufti (2004). "Multi-joint coordination in ballet dancers." Neuroscience letters **369**(1): 80-84.

Tsanaka, A., et al. (2017). "Effects of a modified ballet class on strength and jumping ability in college ballet dancers." Journal of dance medicine & science **21**(3): 97-101.

Verghese, J. (2006). "Cognitive and mobility profile of older social dancers." Journal of the American Geriatrics Society **54**(8): 1241-1244.

Volpe, D., et al. (2013). "A comparison of Irish set dancing and exercises for people with Parkinson's disease: a phase II feasibility study." BMC geriatrics **13**(1): 1-6.

Wallmann, H. W., et al. (2009). "The effect of a senior jazz dance class on static balance in healthy women over 50 years of age: a pilot study." Biological research for nursing **10**(3): 257-266.

White, M. K., et al. (1984). "The effects of exercise on the bones of postmenopausal women." International orthopaedics **7**(4): 209-214.

Wu, H. Y., et al. (2016). "Effects of low-impact dance on blood biochemistry, bone mineral density, the joint range of motion of lower extremities, knee extension torque, and fall in females." Journal of aging and physical activity **24**(1): 1-7.

Wu, H.-Y., et al. (2011). "The effects of low-impact dance on knee torque and lower extremity mobility in middle-aged and older women." Journal of Nursing Research **19**(4): 267-274.

Zhang, J.-G., et al. (2008). "Postural stability and physical performance in social dancers." Gait & posture **27**(4): 697-701.

Zhang, X., et al. (2012). "Study about effects of square dancing on the cognitive function and emotion of the aged." Chin Foreign Med Res **10**: 4-5.

**Included Individual Studies** *(from included reviews)* **– Music**

Anaya, E. M., et al. (2016). "Long-term musical experience and auditory and visual perceptual abilities under adverse conditions." The Journal of the Acoustical Society of America **140**(3): 2074-2081.

Annello, J. A. (1972). "A Comparison of Academic Achievement between Instrumental Music Students and Non-Music Students in the El Dorado and Valencia High Schools of the Placentia United School District." Doctoral Diss., Brigham Young University.

Bae, M. (2011). "Effect of group music therapy on student music therapists' anxiety, mood, job engagement, and self-efficacy." Univ Kansas **132**: 3458209.

Başkent, D. and E. Gaudrain (2016). "Musician advantage for speech-on-speech perception." The Journal of the Acoustical Society of America **139**(3): EL51-EL56.

Beck, R., et al. (2000). "Choral singing, performance perception, and immune system changes in salivary immunoglobulin A and cortisol." Music Perception **18**(1): 87-106.

Beck, R. J., et al. (2006). "Supporting the Health of College Solo Singers: The Relationship of Positive Emotions and Stress to Changes in Salivary IgA and Cortisol during Singing." Journal for Learning through the Arts **2**(1): 19.

Bengtsson, S. L., et al. (2007). "Cortical regions involved in the generation of musical structures during improvisation in pianists." Journal of cognitive neuroscience **19**(5): 830-842.

Berkowitz, A. L. and D. Ansari (2008). "Generation of novel motor sequences: the neural correlates of musical improvisation." Neuroimage **41**(2): 535-543.

Bialystok, E. and A.-M. DePape (2009). "Musical expertise, bilingualism, and executive functioning." Journal of Experimental Psychology: Human Perception and Performance **35**(2): 565.

Bidelman, G. M., et al. (2014). "Coordinated plasticity in brainstem and auditory cortex contributes to enhanced categorical speech perception in musicians." European Journal of Neuroscience **40**(4): 2662-2673.

Bilhartz, T. D., et al. (1999). "The effect of early music training on child cognitive development." Journal of Applied Developmental Psychology **20**(4): 615-636.

Bittman, B., et al. (2003). "Recreational music-making: a cost-effective group interdisciplinary strategy for reducing burnout and improving mood states in long-term care workers." Advances in Mind Body Medicine **19**(3/4): 4-15.

Bittman, B., et al. (2009). "Creative musical expression as a catalyst for quality-of-life improvement in inner-city adolescents placed in a court-referred residential treatment program." Advances in mind-body medicine **24**(1): 8-19.

Bittman, B. B., et al. (2001). "Composite effects of group drumming music therapy on modulation of neuroendocrine-immune parameters in normal subjects." Alternative therapies in health and medicine **7**(1): 38.

Bittman, B. B., et al. (2004). "Recreational music-making: an integrative group intervention for reducing burnout and improving mood states in first year associate degree nursing students: insights and economic impact." International Journal of Nursing Education Scholarship **1**(1).

Boebinger, D., et al. (2015). "Musicians and non-musicians are equally adept at perceiving masked speech." The Journal of the Acoustical Society of America **137**(1): 378-387.

Bolduc, J. and P. Lefebvre (2012). "Using nursery rhymes to foster phonological and musical processing skills in kindergarteners."

Bormann, J. E., et al. (2006). "Relationship of frequent mantram repetition to emotional and spiritual well-being in healthcare workers." The Journal of Continuing Education in Nursing **37**(5): 218-224.

Brandler, S. and T. H. Rammsayer (2003). "Differences in mental abilities between musicians and non-musicians." Psychology of music **31**(2): 123-138.

Brown, S., et al. (2006). "Music and language side by side in the brain: a PET study of the generation of melodies and sentences." European Journal of Neuroscience **23**(10): 2791-2803.

Callan, D. E., et al. (2006). "Song and speech: brain regions involved with perception and covert production." Neuroimage **31**(3): 1327-1342.

Catterall, J. S., et al. (1999). "Involvement in the arts and human development: General involvement and intensive involvement in music and theater arts." Champions of change: The impact of the arts on learning **1**: 1-18.

Chan, A. S., et al. (1998). "Music training improves verbal memory." Nature **396**(6707): 128-128.

Ciepluch, G. M. (1989). "Sightreading achievement in instrumental music performance, learning gifts, and academic achievement: A correlation study."

Clayton, K. K., et al. (2016). "Executive function, visual attention and the cocktail party problem in musicians and non-musicians." PloS one **11**(7): e0157638.

Coffey, E. B., et al. (2017). "Neural correlates of early sound encoding and their relationship to speech-in-noise perception." Frontiers in neuroscience **11**: 479.

Coffey, E. B., et al. (2016). "Cortical contributions to the auditory frequency-following response revealed by MEG." Nature communications **7**(1): 1-11.

Cogo-Moreira, H., et al. (2013). "Effectiveness of music education for the improvement of reading skills and academic achievement in young poor readers: a pragmatic cluster-randomized, controlled clinical trial." PloS one **8**(3): e59984.

Cohen, G. D., et al. (2006). "The impact of professionally conducted cultural programs on the physical health, mental health, and social functioning of older adults." The Gerontologist **46**(6): 726-734.

Cohen, M. L. (2009). "Choral singing and prison inmates: Influences of performing in a prison choir." Journal of Correctional Education (1974-) **60**(1): 52-65.

College Board (1988-1999). College bound seniors profile of SAT and achievement test takers.

Costa-Giomi, E. (1999). "The effects of three years of piano instruction on children's cognitive development." Journal of research in music education **47**(3): 198-212.

Costa-Giomi, E. (2004). "Effects of three years of piano instruction on children’s academic achievement, school performance and self-esteem." Psychology of music **32**(2): 139-152.

Coulton, S., et al. (2015). "Effectiveness and cost-effectiveness of community singing on mental health-related quality of life of older people: randomised controlled trial." The British Journal of Psychiatry **207**(3): 250-255.

de Manzano, Ö. and F. Ullén (2012). "Goal-independent mechanisms for free response generation: Creative and pseudo-random performance share neural substrates." Neuroimage **59**(1): 772-780.

Degé, F. and G. Schwarzer (2011). "The effect of a music program on phonological awareness in preschoolers." Frontiers in psychology **2**: 124.

Douglas, S. and P. Willatts (1994). "The relationship between musical ability and literacy skills." Journal of Research in reading **17**(2): 99-107.

Du, Y. and R. J. Zatorre (2017). "Musical training sharpens and bonds ears and tongue to hear speech better." Proceedings of the National Academy of Sciences **114**(51): 13579-13584.

Dunbar, R. I., et al. (2012). "Performance of music elevates pain threshold and positive affect: implications for the evolutionary function of music." Evolutionary Psychology **10**(4): 147470491201000403.

Engdahl, P. M. (1994). "The effect of pull-out programs on the academic achievement of sixth grade students in South Bend, Indiana."

Fetzer, L. (1996). "Facilitating print awareness and literacy development with familiar children's songs."

Flohr, J. W. (1996). "Children's Electrophysiological Responses to Music."

Flohr, J. W., et al. (1998). Quantitative EEG responses to music stimuli. Music Educators National Conference, Phoenix, Ariz.

Flohr, J. W., et al. (2000). "Brain Research Applied to Music Education." Music Educators National Conference.

Franklin, M. S., et al. (2008). "The effects of musical training on verbal memory." Psychology of music **36**(3): 353-365.

Friedman, B. (1959). An evaluation of the achievement in reading and arithmetic of pupils in elementary school instrumental music classes, New York University, School of Education.

Fuller, C. D., et al. (2014). "The musician effect: does it persist under degraded pitch conditions of cochlear implant simulations?" Frontiers in neuroscience **8**: 179.

George, E. M. and D. Coch (2011). "Music training and working memory: An ERP study." Neuropsychologia **49**(5): 1083-1094.

Gold, C., et al. (2014). "Music therapy for prisoners: Pilot randomised controlled trial and implications for evaluating psychosocial interventions." International journal of offender therapy and comparative criminology **58**(12): 1520-1539.

Grape, C., et al. (2002). "Does singing promote well-being?: An empirical study of professional and amateur singers during a singing lesson." Integrative Physiological & Behavioral Science **38**(1): 65-74.

Graziano, A. B., et al. (1999). "Enhanced learning of proportional math through music training and spatial-temporal training." Neurological Research **21**(2): 139-152.

Groff, F. H. (1963). Effect on academic achievement of excusing elementary school pupils from classes to study instrumental music, University of Connecticut.

Gromko, J. E. (2005). "The effect of music instruction on phonemic awareness in beginning readers." Journal of research in music education **53**(3): 199-209.

Gromko, J. E. and A. S. Poorman (1998). "The effect of music training on preschoolers' spatial-temporal task performance." Journal of research in music education **46**(2): 173-181.

Hansen, M., et al. (2013). "Working memory and musical competence of musicians and non-musicians." Psychology of music **41**(6): 779-793.

Helmbold, N., et al. (2005). "Differences in primary mental abilities between musicians and nonmusicians." Journal of Individual Differences **26**(2): 74-85.

Herrera, L., et al. (2011). "Effects of phonological and musical training on the reading readiness of native- and foreign-Spanish-speaking children." Psychology of music **39**(1): 68-81.

Hilliard, R. E. (2006). "The effect of music therapy sessions on compassion fatigue and team building of professional hospice caregivers." The Arts in Psychotherapy **33**(5): 395-401.

Ho, P., et al. (2011). "The impact of group drumming on social-emotional behavior in low-income children." Evidence-Based Complementary and Alternative Medicine **2011**.

Ho, Y.-C., et al. (2003). "Music training improves verbal but not visual memory: cross-sectional and longitudinal explorations in children." Neuropsychology **17**(3): 439.

Houston, D. M., et al. (1998). "Using humour to promote psychological wellbeing in residential homes for older people." Aging & Mental Health **2**(4): 328-332.

Huang, Z., et al. (2010). "Verbal memory retrieval engages visual cortex in musicians." Neuroscience **168**(1): 179-189.

Hurwitz, I., et al. (1975). "Nonmusicol effects of the kodaly music curriculum in primary grade children." Journal of learning Disabilities **8**(3): 167-174.

Jakobson, L. S., et al. (2008). "Memory for verbal and visual material in highly trained musicians." Music Perception **26**(1): 41-55.

Jeffries, K., et al. (2003). "Words in melody: an H215O PET study of brain activation during singing and speaking." Neuroreport **14**(5): 749-754.

Jentschke, S., et al. (2005). "Investigating the relationship of music and language in children: influences of musical training and language impairment." Annals of the New York Academy of Sciences **1060**(1): 231-242.

Kelley, L. L. (1981). "A combined experimental and descriptive study of the effect of music on reading and language."

Kennedy, R. (1998). The effects of musical performance, rational emotive therapy and vicarious experience on the self-efficacy and self-esteem of juvenile delinquents and disadvantaged children, University of Kansas, Music and Dance.

Kleber, B., et al. (2007). "Overt and imagined singing of an Italian aria." Neuroimage **36**(3): 889-900.

Kleber, B., et al. (2010). "The brain of opera singers: experience-dependent changes in functional activation." Cerebral Cortex **20**(5): 1144-1152.

Koelsch, S., et al. (2010). "Music in the treatment of affective disorders: an exploratory investigation of a new method for music-therapeutic research." Music Perception **27**(4): 307-316.

Koyama, M., et al. (2009). "Recreational music-making modulates immunological responses and mood states in older adults." Journal of medical and dental sciences **56**(2): 79-90.

Kreutz, G. (2014). "Does singing facilitate social bonding." Music Med **6**(2): 51-60.

Kreutz, G., et al. (2004). "Effects of choir singing or listening on secretory immunoglobulin A, cortisol, and emotional state." Journal of behavioral medicine **27**(6): 623-635.

Kuhn, D. (2002). "The effects of active and passive participation in musical activity on the immune system as measured by salivary immunoglobulin A (SlgA)." Journal of music therapy **39**(1): 30-39.

Kvet, E. J. (1985). "Excusing elementary school students from regular classroom activities for the study of instrumental music: The effect on sixth-grade reading, language, and mathematics achievement." Journal of research in music education **33**(1): 45-54.

Laczo, Z. (1985). "The nonmusical outcomes of music education: Influence on intelligence?" Bulletin of the Council for Research in Music Education: 109-118.

Lamar, H. B. (1989). An examination of the congruency of music aptitude scores and mathematics and reading achievement scores of elementary children, University of Southern Mississippi.

Lee, K. M., et al. (2009). "Selective subcortical enhancement of musical intervals in musicians." Journal of Neuroscience **29**(18): 5832-5840.

Lee, Y.-s., et al. (2007). "Effects of skill training on working memory capacity." Learning and Instruction **17**(3): 336-344.

Limb, C. J. and A. R. Braun (2008). "Neural substrates of spontaneous musical performance: An fMRI study of jazz improvisation." PloS one **3**(2): e1679.

Lindblad, F., et al. (2007). "Music intervention for 5th and 6th graders—effects on development and cortisol secretion." Stress and Health: Journal of the International Society for the Investigation of Stress **23**(1): 9-14.

Mallory, M. E. and K. E. Philbrick (1995). "Music training and spatial skills in preschool children." America Psychological Association of Music, New York.

McCarthy, K. J. (1992). "Music Performance Group Membership and Academic Success: A Descriptive Study of One 4-year High School." Colorado Music Educators Association.

Monahan, C. B., et al. (1987). "The effect of melodic and temporal contour on recognition memory for pitch change." Perception & Psychophysics **41**(6): 576-600.

Moreno, S., et al. (2011). "Effect of music training on promoting preliteracy skills: Preliminary causal evidence." Music Perception **29**(2): 165-172.

Moreno, S., et al. (2009). "Musical training influences linguistic abilities in 8-year-old children: more evidence for brain plasticity." Cerebral Cortex **19**(3): 712-723.

Moritz, C., et al. (2013). "Links between early rhythm skills, musical training, and phonological awareness." Reading and Writing **26**(5): 739-769.

Mungas, R. and M. J. Silverman (2014). "Immediate effects of group-based wellness drumming on affective states in university students." The Arts in Psychotherapy **41**(3): 287-292.

Musacchia, G., et al. (2007). "Musicians have enhanced subcortical auditory and audiovisual processing of speech and music." Proceedings of the National Academy of Sciences **104**(40): 15894-15898.

Musacchia, G., et al. (2008). "Relationships between behavior, brainstem and cortical encoding of seen and heard speech in musicians and non-musicians." Hearing research **241**(1-2): 34-42.

Neufeld, K. A. (1986). "Understanding of selected pre-number concepts: Relationships to a formal music program." Alberta Journal of Educational Research.

Okhrei, A., et al. (2016). "Performance of working memory of musicians and non-musicians in tests with letters, digits, and geometrical shapes." Biologija **62**(4).

Olanoff, M. and L. Kirschner (1969). Musical Ability Utilization Program. Final Report, ERIC Clearinghouse.

Oxenham, A. J., et al. (2003). "Informational masking and musical training." The Journal of the Acoustical Society of America **114**(3): 1543-1549.

Pai, I., et al. (2008). "The effect of singing on snoring and daytime somnolence." Sleep and Breathing **12**(3): 265-268.

Pallesen, K. J., et al. (2010). "Cognitive control in auditory working memory is enhanced in musicians." PloS one **5**(6): e11120.

Parbery-Clark, A., et al. (2012). "Musical experience offsets age-related delays in neural timing." Neurobiology of aging **33**(7): 1483. e1481-1483. e1484.

Parbery-Clark, A., et al. (2012). "Musical experience strengthens the neural representation of sounds important for communication in middle-aged adults." Frontiers in aging neuroscience **4**: 30.

Parbery-Clark, A., et al. (2009). "Musician enhancement for speech-in-noise." Ear and hearing **30**(6): 653-661.

Parbery-Clark, A., et al. (2011). "Musical experience and the aging auditory system: implications for cognitive abilities and hearing speech in noise." PloS one **6**(5): e18082.

Parbery-Clark, A., et al. (2011). "Context-dependent encoding in the auditory brainstem subserves enhanced speech-in-noise perception in musicians." Neuropsychologia **49**(12): 3338-3345.

Parente, J. A. and J. J. O'Malley (1975). "Training in musical rhythm and field dependence of children." Perceptual and motor skills **40**(2): 392-394.

Perkins, R. and A. Williamon (2014). "Learning to make music in older adulthood: A mixed-methods exploration of impacts on wellbeing." Psychology of music **42**(4): 550-567.

Perry, D. W., et al. (1999). "Localization of cerebral activity during simple singing." Neuroreport **10**(16): 3453-3458.

Piro, J. M. and C. Ortiz (2009). "The effect of piano lessons on the vocabulary and verbal sequencing skills of primary grade students." Psychology of music **37**(3): 325-347.

Portowitz, A., et al. (2009). "Underlying mechanisms linking music education and cognitive modifiability." Research Studies in Music Education **31**(2): 107-128.

Ramachandra, V., et al. (2012). "The Impact of Musical Training on the Phonological Memory and the Central Executive: A Brief Report." North American Journal of Psychology **14**(3).

Rauscher, F., et al. (1997). "Music training causes long-term enhancement of preschool children's spatial–temporal reasoning." Neurological Research **19**(1): 2-8.

Rauscher, F. H. (1994). "Music and Spatial Task Performance: A Causal Relationship."

Rauscher, F. H. (1999). Music, cognitive development, and the classroom: Franklin Elementary. American Orff-Schulwerk Association National Conference, Phoenix, Ariz.

Rauscher, F. H. and M. A. Zupan (2000). "Classroom keyboard instruction improves kindergarten children's spatial-temporal performance: A field experiment." Early childhood research quarterly **15**(2): 215-228.

Register, D. (2001). "The effects of an early intervention music curriculum on prereading/writing." Journal of music therapy **38**(3): 239-248.

Register, D. (2004). "The effects of live music groups versus an educational children's television program on the emergent literacy of young children." Journal of music therapy **41**(1): 2-27.

Rickard, N. S., et al. (2012). "Absence of widespread psychosocial and cognitive effects of school-based music instruction in 10–13-year-old students." International Journal of Music Education **30**(1): 57-78.

Roberts, D. L. (1978). An experimental study of the relationship between musical note-reading and language reading, University of Missouri-Kansas City, Conservatory of Music.

Rodrigues, A. C., et al. (2014). "Visual memory in musicians and non-musicians." Frontiers in human neuroscience **8**: 424.

Roskam, K. (1979). "Music therapy as an aid for increasing auditory awareness and improving reading skill." Journal of music therapy **16**(1): 31-42.

Ruggles, D. R., et al. (2014). "Influence of musical training on understanding voiced and whispered speech in noise." PloS one **9**(1): e86980.

Ruiz, M. H., et al. (2009). "Detecting wrong notes in advance: neuronal correlates of error monitoring in pianists." Cerebral Cortex **19**(11): 2625-2639.

Sandgren, M. (2009). Evidence for strong immediate well-being effects of choral singing–With more enjoyment for women than for men. ESCOM 2009: 7th Triennial Conference of European Society for the Cognitive Sciences of Music.

Schellenberg, E. G. (2004). "Music lessons enhance IQ." Psychological science **15**(8): 511-514.

Schiavio, A. and R. Timmers (2016). "Motor and audiovisual learning consolidate auditory memory of tonally ambiguous melodies." Music Perception: An Interdisciplinary Journal **34**(1): 21-32.

Schorr-Lesnick, B., et al. (1985). "Pulmonary function in singers and wind-instrument players." Chest **88**(2): 201-205.

Schulze, K., et al. (2011). "Working memory for tonal and atonal sequences during a forward and a backward recognition task." Music Perception **29**(3): 255-267.

Schulze, K., et al. (2011). "Neural correlates of strategy use during auditory working memory in musicians and non-musicians." European Journal of Neuroscience **33**(1): 189-196.

Schulze, K., et al. (2011). "Neuroarchitecture of verbal and tonal working memory in nonmusicians and musicians." Human brain mapping **32**(5): 771-783.

Slater, J. and N. Kraus (2016). "The role of rhythm in perceiving speech in noise: a comparison of percussionists, vocalists and non-musicians." Cognitive processing **17**(1): 79-87.

Slater, J., et al. (2015). "Music training improves speech-in-noise perception: Longitudinal evidence from a community-based music program." Behavioural brain research **291**: 244-252.

Smith, C., et al. (2014). "African drumming: a holistic approach to reducing stress and improving health?" Journal of Cardiovascular Medicine **15**(6): 441-446.

Steurer, M., et al. (1998). "Does choir singing cause noise-induced hearing loss?" Audiology **37**(1): 38-51.

Strait, D. L. and N. Kraus (2011). "Can you hear me now? Musical training shapes functional brain networks for selective auditory attention and hearing speech in noise." Frontiers in psychology **2**: 113.

Strait, D. L., et al. (2010). "Musical experience shapes top-down auditory mechanisms: evidence from masking and auditory attention performance." Hearing research **261**(1-2): 22-29.

Suárez, L., et al. (2016). "Cross-sectional study on the relationship between music training and working memory in adults." Australian Journal of Psychology **68**(1): 38-46.

Swaminathan, J., et al. (2015). "Musical training, individual differences and the cocktail party problem." Scientific reports **5**: 11628.

Taetle, L. D. (1999). "The effect of active and passive music instruction on the spatial ability of kindergarten children."

Talamini, F., et al. (2016). "The working memory of musicians and nonmusicians." Music Perception: An Interdisciplinary Journal **34**(2): 183-191.

Taylor, A. C. and S. A. Dewhurst (2017). "Investigating the influence of music training on verbal memory." Psychology of music **45**(6): 814-820.

Thomson, J. M., et al. (2013). "Auditory processing interventions and developmental dyslexia: a comparison of phonemic and rhythmic approaches." Reading and Writing **26**(2): 139-161.

Tierney, A. T., et al. (2015). "Music training alters the course of adolescent auditory development." Proceedings of the National Academy of Sciences **112**(32): 10062-10067.

Tsang, C. D. and N. J. Conrad (2011). "Music training and reading readiness." Music Perception **29**(2): 157-163.

Unwin, M. M., et al. (2002). "The effects of group singing on mood." Psychology of music **30**(2): 175-185.

Valentine, E. and C. Evans (2001). "The effects of solo singing, choral singing and swimming on mood and physiological indices." British Journal of Medical Psychology **74**(1): 115-120.

Varnet, L., et al. (2015). "How musical expertise shapes speech perception: evidence from auditory classification images." Scientific reports **5**: 14489.

Vasuki, P. R. M., et al. (2016). "Musicians' edge: A comparison of auditory processing, cognitive abilities and statistical learning." Hearing research **342**: 112-123.

Vogt, S., et al. (2007). "Prefrontal involvement in imitation learning of hand actions: effects of practice and expertise." Neuroimage **37**(4): 1371-1383.

Wachi, M., et al. (2007). "Recreational music-making modulates natural killer cell activity, cytokines, and mood states in corporate employees." Medical Science Monitor **13**(2): CR57-CR70.

Weeden, R. E. (1972). A comparison of the academic achievement in reading and mathematics of Negro children whose parents are interested, not interested, or involved in a program of Suzuki violin, ProQuest Information & Learning.

Weiss, A. H., et al. (2014). "Spatial vision is superior in musicians when memory plays a role." Journal of vision **14**(9): 18-18.

Wheeler, L. R. and V. D. Wheeler (1951). "The intelligence of music students." Journal of Educational psychology **42**(4): 223.

Wise, G. W., et al. (1992). "Exploration of the relationship between choral singing and successful aging." Psychological Reports **70**(3_suppl): 1175-1183.

Yazejian, N. and E. S. Peisner-Feinberg (2009). "Effects of a preschool music and movement curriculum on children's language skills." NHSA dialog **12**(4): 327-341.

Zarate, J. M., et al. (2010). "Neural networks involved in voluntary and involuntary vocal pitch regulation in experienced singers." Neuropsychologia **48**(2): 607-618.

Zarate, J. M. and R. J. Zatorre (2008). "Experience-dependent neural substrates involved in vocal pitch regulation during singing." Neuroimage **40**(4): 1871-1887.

Zendel, B. R. and C. Alain (2012). "Musicians experience less age-related decline in central auditory processing." Psychology and aging **27**(2): 410.

Zendel, B. R., et al. (2015). "The impact of musicianship on the cortical mechanisms related to separating speech from background noise." Journal of cognitive neuroscience **27**(5): 1044-1059.

Zuk, J., et al. (2014). "Behavioral and neural correlates of executive functioning in musicians and non-musicians." PloS one **9**(6): e99868.

Zulauf, M. (1993). "Three-year experiment in extended music teaching in Switzerland: The different effects observed in a group of French-speaking pupils." Bulletin of the Council for Research in Music Education: 111-121.

**<u>Excluded Individual Studies</u>** *<u>(from included reviews)</u>* **<u>– Dance</u>**

Ambegaonkar, J. P., et al. (2016). "Lower extremity hypermobility, but not core muscle endurance influences balance in female collegiate dancers." <u>International journal of sports physical therapy</u> **11**(2): 220.

Annino, G., et al. (2007). "Effect of whole body vibration training on lower limb performance in selected high-level ballet students." <u>The Journal of Strength & Conditioning Research</u> **21**(4): 1072-1076.

Arzoglou, D., et al. (2013). "The effect of [alpha] tradinional dance training program on neuromuscular coordination of individuals with autism." <u>Journal of Physical Education and Sport</u> **13**(4): 563.

Barcellos, C. and I. LA (2002). "Alterações posturais e do equilíbrio corporal na primeira posição em ponta do balé clássico." <u>Rev Paul Educ Fís</u> **16**(1): 43-52.

Batson, G. (2010). "Validating a dance-specific screening test for balance: preliminary results from multisite testing." <u>Medical Problems of Performing Artists</u> **25**(3): 110-115.

Belardinelli, R., et al. (2008). "Waltz dancing in patients with chronic heart failure: new form of exercise training." <u>Circulation: Heart Failure</u> **1**(2): 107-114.

Bruyneel, A., et al. (2010). "Organization of postural equilibrium in several planes in ballet dancers." <u>Neuroscience letters</u> **485**(3): 228-232.

Candelario-Gorbea, C., et al. (2014). "Effectiveness of a PNF intervention on active and passive range of motion during "developpe a la second" in ballet dancers." <u>Orthopaedic Physical Therapy Practice</u> **26**(3): 198-202.

Caplan, N. and A. S. C. Gibson (2011). "Single leg postural sway characteristics of dancers during a rotating task: a pilot study." <u>Arts Biomechanics</u> **1**(1): 45.

Cheng, H. S., et al. (2011). "Preliminary results of dancing exercise on postural stability in adolescent females." <u>The Kaohsiung journal of medical sciences</u> **27**(12): 566-572.

Clark, T. and E. Redding (2012). "The relationship between postural stability and dancer's past and future lower-limb injuries." <u>Medical Problems of Performing Artists</u> **27**(4): 197-204.

Cloak, R., et al. (2010). "Vibration training improves balance in unstable ankles." <u>International journal of sports medicine</u> **31**(12): 894.

Coutts, R. A., et al. (2006). "Development and assessment of an incremental fatigue protocol for contemporary dance." <u>Medical Problems of Performing Artists</u> **21**(2): 65-70.

da Costa, P. H. L., et al. (2013). "Single leg balancing in ballet: Effects of shoe conditions and poses." <u>Gait & posture</u> **37**(3): 419-423.

Da Silva, C. C., et al. (2015). "The effects of 17 weeks of ballet training on the autonomic modulation, hormonal and general biochemical profile of female adolescents." <u>Journal of human kinetics</u> **47**(1): 61-71.

Denardi, R. A., et al. (2008). "Informação visual e controlo postural durante a execução da pirouette no ballet." <u>Revista Portuguesa de Ciências do Desporto</u> **8**(2): 241-250.

Doi, T., et al. (2017). "Effects of cognitive leisure activity on cognition in mild cognitive impairment: results of a randomized controlled trial." Journal of the American Medical Directors Association **18**(8): 686-691.

Duncan, R. P. and G. M. Earhart (2014). "Are the effects of community-based dance on Parkinson disease severity, balance, and functional mobility reduced with time? A 2-year prospective pilot study." The Journal of Alternative and Complementary Medicine **20**(10): 757-763.

Engels, H.-J., et al. (1998). "Effects of low-impact, moderate-intensity exercise training with and without wrist weights on functional capacities and mood states in older adults." Gerontology **44**(4): 239-244.

Golomer, E., et al. (1997). "Sex-linked differences in equilibrium reactions among adolescents performing complex sensorimotor tasks." Journal of Physiology-Paris **91**(2): 49-55.

Golomer, E., et al. (1999). "The contribution of vision in dynamic spontaneous sways of male classical dancers according to student or professional level." Journal of Physiology-Paris **93**(3): 233-237.

Golomer, E. M., et al. (2009). "Influence of vision and motor imagery styles on equilibrium control during whole-body rotations." Somatosensory & motor research **26**(4): 105-110.

Hackney, M. E. and G. M. Earhart (2009). "Effects of dance on movement control in Parkinson's disease: a comparison of Argentine tango and American ballroom." Journal of rehabilitation medicine **41**(6): 475-481.

Hackney, M. E. and G. M. Earhart (2009). "Health-related quality of life and alternative forms of exercise in Parkinson disease." Parkinsonism & related disorders **15**(9): 644-648.

Hackney, M. E. and G. M. Earhart (2009). "Short duration, intensive tango dancing for Parkinson disease: an uncontrolled pilot study." Complementary Therapies in Medicine **17**(4): 203-207.

Hackney, M. E., et al. (2013). "Dancing for balance: feasibility and efficacy in oldest-old adults with visual impairment." Nursing research **62**(2): 138-143.

Hackney, M. E., et al. (2007). "A study on the effects of Argentine tango as a form of partnered dance for those with Parkinson disease and the healthy elderly." American Journal of Dance Therapy **29**(2): 109-127.

Holmerová, I., et al. (2010). "Effect of the exercise dance for seniors (EXDASE) program on lower-body functioning among institutionalized older adults." Journal of aging and health **22**(1): 106-119.

Hopkins, D. R., et al. (1990). "Effect of low-impact aerobic dance on the functional fitness of elderly women." The Gerontologist **30**(2): 189-192.

Jeon, M. Y., et al. (2005). "The effects of a Korean traditional dance movement program in elderly women." Journal of Korean Academy of Nursing **35**(7): 1268-1276.

Jeon, M. Y., et al. (2000). "Effect of Korean traditional dance movement training on balance, gait and leg strength in home bound elderly women." Journal of Korean Academy of Nursing **30**(3): 647-658.

Kaltsatou, A. C., et al. (2014). "Functional and psychosocial effects of either a traditional dancing or a formal exercising training program in patients with chronic heart failure: a comparative randomized controlled study." Clinical Rehabilitation **28**(2): 128-138.

Kattenstroth, J.-C., et al. (2013). "Six months of dance intervention enhances postural, sensorimotor, and cognitive performance in elderly without affecting cardio-respiratory functions." Frontiers in aging neuroscience **5**: 5.

Khan, K. M., et al. (2000). "Can 16–18-year-old elite ballet dancers improve their hip and ankle range of motion over a 12-month period?" Clinical Journal of Sport Medicine **10**(2): 98-103.

Kim, C.-G., et al. (2003). "Effects of a health-promotion program on cardiovascular risk factors, health behaviors, and life satisfaction in institutionalized elderly women." International journal of nursing studies **40**(4): 375-381.

Kim, H.-S., et al. (2007). "Effects of nutrition education and exercise programs on perceived dietary behaviors, food intake and serum lipid profiles in elderly Korean women living in residential homes." Asian Nursing Research **1**(1): 35-47.

Kim, S.-H., et al. (2011). "Effect of dance exercise on cognitive function in elderly patients with metabolic syndrome: a pilot study." Journal of sports science & Medicine **10**(4): 671.

Kirkendall, D., et al. (1984). "Isokinetic characteristics of ballet dancers and the response to a season of ballet training." Journal of Orthopaedic & Sports Physical Therapy **5**(4): 207-211.

Koutedakis, Y., et al. (1996). "The effects of strength training in male ballet dancers." Impulse **4**(3): 210-219.

Koutedakis, Y. and N. C. Sharp (2004). "Thigh-muscles strength training, dance exercise, dynamometry, and anthropometry in professional ballerinas." Journal of strength and conditioning research **18**(4): 714-718.

Krampe, J. (2013). "Exploring the effects of dance-based therapy on balance and mobility in older adults." Western journal of nursing research **35**(1): 39-56.

Krampe, J., et al. (2014). "Does dance-based therapy increase gait speed in older adults with chronic lower extremity pain: A feasibility study." Geriatric Nursing **35**(5): 339-344.

Li, Z.-t., et al. (2014). "Biomechanical research on the balance control of pirouettes in classical dance." Journal of Beijing Sport University **1**.

Liederbach, M., et al. (1992). "Monitoring training status in professional ballet dancers." The Journal of sports medicine and physical fitness **32**(2): 187-195.

Lim, S.-T., et al. (2015). "Effects of a healthy life exercise program on arteriosclerosis adhesion molecules in elderly obese women." Journal of physical therapy science **27**(5): 1529-1532.

Lin, C.-F., et al. (2005). "Ankle biomechanics of ballet dancers in relevé en pointé dance." Research in Sports Medicine **13**(1): 23-35.

Lin, C.-W., et al. (2014). "Differences of ballet turns (pirouette) performance between experienced and novice ballet dancers." Research quarterly for exercise and sport **85**(3): 330-340.

López-Ortiz, C., et al. (2012). "Dance program for physical rehabilitation and participation in children with cerebral palsy." Arts & Health **4**(1): 39-54.

Marín, G. H., et al. (2009). "Evaluation of the state intervention project to improve quality of life and reduce the complications associated with aging:" Add health to your years"." Gaceta sanitaria **23**(4): 272-277.

McGill, A., et al. (2019). "Effects of a ballet-based dance intervention on gait variability and balance confidence of people with Parkinson's." Arts & Health **11**(2): 133-146.

McKee, K. E. and M. E. Hackney (2013). "The effects of adapted tango on spatial cognition and disease severity in Parkinson's disease." Journal of motor behavior **45**(6): 519-529.

McMillan, A., et al. (1998). "The effect of Pilates-based training on dancers' dynamic posture." Journal of dance medicine & science **2**(3): 101-107.

Mertz, L. and C. Docherty (2012). "Self-described differences between legs in ballet dancers: do they relate to postural stability and ground reaction force measures?" Journal of dance medicine & science **16**(4): 154-160.

Micheli, L. J., et al. (2005). "Preseason to postseason changes in body composition of professional ballet dancers." Journal of dance medicine & science **9**(2): 56-59.

Morrin, N. and E. Redding (2013). "Acute effects of warm-up stretch protocols on balance, vertical jump height, and range of motion in dancers." Journal of dance medicine & science **17**(1): 34-40.

Müller, P., et al. (2017). "Evolution of neuroplasticity in response to physical activity in old age: the case for dancing." Frontiers in aging neuroscience **9**: 56.

Murrock, C. J. and C. H. Graor (2014). "Effects of dance on depression, physical function, and disability in underserved adults." Journal of aging and physical activity **22**(3): 380-385.

Notarnicola, A., et al. (2014). "Effect of teaching with or without mirror on balance in young female ballet students." BMC research notes **7**(1): 426.

Pappas, E., et al. (2011). "Time to stability differences between male and female dancers after landing from a jump on flat and inclined floors." Clinical Journal of Sport Medicine **21**(4): 325-329.

Rehfeld, K., et al. (2018). "Dance training is superior to repetitive physical exercise in inducing brain plasticity in the elderly." PloS one **13**(7): e0196636.

Romenets, S. R., et al. (2015). "Tango for treatment of motor and non-motor manifestations in Parkinson's disease: a randomized control study." Complementary Therapies in Medicine **23**(2): 175-184.

Scheidler, A. M., et al. (2018). "Targeted ballet program mitigates ataxia and improves balance in females with mild-to-moderate multiple sclerosis." PloS one **13**(10): e0205382.

Shigematsu, R., et al. (2002). "Dance-based aerobic exercise may improve indices of falling risk in older women." Age and ageing **31**(4): 261-266.

Silvestri, L. (1986). "Effects of aerobic dance and progressive relaxation on improving physical fitness of high school girls." Perceptual and motor skills **63**(1): 131-135.

Song, R., et al. (2004). "Comparisons of motivation, health behaviors, and functional status among elders in residential homes in Korea." Public Health Nursing **21**(4): 361-371.

Uusi-Rasi, K., et al. (1999). "Long-term recreational gymnastics, estrogen use, and selected risk factors for osteoporotic fractures." Journal of Bone and Mineral Research **14**(7): 1231-1238.

Ventura, M. I., et al. (2016). "A pilot study to evaluate multi-dimensional effects of dance for people with Parkinson's disease." Contemporary clinical trials **51**: 50-55.

Volpe, D., et al. (2013). "A comparison of Irish set dancing and exercises for people with Parkinson's disease: a phase II feasibility study." BMC geriatrics **13**(1): 1-6.

Wyon, M. A., et al. (2013). "Effect of midsole thickness of dance shoes on dynamic postural stability." Medical Problems of Performing Artists **28**(4): 195-198.

Young, C., et al. (2007). "Simple, novel physical activity maintains proximal femur bone mineral density, and improves muscle strength and balance in sedentary, postmenopausal Caucasian women." Osteoporosis International **18**(10): 1379-1387.

Zaferiou, A. M., et al. (2016). "Whole-body balance regulation during the turn phase of Pique and Pirouette turns with varied rotational demands." Medical Problems of Performing Artists **31**(2): 96-103.

**Excluded Individual Studies** *(from included reviews)* – **Music**

Aloui, A., et al. (2015). "Listening to music during warming-up counteracts the negative effects of ramadan observance on short-term maximal performance." PloS one **10**(8): e0136400.

Anderson, K. and K. Overy (2010). "Engaging Scottish young offenders in education through music and art." International Journal of Community Music **3**(1): 47-64.

Anderson, L. W. and C. U. Gustavson (2016). "The impact of a knitting intervention on compassion fatigue in oncology nurses." Clin J Oncol Nurs **20**(1): 102-104.

Baker, S. and S. Homan (2007). "Rap, recidivism and the creative self: A popular music programme for young offenders in detention." Journal of Youth Studies **10**(4): 459-476.

Baldari, C., et al. (2010). "Effects of music during exercise in different training status." Journal of sports medicine and physical fitness **50**(3): 281.

Ball, T., et al. (2007). "Response properties of human amygdala subregions: evidence based on functional MRI combined with probabilistic anatomical maps." PloS one **2**(3): e307.

Baumgartner, T., et al. (2006). "The emotional power of music: how music enhances the feeling of affective pictures." Brain research **1075**(1): 151-164.

Beck, B. D., et al. (2015). "Coping with work-related stress through Guided Imagery and Music (GIM): Randomized controlled trial." Journal of music therapy **52**(3): 323-352.

Bekiroğlu, T., et al. (2013). "Effect of Turkish classical music on blood pressure: a randomized controlled trial in hypertensive elderly patients." Complementary Therapies in Medicine **21**(3): 147-154.

Bensimon, M., et al. (2015). "The impact of relaxing music on prisoners' levels of anxiety and anger." International journal of offender therapy and comparative criminology **59**(4): 406-423.

Bensimon, M. and A. Gilboa (2010). "The music of my life: The impact of the musical presentation on the sense of purpose in life and on self-consciousness." The Arts in Psychotherapy **37**(3): 172-178.

Berns, G. S., et al. (2010). "Neural mechanisms of the influence of popularity on adolescent ratings of music." Neuroimage **49**(3): 2687-2696.

Blood, A. J. and R. J. Zatorre (2001). "Intensely pleasurable responses to music correlate with activity in brain regions implicated in reward and emotion." Proceedings of the National Academy of Sciences **98**(20): 11818-11823.

Boothby, D. M. and S. J. Robbins (2011). "The effects of music listening and art production on negative mood: A randomized, controlled trial." The Arts in Psychotherapy **38**(3): 204-208.

Brooks, D. M., et al. (2010). "Creative approaches for reducing burnout in medical personnel." The Arts in Psychotherapy **37**(3): 255-263.

Brown, S. and M. J. Martinez (2007). "Activation of premotor vocal areas during musical discrimination." Brain and cognition **63**(1): 59-69.

Brown, S., et al. (2004). "Passive music listening spontaneously engages limbic and paralimbic systems." Neuroreport **15**(13): 2033-2037.

Bungay, H. and A. Skingley (2008). The Silver Song Club Project: Summary of a Formative Evaluation. Canterbury Christ Church University. Viitattu 12.1. 2011.

Burns, J. L., et al. (2002). "The effects of different types of music on perceived and physiological measures of stress." Journal of music therapy **39**(2): 101-116.

Bygren, L. O., et al. (2009). "Cultural participation and health: a randomized controlled trial among medical care staff." Psychosomatic Medicine **71**(4): 469-473.

Cai, G., et al. (2001). "Clinical observation of music therapy combined with anti-tumor drugs in treating 116 cases of tumor patients." Zhongguo Zhong xi yi jie he za zhi Zhongguo Zhongxiyi Jiehe Zazhi= Chinese Journal of Integrated Traditional and Western Medicine **21**(12): 891-894.

Campion, M. and L. Levita (2014). "Enhancing positive affect and divergent thinking abilities: Play some music and dance." The Journal of Positive Psychology **9**(2): 137-145.

Carissoli, C., et al. (2015). "Does a meditation protocol supported by a mobile application help people reduce stress? Suggestions from a controlled pragmatic trial." Cyberpsychology, Behavior, and Social Networking **18**(1): 46-53.

Chan, M. F., et al. (2010). "Effects of music on depression and sleep quality in elderly people: A randomised controlled trial." Complementary Therapies in Medicine **18**(3-4): 150-159.

Chan, M. F., et al. (2012). "Effects of music on depression in older people: a randomised controlled trial." Journal of Clinical Nursing **21**(5-6): 776-783.

Chang, F. Y., et al. (2010). "The effect of a music programme during lunchtime on the problem behaviour of the older residents with dementia at an institution in Taiwan." Journal of Clinical Nursing **19**(7-8): 939-948.

Chang, H.-C., et al. (2015). "The effects of music listening on psychosocial stress and maternal–fetal attachment during pregnancy." Complementary Therapies in Medicine **23**(4): 509-515.

Chang, M. Y., et al. (2008). "Effects of music therapy on psychological health of women during pregnancy." Journal of Clinical Nursing **17**(19): 2580-2587.

Chen, C. J., et al. (2015). "The effects of C hinese five-element music therapy on nursing students with depressed mood." International Journal of Nursing Practice **21**(2): 192-199.

Clift, S. and G. Hancox (2010). "The significance of choral singing for sustaining psychological wellbeing: findings from a survey of choristers in England, Australia and Germany." Music Performance Research **3**.

Clift, S., et al. (2007). Choral singing and psychological wellbeing: Findings from English choirs in a cross-national survey using the WHOQOL-BREF. Proceedings of the international symposium on performance science.

Clift, S., et al. (2009). What do Singers Say About the Effects of Choral Singing on Physical Health?-Findings from a Survey of Choristers in Australia, England and Germany. ESCOM 2009: 7th Triennial Conference of European Society for the Cognitive Sciences of Music.

Clift, S., et al. (2010). "Choral singing and psychological wellbeing: Quantitative and qualitative findings from English choirs in a cross-national survey." Journal of Applied Arts & Health **1**(1): 19-34.

Clift, S. M. and G. Hancox (2001). "The perceived benefits of singing: findings from preliminary surveys of a university college choral society." The journal of the Royal Society for the Promotion of Health **121**(4): 248-256.

Clift, S. M., et al. (2008). "Singing and health: A systematic mapping and review of non-clinical research."

Coen, S. J., et al. (2009). "Negative mood affects brain processing of visceral sensation." Gastroenterology **137**(1): 253-261. e252.

Cohen, G. D., et al. (2006). "The impact of professionally conducted cultural programs on the physical health, mental health, and social functioning of older adults." The Gerontologist **46**(6): 726-734.

Cohen, M. L. (2007). "Explorations of inmate and volunteer choral experiences in a prison-based choir." Australian Journal of Music Education(1): 61.

Cooke, M. L., et al. (2010). "A randomized controlled trial exploring the effect of music on agitated behaviours and anxiety in older people with dementia." Aging and mental health **14**(8): 905-916.

Courey, S. J., et al. (2012). "Academic music: music instruction to engage third-grade students in learning basic fraction concepts." Educational studies in mathematics **81**(2): 251-278.

De Roeper, J. and H. J. Savelsberg (2009). "Challenging the youth policy imperative: engaging young people through the arts." Journal of Youth Studies **12**(2): 209-225.

DeCarlo, A. and E. Hockman (2004). "RAP therapy: A group work intervention method for urban adolescents." Social work with Groups **26**(3): 45-59.

DuRousseau, D. R., et al. (2011). "Operational study to evaluate music-based neurotraining at improving sleep quality, mood, and daytime function in a first responder population." Journal of Neurotherapy **15**(4): 389-398.

Dyck, M., et al. (2011). "Cognitive versus automatic mechanisms of mood induction differentially activate left and right amygdala." Neuroimage **54**(3): 2503-2513.

Eldar, E., et al. (2007). "Feeling the real world: limbic response to music depends on related content." Cerebral Cortex **17**(12): 2828-2840.

Fechir, M., et al. (2008). "Patterns of sympathetic responses induced by different stress tasks." The open neurology journal **2**: 25.

Field, T., et al. (1997). "Job stress reduction therapies." Alternative therapies in health and medicine **3**(4): 54-56.

Flores-Gutiérrez, E. O., et al. (2007). "Metabolic and electric brain patterns during pleasant and unpleasant emotions induced by music masterpieces." International Journal of Psychophysiology **65**(1): 69-84.

Ford, J. H., et al. (2011). "Differential neural activity during search of specific and general autobiographical memories elicited by musical cues." Neuropsychologia **49**(9): 2514-2526.

Foss, A. H., et al. (2007). "Neural correlates of the Pythagorean ratio rules." Neuroreport **18**(15): 1521-1525.

Fujisawa, T. X. and N. D. Cook (2011). "The perception of harmonic triads: an fMRI study." Brain imaging and behavior **5**(2): 109-125.

Gann, E. (2010). "The effects of therapeutic hip-hop activity groups on perception of self and social supports in at-risk urban adolescents." Unpublished doctoral dissertation. The Wright Institute, Berkeley, CA.

Green, A. C., et al. (2008). "Music in minor activates limbic structures: a relationship with dissonance?" Neuroreport **19**(7): 711-715.

Guetin, S., et al. (2009). "Effect of music therapy on anxiety and depression in patients with Alzheimer’s type dementia: randomised, controlled study." Dementia and geriatric cognitive disorders **28**(1): 36-46.

Gupta, U. and B. Gupta (2005). "Psychophysiological responsivity to Indian instrumental music." Psychology of music **33**(4): 363-372.

Harmat, L. and T. Theorell (2010). "Heart rate variability during singing and flute playing." Music and Medicine **2**(1): 10-17.

Haslam, C., et al. (2014). "Social identification moderates cognitive health and well-being following story-and song-based reminiscence." Aging & Mental Health **18**(4): 425-434.

Hays, T. (2005). "Well-being in later life through music." Australasian Journal on Ageing **24**(1): 28-32.

Hays, T. and V. Minichiello (2005). "The contribution of music to quality of life in older people: An Australian qualitative study." Ageing & Society **25**(2): 261-278.

Hicks-Moore, S. L. (2005). "Relaxing music at mealtime in nursing homes." Journal of gerontological nursing **31**(12): 26-32.

Hillman, S. (2002). "Participatory singing for older people: A perception of benefit." Health education.

Huang, R.-H. and Y.-N. Shih (2011). "Effects of background music on concentration of workers." Work **38**(4): 383-387.

Hugdahl, K., et al. (1999). "Brain activation during dichotic presentations of consonant-vowel and musical instrument stimuli: a 15O-PET study." Neuropsychologia **37**(4): 431-440.

Ifrach, E. R. and A. Miller (2016). "Social action art therapy as an intervention for compassion fatigue." The Arts in Psychotherapy **50**: 34-39.

Italia, S., et al. (2008). "Evaluation and art therapy treatment of the burnout syndrome in oncology units." Psycho-Oncology: Journal of the Psychological, Social and Behavioral Dimensions of Cancer **17**(7): 676-680.

James, C. E., et al. (2008). "Early neuronal responses in right limbic structures mediate harmony incongruity processing in musical experts." Neuroimage **42**(4): 1597-1608.

Janata, P. (2009). "The neural architecture of music-evoked autobiographical memories." Cerebral Cortex **19**(11): 2579-2594.

Jerde, T. A., et al. (2011). "Dissociable systems of working memory for rhythm and melody." Neuroimage **57**(4): 1572-1579.

Karpavičiūtė, S. and J. Macijauskienė (2016). "The impact of arts activity on nursing staff well-being: An intervention in the workplace." International Journal of Environmental Research and Public Health **13**(4): 435.

Kenny, D. T., et al. (2004). "Music performance anxiety and occupational stress amongst opera chorus artists and their relationship with state and trait anxiety and perfectionism." Journal of anxiety disorders **18**(6): 757-777.

Kim, D. S., et al. (2011). "Effects of music therapy on mood in stroke patients." Yonsei medical journal **52**(6): 977-981.

Knösche, T. R., et al. (2005). "Perception of phrase structure in music." Human brain mapping **24**(4): 259-273.

Koelsch, S., et al. (2008). "Amygdala activity can be modulated by unexpected chord functions during music listening." Neuroreport **19**(18): 1815-1819.

Koelsch, S., et al. (2006). "Investigating emotion with music: an fMRI study." Human brain mapping **27**(3): 239-250.

Kravits, K., et al. (2010). "Self-care strategies for nurses: A psycho-educational intervention for stress reduction and the prevention of burnout." Applied Nursing Research **23**(3): 130-138.

Lally, E. (2009). "The power to heal us with a smile and a song: Senior well-being, music-based participatory arts and the value of qualitative evidence." Journal of Arts & Communities **1**(1): 25-44.

Lane, D. (1994). "Effects of music therapy on immune function of hospitalized patients." Qual. Life–A Nurs Challenge **3**: 74-80.

Lashua, B. (2005). Making music, remaking leisure in the Beat of Boyle St, PhD thesis. University of Alberta, Edmonton.

Latimer, M. E. (2008). "Our voices enlighten, inspire, heal and empower. A mixed methods investigation of demography, sociology, and identity acquisition in a gay men's chorus." International Journal of Research in Choral Singing **3**(1): 23-38.

Lee, Y.-S., et al. (2011). "Investigation of melodic contour processing in the brain using multivariate pattern-based fMRI." Neuroimage **57**(1): 293-300.

Lee, Y. Y., et al. (2010). "Effectiveness of music intervention on the quality of life of older people." Journal of advanced nursing **66**(12): 2677-2687.

Lerner, Y., et al. (2009). "Eyes wide shut: amygdala mediates eyes-closed effect on emotional experience with music." PloS one **4**(7): e6230.

Lesiuk, T. (2008). "The effect of preferred music listening on stress levels of air traffic controllers." The Arts in Psychotherapy **35**(1): 1-10.

Lesiuk, T. (2010). "The effect of preferred music on mood and performance in a high-cognitive demand occupation." Journal of music therapy **47**(2): 137-154.

Lesta, B. and P. Petocz (2006). "Familiar Group Singing: Addressing Mood and Social Behaviour of Residents with Dementia Displaying Sundowning." Australian Journal of Music Therapy **17**.

Lord, V. M., et al. (2012). "Singing classes for chronic obstructive pulmonary disease: a randomized controlled trial." BMC Pulmonary Medicine **12**(1): 1-7.

Lotter, C. B. (2006). Circles of Courage: music therapy with adolescents in conflict with the law at a community based setting, University of Pretoria.

Louhivuori, J., et al. (2005). "'Singing Together': A Crosscultural Approach to the Meaning of Choirs As a Community." Cultural diversity in music education: 81.

Macpherson, C. F. (2008). "Peer-supported storytelling for grieving pediatric oncology nurses." Journal of Pediatric Oncology Nursing **25**(3): 148-163.

McCaffrey, R. and E. Freeman (2003). "Effect of music on chronic osteoarthritis pain in older people." Journal of advanced nursing **44**(5): 517-524.

McCaffrey, R. and R. Locsin (2006). "The effect of music on pain and acute confusion in older adults undergoing hip and knee surgery." Holistic nursing practice **20**(5): 218-224.

Menon, V. and D. J. Levitin (2005). "The rewards of music listening: response and physiological connectivity of the mesolimbic system." Neuroimage **28**(1): 175-184.

Miskovic, D., et al. (2008). "Randomized controlled trial investigating the effect of music on the virtual reality laparoscopic learning performance of novice surgeons." Surgical endoscopy **22**(11): 2416-2420.

Mitterschiffthaler, M. T., et al. (2007). "A functional MRI study of happy and sad affective states induced by classical music." Human brain mapping **28**(11): 1150-1162.

Mizuno, T. and M. Sugishita (2007). "Neural correlates underlying perception of tonality-related emotional contents." Neuroreport **18**(16): 1651-1655.

Mok, E. and K.-Y. Wong (2003). "Effects of music on patient anxiety." AORN journal **77**(2): 396-410.

Mutschler, I., et al. (2010). "Time scales of auditory habituation in the amygdala and cerebral cortex." Cerebral Cortex **20**(11): 2531-2539.

Nakamura, S., et al. (1999). "Analysis of music–brain interaction with simultaneous measurement of regional cerebral blood flow and electroencephalogram beta rhythm in human subjects." Neuroscience letters **275**(3): 222-226.

Ohnishi, T., et al. (2001). "Functional anatomy of musical perception in musicians." Cerebral Cortex **11**(8): 754-760.

Okada, K., et al. (2009). "Effects of music therapy on autonomic nervous system activity, incidence of heart failure events, and plasma cytokine and catecholamine levels in elderly patients with cerebrovascular disease and dementia." International heart journal **50**(1): 95-110.

Pallesen, K. J., et al. (2005). "Emotion processing of major, minor, and dissonant chords: a functional magnetic resonance imaging study." Annals of the New York Academy of Sciences **1060**(1): 450-453.

Pallesen, K. J., et al. (2009). "Cognitive and emotional modulation of brain default operation." Journal of cognitive neuroscience **21**(6): 1065-1080.

Pallesen, K. J., et al. (2010). "Cognitive control in auditory working memory is enhanced in musicians." PloS one **5**(6): e11120.

Pickles, V. (2003). "Music and the third age." Psychology of music **31**(4): 415-423.

Potash, J. S., et al. (2014). "Can art therapy reduce death anxiety and burnout in end-of-life care workers? A quasi-experimental study." International journal of palliative nursing **20**(5): 233-240.

Rice, K. L., et al. (2014). "Using second life to facilitate peer storytelling for grieving oncology nurses." Ochsner Journal **14**(4): 551-562.

Salzano, A. T., et al. (2013). "The effectiveness of a collaborative art-making task on reducing stress in hospice caregivers." The Arts in Psychotherapy **40**(1): 45-52.

Sands, S. A., et al. (2008). "Pediatric narrative oncology: interprofessional training to promote empathy, build teams, and prevent burnout." J Support Oncol **6**(7): 307-312.

Särkämö, T., et al. (2014). "Cognitive, emotional, and social benefits of regular musical activities in early dementia: randomized controlled study." The Gerontologist **54**(4): 634-650.

Satoh, M., et al. (2001). "Activated brain regions in musicians during an ensemble: a PET study." Cognitive Brain Research **12**(1): 101-108.

Shih, Y.-N., et al. (2016). "Elucidating the relationship between work attention performance and emotions arising from listening to music." Work **55**(2): 489-494.

Shih, Y.-N., et al. (2009). "Correlation between work concentration level and background music: A pilot study." Work **33**(3): 329-333.

Shih, Y.-N., et al. (2012). "Background music: Effects on attention performance." Work **42**(4): 573-578.

Shimizu, N., et al. (2013). "Effects of movement music therapy with the Naruko clapper on psychological, physical and physiological indices among elderly females: a randomized controlled trial." Gerontology **59**(4): 355-367.

Silber, L. (2005). "Bars behind bars: the impact of a women's prison choir on social harmony." Music education research **7**(2): 251-271.

Skingley, A. and H. Bungay (2010). "The Silver Song Club Project: singing to promote the health of older people." British journal of community nursing **15**(3): 135-140.

Snow, S. and M. D'Amico (2010). "The drum circle project: A qualitative study with at-risk youth in a school setting." Canadian Journal of Music Therapy **16**(1).

Solé, C., et al. (2010). "Contributions of music to aging adults' quality of life." Journal of music therapy **47**(3): 264-281.

Speer, S. (2011). "Effect of Background Music, Speech and Silence on Office Workers' Selective Attention."

Suda, M., et al. (2008). "Cortical responses to Mozart's sonata enhance spatial-reasoning ability." Neurological Research **30**(9): 885-888.

Sun, J. and N. Buys (2013). "Participatory community singing program to enhance quality of life and social and emotional well-being in Aboriginal and Torres Strait Islander Australians with chronic diseases." International Journal on Disability and Human Development **12**(3): 317-323.

Sun, J., et al. (2013). "The role of Tai Chi, cultural dancing, playing a musical instrument and singing in the prevention of chronic disease in Chinese older adults: A mind–body meditative approach." International Journal of Mental Health Promotion **15**(4): 227-239.

Sung, H. C., et al. (2010). "A preferred music listening intervention to reduce anxiety in older adults with dementia in nursing homes." Journal of Clinical Nursing **19**(7-8): 1056-1064.

Suzuki, M., et al. (2005). "Behavioral, stress and immunological evaluation methods of music therapy in elderly patients with senile dementia." Nihon Ronen Igakkai zasshi. Japanese journal of geriatrics **42**(1): 74-82.

Thaut, M. H., et al. (2009). "Distinct cortico-cerebellar activations in rhythmic auditory motor synchronization." Cortex **45**(1): 44-53.

Twiss, E., et al. (2006). "The effect of music listening on older adults undergoing cardiovascular surgery." Nursing in critical care **11**(5): 224-231.

Tyson, E. H. (2002). "Hip hop therapy: An exploratory study of a rap music intervention with at-risk and delinquent youth." Journal of Poetry Therapy **15**(3): 131-144.

Warth, M., et al. (2015). "Music therapy in palliative care: A randomized controlled trial to evaluate effects on relaxation." Deutsches Ärzteblatt International **112**(46): 788.

Wlodarczyk, N. (2007). "The effect of music therapy on the spirituality of persons in an in-patient hospice unit as measured by self-report." Journal of music therapy **44**(2): 113-122.

Wlodarczyk, N. (2013). "The effect of a group music intervention for grief resolution on disenfranchised grief of hospice workers." Progress in palliative care **21**(2): 97-106.

Wlodarczyk, N. M. (2010). "Effect of a Single-Session Music Therapy Group Intervention for Grief Resolution on the Disenfranchised Grief of Hospice Workers."

Woodward, S. C., et al. (2007). "South Africa, the arts and youth in conflict with the law." International Journal of Community Music **1**(1): 69-88.

WU, S. M. (2002). "Effects of music therapy on anxiety, depression and self-esteem of undergraduates." Psychologia **45**(2): 104-114.

**Included studies – Non-communicable disease risk**

Balbag, M. A., et al. (2014). "Playing a musical instrument as a protective factor against dementia and cognitive impairment: A population-based twin study." International Journal of Alzheimer's Disease **2014**.

Bygren, L. O., et al. (1996). "Attendance at cultural events, reading books or periodicals, and making music or singing in a choir as determinants for survival: Swedish interview survey of living conditions." BMJ **313**(7072): 1577-1580.

Chen, M., et al. (2013). "Different physical activity subtypes and risk of metabolic syndrome in middle-aged and older Chinese people." PloS one **8**(1): e53258.

Ekholm, O. and L. O. Bonde (2018). Music and Health in Everyday Life in Denmark: Associations Between the Use of Music and Health-Related Outcomes in Adult Danes. Music and Public Health, Springer**:** 15-31.

Hughes, T. F., et al. (2010). "Engagement in reading and hobbies and risk of incident dementia: the MoVIES project." American Journal of Alzheimer's Disease & Other Dementias® **25**(5): 432-438.

Konlaan, B. B., et al. (2000). "Visiting the cinema, concerts, museums or art exhibitions as determinant of survival: a Swedish fourteen-year cohort follow-up." Scandinavian Journal of Public Health **28**(3): 174-178.

Lin, W., et al. (2019). "Performing different kinds of physical exercise differentially attenuates the genetic effects on obesity measures: Evidence from 18,424 Taiwan Biobank participants." PLoS Genetics **15**: e1008277.

Merom, D., et al. (2016). "Dancing participation and cardiovascular disease mortality: a pooled analysis of 11 population-based British cohorts." American journal of preventive medicine **50**(6): 756-760.

Verghese, J., et al. (2003). "Leisure activities and the risk of dementia in the elderly." New England Journal of Medicine **348**(25): 2508-2516.

#### Included studies – Heart Rate, Dance

Aygün, C., et al. (2018). "Active video dancing game provides high-intensity exercise for hip-hop dancers and non-dancers." South African Journal for Research in Sport, Physical Education and Recreation **40**(2): 1-10.

Baillie, Y., et al. (2007). "Highland dance: heart-rate and blood lactate differences between competition and class." International Journal of Sports Physiology and Performance **2**(4): 371-376.

Beck, S., et al. (2018). "Changes in Energy Demand of Dance Activity and Cardiorespiratory Fitness During 1 Year of Vocational Contemporary Dance Training." The Journal of Strength & Conditioning Research **32**(3): 841-848.

Blanksby, B. and P. Reidy (1988). "Heart rate and estimated energy expenditure during ballroom dancing." British Journal of Sports Medicine **22**(2): 57-60.

Bria, S., et al. (2011). "Physiological characteristics of elite sport-dancers." The Journal of sports medicine and physical fitness **51**(2): 194-203.

Bronner, S., et al. (2016). "Physiological and psychophysiological responses to an exer-game training protocol." Journal of science and medicine in sport **19**(3): 267-271.

Bronner, S., et al. (2013). "Energy cost and game flow of 5 exer-games in trained players." American journal of health behavior **37**(3): 369-380.

Bronstein, M., et al. (1990). "Physiological response to low-impact aerobic dance by trained dancers with and without handheld weights." Ann. Sports Med **5**: 74-77.

Çakir-Atabek, H., et al. (2020). "Active Video Games versus Traditional Exercises: Energy Expenditure and Blood Lactate Responses." Research quarterly for exercise and sport **91**(2): 188-196.

Canabrava, K. L., et al. (2018). "Energy expenditure and intensity of active video games in children and adolescents." Research quarterly for exercise and sport **89**(1): 47-56.

Carroll, M. W., et al. (1991). "The metabolic cost of two ranges of arm position height with and without hand weights during low impact aerobic dance." Research quarterly for exercise and sport **62**(4): 420-423.

Cohen, J. L., et al. (1982). "Cardiorespiratory responses to ballet exercise and the VO2max of elite ballet dancers." Medicine and Science in Sports and Exercise **14**(3): 212-217.

Cugusi, L., et al. (2015). "A New Type of Physical Activity from an Ancient Tradition: The Sardinian Folk Dance “Ballu Sardu”." Journal of dance medicine & science **19**(3): 118-123.

Darby, L. A., et al. (1995). "The effects of cadence, impact, and step on physiological responses to aerobic dance exercise." Research quarterly for exercise and sport **66**(3): 231-238.

De Angelis, M., et al. (1998). "Oxygen uptake, heart rate and blood lactate concentration during a normal training session of an aerobic dance class." European journal of applied physiology and occupational physiology **78**(2): 121-127.

Delextrat, A. and E. Neupert (2016). "Physiological load associated with a Zumba® fitness workout: a comparison pilot study between classes and a DVD." Journal of sports sciences **34**(1): 47-55.

Domene, P. A. and C. Easton (2014). "Combined triaxial accelerometry and heart rate telemetry for the physiological characterization of Latin dance in non-professional adults." Journal of dance medicine & science **18**(1): 29-36.

Eason, J. M., et al. (2016). "A Comparison of Energy Expenditure and Heart Rate Response Between a Dance-Based Group Fitness Class and a Dance-Based Video Game on the Xbox Kinect." Cardiopulmonary Physical Therapy Journal **27**(2): 62-67.

Emerenziani, G. P., et al. (2013). "Exercise intensity and gender difference of 3 different salsa dancing conditions." Int J Sports Med **34**(4): 330-335.

Fawkner, S. G., et al. (2010). "Adolescent girls' energy expenditure during dance simulation active computer gaming." Journal of sports sciences **28**(1): 61-65.

Forte, R., et al. (2001). "Cardiovascular response during low-intensity step-aerobic dance in middle-aged subjects." European Journal of Sport Science **1**(3): 1-7.

Gao, Z., et al. (2009). "Middle School Student's Heart Rates during Different Curricular Activities in Physical Education." ICHPER-SD Journal of Research **4**(1): 21-24.

Giancotti, G. F., et al. (2018). "Energy expenditure and perceived exertion during active video games in relation to player mode and gender." Kinesiology **50**(1): 18-24.

Graf, D. L., et al. (2009). "Playing active video games increases energy expenditure in children." Pediatrics **124**(2): 534-540.

Grant, S., et al. (2002). "A comparison of physiological responses and rating of perceived exertion in two modes of aerobic exercise in men and women over 50 years of age." British Journal of Sports Medicine **36**(4): 276-281.

Grier, T. D., et al. (2002). "Metabolic cost of aerobic dance bench stepping at varying cadences and bench heights." Journal of strength and conditioning research **16**(2): 242-249.

Heil, D. P., et al. (2019). "The Energy Expenditure of Tinikling: A Culturally Relevant Filipino Dance." International Journal of Exercise Science **12**(4): 111.

Kraft, J. A., et al. (2011). "Heart rate and perceived exertion during self-selected intensities for exergaming compared to traditional exercise in college-age participants." The Journal of Strength & Conditioning Research **25**(6): 1736-1742.

La Torre, A., et al. (2005). "Cardiovascular responses to aerobic step dance sessions with and without appendicular overload." Journal of sports medicine and physical fitness **45**(3): 264.

Léger, L. A. (1982). "Energy cost of disco dancing." Research quarterly for exercise and sport **53**(1): 46-49.

Liiv, H., et al. (2014). "Physiological characteristics of elite dancers of different dance styles." European Journal of Sport Science **14**(sup1): S429-S436.

Lin, J.-H. (2015). "“Just Dance”: The effects of exergame feedback and controller use on physical activity and psychological outcomes." Games for Health Journal **4**(3): 183-189.

Lowe, A., et al. (2010). "Accuracy of polar F6 in estimating the energy cost of aerobic dance bench stepping in college-age females." Journal of sports medicine and physical fitness **50**(4): 385.

Maciejczyk, M. and A. Feć (2013). "Evaluation of aerobic capacity and energy expenditure in folk dancers." Human movement **14**(1): 76-81.

Marks, D. W., et al. (2015). "Greater physiological responses while playing XBox Kinect compared to Nintendo Wii." International Journal of Exercise Science **8**(2): 7.

Nelson, L., et al. (2011). "Heart rates of elementary physical education students during the dancing classrooms program." Research quarterly for exercise and sport **82**(2): 256-263.

Nicholas, J. C., et al. (2019). "Pole Dancing for Fitness: The Physiological and Metabolic Demand of a 60-Minute Class." The Journal of Strength & Conditioning Research **33**(10): 2704-2710.

Oh, Y., et al. (2017). "Comparisons of Heart Rate and Energy Expenditure During Exergaming in College-age Adults." International Journal of Computer Science in Sport **16**(3): 197-206.

Oliveira, S. M., et al. (2010). "Physiological responses to a tap dance choreography: comparisons with graded exercise test and prescription recommendations." The Journal of Strength & Conditioning Research **24**(7): 1954-1959.

Pennington, C. G. and L. P. Nelson (2020). "Physical Activity Contributions of Dancing Classrooms Program on Middle School Students." The Physical Educator **77**(2).

Pilch, W., et al. (2017). "Energy expenditure and lactate concentration in sports dancers in a simulated final round of the standard style competition." Human movement **18**(2): 62-67.

Powers, M. M., et al. (2016). "A Comparison of Energy Expenditure Across 3 Gaming Platforms Using an Activity-Based Dance Game." Cardiopulmonary Physical Therapy Journal **27**(4): 108-114.

Rauber, S. B., et al. (2018). "Oxygen consumption and energy expenditure during and after street games, active video games and tv." Revista Brasileira de Medicina do Esporte **24**(5): 338-342.

Rixon, K. P., et al. (2006). "Analysis of the assessment of caloric expenditure in four modes of aerobic dance." Journal of strength and conditioning research **20**(3): 593.

Rockefeller, K. A. and E. Burke (1979). "Psycho-physiological analysis of an aerobic dance programme for women." British Journal of Sports Medicine **13**(2): 77-80.

Rodrigues-Krause, J., et al. (2014). "Oxygen consumption and heart rate responses to isolated ballet exercise sets." Journal of dance medicine & science **18**(3): 99-105.

Rodrigues-Krause, J., et al. (2018). "Cardiorespiratory responses of a dance session designed for older women: A cross sectional study." Experimental gerontology **110**: 139-145.

Rodrigues-Krause, J., et al. (2014). "Ballet dancers cardiorespiratory, oxidative and muscle damage responses to classes and rehearsals." European Journal of Sport Science **14**(3): 199-208.

Rousanoglou, E. and K. Boudolos (2005). "Ground reaction forces and heart rate profile of aerobic dance instructors during a low and high impact exercise programme." Journal of sports medicine and physical fitness **45**(2): 162.

Sanders, D. J., et al. (2019). "Training Demands and Physiological Profile of Cross-Disciplined Collegiate Female Dancers." Journal of strength and conditioning research.

Schaefeer, S. A., et al. (1995). "Perceived exertion and metabolic responses of women during aerobic dance exercise." Perceptual and motor skills **81**(2): 691-700.

Schaeffer-Gerschutz, S. A., et al. (2000). "Differentiated ratings of perceived exertion and physiological responses during aerobic dance steps by impact/type of arm movement." Perceptual and motor skills **90**(2): 457-471.

Schneekloth, B. and G. A. Brown (2018). "Comparison of physical activity during Zumba with a human or video game instructor." International Journal of Exercise Science **11**(4): 1019.

Sekulic, D., et al. (2006). "Students'-instructors' physiological responses during step aerobic exercise." Biology of Sport **23**(2): 171.

Seliger, V., et al. (1970). "Evaluation of stage artist's activities on basis of telemetrical measurements of heart rates." Internationale Zeitschrift für angewandte Physiologie einschließlich Arbeitsphysiologie **28**(2): 86-104.

Sell, K., et al. (2008). "Energy expenditure during physically interactive video game playing in male college students with different playing experience." Journal of American College Health **56**(5): 505-512.

Smallwood, S. R., et al. (2012). "Physiologic responses and energy expenditure of kinect active video game play in schoolchildren." Archives of pediatrics & adolescent medicine **166**(11): 1005-1009.

Stephens, N. and M. Wyon (2020). "Physiological Characteristics of Musical Theatre Performers and the Effect on Cardiorespiratory Demand Whilst Singing and Dancing." Medical Problems of Performing Artists **35**(1): 54-58.

Strejcová, B., et al. (2013). "The energetic cost and cardiorespiratory changes during Zumba exercise in middle aged women " International Journal of Sports Sciences & Fitness **3**(2).

Tan, B., et al. (2002). "Aerobic demands of the dance simulation game." International journal of sports medicine **23**(2): 125-129.

Tarrant, K. and L. McNaughton (1997). "A comparison of the metabolic effects of high and low impact aerobic dance exercise." Research in Sports Medicine: An International Journal **7**(3-4): 255-264.

Thompson, W. R., et al. (1991). "The effect of hand-held weights on the physiological responses to aerobic dance." The Journal of Strength & Conditioning Research **5**(4): 208-212.

Thomsen, D. and D. L. Ballor (1991). "Physiological responses during aerobic dance of individuals grouped by aerobic capacity and dance experience." Research quarterly for exercise and sport **62**(1): 68-72.

Unnithan, V. B., et al. (2006). "Evaluation of the energy cost of playing a dance simulation video game in overweight and non-overweight children and adolescents." International journal of sports medicine **27**(10): 804-809.

Usagawa, T., et al. (2014). "Metabolic equivalent determination in the cultural dance of hula." International journal of sports medicine **35**(5): 399.

Ušpurienė, A. B. and A. Čepulėnas (2012). "Physical Load Intensity in Standard and Latin American Sports Dancing Programmes for Juvenile Dancers." Baltic Journal of Sport and Health Sciences **2**(85).

Wigaeus, E. (1980). "Physical demands during folk dancing." European journal of applied physiology and occupational physiology **45**(2-3): 177-183.

Wyon, M., et al. (2002). "The cardiorespiratory responses to modern dance classes: differences between university, graduate, and professional classes." Journal of dance medicine & science **6**(2): 41-45.

Wyon, M. A., et al. (2004). "Oxygen uptake during modern dance class, rehearsal, and performance." The Journal of Strength & Conditioning Research **18**(3): 646-649.

Wyon, M. A., et al. (2018). "Cardiorespiratory profile and performance demands of elite hip-hop dancers: breaking and new style." Medical Problems of Performing Artists **33**(3): 198-204.

Zhu, W., et al. (2018). "Characterizing the Aerobic and Anaerobic Energy Costs of Polynesian Dances." International Journal of Exercise Science **11**(4): 1156.

**Included studies – Heart Rate, Music**

Barr, D., et al. (2000). "Physiological assessment of bagpipers: a preliminary study of cardiopulmonary parameters." Medical Problems of Performing Artists **15**(2): 51-55.

De La Rue, S., et al. (2013). "Energy expenditure in rock/pop drumming." International journal of sports medicine **34**(10): 868-872.

Dye, D. and V. Barry (2016). "The Effect of Metronome Use on Heart Rates and RPE for Drum Corps Percussionists." International Journal of Exercise Science **9**(4): 3.

Erdmann, L. D., et al. (2003). "Adolescents' energy cost in marching band." Perceptual and motor skills **97**(2): 639-646.

Hahnengress, M. L. and D. Böning (2010). "Cardiopulmonary changes during clarinet playing." European journal of applied physiology **110**(6): 1199-1208.

Harmat, L. and T. Theorell (2010). "Heart rate variability during singing and flute playing." Music and Medicine **2**(1): 10-17.

Horswill, C., et al. (1995). "Energy expenditure in adolescents during low intensity, leisure activities." Medicine & Science in Sports & Exercise.

Hunsaker, L. A. (1994). "Heart rate and rhythm responses during trumpet playing." Medical Problems of Performing Artists **9**: 69-69.

Iñesta, C., et al. (2008). "Heart rate in professional musicians." Journal of Occupational Medicine and Toxicology **3**(1): 1-11.

Mulcahy, D., et al. (1990). "Circadian variation of heart rate is affected by environment: a study of continuous electrocardiographic monitoring in members of a symphony orchestra." Heart **64**(6): 388-392.

Robertson, D. U., et al. (2010). "Cardiac response during trumpet playing." Medical Problems of Performing Artists **25**(1): 16-21.

Romero, B., et al. (2016). "Metabolic demands of heavy metal drumming." International Journal of Kinesiology and Sports Science **4**(3): 32-36.

Seliger, V., et al. (1970). "Evaluation of stage artist's activities on basis of telemetrical measurements of heart rates." Internationale Zeitschrift für angewandte Physiologie einschließlich Arbeitsphysiologie **28**(2): 86-104.

Strand, B. and C. Sommer (2005). "Should marching band be allowed to replace physical education credits: An analysis." The Physical Educator **62**(3).

Studer, R. K., et al. (2014). "Psychophysiological activation during preparation, performance, and recovery in high-and low-anxious music students." Applied Psychophysiology and Biofeedback **39**(1): 45-57.

Vellers, H. L., et al. (2015). "Heart rate response of professional musicians when playing music." Medical Problems of Performing Artists **30**(2): 100-105.
